# Quantitative bias analysis methods for summary level epidemiologic data in the peer-reviewed literature: a systematic review

**DOI:** 10.1101/2024.04.23.24306205

**Authors:** Xiaoting Shi, Ziang Liu, Mingfeng Zhang, Wei Hua, Jie Li, Joo-Yeon Lee, Sai Dharmarajan, Kate Nyhan, Ashley Naimi, Timothy L. Lash, Molly M. Jeffery, Joseph S. Ross, Zeyan Liew, Joshua D. Wallach

## Abstract

**Objective:** Quantitative bias analysis (QBA) methods evaluate the impact of biases arising from systematic errors on observational study results. This systematic review aimed to summarize the range and characteristics of quantitative bias analysis (QBA) methods for summary level data published in the peer-reviewed literature.

**Study Design and Setting:** We searched MEDLINE, Embase, Scopus, and Web of Science for English-language articles describing QBA methods. For each QBA method, we recorded key characteristics, including applicable study designs, bias(es) addressed; bias parameters, and publicly available software. The study protocol was pre-registered on the Open Science Framework (https://osf.io/ue6vm/).

**Results:** Our search identified 10,249 records, of which 53 were articles describing 57 QBA methods for summary level data. Of the 57 QBA methods, 51 (89%) were explicitly designed for observational studies, 2 (4%) for non-randomized interventional studies, and 4 (7%) for meta-analyses. There were 29 (51%) QBA methods that addressed unmeasured confounding, 20 (35%) misclassification bias, 5 (9%) selection bias, and 3 (5%) multiple biases. 38 (67%) QBA methods were designed to generate bias-adjusted effect estimates and 18 (32%) were designed to describe how bias could explain away observed findings. 22 (39%) articles provided code or online tools to implement the QBA methods.

**Conclusion:** In this systematic review, we identified a total of 57 QBA methods for summary level epidemiologic data published in the peer-reviewed literature. Future investigators can use this systematic review to identify different QBA methods for summary level epidemiologic data.

**What is New?:** *Key findings:* This systematic review identified 57 quantitative bias analysis (QBA) methods for summary level data from observational and non-randomized interventional studies. Overall, there were 29 QBA methods that addressed unmeasured confounding, 20 that addressed misclassification bias, 5 that addressed selection bias, and 3 that addressed multiple biases.

**What this adds to what is known related to methods research within the field of clinical epidemiology?:** This systematic review provides an overview of the range and characteristics of QBA methods for summary level epidemiologic that are published in the peer-reviewed literature and that can be used by researchers within the field of clinical epidemiology.

**What is the implication, what should change now?:** This systematic review may help future investigators identify different QBA methods for summary level data. However, investigators should carefully review the original manuscripts to ensure that any assumptions are fulfilled, that the necessary bias parameters are available and accurate, and that all interpretations and conclusions are made with caution.

## Introduction

Randomized controlled trials (RCTs) are often considered the gold standard for estimating causal effects in clinical research. However, RCTs are not feasible for all clinical questions (e.g., when randomization is not ethical and for estimating treatment effects in populations not included in efficacy trials), often have strict inclusion and exclusion criteria, face recruitment and retention difficulties, and take a long time to complete.^1^ These limitations and operational challenges, which can lead to higher costs and lower generalizability to real-world settings, highlight the important role of observational studies and non-randomized interventional studies.^2^ Although these study designs can overcome some of the challenges faced by RCTs, they are more susceptible to systematic errors (i.e., uncontrolled confounding, misclassification, and selection bias), which can contribute to the uncertainty of a study’s results.^3^ Therefore, analytical methods are needed to help assess the impact of systematic errors on the findings from observational and non-randomized interventional studies.

Quantitative bias analysis (QBA) methods estimate the direction, magnitude, and uncertainty resulting from systematic errors in a study, and can be used to explore how sensitive study findings are to assumptions and bias parameters.^4,5^ QBA methods, which are often classified across six categories - simple sensitivity analysis, multidimensional analysis, probabilistic analysis, direct bias modeling and missing data methods, Bayesian analysis, and multiple bias modeling - can be used to estimate what the observed association from a study would have been in the absence of systematic error (**Table 1)**.^4,5^ Although numerous QBA methods have been published in the literature, there are several challenges that have limited the widespread application of QBA methods in observational and non-randomized interventional studies. First, some QBA methods require more extensive statistical and programming expertise.^4,6,7,8^ Second, it may be difficult to assign reasonable values to the bias parameters and priors for QBA methods. Third, some QBA methods can only be conducted using the individual participant level data from a study.^4^ However, certain QBA methods can be conducted using simple equations and summary level data based on published study results, including two-by-two contingency tables, effect estimates and corresponding 95% confidence intervals, bias parameters from the literature, assumptions, and educated guesses.^4^ These QBA methods for summary level data may be more straightforward to include as sensitivity analyses in observational studies. However, little is known about the full range and characteristics of available QBA methods in the peer-reviewed literature that only require summary level data.

**Table 1.** Common classification system for quantitative bias analysis methods^4,9^.

| **Classification** | **Assignment of bias parameters** | **Number of biases accounted for** | **Output** |
| --- | --- | --- | --- |
| Simple sensitivity analysis | One fixed value assigned to each bias parameter | One at a time | Single bias-adjusted effect estimate |
| Multidimensional analysis | More than one value assigned to each bias parameter | One at a time | Range of bias-adjusted effect estimates |
| Probabilistic analysis | Probability distributions assigned to each bias parameter | One at a time | Frequency distribution of bias-adjusted effect estimates |
| Direct bias modelling and missing data methods | Estimate and variance obtained from information internal or external to dataset | One at a time | Distribution of bias-adjusted effect estimates |
| Bayesian analysis | Probability distributions assigned to each bias parameter | Multiple biases at a time | Distribution of bias-adjusted effect estimates |
| Multiple bias modelling | Probability distributions assigned to each bias parameter | Multiple biases at a time | Frequency distribution of bias-adjusted effect estimates |

To address these knowledge gaps, we conducted a systematic review to comprehensively identify and summarize QBA methods for summary level data from observational and non-randomized interventional studies that have been proposed in the peer-reviewed literature.

## Methods

This review was reported following the Preferred Reporting Items for Systematic Reviews and Meta-Analyses (PRISMA) 2020 statement (**Appendix 1**).^10^ We pre-registered our study protocol on the Open Science Framework (https://osf.io/ue6vm/).

### Literature search and study selection

Working with an experienced librarian (KN), we developed a systematic literature search capturing the broad concepts of bias analysis and epidemiologic methods (**Appendix 2**). On January 10^th^ 2022, a research librarian (KN) performed a comprehensive search of multiple databases: MEDLINE (Ovid ALL, from 1949), Embase (Ovid, from 1974), Scopus, and Web of Science Core Collection as licensed by Yale University. No date limit was applied. The search retrieved a total of 13,356 records, which we pooled in EndNote (https://endnote.com/), deduplicated, and uploaded to Covidence (https://covidence.org/). On October 14^th^ 2022, 2,702 additional records were retrieved through backwards reference chaining. At least two independent investigators (XS, ZLiu, and/or JDW) screened records at the title-abstract and then full-text level. All uncertainties were discussed and reviewed by two additional authors (ZLiew and JDW).

### Eligibility criteria

Articles were considered eligible for inclusion if they were peer-reviewed English-language publications describing, evaluating, and/or comparing QBA methods for summary level data from observational (cohort, case-control, and cross-sectional studies), nonrandomized interventional studies (nonrandomized interventional studies, including single-arm studies with or without external controls, and quasi-experimental studies), and meta-analyses of observational study designs with no date limits applied. We included nonrandomized interventional studies because these study designs are also susceptible to systematic errors.^2^ We included methodological articles that described significant or slight modifications of previously published QBA methods focused on unmeasured confounding, misclassification (information bias), and selection bias. We excluded all conference abstracts, corrigendum, and non-peer-reviewed articles. Analytical methods that were designed to address biases but were not defined as QBA methods were excluded (i.e., inverse probability weighting, marginal structural models, g-estimation, covariate regression adjustment, propensity scores, missing data imputation, negative controls, instrumental variable analyses, restriction and mediation).^5^ We further excluded articles that only applied QBA methods in primary or sensitivity analyses, but did not propose new methods or modification to the existing methods, since a previous systematic review already focused on the application of QBA in observational studies.^5^ QBA methods that required individual participant level data (i.e., record-level data, raw data) were also excluded.

### Data collection

For all eligible articles, two independent investigators (XS and ZLiu) abstracted the following article characteristics: study title, first author, publication year, and digital object identifier (DOI).

For each eligible QBA method for summary level data, we recorded the following study characteristics: name of the method, applicable study design scenarios (i.e., cohort only, case-control only, cohort and case-control, non-randomized interventional studies and observational studies, or meta-analyses); sources of bias(es) addressed (i.e., unmeasured confounding, misclassification bias, selection bias, or multiple biases); bias parameters required to conduct the analysis; required data format for the exposure, confounder, and outcome (i.e., categorical, continuous, time-to-event/survival, multiple data types, or unclear); effect measure of interest (i.e., ratio measures [e.g., risk ratio, odds ratio, rate ratio, hazard ratio] and/or absolute measures [e.g., mean difference, risk difference] measures); output and type of output obtained from each method (i.e., explain-away [i.e., if the observed exposure-outcome relationship is explained away by the bias] or a corrected effect estimate); stated data assumptions and additional required features to implement each method; and the availability of publicly available software, tools, or websites to conduct the analyses. For study design, effect measure of interest, and stated data assumptions, we only recorded the information explicitly mentioned or used by the authors. Next, we recorded the main formula(s) and any explicitly mentioned considerations relevant to each QBA method. We then determined the relevant interpretation of the output of each method. Each article describing the eligible QBA methods were then reviewed for explicit discussions regarding the key similarities and differences between the eligible QBA methods. Last, we reviewed a prominent textbook on QBA methods to determine which of the identified methods were referenced and/or explained.^4^ All abstractions were reviewed and arbitrated by two reviewers (ZLiew and JDW).

### Analyses

Eligible QBA methods were classified into previously developed categories: simple sensitivity analysis, multidimensional analysis, probabilistic analysis, Bayesian analysis, direct bias modelling and missing data methods, and multiple bias modelling (**Table 1)**.^4^ Key characteristics were summarized using descriptive statistics.

### QBA method clusters

We chronologically grouped clusters of QBA methods that addressed the same systematic errors and where the authors noted that they were derivations or modified versions of previously developed methods. Methods that only mentioned or compared previous methods but did not lead into new methods or were not derived from previous methods were not considered (**Appendix 2**). Next, we classified the QBA methods based on their study characteristics (**Appendix 3**): study design (i.e., cohort study, case-control study, other observational study, nonrandomized interventional study or meta-analysis), bias type (i.e., unmeasured confounding, misclassification bias, or selection bias), result of interest (i.e., if the goal to explain-away or bias adjust the observed effect estimates), and the exposure, outcome, and confounder data types. For bias types with unique characteristics (e.g., nondifferential or differential misclassification bias), additional clusters were generated.

## Results

### Study selection

Of the 16,058 records that were identified (**Figure 1**), 5,809 were excluded as duplicates, leaving 10,249 articles for initial screening. We excluded 9,662 articles based on title and abstract. Among the 587 full-text articles assessed for eligibility, 534 articles were excluded, mostly because they focused on non-QBA methods (e.g., propensity scores and other approaches: 218, 41%) or described QBA methods for individual participant level data (170, 32%). We were left with 53 articles with 57 QBA methods that met the inclusion criteria.

**Figure 1.**
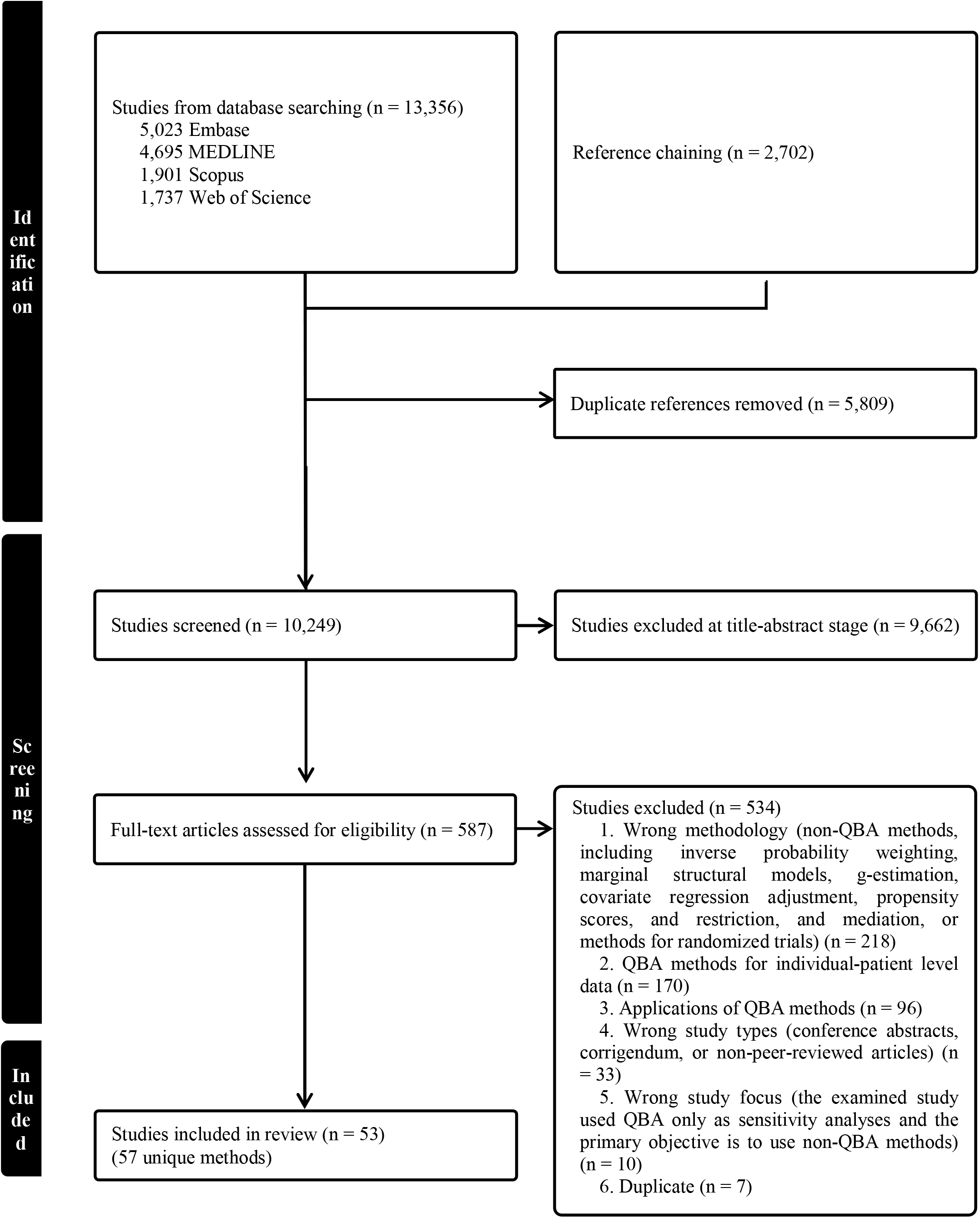
PRISMA flowchart. Footnotes: QBA: quantitative bias analysis.

### Description of included QBA methods

The 53 eligible articles described 57 QBA methods for summary level data (**Table 2**). Of these, we classified 36 (63%) as simple sensitivity analysis methods, 7 (12%) as multidimensional analysis methods, 4 (7%) as Bayesian analysis methods, 3 (5%) as probabilistic analysis methods, 3 (5%) as multiple bias modeling methods, and 1 (2%) as direct bias modeling (classification scheme in **Table 1**). There were 3 (5%) methods that were classified as simple or multidimensional analysis methods because it was possible to assign one or multiple values to the bias parameters. Overall, 21 (37%) methods were referenced in a QBA textbook,^4^ of which 11 (52%) were also described in detail.

**Table 2.** Summary of quantitative bias analysis methods for summary level data (n=57 methods).

| Study characteristics |  | n | % |
| --- | --- | --- | --- |
| Publication year | Median (interquartile range) | 2007<br>(1993-2018) |  |
|  | Full range | 1959-2021 |  |
| Method classification | Simple sensitivity analysis | 36 | 63 |
|  | Multidimensional analysis | 7 | 12 |
|  | Bayesian analysis | 4 | 7 |
|  | Probabilistic analysis | 3 | 5 |
|  | Multiple bias modeling | 3 | 5 |
|  | Direct bias modeling and missing data methods | 1 | 2 |
|  | Simple or multidimensional analysis | 3 | 5 |
| Applicable study scenarios | Observational study designs only | 51 | 89 |
|  | *Cohort only* | 6 | 12 |
|  | *Case-control only* | 13 | 26 |
|  | *Cohort and case-control* | 31 | 61 |
|  | *Other observational studies* | 1 | 2 |
|  | Non-randomized interventional studies and/or other observational study designs | 2 | 4 |
|  | Meta-analyses | 4 | 7 |
| Source(s) of bias addressed | Unmeasured confounding | 29 | 51 |
|  | *Single confounder only* | 22 | 76 |
|  | *Single and multiple confounders* | 7 | 24 |
|  | Misclassification bias | 20 | 35 |
|  | Exposure misclassification | 13 | 65 |
|  | *Differential only* | 1 | 8 |
|  | *Nondifferential only* | 6 | 46 |
|  | *Both differential and nondifferential* | 6 | 46 |
|  | Confounder misclassification | 2 | 10 |
|  | *Differential only* | 0 | 0 |
|  | *Nondifferential only* | 2 | 100 |
|  | *Both differential and nondifferential* | 0 | 0 |
|  | Outcome misclassification | 6 | 30 |
|  | *Differential only* | 1 | 17 |
|  | *Nondifferential only* | 3 | 50 |
|  | *Both differential and nondifferential* | 2 | 33 |
|  | Selection bias | 5 | 9 |
|  | Multiple biases | 3 | 5 |
| If cross-referenced by the textbook<sup>a</sup> | Not referenced | 36 | 63 |
|  | Referenced but not explained | 10 | 18 |
|  | Referenced and explained | 11 | 19 |
| Exposure data type | Categorical | 47 | 82 |
|  | Continuous only | 0 | 0 |
|  | Multiple data types | 10 | 18 |
| Outcome data type | Categorical | 37 | 65 |
|  | Continuous only | 2 | 4 |
|  | Time-to-event only | 0 | 0 |
|  | Multiple data types | 18 | 32 |
| Confounder data type | Categorical | 29 | 51 |
|  | Continuous only | 0 | 0 |
|  | Multiple data types | 14 | 25 |
|  | Unclear or not applicable | 14 | 25 |
| Effect estimates | Ratio measures<sup>b</sup> | 42 | 74 |
|  | Difference measures | 1 | 2 |
|  | Both | 11 | 19 |
|  | Other<sup>c</sup> | 2 | 4 |
|  | Unclear<sup>d</sup> | 1 | 2 |
| Result of interest | Corrected estimate(s) | 38 | 67 |
|  | Explain-away | 18 | 32 |
|  | Bias illustration<sup>d</sup> | 1 | 2 |
| Recommended software/tools | One tool | 16 | 28 |
|  | *Code/software package (R, SAS) only* | 10 | 63 |
|  | *Online tools only* | 1 | 6 |
|  | *Excel only* | 5 | 31 |
|  | Multiple tools | 6 | 11 |
|  | No tool | 35 | 61 |

Footnotes: <sup>a</sup>Fox et al 2021, Applying Quantitative Bias Analysis to Epidemiologic Data (2<sup>nd</sup> edition); <sup>b</sup>ratio measures include relative risk, rate ratio, odds ratio, and hazard ratio; difference measures include mean difference, risk difference and attributable fraction; <sup>c</sup>two methods work for regression coefficients; <sup>d</sup>this method uses bias plots to illustrate the potential range of bias, and has an unclear result of interest.

There were 51 (89%) QBA methods that were explicitly described as being suitable to use for observational studies, 2 (4%) for non-randomized interventional studies, and 4 (7%) for meta-analyses (**Table 2**).

### Sources of bias

There were 29 (51%) QBA methods designed to address unmeasured confounding, of which 22 (76%) were for studies that focused on examining a single unmeasured confounder (**Table 2)**. There were 20 (35%) methods for misclassification bias, of which 13 (65%) were for exposure misclassification, 2 (10%) for confounder misclassification, and 6 (30%) for outcome misclassification. There were 5 (9%) methods for selection bias and 3 (5%) for multiple biases at a time.

### Data types and effect estimates

There were 47 (82%) methods explicitly designed for studies where the exposure can be treated as a categorical variable and 10 (18%) for studies where the exposure can be treated as either categorical or continuous (i.e., multiple data types) (**Table 2**). There were 37 (65%) methods that were explicitly designed to accommodate only categorical outcome variables and 18 (32%) methods were described for multiple data types.

There were 42 (74%) QBA methods explicitly designed for studies with only ratio measures, 1 (2%) for only difference measures, and 11 (19%) for studies with both ratio and difference measures. Two-thirds (38, 67%) of the methods were designed to generate bias-adjusted effect estimates and 18 (32%) to describe how bias could fully explain away observed findings (i.e., to bias adjust non-null findings to the null).

### Software, tools, and code

Among the 53 articles describing the 57 QBA methods, 22 (39%) provided publicly available supplementary code or tools to implement the QBA methods; 3 noted that their code was available upon request.

### QBA method clusters

We identified two distinct clusters of QBA methods with the same fundamental form - confounding methods derived from Cornfield 1959 and Bross 1966 (15 [52%] of the 29 methods for unmeasured confounding) and the matrix correction methods for misclassification bias (6 [30%] of the 20 methods for misclassification bias) (**Appendix 2**).^11,12^

## Discussion

In this systematic review, we identified 53 articles describing 57 QBA methods for summary level data from observational studies and nonrandomized interventional studies in the peer-reviewed literature. While over 50% of these methods were designed to address unmeasured confounding, fewer than 10% were for selection bias. Approximately two-thirds of the QBA methods for summary level data were designed to generate bias-adjusted effect estimates and one-third were designed to describe how bias can explain away the observed findings from a study. Although this systematic review can be used to identify different QBA methods for summary level epidemiologic data, investigators should carefully review the original manuscripts to ensure that any assumptions are fulfilled, that the necessary bias parameters are available and accurate, and that all interpretations and conclusions are made with caution.

We found that most QBA methods for summary level epidemiologic data were for unmeasured confounding. In fact, over half of the QBA methods for confounding identified outlined that they were derived from Cornfield in 1959, who described methods to assess the impact of uncontrolled confounding when evaluating the association between smoking and lung cancer in two-by-two tables,^11^ and Bross 1966, who introduced a framework to analyze the bias due to a binary unmeasured confounder by relating an observed effect estimate to a bias-adjusted effect estimate (i.e., the size rule/array approach).^12^ Subsequent approaches that build upon these methods account for additional types of effect estimates,^13 14^ data types, ^15^ and assumptions.^16,17^ More recent QBA methods, including the E-value, require fewer assumptions and specifications, and allow for the estimation of the minimum strength of association, on the relative risk scale, that an unmeasured confounder would need to have with both the exposure and outcome to fully explain away observed findings.^18,19^ Since 2017, the E-value method has been extended to further minimize the number of required assumptions.^18,19^

Evidence suggests that many epidemiologic studies conducting bias analyses evaluate how strong a potential unmeasured confounder would need to be to fully explain away an observed non-null effect estimate.^5^ Unlike QBA methods that generate bias-adjusted effect estimates, which can be compared with the crude estimates from a study to determine the potential magnitude and direction of the bias,^4,5^ these methods may not always be as informative (e.g., when the magnitude of the effect estimate in a study is large or when the objective is to determine whether potential confounders are likely to change effect estimates by a specific amount).^5^ Given that many of the related methods for unmeasured confounding require relatively few assumptions and parameters, and have online tools to facilitate analyses,^18-21^ investigators should carefully consider the characteristics of their studies prior to determining whether it is more appropriate to measure the potential magnitude and direction of unmeasured confounding or describe how unmeasured confounding could explain away observed findings.

In our study, a third of the QBA methods for summary level data that we identified were for misclassification bias and fewer than 10% were for selection bias. Many QBA methods for misclassification fall under the matrix correction cluster of methods and can be used to adjust for the effects of nondifferential or differential misclassification using summary level data from contingency tables. The earliest approach that we identified was the matrix correction method from Barron (1977),^22^ which can be used to evaluate the effect of nondifferential misclassification in studies with categorical exposures and outcomes. Subsequent methods have extended this approach to accommodate differential misclassification bias, matched data, arbitrary two-way tables,^23^ and multilevel exposures.^24^ Given the availability of methods for misclassification bias, it may not be surprising that misclassification bias is modelled more often than selection bias in epidemiologic studies.^5^ Selection bias is often considered more challenging to understand than confounding or misclassification bias,^25^ with parameters that may not be as easy to identify.^5^ These findings highlight the need for greater guidance on the approaches, assumptions, required parameters, and interpretations of QBA methods for selection bias.

For this review, we generated detailed supplementary tables that outline any explicitly described assumptions, required bias parameters, formulas, and characteristics necessary to interpret the results from QBA methods. Previous studies have highlighted that one of the possible reasons for the limited application of the QBA methods in the epidemiologic literature is the fact that investigators may not be aware of the methods that are most straightforward to conduct.^5,9^ We hope that our review will help researchers identify methods that may be appropriate for their studies, including those with publicly available code or online tools. However, it is worth noting that not all assumptions and parameters are explicitly specified in the manuscripts describing the identified QBA methods. This is particularly concerning because it can lead to the misuse and misinterpretation of QBA analyses. Moving forward, it is crucial that manuscripts describing QBA methods clearly outline their assumptions and required parameters. Furthermore, anyone considering using QBA methods, including those identified by this review, should carefully review the original manuscripts to ensure the approach is appropriate given the study characteristics, that any assumptions are fulfilled, that the necessary bias parameters are available and accurate, and that interpretations and conclusions are made with caution.^9^

This study has a few limitations. First, the terminology used to describe various biases and bias analysis methods is largely unstandardized, which can make it difficult to identify articles developing QBA methods. However, we conducted a comprehensive search, with broad concepts across multiple databases, and performed reference chaining. Second, we restricted our search for QBA methods to the peer-reviewed literature, which did not include QBA methods described in preprints, conferences abstracts, working papers, dissertations, or textbooks. Third, we did not include QBA methods that can only be conducted using individual participant level data. Fourth, we relied on the information explicitly described in the manuscripts for each QBA method. However, it is possible that QBA methods could be extended to accommodate different study designs and data formats that are not described in the articles. Therefore, the information reported in the supplementary tables describing the QBA methods could change based on more comprehensive statistical evaluations. Fifth, our review does not provide information outlining the most appropriate QBA method for specific scenarios. Authors selecting QBA methods should carefully consider the stated and unstated assumptions, the feasibility of identifying required parameters, and the actual performance of the methods.

## Conclusion

In this systematic review, we identified a total of 57 QBA methods for summary level epidemiologic data published in the peer-reviewed literature. Future investigators can use this review to identify potential QBA methods that could be evaluated and then used for different study designs and biases. However, appropriate interpretation and implementation of these methods is necessary.

## Data Availability

All data produced in the present work are contained in the manuscript.

## Author contributions

JDW, ZLiew, and XS initiated the study. XS and ZLiu screened the manuscripts. XS abstracted and analyzed the data, which was verified by ZLiu, ZLiew, and JDW. All authors interpreted the results. XS wrote the first draft of the manuscript. All authors revised the manuscript and read and approved the final version of the manuscript. JDW supervised the research. XS and JDW had full access to all the data (including statistical reports and tables) in the study and can take responsibility for the integrity of the data and the accuracy of the analysis. JDW is the guarantor. XS and JDW have checked the references for accuracy and completeness.

## References

1. Hariton E, Locascio JJ. Randomised controlled trials - the gold standard for effectiveness research: Study design: randomised controlled trials. BJOG. 2018;125(13):1716.

2. Concato J, Corrigan-Curay J. Real-World Evidence — Where Are We Now? N. Engl. J. Med. 2022;386(18):1680–1682.

3. Lash TL, Fox MP, Cooney D, Lu Y, Forshee RA. Quantitative Bias Analysis in Regulatory Settings. Am. J. Public Health. 2016;106(7):1227–1230.

4. Fox MP, MacLehose RF, Lash TL. Applying Quantitative Bias Analysis to Epidemiologic Data. Springer Nature; 2021.

5. Petersen JM, Ranker LR, Barnard-Mayers R, MacLehose RF, Fox MP. A systematic review of quantitative bias analysis applied to epidemiological research. Int. J. Epidemiol. 2021.

6. Lash TL, Fox MP, MacLehose RF, Maldonado G, McCandless LC, Greenland S. Good practices for quantitative bias analysis. Int. J. Epidemiol. 2014;43(6):1969–1985.

7. Orsini N, Bellocco R, Bottai M, Wolk A, Greenland S. A Tool for Deterministic and Probabilistic Sensitivity Analysis of Epidemiologic Studies. The Stata Journal. 2008;8(1):29–48.

8. Smith LH, Mathur MB, VanderWeele TJ. Multiple-bias Sensitivity Analysis Using Bounds. Epidemiology. 2021;32(5):625–634.

9. Lash TL, Fox MP, MacLehose RF, Maldonado G, McCandless LC, Greenland S. Good practices for quantitative bias analysis. Int. J. Epidemiol. 2014;43(6):1969–1985.

10. Page MJ, McKenzie JE, Bossuyt PM, et al. The PRISMA 2020 statement: an updated guideline for reporting systematic reviews. BMJ. 2021;372:n71.

11. Cornfield J, Haenszel W, Hammond EC, Lilienfeld AM, Shimkin MB, Wynder EL. Smoking and Lung Cancer: Recent Evidence and a Discussion of Some Questions. JNCI: Journal of the National Cancer Institute. 1959;22(1):173–203.

12. Bross ID. Spurious effects from an extraneous variable. J. Chronic Dis. 1966;19(6):637–647.

13. Arah OA, Chiba Y, Greenland S. Bias Formulas for External Adjustment and Sensitivity Analysis of Unmeasured Confounders. Ann. Epidemiol. 2008;18(8):637–646.

14. Vanderweele TJ, Arah OA. Bias formulas for sensitivity analysis of unmeasured confounding for general outcomes, treatments, and confounders. Epidemiology. 2011;22(1):42–52.

15. Lin DY, Psaty BM, Kronmal RA. Assessing the sensitivity of regression results to unmeasured confounders in observational studies. Biometrics. 1998;54(3):948–963.

16. Schlesselman JJ. Assessing effects of confounding variables. Am. J. Epidemiol. 1978;108(1):3–8.

17. Rosenbaum PR, Rubin DB. Assessing Sensitivity to an Unobserved Binary Covariate in an Observational Study with Binary Outcome. Journal of the Royal Statistical Society. Series B (Methodological). 1983;45(2):212–218.

18. VanderWeele TJ, Ding P. Sensitivity Analysis in Observational Research: Introducing the E-Value. Ann. Intern. Med. 2017;167(4):268–274.

19. Ding P, VanderWeele TJ. Sensitivity Analysis Without Assumptions. Epidemiology. 2016;27(3):368–377.

20. MacLehose RF, Ahern TP, Lash TL, Poole C, Greenland S. The Importance of Making Assumptions in Bias Analysis. Epidemiology. 2021;32(5).

21. Cusson A, Infante-Rivard C. Bias factor, maximum bias and the E-value: insight and extended applications. Int. J. Epidemiol. 2020;49(5):1509–1516.

22. Barron BA. The effects of misclassification on the estimation of relative risk. Biometrics. 1977;33(2):414–418.

23. Greenland S, Kleinbaum DG. Correcting for Misclassification in Two-Way Tables and Matched-Pair Studies. Int. J. Epidemiol. 1983;12(1):93–97.

24. Weinkam JJ, Rosenbaum WL, Sterling TD. Recovering true risks when multilevel exposure and covariables are both misclassified. Am. J. Epidemiol. 1999;150(8):886–891.

25. Infante-Rivard C, Cusson A. Reflection on modern methods: selection bias—a review of recent developments. Int. J. Epidemiol. 2018;47(5):1714–1722.

