## Appendix 2 for "Quantitative bias analysis methods for summary level epidemiologic data in the peer-reviewed literature: a systematic review"

**Detailed search characteristics**

| **Ovid MEDLINE(R) ALL <1946 to December 10, 2021>** | | **#studies retrieved** |
| --- | --- | --- |
| 1 | quantitative bias analys#s.mp. | 83 |
| 2 | ("probabilistic* bias" or "probabilistic* analys#s").mp. | 769 |
| 3 | ("simple bias" or "simple sensitivity").mp. | 71 |
| 4 | ("multidimensional bias analys#s" or "multidimensional analys#s").mp. | 679 |
| 5 | multiple bias model*.mp. | 4 |
| 6 | (("sensitivity analys#s" or "quantitative analys#s") adj2 ("bias*" or "error*")).mp. | 587 |
| 7 | ("bias analys#s" or "uncertainty analys#s" or "sensitivity analys#s" or "sensitivity evaluation" or "quantitative analys#s").ti. and (bias*.ab. /freq=2 or (Confounding Factors, Epidemiologic/ or *Epidemiologic Methods/)) | 253 |
| 8 | (("bias analys#s" or "uncertainty analys#s" or "sensitivity analys#s" or "sensitivity evaluation") adj10 (confound* or systematic error* or quanti*)).ab. | 1384 |
| 9 | (("bias analys#s" or "uncertainty analys#s" or "sensitivity analys#s" or "sensitivity evaluation") and (confound* or systematic error* or quanti*)).ti. | 143 |
| 10 | ("selection bias" or "uncontrolled confound*" or "unmeasured confound*" or "misclassification" or "information bias" or "loss to followup" or "loss to follow up" or "lost to followup" or "lost to follow up" or "participation bias*" or "attrition bias*" or "measurement error" or "residual confound*").ti. and (bias*.ab. /freq=2 or (Confounding Factors, Epidemiologic/ or *Epidemiologic Methods/)) | 1083 |
| 11 | 1 or 2 or 3 or 4 or 5 or 6 or 7 or 8 or 9 or 10 | 4623 |

| **Embase <1974 to 2021 December 10>** | |  |
| --- | --- | --- |
| 1 | quantitative bias analys#s.mp. | 105 |
| 2 | ("probabilistic* bias" or "probabilistic* analys#s").mp. | 1108 |
| 3 | ("simple bias" or "simple sensitivity").mp. | 71 |
| 4 | ("multidimensional bias analys#s" or "multidimensional analys#s").mp. | 817 |
| 5 | multiple bias model*.mp. | 7 |
| 6 | (("sensitivity analys#s" or "quantitative analys#s") adj2 ("bias*" or "error*")).mp. | 692 |
| 7 | ("bias analys#s" or "uncertainty analys#s" or "sensitivity analys#s" or "sensitivity evaluation" or "quantitative analys#s").ti. and (bias*.ab. /freq=2 or *sensitivity analysis/) | 1161 |
| 8 | (("bias analys#s" or "uncertainty analys#s" or "sensitivity analys#s" or "sensitivity evaluation") adj10 (confound* or systematic error* or quanti*)).ab. | 1791 |
| 9 | (("bias analys#s" or "uncertainty analys#s" or "sensitivity analys#s" or "sensitivity evaluation") and (confound* or systematic error* or quanti*)).ti. | 166 |
| 10 | ("selection bias" or "uncontrolled confound*" or "unmeasured confound*" or "misclassification" or "information bias" or "loss to followup" or "loss to follow up" or "lost to followup" or "lost to follow up" or "participation bias*" or "attrition bias*" or "measurement error").ti. and (bias*.ab. /freq=2 or *sensitivity analysis/) | 1040 |
| 11 | 1 or 2 or 3 or 4 or 5 or 6 or 7 or 8 or 9 or 10 | 6410 |

**Web Of Science***

| **#studies retrieved** | |
| --- | --- |
| [1,932](https://www.webofscience.com/wos/woscc/summary/9a2d1713-7818-4796-8dfc-d2f905f0ec9f-20149eaa/relevance/1) | TI=("selection bias" or "uncontrolled confound*" or "unmeasured confound*" or "misclassification" or "information bias" or "loss to followup" or "loss to follow up" or "lost to followup" or "lost to follow up" or "participation bias*" or "attrition bias*" or "measurement error" or "residual confound*") and AB=(bias*) |
| [425](https://www.webofscience.com/wos/woscc/summary/719651b4-6c2c-49ba-b7a9-0fb5b8a0f976-20149a85/relevance/1) | TI=(("bias analys?s" or "uncertainty analys?s" or "sensitivity analys?s" or "sensitivity evaluation") and (confound* or "systematic error*" or quanti*)) |
| [3,515](https://www.webofscience.com/wos/woscc/summary/77e6f5b7-3dc6-4fd2-9062-6cc90218326e-201497c7/relevance/1) | AB=(("bias analys?s" or "uncertainty analys?s" or "sensitivity analys?s" or "sensitivity evaluation") NEAR/10 (confound* or "systematic error*" or quanti*)) |
| 913 | TI=("bias analys?s" or "uncertainty analys?s" or "sensitivity analys?s" or "sensitivity evaluation" or "quantitative analys?s") AND AB= (bias*) |
| 10692 | TS=("quantitative bias analys?s" OR "probabilistic* bias" OR "probabilistic* analys?s" OR "simple bias" or "simple sensitivity" OR "multidimensional bias analys?s" OR "multidimensional analys?s" OR "multiple bias model*" OR ( ("sensitivity analys?s" OR "quantitative analys?s") NEAR/2 (bias* OR error*) ) ) |
| 16888 | #1 OR #2 OR #3 OR #4 OR #5 |

*Web of Science Core Collection as licensed by Yale University (Science Citation Index Expanded 1900-present, Social Sciences Citation Index 1900-present, Arts & Humanities Citation Index 1975-present, Conference Proceedings Citation Index – Science 1991-present, Conference Proceedings Citation Index – Social Science & Humanities 1991-present, Book Citation Index – Science 2005-present, Book Citation Index – Social Sciences & Humanities 2005-present, Emerging Sources Citation Index 2018-present, Current Chemical Reactions 1985-present, Index Chemicus 1993-present).

**Scopus**

| #studies retrieved | |
| --- | --- |
| 22230 | ( TITLE-ABS-KEY ( "quantitative bias analys?s"  OR  "probabilistic* bias"  OR  "probabilistic* analys?s"  OR  "simple bias"  OR  "simple sensitivity"  OR  "multidimensional bias analys?s"  OR  "multidimensional analys?s"  OR  "multiple bias model*"  OR  ( ( "sensitivity analys?s"  OR  "quantitative analys?s" )  W/2  ( bias*  OR  error* ) ) ) )  OR  ( TITLE ( "bias analys?s"  OR  "uncertainty analys?s"  OR  "sensitivity analys?s"  OR  "sensitivity evaluation"  OR  "quantitative analys?s" )  AND  ABS ( bias* ) )  OR  ( ABS ( ( "bias analys?s"  OR  "uncertainty analys?s"  OR  "sensitivity analys?s"  OR  "sensitivity evaluation" )  W/10  ( confound*  OR  "systematic error*"  OR  quanti* ) ) )  OR  ( TITLE ( ( "bias analys?s"  OR  "uncertainty analys?s"  OR  "sensitivity analys?s"  OR  "sensitivity evaluation" )  AND  ( confound*  OR  "systematic error*"  OR  quanti* ) ) )  OR  ( TITLE ( "selection bias"  OR  "uncontrolled confound*"  OR  "unmeasured confound*"  OR  "misclassification"  OR  "information bias"  OR  "loss to followup"  OR  "loss to follow up"  OR  "lost to followup"  OR  "lost to follow up"  OR  "participation bias*"  OR  "attrition bias*"  OR  "measurement error"  OR  "residual confound*" )  AND  ABS ( bias* ) ) |
| Notes | For Web of science and Scopus, we excluded papers from the following areas: polymer science, spectroscopy, communication, information science library science, physics fluids plasmas, zoology, criminology, physics nuclear, forestry, physics particles fields, fisheries, veterinary sciences, chemistry applied, agricultural engineering, meterological atmosphere sciences, engineering chemical, marine freshwater bioogy, biodiversity conservation, robotics, regional urban planning, soil science, computer science cubernetics, medical legal, urban studies, biotechmology applied, biochemical research methods, geographic phsyical, engineering petroleun, language inguistics, physics condensed matter, dentistry oral, agriculture, ethics, geology, logic, parasitology, materials, international relations, law, medical la tech, antro, industrial relations, education, paleontology, public admin, history philosophy of science, material science ceramics, philosophy, demography |

**Besides database searching, we retrieved additional records by backwards reference chaining using Citationchaser,^1^ a tool for transparent and efficient citation chasing in systematic searching. This tool makes use of the Lens.org bibliographic database, which is a comprehensive database that** **collates content across multiple bibliographic resources.**

**Supplementary Table 1. Confounding methods derived from Cornfield (1959) and Bross (1966)**

| **Method** | **Name year** | **Brief descriptions and relationships with other methods** |
| --- | --- | --- |
| Sensitivity analysis for unmeasured confounding | Cornfield et al 1959^2^ | This paper, which is often referred to as describing the first sensitivity analysis, demonstrates a way to assess the impact of uncontrolled confounding when evaluating the association between smoking and lung cancer using data from two-by-two tables. |
| Size rule/Array approach for unmeasured confounding | Bross 1966^3^ | This paper, which initiated a framework for analyzing the bias due to an unmeasured confounder, provides formulas to quantify the effect of only one binary unmeasured confounder. |
| Method for unmeasured confounding | Schlesselman 1978^4^ | This paper, which builds upon Bross 1966, provides a formula that can be used for studies that do not fulfill the no interaction between confounder and exposure assumption. When the no interaction assumption is met, the formula is equivalent to Cornfield et al 1959 and Bross 1966. |
| Method for unmeasured confounding | Rosenbaum and Rubin 1983^5^ | This paper provides a formula to obtain bias-adjusted estimates accounting for measured and unmeasured confounding. While the method does not require the assumption for no interaction between unmeasured confounder and exposure, it is restricted to studies evaluating binary outcomes. |
| Indirect assessment of confounding | Flanders and Khoury 1990^6^ | This paper provides a formula to obtain bias-adjusted estimates when little is known about the unmeasured confounder. |
| Basic methods for sensitivity analysis of unmeasured confounding | Greenland 1996^7^ | This paper describes a method to account for unmeasured confounding that can be implemented with computer programming. This method has the assumption that the odds ratios or relative risks between the exposure and the outcome are constant across strata of the unmeasured confounder. |
| Sensitivity analysis for unmeasured confounders for regression results | Lin et al 1998^8^ | This paper describes a method that builds upon Schlesselman 1978^4^ and can be used for regression model results. While the method can be applied to different types of outcomes (binary, continuous, or survival time), it requires conditional independence of unmeasured confounders and measured confounders given exposure, and no interaction between exposure and unmeasured confounding. |
| Method for unmeasured confounders | Arah et al 2008^9^ | This paper describes a method that builds upon the framework developed by Schlesselman 1978^4^ and Flanders and Khoury 1990^6^, and can be applied to studies estimating relative risks, odds ratios, or risk differences. The formula for relative risk is equivalent to the formula derived by Flanders and Khoury 1990.^6^ |
| Method for unmeasured confounding for general outcomes, treatments, and confounders | VanderWeele and Arah 2011^10^ | This paper describes a method that builds upon Rosenbaum and Rubin 1983,^5^ and can accommodate categorical or continuous exposures, unmeasured confounders, and outcomes, and works for additive, risk-ratio, and odds-ratio scales. It also does not assume that the unmeasured confounders are independent from the measured confounders. |
| Bayesian method for unmeasured confounding in meta-analyses | McCandless 2012^11^ | This paper describes a Bayesian method for meta-analyses that builds upon the framework and formula from Lin et al 1998.^8^ |
| Bounding factor | Ding and VanderWeele 2016^12^ | This paper describes a method that generates bounding factors, which can be considered a measure of the strength of confounding between the exposure and the outcome induced by a confounder. This method requires no assumptions about the unmeasured confound and was further developed into the E-value method. |
| The E-value method | VanderWeele and Ding 2017^13^ | The paper describes the E-value method which focuses on estimating the minimum strength of association, on the relative risk scale, that an unmeasured confounder would need to have with both the exposure and outcome to fully explain away a specific exposure-outcome relationship. It has only one explicitly stated assumption (i.e., that the association between unmeasured confounder and exposure is of the same strength as the association between confounder and outcome) and can be applied to different exposure and outcome types. |
| Robustness value | Cinelli and Hazlett 2019^14^ | This paper describes a method that is similar to the E-value method that can be used for linear regression models. It requires equal associations between the confounder and the exposure, and the confounder and the outcome. |
| E-value extensions | Cusson and Infante-Rivard 2020^15^ | This paper extends the E-value method to accommodate more general situations. First, it does not require the equal association between unmeasured confounder and exposure, and the association between confounder and outcome. Second, although the E-value method could be applied to studies with odds ratios, the measure was only interpretable on the relative risk scale. Therefore, Cusson and Infante-Rivard extend the E-value formula so that it can also be interpreted on the odds ratio scale. |
| Generalized E-value (G-value) | MacLehose et al 2021^16^ | This paper extends the E-value method by removing the unstated assumption that the prevalence of the uncontrolled confounder among the exposed is 100%. The G-value requires one more parameter that the E-value - prevalence of confounder among the exposed. However, when this prevalence is 100%, the E-value and G-value are identical for the same exposure-outcome association. |

**Supplementary Table 2. Matrix correction method for misclassification bias**

| **Method** | **Name year** | **Brief descriptions and relationships with other methods** |
| --- | --- | --- |
| Method for misclassification for relative risks | Barron 1977^17^ | This paper describes a matrix formula that allows for the correction of nondifferential misclassification using summary data from two-by-two tables. While this method can be applied to nondifferential exposure and/or outcome misclassification, it is restricted to categorical (binary or nonbinary) exposures and outcomes. |
| Method for misclassification bias in the estimation of relative risk | Copeland et al 1977^18^ | This paper describes a correction formula for differential or nondifferential misclassification of study subjects (i.e., disease occurrence in cohorts or exposure history in case-control studies,) using summary data from two-by-two tables. It is restricted to only binary exposures and outcomes. |
| Generalizations of Barron’s matrix correction method | Greenland and Kleinbaum 1983^19^ | This paper, which builds upon Barron 1977^17^, provides a methods that can accommodate studies with differential misclassification, matched-pair data, and arbitrary two-way tables. While this method can be applied to nondifferential and differential exposure and/or outcome misclassification on the estimation of odds ratios, it is restricted to categorical (binary or nonbinary) exposures and outcomes. |
| Sensitivity analysis for validation studies using an alloyed gold standard | Wacholder et al 1993^20^ | This paper, which upon Barron 1977,^17^ explores the influence of the error in an alloyed gold standard when a validation study is used to correct estimates of effect. This method only works for nondifferential misclassification bias and is restricted to a categorical (binary or nonbinary) exposure and a binary outcome. |
| Alloyed gold standard for exposure misclassification | Brenner 1996^21^ | This paper describes a method that can be used to correct for exposure misclassification using an alloyed gold standard, which was proposed by Wacholder et al 1993^20^. Unlike Wacholder et al 1993, this method is restricted to a binary exposure and a categorical outcome. It also requires that a validation study is carried out in which routinely employed measures of a binary exposure variable are validated by an alloyed gold standard in a group of study participants. |
| Sensitivity analysis when multilevel exposure and covariates are both misclassified | Weinkam et al 1999^22^ | The paper, which builds upon Barron 1977^17^, can be used for studies with multilevel exposures and covariables are both subject to nondifferential misclassification with respect to a binary outcome variable known without error. This method is restricted to studies with categorical exposures (binary or nonbinary), confounders (binary or nonbinary), and outcomes (binary only) |

References

1.	Haddaway NR, Grainger MJ, Gray CT. Citationchaser: A tool for transparent and efficient forward and backward citation chasing in systematic searching. *Research Synthesis Methods.* 2022;13(4):533-545.

2.	Cornfield J, Haenszel W, Hammond EC, Lilienfeld AM, Shimkin MB, Wynder EL. Smoking and Lung Cancer: Recent Evidence and a Discussion of Some Questions. *JNCI: Journal of the National Cancer Institute.* 1959;22(1):173-203.

3.	Bross ID. Spurious effects from an extraneous variable. *J. Chronic Dis.* 1966;19(6):637-647.

4.	Schlesselman JJ. Assessing effects of confounding variables. *Am. J. Epidemiol.* 1978;108(1):3-8.

5.	Rosenbaum PR, Rubin DB. Assessing Sensitivity to an Unobserved Binary Covariate in an Observational Study with Binary Outcome. *Journal of the Royal Statistical Society. Series B (Methodological).* 1983;45(2):212-218.

6.	Flanders WD, Khoury MJ. Indirect assessment of confounding: graphic description and limits on effect of adjusting for covariates. *Epidemiology.* 1990;1(3):239-246.

7.	GREENLAND S. Basic Methods for Sensitivity Analysis of Biases. *Int. J. Epidemiol.* 1996;25(6):1107-1116.

8.	Lin DY, Psaty BM, Kronmal RA. Assessing the sensitivity of regression results to unmeasured confounders in observational studies. *Biometrics.* 1998;54(3):948-963.

9.	Arah OA, Chiba Y, Greenland S. Bias Formulas for External Adjustment and Sensitivity Analysis of Unmeasured Confounders. *Ann. Epidemiol.* 2008;18(8):637-646.

10.	Vanderweele TJ, Arah OA. Bias formulas for sensitivity analysis of unmeasured confounding for general outcomes, treatments, and confounders. *Epidemiology.* 2011;22(1):42-52.

11.	McCandless LC. Meta-analysis of observational studies with unmeasured confounders. *Int. J. Biostat.* 2012;8(2).

12.	Ding P, VanderWeele TJ. Sensitivity Analysis Without Assumptions. *Epidemiology.* 2016;27(3):368-377.

13.	VanderWeele TJ, Ding P. Sensitivity Analysis in Observational Research: Introducing the E-Value. *Ann. Intern. Med.* 2017;167(4):268-274.

14.	Cinelli C, Hazlett C. Making sense of sensitivity: Extending omitted variable bias. *Journal of the Royal Statistical Society: Series B (Statistical Methodology).* 2020;82(1):39-67.

15.	Cusson A, Infante-Rivard C. Bias factor, maximum bias and the E-value: insight and extended applications. *Int. J. Epidemiol.* 2020;49(5):1509-1516.

16.	MacLehose RF, Ahern TP, Lash TL, Poole C, Greenland S. The Importance of Making Assumptions in Bias Analysis. *Epidemiology.* 2021;32(5).

17.	Barron BA. The effects of misclassification on the estimation of relative risk. *Biometrics.* 1977;33(2):414-418.

18.	Copeland KT, Checkoway H, McMichael AJ, Holbrook RH. Bias due to misclassification in the estimation of relative risk. *Am. J. Epidemiol.* 1977;105(5):488-495.

19.	GREENLAND S, KLEINBAUM DG. Correcting for Misclassification in Two-Way Tables and Matched-Pair Studies. *Int. J. Epidemiol.* 1983;12(1):93-97.

20.	Wacholder S, Armstrong B, Hartge P. Validation Studies using an Alloyed Gold Standard. *Am. J. Epidemiol.* 1993;137(11):1251-1258.

21.	Brenner H. Correcting for exposure misclassification using an alloyed gold standard. *Epidemiology.* 1996;7(4):406-410.

22.	Weinkam JJ, Rosenbaum WL, Sterling TD. Recovering true risks when multilevel exposure and covariables are both misclassified. *Am. J. Epidemiol.* 1999;150(8):886-891.
