## Appendix 3 for "Quantitative bias analysis methods for summary level epidemiologic data in the peer-reviewed literature: a systematic review"

**eAppendix 3. Classification of quantitative bias analysis methods across key study characteristics**

**Abbreviations**

CI: confidence interval;

DOI: digital object identifier;

HR: hazard ratio;

MD: mean difference;

OR: odds ratio;

PMID: the PubMed reference number;

QBA: quantitative bias analysis;

RD: risk difference;

RR: risk ratio;

SMR: standardized mortality ratio.

Contents of QBA methods for summary level data

**Classification tool for** **QBA methods for summary level data (*indicates availability for software or online tools)**

**1. Study design (55)**

**1.1 Non-randomized interventional studies (2)**

***2. Bias type***

***2.1 Unmeasured confounding (0)***

***2.2 Misclassification bias (2)***

Method for misclassification bias in cohort studies of vaccine efficacy (Farrington 1990), this method can be used to quantify the bias due to misclassification and generate bias-adjusted estimates in prospective cohort studies of vaccine efficacy. Analyses can be conducted using multiple parameters, including the predictive value and sensitivities among the vaccinated group.

Response probability ratio for missing outcome (Magder 2003), this method can be used to assess the impact of missing outcome information among experimental or observational study designs. Analyses can be conducted using the following information: the cells in a 2 by 2 table and the probability that a patient will not have a missing value for the outcome variable for two treatments.

***2.3 Selection bias (0)***

**1.2 Observational studies (53)**

**1.2.1 Cohort studies (39)**

***2. Bias type***

***2.1 Unmeasured confounding (23)***

***2.1.1 Multiple unmeasured confounders (8)***

3. Result of interest

3.1 Explain-away (6)

4. Confounder data format

4.1 Categorical (6)

5. Exposure data format

5.1 Categorical (6)

6. Outcome data format

6.1 Categorical (5)

Impact thresholds for regression models (Frank 2000), this method can be used to quantify the impact of a confounding variable on the inference of a regression coefficient. Analyses can be conducted using the following information: the sample correlation between the outcome and predictor of interest, the estimated learn regression coefficient and standard error, the sample size, and the number of parameters estimate in the regression model (note: the approach can be extended beyond the general linear model (e.g., logistic regression), but the information provided is for linear models);

Bounding formulas for unmeasured confounding with effect-modifying potentials (Lee 2011), this method can be used to assess if the unmeasured confounder that may exhibit effect modification can explain away the findings. Analyses can be conducted using the following information: the confounded effect estimate, the association between an unmeasured confounder and outcome, the association between an unmeasured confounder and exposure, and the modifying effect that an unmeasured confounder has on the association between an exposure and outcome;

E-value (VanderWeele and Ding 2017)*, this method can be used to assess the minimum strength of potential unmeasured confounding needed to explain away an observed effect estimate. this method can be conducted using only reported effect estimates, and explicitly requires that the association between confounder and exposure is of the same strength as the association between confounder and outcome and that the prevalence of the uncontrolled confound among the exposed is 100%;

E-value extensions (Cusson and Infante-Rivard 2020), this method can be used as an extension of the E-value method to accommodate more general situations, including studies where it is not possible to assume that the association between unmeasured confounder and exposure and the association between confounder and outcome are equal. Analyses can be conducted using the observed relative risk or odds ratio;

Generalized E-value (G-value) (MacLehose et al 2021), this method can be used to assess the minimum strength of potential unmeasured confounding needed to explain away an effect, and extends the E-value method by removing the assumption that the prevalence of the uncontrolled confounder among the exposed is 100%. Analyses can be conducted using the following information: the effect estimate and the prevalence of the confounder among the exposed.

6.2 Continuous (3)

Impact thresholds for regression models (Frank 2000), this method can be used to quantify the impact of a confounding variable on the inference of a regression coefficient. Analyses can be conducted using the following information: the sample correlation between the outcome and predictor of interest, the estimated learn regression coefficient and standard error, the sample size, and the number of parameters estimate in the regression model (note: the approach can be extended beyond the general linear model (e.g., logistic regression), but the information provided is for linear models);

E-value (VanderWeele and Ding 2017)*, this method can be used to assess the minimum strength of potential unmeasured confounding needed to explain away an observed effect estimate. this method can be conducted using only reported effect estimates, and explicitly requires that the association between confounder and exposure is of the same strength as the association between confounder and outcome and that the prevalence of the uncontrolled confound among the exposed is 100%;

Robustness value (Cinelli and Hazlett 2019)*, this method can be used as an extension of the E-value method for regression models. Analyses can be conducted using the following information: the coefficient estimate, standard error, and degrees of freedom from a linear regression model.

6.3 Time-to-event (3)

E-value (VanderWeele and Ding 2017)*, this method can be used to assess the minimum strength of potential unmeasured confounding needed to explain away an observed effect estimate. this method can be conducted using only reported effect estimates, and explicitly requires that the association between confounder and exposure is of the same strength as the association between confounder and outcome and that the prevalence of the uncontrolled confound among the exposed is 100%;

E-value extensions (Cusson and Infante-Rivard 2020), this method can be used as an extension of the E-value method to accommodate more general situations, including studies where it is not possible to assume that the association between unmeasured confounder and exposure and the association between confounder and outcome are equal. Analyses can be conducted using the observed relative risk or odds ratio;

Generalized E-value (G-value) (MacLehose et al 2021), this method can be used to assess the minimum strength of potential unmeasured confounding needed to explain away an effect, and extends the E-value method by removing the assumption that the prevalence of the uncontrolled confounder among the exposed is 100%. Analyses can be conducted using the following information: the effect estimate and the prevalence of the confounder among the exposed.

5.2 Continuous (2)

Impact thresholds for regression models (Frank 2000), t his method can be used to quantify the impact of a confounding variable on the inference of a regression coefficient. Analyses can be conducted using the following information: the sample correlation between the outcome and predictor of interest, the estimated learn regression coefficient and standard error, the sample size, and the number of parameters estimate in the regression model (note: the approach can be extended beyond the general linear model (e.g., logistic regression), but the information provided is for linear models);

Robustness value (Cinelli and Hazlett 2019)*, this method can be used as an extension of the E-value method for regression models. Analyses can be conducted using the following information: the coefficient estimate, standard error, and degrees of freedom from a linear regression model.

4.2 Categorical (non-binary) (6)

5. Exposure data format

5.1 Categorical (6)

6. Outcome data format

6.1 Categorical (5)

Impact thresholds for regression models (Frank 2000), this method can be used to quantify the impact of a confounding variable on the inference of a regression coefficient. Analyses can be conducted using the following information: the sample correlation between the outcome and predictor of interest, the estimated learn regression coefficient and standard error, the sample size, and the number of parameters estimate in the regression model (note: the approach can be extended beyond the general linear model (e.g., logistic regression), but the information provided is for linear models);

Bounding formulas for unmeasured confounding with effect-modifying potentials (Lee 2011), this method can be used to assess if the unmeasured confounder that may exhibit effect modification can explain away the findings. Analyses can be conducted using the following information: the confounded effect estimate, the association between an unmeasured confounder and outcome, the association between an unmeasured confounder and exposure, and the modifying effect that an unmeasured confounder has on the association between an exposure and outcome;

E-value (VanderWeele and Ding 2017)*, this method can be used to assess the minimum strength of potential unmeasured confounding needed to explain away an observed effect estimate. this method can be conducted using only reported effect estimates, and explicitly requires that the association between confounder and exposure is of the same strength as the association between confounder and outcome and that the prevalence of the uncontrolled confound among the exposed is 100%;

E-value extensions (Cusson and Infante-Rivard 2020), this method can be used as an extension of the E-value method to accommodate more general situations, including studies where it is not possible to assume that the association between unmeasured confounder and exposure and the association between confounder and outcome are equal. Analyses can be conducted using the observed relative risk or odds ratio;

Generalized E-value (G-value) (MacLehose et al 2021), this method can be used to assess the minimum strength of potential unmeasured confounding needed to explain away an effect, and extends the E-value method by removing the assumption that the prevalence of the uncontrolled confounder among the exposed is 100%. Analyses can be conducted using the following information: the effect estimate and the prevalence of the confounder among the exposed.

6.2 Continuous (3)

Impact thresholds for regression models (Frank 2000), this method can be used to quantify the impact of a confounding variable on the inference of a regression coefficient. Analyses can be conducted using the following information: the sample correlation between the outcome and predictor of interest, the estimated learn regression coefficient and standard error, the sample size, and the number of parameters estimate in the regression model (note: the approach can be extended beyond the general linear model (e.g., logistic regression), but the information provided is for linear models);

E-value (VanderWeele and Ding 2017)*, this method can be used to assess the minimum strength of potential unmeasured confounding needed to explain away an observed effect estimate. this method can be conducted using only reported effect estimates, and explicitly requires that the association between confounder and exposure is of the same strength as the association between confounder and outcome and that the prevalence of the uncontrolled confound among the exposed is 100%;

Robustness value (Cinelli and Hazlett 2019)*, this method can be used as an extension of the E-value method for regression models. Analyses can be conducted using the following information: the coefficient estimate, standard error, and degrees of freedom from a linear regression model.

6.3 Time-to-event (3)

E-value (VanderWeele and Ding 2017)*, this method can be used to assess the minimum strength of potential unmeasured confounding needed to explain away an observed effect estimate. this method can be conducted using only reported effect estimates, and explicitly requires that the association between confounder and exposure is of the same strength as the association between confounder and outcome and that the prevalence of the uncontrolled confound among the exposed is 100%;

E-value extensions (Cusson and Infante-Rivard 2020), this method can be used as an extension of the E-value method to accommodate more general situations, including studies where it is not possible to assume that the association between unmeasured confounder and exposure and the association between confounder and outcome are equal. Analyses can be conducted using the observed relative risk or odds ratio;

Generalized E-value (G-value) (MacLehose et al 2021), this method can be used to assess the minimum strength of potential unmeasured confounding needed to explain away an effect, and extends the E-value method by removing the assumption that the prevalence of the uncontrolled confounder among the exposed is 100%. Analyses can be conducted using the following information: the effect estimate and the prevalence of the confounder among the exposed.

5.2 Continuous (2)

Impact thresholds for regression models (Frank 2000), this method can be used to quantify the impact of a confounding variable on the inference of a regression coefficient. Analyses can be conducted using the following information: the sample correlation between the outcome and predictor of interest, the estimated learn regression coefficient and standard error, the sample size, and the number of parameters estimate in the regression model (note: the approach can be extended beyond the general linear model (e.g., logistic regression), but the information provided is for linear models);

Robustness value (Cinelli and Hazlett 2019)*, this method can be used as an extension of the E-value method for regression models. Analyses can be conducted using the following information: the coefficient estimate, standard error, and degrees of freedom from a linear regression model.

4.3 Continuous (4)

5. Exposure data format

5.1 Categorical (4)

6. Outcome data format

6.1 Categorical (3)

Impact thresholds for regression models (Frank 2000), this method can be used to quantify the impact of a confounding variable on the inference of a regression coefficient. Analyses can be conducted using the following information: the sample correlation between the outcome and predictor of interest, the estimated learn regression coefficient and standard error, the sample size, and the number of parameters estimate in the regression model (note: the approach can be extended beyond the general linear model (e.g., logistic regression), but the information provided is for linear models);

Bounding formulas for unmeasured confounding with effect-modifying potentials (Lee 2011), this method can be used to assess if the unmeasured confounder that may exhibit effect modification can explain away the findings. Analyses can be conducted using the following information: the confounded effect estimate, the association between an unmeasured confounder and outcome, the association between an unmeasured confounder and exposure, and the modifying effect that an unmeasured confounder has on the association between an exposure and outcome;

E-value (VanderWeele and Ding 2017)*, this method can be used to assess the minimum strength of potential unmeasured confounding needed to explain away an observed effect estimate. this method can be conducted using only reported effect estimates, and explicitly requires that the association between confounder and exposure is of the same strength as the association between confounder and outcome and that the prevalence of the uncontrolled confound among the exposed is 100%.

6.2 Continuous (2)

Impact thresholds for regression models (Frank 2000), this method can be used to quantify the impact of a confounding variable on the inference of a regression coefficient. Analyses can be conducted using the following information: the sample correlation between the outcome and predictor of interest, the estimated learn regression coefficient and standard error, the sample size, and the number of parameters estimate in the regression model (note: the approach can be extended beyond the general linear model (e.g., logistic regression), but the information provided is for linear models);

Robustness value (Cinelli and Hazlett 2019)*, this method can be used as an extension of the E-value method for regression models. Analyses can be conducted using the following information: the coefficient estimate, standard error, and degrees of freedom from a linear regression model.

6.3 Time-to-event (1)

E-value (VanderWeele and Ding 2017)*, this method can be used to assess the minimum strength of potential unmeasured confounding needed to explain away an observed effect estimate. this method can be conducted using only reported effect estimates, and explicitly requires that the association between confounder and exposure is of the same strength as the association between confounder and outcome and that the prevalence of the uncontrolled confound among the exposed is 100%.

5.2 Continuous (3)

Impact thresholds for regression models (Frank 2000), this method can be used to quantify the impact of a confounding variable on the inference of a regression coefficient. Analyses can be conducted using the following information: the sample correlation between the outcome and predictor of interest, the estimated learn regression coefficient and standard error, the sample size, and the number of parameters estimate in the regression model (note: the approach can be extended beyond the general linear model (e.g., logistic regression), but the information provided is for linear models);

E-value (VanderWeele and Ding 2017)*, this method can be used to assess the minimum strength of potential unmeasured confounding needed to explain away an observed effect estimate. this method can be conducted using only reported effect estimates, and explicitly requires that the association between confounder and exposure is of the same strength as the association between confounder and outcome and that the prevalence of the uncontrolled confound among the exposed is 100%;

Robustness value (Cinelli and Hazlett 2019)*, this method can be used as an extension of the E-value method for regression models. Analyses can be conducted using the following information: the coefficient estimate, standard error, and degrees of freedom from a linear regression model.

3.2 Corrected estimates (2)

Sensitivity analysis for unmeasured confounders for regression results (Lin et al 1998), this method can be used to assess the sensitivity of regression results to unmeasured confounders by generating bias-adjusted estimates. Analyses can be conducted using the following information: the association between the unmeasured confounder and outcome among the unexposed and exposed, the prevalence of the unmeasured confounder among the unexposed and exposed, and the confounded effect estimate;

Sensitivity analysis for interactions under unmeasured confounding (VanderWeele et al 2012), this method can be used to assess the sensitivity of interaction analyses to unmeasured confounding by generating bias-adjusted estimates. Depending on the type of interaction (additive or multiplicative), analyses can be conducted using different parameters, including the prevalence of the unmeasured confounder.

***2.1.2 Single unmeasured confounder (23)***

3. Result of interest

3.1 Explain-away (9)

4. Confounder data format

4.1 Categorical (9)

5. Exposure data format

5.1 Categorical (9)

6. Outcome data format

6.1 Categorical (8)

Sensitivity analysis for unmeasured confounding (Cornfield et al 1959), this method can be used to understand the impact of unmeasured confounding played on the exposure-outcome relationship. Analyses can be conducted using the following information: the prevalence of the confounder among the exposed, relative to the prevalence among those unexposed, and crude effect estimate between the exposure and outcome;

Basic methods for sensitivity analysis of unmeasured confounding (Greenland 1996), this method can be used to generate effect estimates that are corrected for potential unmeasured confounders. Analyses can be conducted using the following information: the information reported in 2 by 2 tables, the effect estimate between an unmeasured confounder and outcome, and the prevalence of unmeasured confounder among the unexposed and the exposed;

Impact thresholds for regression models (Frank 2000), this method can be used to quantify the impact of a confounding variable on the inference of a regression coefficient. Analyses can be conducted using the following information: the sample correlation between the outcome and predictor of interest, the estimated learn regression coefficient and standard error, the sample size, and the number of parameters estimate in the regression model (note: the approach can be extended beyond the general linear model (e.g., logistic regression), but the information provided is for linear models);

Rule-out approach/target-adjustment sensitivity analysis (Schneeweiss 2006), this method can be used to understand if the observed exposure-outcome association can be explained away by the unmeasured confounder. Analyses can be conducted using the following information: the prevalence of the exposure and confounder in the total population, the confounded effect estimate, and the effect estimate between the confounder and outcome;

Bounding formulas for unmeasured confounding with effect-modifying potentials (Lee 2011), this method can be used to assess if the unmeasured confounder that may exhibit effect modification can explain away the findings. Analyses can be conducted using the following information: the confounded effect estimate, the association between an unmeasured confounder and outcome, the association between an unmeasured confounder and exposure, and the modifying effect that an unmeasured confounder has on the association between an exposure and outcome;

E-value (VanderWeele and Ding 2017)*, this method can be used to assess the minimum strength of potential unmeasured confounding needed to explain away an observed effect estimate. this method can be conducted using only reported effect estimates, and explicitly requires that the association between confounder and exposure is of the same strength as the association between confounder and outcome and that the prevalence of the uncontrolled confound among the exposed is 100%;

E-value extensions (Cusson and Infante-Rivard 2020), this method can be used as an extension of the E-value method to accommodate more general situations, including studies where it is not possible to assume that the association between unmeasured confounder and exposure and the association between confounder and outcome are equal. Analyses can be conducted using the observed relative risk or odds ratio;

Generalized E-value (G-value) (MacLehose et al 2021), this method can be used to assess the minimum strength of potential unmeasured confounding needed to explain away an effect, and extends the E-value method by removing the assumption that the prevalence of the uncontrolled confounder among the exposed is 100%. Analyses can be conducted using the following information: the effect estimate and the prevalence of the confounder among the exposed.

6.2 Continuous (3)

Impact thresholds for regression models (Frank 2000), this method can be used to quantify the impact of a confounding variable on the inference of a regression coefficient. Analyses can be conducted using the following information: the sample correlation between the outcome and predictor of interest, the estimated learn regression coefficient and standard error, the sample size, and the number of parameters estimate in the regression model (note: the approach can be extended beyond the general linear model (e.g., logistic regression), but the information provided is for linear models);

E-value (VanderWeele and Ding 2017)*, this method can be used to assess the minimum strength of potential unmeasured confounding needed to explain away an observed effect estimate. this method can be conducted using only reported effect estimates, and explicitly requires that the association between confounder and exposure is of the same strength as the association between confounder and outcome and that the prevalence of the uncontrolled confound among the exposed is 100%;

Robustness value (Cinelli and Hazlett 2019)*, this method can be used as an extension of the E-value method for regression models. Analyses can be conducted using the following information: the coefficient estimate, standard error, and degrees of freedom from a linear regression model.

6.3 Time-to-event (3)

E-value (VanderWeele and Ding 2017)*, this method can be used to assess the minimum strength of potential unmeasured confounding needed to explain away an observed effect estimate. this method can be conducted using only reported effect estimates, and explicitly requires that the association between confounder and exposure is of the same strength as the association between confounder and outcome and that the prevalence of the uncontrolled confound among the exposed is 100%;

E-value extensions (Cusson and Infante-Rivard 2020), this method can be used as an extension of the E-value method to accommodate more general situations, including studies where it is not possible to assume that the association between unmeasured confounder and exposure and the association between confounder and outcome are equal. Analyses can be conducted using the observed relative risk or odds ratio;

Generalized E-value (G-value) (MacLehose et al 2021), this method can be used to assess the minimum strength of potential unmeasured confounding needed to explain away an effect, and extends the E-value method by removing the assumption that the prevalence of the uncontrolled confounder among the exposed is 100%. Analyses can be conducted using the following information: the effect estimate and the prevalence of the confounder among the exposed.

5.2 Continuous (2)

Impact thresholds for regression models (Frank 2000), this method can be used to quantify the impact of a confounding variable on the inference of a regression coefficient. Analyses can be conducted using the following information: the sample correlation between the outcome and predictor of interest, the estimated learn regression coefficient and standard error, the sample size, and the number of parameters estimate in the regression model (note: the approach can be extended beyond the general linear model (e.g., logistic regression), but the information provided is for linear models);

Robustness value (Cinelli and Hazlett 2019)*, this method can be used as an extension of the E-value method for regression models. Analyses can be conducted using the following information: the coefficient estimate, standard error, and degrees of freedom from a linear regression model.

4.2 Categorical (non-binary) (6)

5. Exposure data format

5.1 Categorical (6)

6. Outcome data format

6.1 Categorical (5)

Impact thresholds for regression models (Frank 2000), this method can be used to quantify the impact of a confounding variable on the inference of a regression coefficient. Analyses can be conducted using the following information: the sample correlation between the outcome and predictor of interest, the estimated learn regression coefficient and standard error, the sample size, and the number of parameters estimate in the regression model (note: the approach can be extended beyond the general linear model (e.g., logistic regression), but the information provided is for linear models);

Bounding formulas for unmeasured confounding with effect-modifying potentials (Lee 2011), this method can be used to assess if the unmeasured confounder that may exhibit effect modification can explain away the findings. Analyses can be conducted using the following information: the confounded effect estimate, the association between an unmeasured confounder and outcome, the association between an unmeasured confounder and exposure, and the modifying effect that an unmeasured confounder has on the association between an exposure and outcome;

E-value (VanderWeele and Ding 2017)*, this method can be used to assess the minimum strength of potential unmeasured confounding needed to explain away an observed effect estimate. this method can be conducted using only reported effect estimates, and explicitly requires that the association between confounder and exposure is of the same strength as the association between confounder and outcome and that the prevalence of the uncontrolled confound among the exposed is 100%;

E-value extensions (Cusson and Infante-Rivard 2020), this method can be used as an extension of the E-value method to accommodate more general situations, including studies where it is not possible to assume that the association between unmeasured confounder and exposure and the association between confounder and outcome are equal. Analyses can be conducted using the observed relative risk or odds ratio;

Generalized E-value (G-value) (MacLehose et al 2021), this method can be used to assess the minimum strength of potential unmeasured confounding needed to explain away an effect, and extends the E-value method by removing the assumption that the prevalence of the uncontrolled confounder among the exposed is 100%. Analyses can be conducted using the following information: the effect estimate and the prevalence of the confounder among the exposed.

6.2 Continuous (3)

Impact thresholds for regression models (Frank 2000), this method can be used to quantify the impact of a confounding variable on the inference of a regression coefficient. Analyses can be conducted using the following information: the sample correlation between the outcome and predictor of interest, the estimated learn regression coefficient and standard error, the sample size, and the number of parameters estimate in the regression model (note: the approach can be extended beyond the general linear model (e.g., logistic regression), but the information provided is for linear models);

E-value (VanderWeele and Ding 2017)*, this method can be used to assess the minimum strength of potential unmeasured confounding needed to explain away an observed effect estimate. this method can be conducted using only reported effect estimates, and explicitly requires that the association between confounder and exposure is of the same strength as the association between confounder and outcome and that the prevalence of the uncontrolled confound among the exposed is 100%;

Robustness value (Cinelli and Hazlett 2019)*, this method can be used as an extension of the E-value method for regression models. Analyses can be conducted using the following information: the coefficient estimate, standard error, and degrees of freedom from a linear regression model.

6.3 Time-to-event (3)

E-value (VanderWeele and Ding 2017)*, this method can be used to assess the minimum strength of potential unmeasured confounding needed to explain away an observed effect estimate. this method can be conducted using only reported effect estimates, and explicitly requires that the association between confounder and exposure is of the same strength as the association between confounder and outcome and that the prevalence of the uncontrolled confound among the exposed is 100%;

E-value extensions (Cusson and Infante-Rivard 2020), this method can be used as an extension of the E-value method to accommodate more general situations, including studies where it is not possible to assume that the association between unmeasured confounder and exposure and the association between confounder and outcome are equal. Analyses can be conducted using the observed relative risk or odds ratio;

Generalized E-value (G-value) (MacLehose et al 2021), this method can be used to assess the minimum strength of potential unmeasured confounding needed to explain away an effect, and extends the E-value method by removing the assumption that the prevalence of the uncontrolled confounder among the exposed is 100%. Analyses can be conducted using the following information: the effect estimate and the prevalence of the confounder among the exposed.

5.2 Continuous (2)

Impact thresholds for regression models (Frank 2000), this method can be used to quantify the impact of a confounding variable on the inference of a regression coefficient. Analyses can be conducted using the following information: the sample correlation between the outcome and predictor of interest, the estimated learn regression coefficient and standard error, the sample size, and the number of parameters estimate in the regression model (note: the approach can be extended beyond the general linear model (e.g., logistic regression), but the information provided is for linear models);

Robustness value (Cinelli and Hazlett 2019)*, this method can be used as an extension of the E-value method for regression models. Analyses can be conducted using the following information: the coefficient estimate, standard error, and degrees of freedom from a linear regression model.

4.3 Continuous (4)

5. Exposure data format

5.1 Categorical (4)

6. Outcome data format

6.1 Categorical (3)

Impact thresholds for regression models (Frank 2000), this method can be used to quantify the impact of a confounding variable on the inference of a regression coefficient. Analyses can be conducted using the following information: the sample correlation between the outcome and predictor of interest, the estimated learn regression coefficient and standard error, the sample size, and the number of parameters estimate in the regression model (note: the approach can be extended beyond the general linear model (e.g., logistic regression), but the information provided is for linear models);

Bounding formulas for unmeasured confounding with effect-modifying potentials (Lee 2011), this method can be used to assess if the unmeasured confounder that may exhibit effect modification can explain away the findings. Analyses can be conducted using the following information: the confounded effect estimate, the association between an unmeasured confounder and outcome, the association between an unmeasured confounder and exposure, and the modifying effect that an unmeasured confounder has on the association between an exposure and outcome;

E-value (VanderWeele and Ding 2017)*, this method can be used to assess the minimum strength of potential unmeasured confounding needed to explain away an observed effect estimate. this method can be conducted using only reported effect estimates, and explicitly requires that the association between confounder and exposure is of the same strength as the association between confounder and outcome and that the prevalence of the uncontrolled confound among the exposed is 100%.

6.2 Continuous (3)

Impact thresholds for regression models (Frank 2000), this method can be used to quantify the impact of a confounding variable on the inference of a regression coefficient. Analyses can be conducted using the following information: the sample correlation between the outcome and predictor of interest, the estimated learn regression coefficient and standard error, the sample size, and the number of parameters estimate in the regression model (note: the approach can be extended beyond the general linear model (e.g., logistic regression), but the information provided is for linear models);

E-value (VanderWeele and Ding 2017)*, this method can be used to assess the minimum strength of potential unmeasured confounding needed to explain away an observed effect estimate. this method can be conducted using only reported effect estimates, and explicitly requires that the association between confounder and exposure is of the same strength as the association between confounder and outcome and that the prevalence of the uncontrolled confound among the exposed is 100%;

Robustness value (Cinelli and Hazlett 2019)*, this method can be used as an extension of the E-value method for regression models. Analyses can be conducted using the following information: the coefficient estimate, standard error, and degrees of freedom from a linear regression model.

6.3 Time-to-event (1)

E-value (VanderWeele and Ding 2017)*, this method can be used to assess the minimum strength of potential unmeasured confounding needed to explain away an observed effect estimate. this method can be conducted using only reported effect estimates, and explicitly requires that the association between confounder and exposure is of the same strength as the association between confounder and outcome and that the prevalence of the uncontrolled confound among the exposed is 100%.

5.2 Continuous (2)

Impact thresholds for regression models (Frank 2000), this method can be used to quantify the impact of a confounding variable on the inference of a regression coefficient. Analyses can be conducted using the following information: the sample correlation between the outcome and predictor of interest, the estimated learn regression coefficient and standard error, the sample size, and the number of parameters estimate in the regression model (note: the approach can be extended beyond the general linear model (e.g., logistic regression), but the information provided is for linear models);

Robustness value (Cinelli and Hazlett 2019)*, this method can be used as an extension of the E-value method for regression models. Analyses can be conducted using the following information: the coefficient estimate, standard error, and degrees of freedom from a linear regression model.

3.2 Corrected estimates (14)

4. Confounder data format

4.1 Categorical (14)

5. Exposure data format

5.1 Categorical (14)

6. Outcome data format

6.1 Categorical (14)

Size rule/array approach for unmeasured confounding (Bross 1966)*, this method can be used to generate bias-adjusted estimates corrected for an unmeasured confounder. Analyses can be conducted using the following information: the prevalence of a confounder in the exposed and the unexposed, the confounded effect estimate, and the association between a confounder and an outcome;

Method for unmeasured confounding (Schlesselman 1978), this method can be used to generate bias-adjusted estimates corrected for an unmeasured confounder. Analyses can be conducted using the following information: the prevalence of a confounder in the exposed, the prevalence of a confounder in the unexposed, and the confounded effect estimate;

Method for unmeasured confounding (Rosenbaum and Rubin 1983), this method can be used to generate bias-adjusted estimates corrected for an unmeasured confounder. Analyses can be conducted using the following information: the proportion of the unexposed, the prevalence of the unmeasured confounder, the association between the unmeasured confounder and the outcome among the unexposed and exposed, and the association between the unmeasured confounder and the exposure;

Indirect assessment of confounding (Flanders and Khoury 1990), this method can be used to generate bias-adjusted estimates when little is known about the unmeasured confounder. Analyses can be conducted using the following information: only one or two of the following parameters - the prevalence of the confounder among the unexposed in the overall population, the association between the exposure and the confounder, the effect of the confounder on disease, and the unadjusted association between the exposure and the outcome;

Sensitivity analysis for unmeasured confounders for regression results (Lin et al 1998), this method can be used to assess the sensitivity of regression results to unmeasured confounders by generating bias-adjusted estimates. Analyses can be conducted using the following information: the association between the unmeasured confounder and outcome among the unexposed and exposed, the prevalence of the unmeasured confounder among the unexposed and exposed, and the confounded effect estimate;

Quantifying biases for classical confounding and (or) collider-stratification bias (Greenland 2003), this method can be used to examine the potential impact of confounding with or without selection bias under simple causal models. Analyses can be conducted using the following information: the reported associations between exposure, outcome, confounder, and collider. For unmeasured confounding, the analyses also require the confounded effect estimate and the prevalence of the confounder among the unexposed controls;

Monte Carlo sensitivity analysis and Bayesian analysis for unmeasured confounding (Steenland and Greenland 2004)*, this method can be used to generate a range of effect measures given the likely range of bias using Monte Carlo sensitivity analysis and/or Bayesian analysis. Typically, researchers need to combine the priors for unknown parameters with the probability of the observed data, using Bayes’ theorem, to produce a posterior distribution for the parameter of interest (i.e., the bias-adjusted rate ratio);

Integration of Rosenbaum and Greenland method for confounding (Cabral 2007)*, this method, which integrates two previous sensitivity analyses methods, can be used to generate effect estimates that are corrected for potential unmeasured confounders. Analyses can be conducted using the following information: the cells from a 2 by 2 table and odds ratios between the exposure and the confounder, and the confounder and the outcome;

Method for unmeasured confounders (Arah et al 2008), this method can be used to generate effect estimates that are corrected for potential unmeasured confounders. Analyses can be conducted using the following information: the prevalence of the confounder among the exposed, unexposed, and total population, and the effect estimates for confounder-stratified exposure-outcome relationships in the total population;

Probabilistic bias analysis for two biases (Lash et al 2010)*, this method can be used to generate bias-adjusted estimates that are corrected for potential unmeasured confounders. Analyses can be conducted using the following information: the distribution of the association between the confounder and the outcome, and the distribution of prevalence of the confounder in the exposed and the unexposed;

Sensitivity analysis for unmeasured confounding of attributable fraction (Chiba 2011), this method can be used to generate bias-adjusted estimates corrected for unmeasured confounding when the effect measure of interest is attributable fraction. Analyses can be conducted using the following information: the confounded effect estimate, sensitivity parameter, average difference in potential outcomes between those exposed and those unexposed, disease prevalence, and exposure prevalence;

Method for unmeasured confounding for general outcomes, treatments, and confounders (VanderWeele and Arah 2011), this method can be used to control for the impact of unmeasured confounders under different situations, including studies with categorical or continuous exposures, unmeasured confounders, and outcomes, and effect estimates that are on the additive, risk-ratio, and odds-ratio scales. Analyses can be conducted using the following information: the associations between the unmeasured confounder and outcome and the distribution of the unmeasured confounder among different exposure levels;

Sensitivity analysis for interactions under unmeasured confounding (VanderWeele et al 2012), this method can be used to assess the sensitivity of interaction analyses to unmeasured confounding by generating bias-adjusted estimates. Depending on the type of interaction (additive or multiplicative), analyses can be conducted using different parameters, including the prevalence of the unmeasured confounder;

Indirect adjustment of relative risks of an exposure with multiple categories for an unmeasured confounder (Lubin et al 2018)*, this method can be used to conduct indirect adjustment of effect estimates for categorical unmeasured confounding. Analyses can be conducted using the following information: the reported associations between exposure and outcome among the exposed and unexposed, and the same effect estimates in individuals that have the unmeasured confounder among the exposed and unexposed.

6.2 Continuous (4)

Sensitivity analysis for unmeasured confounders for regression results (Lin et al 1998), this method can be used to assess the sensitivity of regression results to unmeasured confounders by generating bias-adjusted estimates. Analyses can be conducted using the following information: the association between the unmeasured confounder and outcome among the unexposed and exposed, the prevalence of the unmeasured confounder among the unexposed and exposed, and the confounded effect estimate;

Integration of Rosenbaum and Greenland method for confounding (Cabral 2007)*, this method, which integrates two previous sensitivity analyses methods, can be used to generate effect estimates that are corrected for potential unmeasured confounders. Analyses can be conducted using the following information: the cells from a 2 by 2 table and odds ratios between the exposure and the confounder, and the confounder and the outcome;

Method for unmeasured confounding for general outcomes, treatments, and confounders (VanderWeele and Arah 2011), this method can be used to control for the impact of unmeasured confounders under different situations, including studies with categorical or continuous exposures, unmeasured confounders, and outcomes, and effect estimates that are on the additive, risk-ratio, and odds-ratio scales. Analyses can be conducted using the following information: the associations between the unmeasured confounder and outcome and the distribution of the unmeasured confounder among different exposure levels;

Sensitivity analysis for interactions under unmeasured confounding (VanderWeele et al 2012), this method can be used to assess the sensitivity of interaction analyses to unmeasured confounding by generating bias-adjusted estimates. Depending on the type of interaction (additive or multiplicative), analyses can be conducted using different parameters, including the prevalence of the unmeasured confounder.

6.3 Time-to-event (3)

Sensitivity analysis for unmeasured confounders for regression results (Lin et al 1998), this method can be used to assess the sensitivity of regression results to unmeasured confounders by generating bias-adjusted estimates. Analyses can be conducted using the following information: the association between the unmeasured confounder and outcome among the unexposed and exposed, the prevalence of the unmeasured confounder among the unexposed and exposed, and the confounded effect estimate;

Probabilistic bias analysis for two biases (Lash et al 2010)*, this method can be used to generate bias-adjusted estimates that are corrected for potential unmeasured confounders. Analyses can be conducted using the following information: the distribution of the association between the confounder and the outcome, and the distribution of prevalence of the confounder in the exposed and the unexposed;

Indirect adjustment of relative risks of an exposure with multiple categories for an unmeasured confounder (Lubin et al 2018)*, this method can be used to conduct indirect adjustment of effect estimates for categorical unmeasured confounding. Analyses can be conducted using the following information: the reported associations between exposure and outcome among the exposed and unexposed, and the same effect estimates in individuals that have the unmeasured confounder among the exposed and unexposed.

5.2 Continuous (3)

6. Outcome data format

6.1 Categorical (3)

Integration of Rosenbaum and Greenland method for confounding (Cabral 2007)*, this method, which integrates two previous sensitivity analyses methods, can be used to generate effect estimates that are corrected for potential unmeasured confounders. Analyses can be conducted using the following information: the cells from a 2 by 2 table and odds ratios between the exposure and the confounder, and the confounder and the outcome;

Method for unmeasured confounding for general outcomes, treatments, and confounders (VanderWeele and Arah 2011), this method can be used to control for the impact of unmeasured confounders under different situations, including studies with categorical or continuous exposures, unmeasured confounders, and outcomes, and effect estimates that are on the additive, risk-ratio, and odds-ratio scales. Analyses can be conducted using the following information: the associations between the unmeasured confounder and outcome and the distribution of the unmeasured confounder among different exposure levels;

Sensitivity analysis for interactions under unmeasured confounding (VanderWeele et al 2012), this method can be used to assess the sensitivity of interaction analyses to unmeasured confounding by generating bias-adjusted estimates. Depending on the type of interaction (additive or multiplicative), analyses can be conducted using different parameters, including the prevalence of the unmeasured confounder.

6.2 Continuous (3)

Integration of Rosenbaum and Greenland method for confounding (Cabral 2007)*, this method, which integrates two previous sensitivity analyses methods, can be used to generate effect estimates that are corrected for potential unmeasured confounders. Analyses can be conducted using the following information: the cells from a 2 by 2 table and odds ratios between the exposure and the confounder, and the confounder and the outcome;

Method for unmeasured confounding for general outcomes, treatments, and confounders (VanderWeele and Arah 2011), this method can be used to control for the impact of unmeasured confounders under different situations, including studies with categorical or continuous exposures, unmeasured confounders, and outcomes, and effect estimates that are on the additive, risk-ratio, and odds-ratio scales. Analyses can be conducted using the following information: the associations between the unmeasured confounder and outcome and the distribution of the unmeasured confounder among different exposure levels;

Sensitivity analysis for interactions under unmeasured confounding (VanderWeele et al 2012), this method can be used to assess the sensitivity of interaction analyses to unmeasured confounding by generating bias-adjusted estimates. Depending on the type of interaction (additive or multiplicative), analyses can be conducted using different parameters, including the prevalence of the unmeasured confounder.

6.3 Time-to-event (0)

4.2 Categorical (non-binary) (9)

5. Exposure data format

5.1 Categorical (9)

6. Outcome data format

6.1 Categorical (9)

Indirect assessment of confounding (Flanders and Khoury 1990), this method can be used to generate bias-adjusted estimates when little is known about the unmeasured confounder. Analyses can be conducted using the following information: only one or two of the following parameters - the prevalence of the confounder among the unexposed in the overall population, the association between the exposure and the confounder, the effect of the confounder on disease, and the unadjusted association between the exposure and the outcome;

Sensitivity analysis for unmeasured confounders for regression results (Lin et al 1998), this method can be used to assess the sensitivity of regression results to unmeasured confounders by generating bias-adjusted estimates. Analyses can be conducted using the following information: the association between the unmeasured confounder and outcome among the unexposed and exposed, the prevalence of the unmeasured confounder among the unexposed and exposed, and the confounded effect estimate;

Monte Carlo sensitivity analysis and Bayesian analysis for unmeasured confounding (Steenland and Greenland 2004)*, this method can be used to generate a range of effect measures given the likely range of bias using Monte Carlo sensitivity analysis and/or Bayesian analysis. Typically, researchers need to combine the priors for unknown parameters with the probability of the observed data, using Bayes’ theorem, to produce a posterior distribution for the parameter of interest (i.e., the bias-adjusted rate ratio);

Method for unmeasured confounders (Arah et al 2008), this method can be used to generate effect estimates that are corrected for potential unmeasured confounders. Analyses can be conducted using the following information: the prevalence of the confounder among the exposed, unexposed, and total population, and the effect estimates for confounder-stratified exposure-outcome relationships in the total population;

Probabilistic bias analysis for two biases (Lash et al 2010)*, this method can be used to generate bias-adjusted estimates that are corrected for potential unmeasured confounders. Analyses can be conducted using the following information: the distribution of the association between the confounder and the outcome, and the distribution of prevalence of the confounder in the exposed and the unexposed;

Sensitivity analysis for unmeasured confounding of attributable fraction (Chiba 2011), this method can be used to generate bias-adjusted estimates corrected for unmeasured confounding when the effect measure of interest is attributable fraction. Analyses can be conducted using the following information: the confounded effect estimate, sensitivity parameter, average difference in potential outcomes between those exposed and those unexposed, disease prevalence, and exposure prevalence;

Method for unmeasured confounding for general outcomes, treatments, and confounders (VanderWeele and Arah 2011), this method can be used to control for the impact of unmeasured confounders under different situations, including studies with categorical or continuous exposures, unmeasured confounders, and outcomes, and effect estimates that are on the additive, risk-ratio, and odds-ratio scales. Analyses can be conducted using the following information: the associations between the unmeasured confounder and outcome and the distribution of the unmeasured confounder among different exposure levels;

Sensitivity analysis for interactions under unmeasured confounding (VanderWeele et al 2012), this method can be used to assess the sensitivity of interaction analyses to unmeasured confounding by generating bias-adjusted estimates. Depending on the type of interaction (additive or multiplicative), analyses can be conducted using different parameters, including the prevalence of the unmeasured confounder;

Indirect adjustment of relative risks of an exposure with multiple categories for an unmeasured confounder (Lubin et al 2018)*, this method can be used to conduct indirect adjustment of effect estimates for categorical unmeasured confounding. Analyses can be conducted using the following information: the reported associations between exposure and outcome among the exposed and unexposed, and the same effect estimates in individuals that have the unmeasured confounder among the exposed and unexposed.

6.2 Continuous (3)

Integration of Rosenbaum and Greenland method for confounding (Cabral 2007)*, this method, which integrates two previous sensitivity analyses methods, can be used to generate effect estimates that are corrected for potential unmeasured confounders. Analyses can be conducted using the following information: the cells from a 2 by 2 table and odds ratios between the exposure and the confounder, and the confounder and the outcome;

Method for unmeasured confounding for general outcomes, treatments, and confounders (VanderWeele and Arah 2011), this method can be used to control for the impact of unmeasured confounders under different situations, including studies with categorical or continuous exposures, unmeasured confounders, and outcomes, and effect estimates that are on the additive, risk-ratio, and odds-ratio scales. Analyses can be conducted using the following information: the associations between the unmeasured confounder and outcome and the distribution of the unmeasured confounder among different exposure levels;

Sensitivity analysis for interactions under unmeasured confounding (VanderWeele et al 2012), this method can be used to assess the sensitivity of interaction analyses to unmeasured confounding by generating bias-adjusted estimates. Depending on the type of interaction (additive or multiplicative), analyses can be conducted using different parameters, including the prevalence of the unmeasured confounder.

6.3 Time-to-event (3)

Sensitivity analysis for unmeasured confounders for regression results (Lin et al 1998), this method can be used to assess the sensitivity of regression results to unmeasured confounders by generating bias-adjusted estimates. Analyses can be conducted using the following information: the association between the unmeasured confounder and outcome among the unexposed and exposed, the prevalence of the unmeasured confounder among the unexposed and exposed, and the confounded effect estimate;

Probabilistic bias analysis for two biases (Lash et al 2010)*, this method can be used to generate bias-adjusted estimates that are corrected for potential unmeasured confounders. Analyses can be conducted using the following information: the distribution of the association between the confounder and the outcome, and the distribution of prevalence of the confounder in the exposed and the unexposed;

Indirect adjustment of relative risks of an exposure with multiple categories for an unmeasured confounder (Lubin et al 2018)*, this method can be used to conduct indirect adjustment of effect estimates for categorical unmeasured confounding. Analyses can be conducted using the following information: the reported associations between exposure and outcome among the exposed and unexposed, and the same effect estimates in individuals that have the unmeasured confounder among the exposed and unexposed.

5.2 Continuous (2)

Method for unmeasured confounding for general outcomes, treatments, and confounders (VanderWeele and Arah 2011), this method can be used to control for the impact of unmeasured confounders under different situations, including studies with categorical or continuous exposures, unmeasured confounders, and outcomes, and effect estimates that are on the additive, risk-ratio, and odds-ratio scales. Analyses can be conducted using the following information: the associations between the unmeasured confounder and outcome and the distribution of the unmeasured confounder among different exposure levels;

Sensitivity analysis for interactions under unmeasured confounding (VanderWeele et al 2012), this method can be used to assess the sensitivity of interaction analyses to unmeasured confounding by generating bias-adjusted estimates. Depending on the type of interaction (additive or multiplicative), analyses can be conducted using different parameters, including the prevalence of the unmeasured confounder.

4.3 Continuous (3)

5. Exposure data format

5.1 Categorical (3)

6. Outcome data format

6.1 Categorical (3)

Sensitivity analysis for unmeasured confounders for regression results (Lin et al 1998), this method can be used to assess the sensitivity of regression results to unmeasured confounders by generating bias-adjusted estimates. Analyses can be conducted using the following information: the association between the unmeasured confounder and outcome among the unexposed and exposed, the prevalence of the unmeasured confounder among the unexposed and exposed, and the confounded effect estimate;

Sensitivity analysis for unmeasured confounding of attributable fraction (Chiba 2011), this method can be used to generate bias-adjusted estimates corrected for unmeasured confounding when the effect measure of interest is attributable fraction. Analyses can be conducted using the following information: the confounded effect estimate, sensitivity parameter, average difference in potential outcomes between those exposed and those unexposed, disease prevalence, and exposure prevalence;

Method for unmeasured confounding for general outcomes, treatments, and confounders (VanderWeele and Arah 2011), this method can be used to control for the impact of unmeasured confounders under different situations, including studies with categorical or continuous exposures, unmeasured confounders, and outcomes, and effect estimates that are on the additive, risk-ratio, and odds-ratio scales. Analyses can be conducted using the following information: the associations between the unmeasured confounder and outcome and the distribution of the unmeasured confounder among different exposure levels.

6.2 Continuous (2)

Sensitivity analysis for unmeasured confounders for regression results (Lin et al 1998), this method can be used to assess the sensitivity of regression results to unmeasured confounders by generating bias-adjusted estimates. Analyses can be conducted using the following information: the association between the unmeasured confounder and outcome among the unexposed and exposed, the prevalence of the unmeasured confounder among the unexposed and exposed, and the confounded effect estimate;

Method for unmeasured confounding for general outcomes, treatments, and confounders (VanderWeele and Arah 2011), this method can be used to control for the impact of unmeasured confounders under different situations, including studies with categorical or continuous exposures, unmeasured confounders, and outcomes, and effect estimates that are on the additive, risk-ratio, and odds-ratio scales. Analyses can be conducted using the following information: the associations between the unmeasured confounder and outcome and the distribution of the unmeasured confounder among different exposure levels.

6.3 Time-to-event (1)

Sensitivity analysis for unmeasured confounders for regression results (Lin et al 1998), this method can be used to assess the sensitivity of regression results to unmeasured confounders by generating bias-adjusted estimates. Analyses can be conducted using the following information: the association between the unmeasured confounder and outcome among the unexposed and exposed, the prevalence of the unmeasured confounder among the unexposed and exposed, and the confounded effect estimate.

5.2 Continuous (1)

Method for unmeasured confounding for general outcomes, treatments, and confounders (VanderWeele and Arah 2011), this method can be used to control for the impact of unmeasured confounders under different situations, including studies with categorical or continuous exposures, unmeasured confounders, and outcomes, and effect estimates that are on the additive, risk-ratio, and odds-ratio scales. Analyses can be conducted using the following information: the associations between the unmeasured confounder and outcome and the distribution of the unmeasured confounder among different exposure levels.

***2.2 Misclassification bias (8)***

***2.2.1 Exposure misclassification bias (6)***

3. Misclassification type

3.1 Differential exposure misclassification bias (2)

Method for misclassification bias in the estimation of relative risk (Copeland et al 1977), this method can be used to correct for differential or nondifferential misclassification of study subjects (i.e., disease occurrence in cohorts or exposure history in case-control studies,) in 2 by 2 tables. Analyses can be conducted using the following information: the four cells from 2 by 2 tables and estimates of specificity of the classification procedure;

Method for differential measurement error (VanderWeele and Li 2019), this method can be used to assess how strong differential measurement error would have to be to completely explain away an observed exposure-outcome association. Analyses can be conducted using the following information: the observed and potential true risk ratio between exposure and outcome.

3.2 Nondifferential exposure misclassification bias (5)

4. Exposure data type

4.1 Categorical (5)

Method for misclassification bias in the estimation of relative risk (Copeland et al 1977), this method can be used to correct for differential or nondifferential misclassification of study subjects (i.e., disease occurrence in cohorts or exposure history in case-control studies,) in 2 by 2 tables. Analyses can be conducted using the following information: the four cells from 2 by 2 tables and estimates of specificity of the classification procedure;

Method for nondifferential exposure misclassification (Checkoway et al 1991), this method can be used to assess the effects of nondifferential misclassification of exposures in occupational studies. Analyses can be conducted using the following information: the four misclassified cells in a 2 by 2 table, sensitivity, and specificity;

Sensitivity analysis for validation studies using an alloyed gold standard (Wacholder et al 1993), this method can be used to explore the influence of misclassification bias in an alloyed gold standard when a validation study is used to correct effect estimates. Analyses can be conducted using the following information: the coefficient estimate of the regression of the outcome on the measured exposure, and the correction factor from the validation study with the alloyed gold standard;

Sensitivity analysis when multilevel exposure and covariates are both misclassified (Weinkam 1999), this method can be used to recover the true relative risks from estimates biased by misclassification in which multilevel exposure and covariables are both subject to nondifferential misclassification. Analyses can be conducted using the following information: the effect estimates for the exposure and confounder in different strata by the exposure and confounder/effect modifier level;

Method for exposure misclassification bias in vaccine effectiveness (Baum et al 2021), this method can be used to assess biases under non-differential exposure misclassification when estimating vaccine effectiveness. Analyses can be conducted using various reported parameters, including the risk of the disease in those observed as vaccinated and unvaccinated, and the sensitivity and specificity of the vaccine.

4.2 Continuous (1)

Sensitivity analysis for validation studies using an alloyed gold standard (Wacholder et al 1993), this method can be used to explore the influence of misclassification bias in an alloyed gold standard when a validation study is used to correct effect estimates. Analyses can be conducted using the following information: the coefficient estimate of the regression of the outcome on the measured exposure, and the correction factor from the validation study with the alloyed gold standard.

***2.2.2 Outcome misclassification bias (1)***

Probabilistic bias analysis for two biases (Lash et al 2010), this method can be used to generate effect estimates that are corrected for bias from outcome misclassification. Analyses can be conducted using the following information: the number of observed cases and the distribution of sensitivities of the exposed and unexposed.

***2.2.3 Confounder misclassification bias (1)***

Bias analysis for a misclassified confounder (Nab et al 2020), this method can be used to assess the bias due to confounder misclassification in analyses that aim to estimate the average treatment effect using a marginal structural model estimated using inverse probability weighting. Analyses can be conducted using multiple parameters, including prevalence of the observed confounder.

***2.3 Selection bias (4)***

3. Result of interest

3.1 Explain-away (2)

Bounding formulas for m-bias and other structural selection bias (Flanders and Ye 2019)*, this method can be used to estimate the bound for M-bias, a structural selection bias. Analyses can be conducted using the associations between all the variables in the causal pathway for M bias;

E-value for selection bias (Smith and VanderWeele 2019, explain-away)*, this method can be used to assess the minimum strength of potential selection bias needed to explain away an observed effect. Analyses can be conducted using the crude effect estimate and corresponding 95% CIs.

3.2 Corrected estimates (2)

Quantifying biases for classical confounding and (or) collider-stratification bias (Greenland 2003), this method can be used to examine the relative magnitudes of selection biases under some simple causal models in which the stratification variable is graphically depicted as a collider. Analyses can be conducted using the following information: the reported associations between exposure, outcome, confounder, and collider;

E-value for selection bias (Smith and VanderWeele 2019, corrected estimates)*, this method can be used to generate selection bias-adjusted effect estimates. Analyses can be conducted using the following information: the reported crude effect estimate and various parameters related to the selection process.

***2.4 Multiple biases (2)***

3. Result of interest

3.1 Explain-away (1)

Bounding formulas for selection bias (Huang and Lee 2015), this method, which extends previous bounding formulas for unmeasured confounding and effect modification, can be used to assess if selection bias can explain away an observed finding. Analyses can be conducted using the associations between all variables, including the exposure, outcome, variable indicating the selection of study subjects, and unmeasured confounder.

3.2 Corrected estimates (1)

Bounding method for multiple biases (Smith et al 2021), this method can be used to estimate bounds for the total composite bias (selection bias, unmeasured confounding, and misclassification bias) under a variety of scenarios. Analyses can be conducted using different parameters related to the biases of interest.

**1.2.2 Case-control studies (43)**

***2. Bias type***

***2.1 Unmeasured confounding (30)***

***2.1.1 Multiple unmeasured confounders (8)***

3. Result of interest

3.1 Explain-away (6)

4. Confounder data format

4.1 Categorical (6)

5. Exposure data format

5.1 Categorical (6)

6. Outcome data format

6.1 Categorical (5)

Impact thresholds for regression models (Frank 2000), this method can be used to quantify the impact of a confounding variable on the inference of a regression coefficient. Analyses can be conducted using the following information: the sample correlation between the outcome and predictor of interest, the estimated learn regression coefficient and standard error, the sample size, and the number of parameters estimate in the regression model (note: the approach can be extended beyond the general linear model (e.g., logistic regression), but the information provided is for linear models);

Bounding formulas for unmeasured confounding with effect-modifying potentials (Lee 2011), this method can be used to assess if the unmeasured confounder that may exhibit effect modification can explain away the findings. Analyses can be conducted using the following information: the confounded effect estimate, the association between an unmeasured confounder and outcome, the association between an unmeasured confounder and exposure, and the modifying effect that an unmeasured confounder has on the association between an exposure and outcome;

E-value (VanderWeele and Ding 2017)*, this method can be used to assess the minimum strength of potential unmeasured confounding needed to explain away an observed effect estimate. this method can be conducted using only reported effect estimates, and explicitly requires that the association between confounder and exposure is of the same strength as the association between confounder and outcome and that the prevalence of the uncontrolled confound among the exposed is 100%;

E-value extensions (Cusson and Infante-Rivard 2020), this method can be used as an extension of the E-value method to accommodate more general situations, including studies where it is not possible to assume that the association between unmeasured confounder and exposure and the association between confounder and outcome are equal. Analyses can be conducted using the observed relative risk or odds ratio;

Generalized E-value (G-value) (MacLehose et al 2021), this method can be used to assess the minimum strength of potential unmeasured confounding needed to explain away an effect, and extends the E-value method by removing the assumption that the prevalence of the uncontrolled confounder among the exposed is 100%. Analyses can be conducted using the following information: the effect estimate and the prevalence of the confounder among the exposed.

6.2 Continuous (3)

Impact thresholds for regression models (Frank 2000), this method can be used to quantify the impact of a confounding variable on the inference of a regression coefficient. Analyses can be conducted using the following information: the sample correlation between the outcome and predictor of interest, the estimated learn regression coefficient and standard error, the sample size, and the number of parameters estimate in the regression model (note: the approach can be extended beyond the general linear model (e.g., logistic regression), but the information provided is for linear models);

E-value (VanderWeele and Ding 2017)*, this method can be used to assess the minimum strength of potential unmeasured confounding needed to explain away an observed effect estimate. this method can be conducted using only reported effect estimates, and explicitly requires that the association between confounder and exposure is of the same strength as the association between confounder and outcome and that the prevalence of the uncontrolled confound among the exposed is 100%;

Robustness value (Cinelli and Hazlett 2019)*, this method can be used as an extension of the E-value method for regression models. Analyses can be conducted using the following information: the coefficient estimate, standard error, and degrees of freedom from a linear regression model.

6.3 Time-to-event (3)

E-value (VanderWeele and Ding 2017)*, this method can be used to assess the minimum strength of potential unmeasured confounding needed to explain away an observed effect estimate. this method can be conducted using only reported effect estimates, and explicitly requires that the association between confounder and exposure is of the same strength as the association between confounder and outcome and that the prevalence of the uncontrolled confound among the exposed is 100%;

E-value extensions (Cusson and Infante-Rivard 2020), this method can be used as an extension of the E-value method to accommodate more general situations, including studies where it is not possible to assume that the association between unmeasured confounder and exposure and the association between confounder and outcome are equal. Analyses can be conducted using the observed relative risk or odds ratio;

Generalized E-value (G-value) (MacLehose et al 2021), this method can be used to assess the minimum strength of potential unmeasured confounding needed to explain away an effect, and extends the E-value method by removing the assumption that the prevalence of the uncontrolled confounder among the exposed is 100%. Analyses can be conducted using the following information: the effect estimate and the prevalence of the confounder among the exposed.

5.2 Continuous (2)

Impact thresholds for regression models (Frank 2000), this method can be used to quantify the impact of a confounding variable on the inference of a regression coefficient. Analyses can be conducted using the following information: the sample correlation between the outcome and predictor of interest, the estimated learn regression coefficient and standard error, the sample size, and the number of parameters estimate in the regression model (note: the approach can be extended beyond the general linear model (e.g., logistic regression), but the information provided is for linear models);

Robustness value (Cinelli and Hazlett 2019)*, this method can be used as an extension of the E-value method for regression models. Analyses can be conducted using the following information: the coefficient estimate, standard error, and degrees of freedom from a linear regression model.

4.2 Categorical (non-binary) (6)

5. Exposure data format

5.1 Categorical (6)

6. Outcome data format

6.1 Categorical (5)

Impact thresholds for regression models (Frank 2000), this method can be used to quantify the impact of a confounding variable on the inference of a regression coefficient. Analyses can be conducted using the following information: the sample correlation between the outcome and predictor of interest, the estimated learn regression coefficient and standard error, the sample size, and the number of parameters estimate in the regression model (note: the approach can be extended beyond the general linear model (e.g., logistic regression), but the information provided is for linear models);

Bounding formulas for unmeasured confounding with effect-modifying potentials (Lee 2011), this method can be used to assess if the unmeasured confounder that may exhibit effect modification can explain away the findings. Analyses can be conducted using the following information: the confounded effect estimate, the association between an unmeasured confounder and outcome, the association between an unmeasured confounder and exposure, and the modifying effect that an unmeasured confounder has on the association between an exposure and outcome;

E-value (VanderWeele and Ding 2017)*, this method can be used to assess the minimum strength of potential unmeasured confounding needed to explain away an observed effect estimate. this method can be conducted using only reported effect estimates, and explicitly requires that the association between confounder and exposure is of the same strength as the association between confounder and outcome and that the prevalence of the uncontrolled confound among the exposed is 100%;

E-value extensions (Cusson and Infante-Rivard 2020), this method can be used as an extension of the E-value method to accommodate more general situations, including studies where it is not possible to assume that the association between unmeasured confounder and exposure and the association between confounder and outcome are equal. Analyses can be conducted using the observed relative risk or odds ratio;

Generalized E-value (G-value) (MacLehose et al 2021), this method can be used to assess the minimum strength of potential unmeasured confounding needed to explain away an effect, and extends the E-value method by removing the assumption that the prevalence of the uncontrolled confounder among the exposed is 100%. Analyses can be conducted using the following information: the effect estimate and the prevalence of the confounder among the exposed.

6.2 Continuous (3)

Impact thresholds for regression models (Frank 2000), this method can be used to quantify the impact of a confounding variable on the inference of a regression coefficient. Analyses can be conducted using the following information: the sample correlation between the outcome and predictor of interest, the estimated learn regression coefficient and standard error, the sample size, and the number of parameters estimate in the regression model (note: the approach can be extended beyond the general linear model (e.g., logistic regression), but the information provided is for linear models);

E-value (VanderWeele and Ding 2017)*, this method can be used to assess the minimum strength of potential unmeasured confounding needed to explain away an observed effect estimate. this method can be conducted using only reported effect estimates, and explicitly requires that the association between confounder and exposure is of the same strength as the association between confounder and outcome and that the prevalence of the uncontrolled confound among the exposed is 100%;

Robustness value (Cinelli and Hazlett 2019)*, this method can be used as an extension of the E-value method for regression models. Analyses can be conducted using the following information: the coefficient estimate, standard error, and degrees of freedom from a linear regression model.

6.3 Time-to-event (3)

E-value (VanderWeele and Ding 2017)*, this method can be used to assess the minimum strength of potential unmeasured confounding needed to explain away an observed effect estimate. this method can be conducted using only reported effect estimates, and explicitly requires that the association between confounder and exposure is of the same strength as the association between confounder and outcome and that the prevalence of the uncontrolled confound among the exposed is 100%;

E-value extensions (Cusson and Infante-Rivard 2020), this method can be used as an extension of the E-value method to accommodate more general situations, including studies where it is not possible to assume that the association between unmeasured confounder and exposure and the association between confounder and outcome are equal. Analyses can be conducted using the observed relative risk or odds ratio;

Generalized E-value (G-value) (MacLehose et al 2021), this method can be used to assess the minimum strength of potential unmeasured confounding needed to explain away an effect, and extends the E-value method by removing the assumption that the prevalence of the uncontrolled confounder among the exposed is 100%. Analyses can be conducted using the following information: the effect estimate and the prevalence of the confounder among the exposed.

5.2 Continuous (2)

Impact thresholds for regression models (Frank 2000), t his method can be used to quantify the impact of a confounding variable on the inference of a regression coefficient. Analyses can be conducted using the following information: the sample correlation between the outcome and predictor of interest, the estimated learn regression coefficient and standard error, the sample size, and the number of parameters estimate in the regression model (note: the approach can be extended beyond the general linear model (e.g., logistic regression), but the information provided is for linear models);

Robustness value (Cinelli and Hazlett 2019)*, this method can be used as an extension of the E-value method for regression models. Analyses can be conducted using the following information: the coefficient estimate, standard error, and degrees of freedom from a linear regression model.

4.3 Continuous (4)

5. Exposure data format

5.1 Categorical (4)

6. Outcome data format

6.1 Categorical (3)

Impact thresholds for regression models (Frank 2000), this method can be used to quantify the impact of a confounding variable on the inference of a regression coefficient. Analyses can be conducted using the following information: the sample correlation between the outcome and predictor of interest, the estimated regression coefficient and standard error, the sample size, and the number of parameters estimate in the regression model;

Bounding formulas for unmeasured confounding with effect-modifying potentials (Lee 2011), this method can be used to assess if the unmeasured confounder that may exhibit effect modification can explain away the findings. Analyses can be conducted using the following information: the confounded effect estimate, the association between an unmeasured confounder and outcome, the association between an unmeasured confounder and exposure, and the modifying effect that an unmeasured confounder has on the association between an exposure and outcome;

E-value (VanderWeele and Ding 2017)*, this method can be used to assess the minimum strength of potential unmeasured confounding needed to explain away an observed effect estimate. this method can be conducted using only reported effect estimates, and explicitly requires that the association between confounder and exposure is of the same strength as the association between confounder and outcome and that the prevalence of the uncontrolled confound among the exposed is 100%.

6.2 Continuous (3)

Impact thresholds for regression models (Frank 2000), this method can be used to quantify the impact of a confounding variable on the inference of a regression coefficient. Analyses can be conducted using the following information: the sample correlation between the outcome and predictor of interest, the estimated regression coefficient and standard error, the sample size, and the number of parameters estimate in the regression model;

E-value (VanderWeele and Ding 2017)*, this method can be used to assess the minimum strength of potential unmeasured confounding needed to explain away an observed effect estimate. this method can be conducted using only reported effect estimates, and explicitly requires that the association between confounder and exposure is of the same strength as the association between confounder and outcome and that the prevalence of the uncontrolled confound among the exposed is 100%;

Robustness value (Cinelli and Hazlett 2019)*, this method can be used as an extension of the E-value method for regression models. Analyses can be conducted using the following information: the coefficient estimate, standard error, and degrees of freedom from a linear regression model.

6.3 Time-to-event (1)

E-value (VanderWeele and Ding 2017)*, this method can be used to assess the minimum strength of potential unmeasured confounding needed to explain away an observed effect estimate. this method can be conducted using only reported effect estimates, and explicitly requires that the association between confounder and exposure is of the same strength as the association between confounder and outcome and that the prevalence of the uncontrolled confound among the exposed is 100%.

5.2 Continuous (2)

Impact thresholds for regression models (Frank 2000), this method can be used to quantify the impact of a confounding variable on the inference of a regression coefficient. Analyses can be conducted using the following information: the sample correlation between the outcome and predictor of interest, the estimated regression coefficient and standard error, the sample size, and the number of parameters estimate in the regression model;

Robustness value (Cinelli and Hazlett 2019)*, this method can be used as an extension of the E-value method for regression models. Analyses can be conducted using the following information: the coefficient estimate, standard error, and degrees of freedom from a linear regression model.

3.2 Corrected estimates (2)

Sensitivity analysis for unmeasured confounders for regression results (Lin et al 1998), this method can be used to assess the sensitivity of regression results to unmeasured confounders by generating bias-adjusted estimates. Analyses can be conducted using the following information: the association between the unmeasured confounder and outcome among the unexposed and exposed, the prevalence of the unmeasured confounder among the unexposed and exposed, and the confounded effect estimate;

Sensitivity analysis for interactions under unmeasured confounding (VanderWeele et al 2012), this method can be used to assess the sensitivity of interaction analyses to unmeasured confounding by generating bias-adjusted estimates. Depending on the type of interaction (additive or multiplicative), analyses can be conducted using different parameters, including the prevalence of the unmeasured confounder.

***2.1.2 Single unmeasured confounder (25)***

3. Result of interest

3.1 Explain-away (12)

4. Confounder data format

4.1 Categorical (12)

5. Exposure data format

5.1 Categorical (12)

6. Outcome data format

6.1 Categorical (11)

Sensitivity analysis for unmeasured confounding (Cornfield et al 1959), this method can be used to understand the impact of unmeasured confounding played on the exposure-outcome relationship. Analyses can be conducted using the following information: the prevalence of the confounder among the exposed, relative to the prevalence among those unexposed, and crude effect estimate between the exposure and outcome;

Basic methods for sensitivity analysis of unmeasured confounding (Greenland 1996), this method can be used to generate effect estimates that are corrected for potential unmeasured confounders. Analyses can be conducted using the following information: the information reported in 2 by 2 tables, the effect estimate between an unmeasured confounder and outcome, and the prevalence of unmeasured confounder among the unexposed and the exposed;

Impact thresholds for regression models (Frank 2000), this method can be used to quantify the impact of a confounding variable on the inference of a regression coefficient. Analyses can be conducted using the following information: the sample correlation between the outcome and predictor of interest, the estimated learn regression coefficient and standard error, the sample size, and the number of parameters estimate in the regression model (note: the approach can be extended beyond the general linear model (e.g., logistic regression), but the information provided is for linear models);

Sensitivity analysis for trend tests (Yu and Gastwirth 2005), this method can be used to assess the sensitivity of association between a multi-level exposure and a binary outcome to one unmeasured confounder. Analyses can be conducted using the following information: association between exposure and unmeasured confounder, association between unmeasured confounder and outcome, and prevalence of unmeasured confounder;

Rule-out approach/target-adjustment sensitivity analysis (Schneeweiss 2006), this method can be used to understand if the observed exposure-outcome association can be explained away by the unmeasured confounder. Analyses can be conducted using the following information: the prevalence of the exposure and confounder in the total population, the confounded effect estimate, and the effect estimate between the confounder and outcome;

Bounding formulas for population stratification bias (Lee and Wang 2007), this method can be used to generate effect estimates that are corrected for bias due to population stratification in case-control studies for genetic association studies. Analyses can be conducted using the following information: the background disease rate and the odds of the susceptibility genotype;

Bounding formulas for unmeasured confounding with effect-modifying potentials (Lee 2011), this method can be used to assess if the unmeasured confounder that may exhibit effect modification can explain away the findings. Analyses can be conducted using the following information: the confounded effect estimate, the association between an unmeasured confounder and outcome, the association between an unmeasured confounder and exposure, and the modifying effect that an unmeasured confounder has on the association between an exposure and outcome;

Average case calibration (Hasegawa and Small 2017), this method can be used to assess if the unmeasured confounding can explain away the findings according to the average bias in a pair in pair-matched studies. Analyses can be conducted using the probability that a unit with a positive outcome receives treatment;

E-value (VanderWeele and Ding 2017)*, this method can be used to assess the minimum strength of potential unmeasured confounding needed to explain away an observed effect estimate. this method can be conducted using only reported effect estimates, and explicitly requires that the association between confounder and exposure is of the same strength as the association between confounder and outcome and that the prevalence of the uncontrolled confound among the exposed is 100%;

E-value extensions (Cusson and Infante-Rivard 2020), this method can be used as an extension of the E-value method to accommodate more general situations, including studies where it is not possible to assume that the association between unmeasured confounder and exposure and the association between confounder and outcome are equal. Analyses can be conducted using the observed relative risk or odds ratio;

Generalized E-value (G-value) (MacLehose et al 2021), this method can be used to assess the minimum strength of potential unmeasured confounding needed to explain away an effect, and extends the E-value method by removing the assumption that the prevalence of the uncontrolled confounder among the exposed is 100%. Analyses can be conducted using the following information: the effect estimate and the prevalence of the confounder among the exposed.

6.2 Continuous (3)

Impact thresholds for regression models (Frank 2000), this method can be used to quantify the impact of a confounding variable on the inference of a regression coefficient. Analyses can be conducted using the following information: the sample correlation between the outcome and predictor of interest, the estimated learn regression coefficient and standard error, the sample size, and the number of parameters estimate in the regression model (note: the approach can be extended beyond the general linear model (e.g., logistic regression), but the information provided is for linear models);

E-value (VanderWeele and Ding 2017)*, this method can be used to assess the minimum strength of potential unmeasured confounding needed to explain away an observed effect estimate. this method can be conducted using only reported effect estimates, and explicitly requires that the association between confounder and exposure is of the same strength as the association between confounder and outcome and that the prevalence of the uncontrolled confound among the exposed is 100%;

Robustness value (Cinelli and Hazlett 2019)*, this method can be used as an extension of the E-value method for regression models. Analyses can be conducted using the following information: the coefficient estimate, standard error, and degrees of freedom from a linear regression model.

6.3 Time-to-event (3)

E-value (VanderWeele and Ding 2017)*, this method can be used to assess the minimum strength of potential unmeasured confounding needed to explain away an observed effect estimate. this method can be conducted using only reported effect estimates, and explicitly requires that the association between confounder and exposure is of the same strength as the association between confounder and outcome and that the prevalence of the uncontrolled confound among the exposed is 100%;

E-value extensions (Cusson and Infante-Rivard 2020), this method can be used as an extension of the E-value method to accommodate more general situations, including studies where it is not possible to assume that the association between unmeasured confounder and exposure and the association between confounder and outcome are equal. Analyses can be conducted using the observed relative risk or odds ratio;

Generalized E-value (G-value) (MacLehose et al 2021), this method can be used to assess the minimum strength of potential unmeasured confounding needed to explain away an effect, and extends the E-value method by removing the assumption that the prevalence of the uncontrolled confounder among the exposed is 100%. Analyses can be conducted using the following information: the effect estimate and the prevalence of the confounder among the exposed.

5.2 Continuous (2)

Impact thresholds for regression models (Frank 2000), this method can be used to quantify the impact of a confounding variable on the inference of a regression coefficient. Analyses can be conducted using the following information: the sample correlation between the outcome and predictor of interest, the estimated learn regression coefficient and standard error, the sample size, and the number of parameters estimate in the regression model (note: the approach can be extended beyond the general linear model (e.g., logistic regression), but the information provided is for linear models);

Robustness value (Cinelli and Hazlett 2019)*, this method can be used as an extension of the E-value method for regression models. Analyses can be conducted using the following information: the coefficient estimate, standard error, and degrees of freedom from a linear regression model.

4.2 Categorical (non-binary) (8)

5. Exposure data format

5.1 Categorical (8)

6. Outcome data format

6.1 Categorical (7)

Impact thresholds for regression models (Frank 2000), this method can be used to quantify the impact of a confounding variable on the inference of a regression coefficient. Analyses can be conducted using the following information: the sample correlation between the outcome and predictor of interest, the estimated learn regression coefficient and standard error, the sample size, and the number of parameters estimate in the regression model (note: the approach can be extended beyond the general linear model (e.g., logistic regression), but the information provided is for linear models);

Sensitivity analysis for trend tests (Yu and Gastwirth 2005), this method can be used to assess the sensitivity of association between a multi-level exposure and a binary outcome to one unmeasured confounder. Analyses can be conducted using the following information: association between exposure and unmeasured confounder, association between unmeasured confounder and outcome, and prevalence of unmeasured confounder;

Bounding formulas for population stratification bias (Lee and Wang 2007), this method can be used to generate effect estimates that are corrected for bias due to population stratification in case-control studies for genetic association studies. Analyses can be conducted using the following information: the background disease rate and the odds of the susceptibility genotype;

Bounding formulas for unmeasured confounding with effect-modifying potentials (Lee 2011), this method can be used to assess if the unmeasured confounder that may exhibit effect modification can explain away the findings. Analyses can be conducted using the following information: the confounded effect estimate, the association between an unmeasured confounder and outcome, the association between an unmeasured confounder and exposure, and the modifying effect that an unmeasured confounder has on the association between an exposure and outcome;

E-value (VanderWeele and Ding 2017)*, this method can be used to assess the minimum strength of potential unmeasured confounding needed to explain away an observed effect estimate. this method can be conducted using only reported effect estimates, and explicitly requires that the association between confounder and exposure is of the same strength as the association between confounder and outcome and that the prevalence of the uncontrolled confound among the exposed is 100%;

E-value extensions (Cusson and Infante-Rivard 2020), this method can be used as an extension of the E-value method to accommodate more general situations, including studies where it is not possible to assume that the association between unmeasured confounder and exposure and the association between confounder and outcome are equal. Analyses can be conducted using the observed relative risk or odds ratio;

Generalized E-value (G-value) (MacLehose et al 2021), this method can be used to assess the minimum strength of potential unmeasured confounding needed to explain away an effect, and extends the E-value method by removing the assumption that the prevalence of the uncontrolled confounder among the exposed is 100%. Analyses can be conducted using the following information: the effect estimate and the prevalence of the confounder among the exposed.

6.2 Continuous (3)

Impact thresholds for regression models (Frank 2000), this method can be used to quantify the impact of a confounding variable on the inference of a regression coefficient. Analyses can be conducted using the following information: the sample correlation between the outcome and predictor of interest, the estimated learn regression coefficient and standard error, the sample size, and the number of parameters estimate in the regression model (note: the approach can be extended beyond the general linear model (e.g., logistic regression), but the information provided is for linear models);

E-value (VanderWeele and Ding 2017)*, this method can be used to assess the minimum strength of potential unmeasured confounding needed to explain away an observed effect estimate. this method can be conducted using only reported effect estimates, and explicitly requires that the association between confounder and exposure is of the same strength as the association between confounder and outcome and that the prevalence of the uncontrolled confound among the exposed is 100%;

Robustness value (Cinelli and Hazlett 2019)*, this method can be used as an extension of the E-value method for regression models. Analyses can be conducted using the following information: the coefficient estimate, standard error, and degrees of freedom from a linear regression model.

6.3 Time-to-event (3)

E-value (VanderWeele and Ding 2017)*, this method can be used to assess the minimum strength of potential unmeasured confounding needed to explain away an observed effect estimate. this method can be conducted using only reported effect estimates, and explicitly requires that the association between confounder and exposure is of the same strength as the association between confounder and outcome and that the prevalence of the uncontrolled confound among the exposed is 100%;

E-value extensions (Cusson and Infante-Rivard 2020), this method can be used as an extension of the E-value method to accommodate more general situations, including studies where it is not possible to assume that the association between unmeasured confounder and exposure and the association between confounder and outcome are equal. Analyses can be conducted using the observed relative risk or odds ratio;

Generalized E-value (G-value) (MacLehose et al 2021), this method can be used to assess the minimum strength of potential unmeasured confounding needed to explain away an effect, and extends the E-value method by removing the assumption that the prevalence of the uncontrolled confounder among the exposed is 100%. Analyses can be conducted using the following information: the effect estimate and the prevalence of the confounder among the exposed.

5.2 Continuous (2)

Impact thresholds for regression models (Frank 2000), this method can be used to quantify the impact of a confounding variable on the inference of a regression coefficient. Analyses can be conducted using the following information: the sample correlation between the outcome and predictor of interest, the estimated learn regression coefficient and standard error, the sample size, and the number of parameters estimate in the regression model (note: the approach can be extended beyond the general linear model (e.g., logistic regression), but the information provided is for linear models);

Robustness value (Cinelli and Hazlett 2019)*, this method can be used as an extension of the E-value method for regression models. Analyses can be conducted using the following information: the coefficient estimate, standard error, and degrees of freedom from a linear regression model.

4.3 Continuous (5)

5. Exposure data format

5.1 Categorical (5)

6. Outcome data format

6.1 Categorical (4)

Impact thresholds for regression models (Frank 2000), this method can be used to quantify the impact of a confounding variable on the inference of a regression coefficient. Analyses can be conducted using the following information: the sample correlation between the outcome and predictor of interest, the estimated learn regression coefficient and standard error, the sample size, and the number of parameters estimate in the regression model (note: the approach can be extended beyond the general linear model (e.g., logistic regression), but the information provided is for linear models);

Sensitivity analysis for trend tests (Yu and Gastwirth 2005), this method can be used to assess the sensitivity of association between a multi-level exposure and a binary outcome to one unmeasured confounder. Analyses can be conducted using the following information: association between exposure and unmeasured confounder, association between unmeasured confounder and outcome, and prevalence of unmeasured confounder;

Bounding formulas for unmeasured confounding with effect-modifying potentials (Lee 2011), this method can be used to assess if the unmeasured confounder that may exhibit effect modification can explain away the findings. Analyses can be conducted using the following information: the confounded effect estimate, the association between an unmeasured confounder and outcome, the association between an unmeasured confounder and exposure, and the modifying effect that an unmeasured confounder has on the association between an exposure and outcome;

E-value (VanderWeele and Ding 2017)*, this method can be used to assess the minimum strength of potential unmeasured confounding needed to explain away an observed effect estimate. this method can be conducted using only reported effect estimates, and explicitly requires that the association between confounder and exposure is of the same strength as the association between confounder and outcome and that the prevalence of the uncontrolled confound among the exposed is 100%.

6.2 Continuous (2)

Impact thresholds for regression models (Frank 2000), this method can be used to quantify the impact of a confounding variable on the inference of a regression coefficient. Analyses can be conducted using the following information: the sample correlation between the outcome and predictor of interest, the estimated learn regression coefficient and standard error, the sample size, and the number of parameters estimate in the regression model (note: the approach can be extended beyond the general linear model (e.g., logistic regression), but the information provided is for linear models);

Robustness value (Cinelli and Hazlett 2019)*, this method can be used as an extension of the E-value method for regression models. Analyses can be conducted using the following information: the coefficient estimate, standard error, and degrees of freedom from a linear regression model.

6.3 Time-to-event (1)

E-value (VanderWeele and Ding 2017)*, this method can be used to assess the minimum strength of potential unmeasured confounding needed to explain away an observed effect estimate. this method can be conducted using only reported effect estimates, and explicitly requires that the association between confounder and exposure is of the same strength as the association between confounder and outcome and that the prevalence of the uncontrolled confound among the exposed is 100%.

5.2 Continuous (2)

Impact thresholds for regression models (Frank 2000), this method can be used to quantify the impact of a confounding variable on the inference of a regression coefficient. Analyses can be conducted using the following information: the sample correlation between the outcome and predictor of interest, the estimated learn regression coefficient and standard error, the sample size, and the number of parameters estimate in the regression model (note: the approach can be extended beyond the general linear model (e.g., logistic regression), but the information provided is for linear models);

Robustness value (Cinelli and Hazlett 2019)*, this method can be used as an extension of the E-value method for regression models. Analyses can be conducted using the following information: the coefficient estimate, standard error, and degrees of freedom from a linear regression model.

3.2 Corrected estimates (12)

4. Confounder data format

4.1 Categorical (12)

5. Exposure data format

5.1 Categorical (12)

6. Outcome data format

6.1 Categorical (12)

Size rule/array approach for unmeasured confounding (Bross 1966)*, this method can be used to generate bias-adjusted estimates corrected for an unmeasured confounder. Analyses can be conducted using the following information: the prevalence of a confounder in the exposed and the unexposed, the confounded effect estimate, and the association between a confounder and an outcome;

Method for unmeasured confounding (Schlesselman 1978), this method can be used to generate bias-adjusted estimates corrected for an unmeasured confounder. Analyses can be conducted using the following information: the prevalence of a confounder in the exposed, the prevalence of a confounder in the unexposed, and the confounded effect estimate;

Method for unmeasured confounding (Rosenbaum and Rubin 1983), this method can be used to generate bias-adjusted estimates corrected for an unmeasured confounder. Analyses can be conducted using the following information: the proportion of the unexposed, the prevalence of the unmeasured confounder, the association between the unmeasured confounder and the outcome among the unexposed and exposed, and the association between the unmeasured confounder and the exposure;

Sensitivity analysis for unmeasured confounders for regression results (Lin et al 1998), this method can be used to assess the sensitivity of regression results to unmeasured confounders by generating bias-adjusted estimates. Analyses can be conducted using the following information: the association between the unmeasured confounder and outcome among the unexposed and exposed, the prevalence of the unmeasured confounder among the unexposed and exposed, and the confounded effect estimate;

Quantifying biases for classical confounding and (or) collider-stratification bias (Greenland 2003), this method can be used to examine the potential impact of confounding with or without selection bias under simple causal models. Analyses can be conducted using the following information: the reported associations between exposure, outcome, confounder, and collider. For unmeasured confounding, the analyses also require the confounded effect estimate and the prevalence of the confounder among the unexposed controls;

Monte Carlo sensitivity analysis and Bayesian analysis for unmeasured confounding (Steenland and Greenland 2004)*, this method can be used to generate a range of effect measures given the likely range of bias using Monte Carlo sensitivity analysis and/or Bayesian analysis. Typically, researchers need to combine the priors for unknown parameters with the probability of the observed data, using Bayes’ theorem, to produce a posterior distribution for the parameter of interest (i.e., the bias-adjusted rate ratio);

Integration of Rosenbaum and Greenland method for confounding (Cabral 2007)*, this method, which integrates two previous sensitivity analyses methods, can be used to generate effect estimates that are corrected for potential unmeasured confounders. Analyses can be conducted using the following information: the cells from a 2 by 2 table and odds ratios between the exposure and the confounder, and the confounder and the outcome;

Method for unmeasured confounders (Arah et al 2008), this method can be used to generate effect estimates that are corrected for potential unmeasured confounders. Analyses can be conducted using the following information: the prevalence of the confounder among the exposed, unexposed, and total population, and the effect estimates for confounder-stratified exposure-outcome relationships in the total population;

Probabilistic bias analysis for two biases (Lash et al 2010)*, this method can be used to generate bias-adjusted estimates that are corrected for potential unmeasured confounders. Analyses can be conducted using the following information: the distribution of the association between the confounder and the outcome, and the distribution of prevalence of the confounder in the exposed and the unexposed;

Sensitivity analysis for unmeasured confounding of attributable fraction (Chiba 2011), this method can be used to generate bias-adjusted estimates corrected for unmeasured confounding when the effect measure of interest is attributable fraction. Analyses can be conducted using the following information: the confounded effect estimate, sensitivity parameter, average difference in potential outcomes between those exposed and those unexposed, disease prevalence, and exposure prevalence;

Method for unmeasured confounding for general outcomes, treatments, and confounders (VanderWeele and Arah 2011), this method can be used to control for the impact of unmeasured confounders under different situations, including studies with categorical or continuous exposures, unmeasured confounders, and outcomes, and effect estimates that are on the additive, risk-ratio, and odds-ratio scales. Analyses can be conducted using the following information: the associations between the unmeasured confounder and outcome and the distribution of the unmeasured confounder among different exposure levels;

Sensitivity analysis for interactions under unmeasured confounding (VanderWeele et al 2012), this method can be used to assess the sensitivity of interaction analyses to unmeasured confounding by generating bias-adjusted estimates. Depending on the type of interaction (additive or multiplicative), analyses can be conducted using different parameters, including the prevalence of the unmeasured confounder.

6.2 Continuous (4)

Sensitivity analysis for unmeasured confounders for regression results (Lin et al 1998), this method can be used to assess the sensitivity of regression results to unmeasured confounders by generating bias-adjusted estimates. Analyses can be conducted using the following information: the association between the unmeasured confounder and outcome among the unexposed and exposed, the prevalence of the unmeasured confounder among the unexposed and exposed, and the confounded effect estimate;

Integration of Rosenbaum and Greenland method for confounding (Cabral 2007)*, this method, which integrates two previous sensitivity analyses methods, can be used to generate effect estimates that are corrected for potential unmeasured confounders. Analyses can be conducted using the following information: the cells from a 2 by 2 table and odds ratios between the exposure and the confounder, and the confounder and the outcome;

Method for unmeasured confounding for general outcomes, treatments, and confounders (VanderWeele and Arah 2011), this method can be used to control for the impact of unmeasured confounders under different situations, including studies with categorical or continuous exposures, unmeasured confounders, and outcomes, and effect estimates that are on the additive, risk-ratio, and odds-ratio scales. Analyses can be conducted using the following information: the associations between the unmeasured confounder and outcome and the distribution of the unmeasured confounder among different exposure levels;

Sensitivity analysis for interactions under unmeasured confounding (VanderWeele et al 2012), this method can be used to assess the sensitivity of interaction analyses to unmeasured confounding by generating bias-adjusted estimates. Depending on the type of interaction (additive or multiplicative), analyses can be conducted using different parameters, including the prevalence of the unmeasured confounder.

6.3 Time-to-event (2)

Sensitivity analysis for unmeasured confounders for regression results (Lin et al 1998), this method can be used to assess the sensitivity of regression results to unmeasured confounders by generating bias-adjusted estimates. Analyses can be conducted using the following information: the association between the unmeasured confounder and outcome among the unexposed and exposed, the prevalence of the unmeasured confounder among the unexposed and exposed, and the confounded effect estimate;

Probabilistic bias analysis for two biases (Lash et al 2010)*, this method can be used to generate bias-adjusted estimates that are corrected for potential unmeasured confounders. Analyses can be conducted using the following information: the distribution of the association between the confounder and the outcome, and the distribution of prevalence of the confounder in the exposed and the unexposed.

5.2 Continuous (3)

Integration of Rosenbaum and Greenland method for confounding (Cabral 2007)*, this method, which integrates two previous sensitivity analyses methods, can be used to generate effect estimates that are corrected for potential unmeasured confounders. Analyses can be conducted using the following information: the cells from a 2 by 2 table and odds ratios between the exposure and the confounder, and the confounder and the outcome;

Method for unmeasured confounding for general outcomes, treatments, and confounders (VanderWeele and Arah 2011), this method can be used to control for the impact of unmeasured confounders under different situations, including studies with categorical or continuous exposures, unmeasured confounders, and outcomes, and effect estimates that are on the additive, risk-ratio, and odds-ratio scales. Analyses can be conducted using the following information: the associations between the unmeasured confounder and outcome and the distribution of the unmeasured confounder among different exposure levels;

Sensitivity analysis for interactions under unmeasured confounding (VanderWeele et al 2012), this method can be used to assess the sensitivity of interaction analyses to unmeasured confounding by generating bias-adjusted estimates. Depending on the type of interaction (additive or multiplicative), analyses can be conducted using different parameters, including the prevalence of the unmeasured confounder.

4.2 Categorical (non-binary) (7)

5. Exposure data format

5.1 Categorical (7)

6. Outcome data format

6.1 Categorical (7)

Sensitivity analysis for unmeasured confounders for regression results (Lin et al 1998), this method can be used to assess the sensitivity of regression results to unmeasured confounders by generating bias-adjusted estimates. Analyses can be conducted using the following information: the association between the unmeasured confounder and outcome among the unexposed and exposed, the prevalence of the unmeasured confounder among the unexposed and exposed, and the confounded effect estimate;

Monte Carlo sensitivity analysis and Bayesian analysis for unmeasured confounding (Steenland and Greenland 2004)*, this method can be used to generate a range of effect measures given the likely range of bias using Monte Carlo sensitivity analysis and/or Bayesian analysis. Typically, researchers need to combine the priors for unknown parameters with the probability of the observed data, using Bayes’ theorem, to produce a posterior distribution for the parameter of interest (i.e., the bias-adjusted rate ratio);

Method for unmeasured confounders (Arah et al 2008), this method can be used to generate effect estimates that are corrected for potential unmeasured confounders. Analyses can be conducted using the following information: the prevalence of the confounder among the exposed, unexposed, and total population, and the effect estimates for confounder-stratified exposure-outcome relationships in the total population;

Probabilistic bias analysis for two biases (Lash et al 2010)*, this method can be used to generate bias-adjusted estimates that are corrected for potential unmeasured confounders. Analyses can be conducted using the following information: the distribution of the association between the confounder and the outcome, and the distribution of prevalence of the confounder in the exposed and the unexposed;

Sensitivity analysis for unmeasured confounding of attributable fraction (Chiba 2011), this method can be used to generate bias-adjusted estimates corrected for unmeasured confounding when the effect measure of interest is attributable fraction. Analyses can be conducted using the following information: the confounded effect estimate, sensitivity parameter, average difference in potential outcomes between those exposed and those unexposed, disease prevalence, and exposure prevalence;

Method for unmeasured confounding for general outcomes, treatments, and confounders (VanderWeele and Arah 2011), this method can be used to control for the impact of unmeasured confounders under different situations, including studies with categorical or continuous exposures, unmeasured confounders, and outcomes, and effect estimates that are on the additive, risk-ratio, and odds-ratio scales. Analyses can be conducted using the following information: the associations between the unmeasured confounder and outcome and the distribution of the unmeasured confounder among different exposure levels;

Sensitivity analysis for interactions under unmeasured confounding (VanderWeele et al 2012), this method can be used to assess the sensitivity of interaction analyses to unmeasured confounding by generating bias-adjusted estimates. Depending on the type of interaction (additive or multiplicative), analyses can be conducted using different parameters, including the prevalence of the unmeasured confounder.

6.2 Continuous (3)

Sensitivity analysis for unmeasured confounders for regression results (Lin et al 1998), this method can be used to assess the sensitivity of regression results to unmeasured confounders by generating bias-adjusted estimates. Analyses can be conducted using the following information: the association between the unmeasured confounder and outcome among the unexposed and exposed, the prevalence of the unmeasured confounder among the unexposed and exposed, and the confounded effect estimate;

Method for unmeasured confounding for general outcomes, treatments, and confounders (VanderWeele and Arah 2011), this method can be used to control for the impact of unmeasured confounders under different situations, including studies with categorical or continuous exposures, unmeasured confounders, and outcomes, and effect estimates that are on the additive, risk-ratio, and odds-ratio scales. Analyses can be conducted using the following information: the associations between the unmeasured confounder and outcome and the distribution of the unmeasured confounder among different exposure levels;

Sensitivity analysis for interactions under unmeasured confounding (VanderWeele et al 2012), this method can be used to assess the sensitivity of interaction analyses to unmeasured confounding by generating bias-adjusted estimates. Depending on the type of interaction (additive or multiplicative), analyses can be conducted using different parameters, including the prevalence of the unmeasured confounder.\

6.3 Time-to-event (0)

5.2 Continuous (2)

Method for unmeasured confounding for general outcomes, treatments, and confounders (VanderWeele and Arah 2011), this method can be used to control for the impact of unmeasured confounders under different situations, including studies with categorical or continuous exposures, unmeasured confounders, and outcomes, and effect estimates that are on the additive, risk-ratio, and odds-ratio scales. Analyses can be conducted using the following information: the associations between the unmeasured confounder and outcome and the distribution of the unmeasured confounder among different exposure levels;

Sensitivity analysis for interactions under unmeasured confounding (VanderWeele et al 2012), this method can be used to assess the sensitivity of interaction analyses to unmeasured confounding by generating bias-adjusted estimates. Depending on the type of interaction (additive or multiplicative), analyses can be conducted using different parameters, including the prevalence of the unmeasured confounder.

4.3 Continuous (3)

5. Exposure data format

5.1 Categorical (3)

6. Outcome data format

6.1 Categorical (3)

Sensitivity analysis for unmeasured confounders for regression results (Lin et al 1998), this method can be used to assess the sensitivity of regression results to unmeasured confounders by generating bias-adjusted estimates. Analyses can be conducted using the following information: the association between the unmeasured confounder and outcome among the unexposed and exposed, the prevalence of the unmeasured confounder among the unexposed and exposed, and the confounded effect estimate;

Sensitivity analysis for unmeasured confounding of attributable fraction (Chiba 2011), this method can be used to generate bias-adjusted estimates corrected for unmeasured confounding when the effect measure of interest is attributable fraction. Analyses can be conducted using the following information: the confounded effect estimate, sensitivity parameter, average difference in potential outcomes between those exposed and those unexposed, disease prevalence, and exposure prevalence;

Method for unmeasured confounding for general outcomes, treatments, and confounders (VanderWeele and Arah 2011), this method can be used to control for the impact of unmeasured confounders under different situations, including studies with categorical or continuous exposures, unmeasured confounders, and outcomes, and effect estimates that are on the additive, risk-ratio, and odds-ratio scales. Analyses can be conducted using the following information: the associations between the unmeasured confounder and outcome and the distribution of the unmeasured confounder among different exposure levels.

6.2 Continuous (2)

Sensitivity analysis for unmeasured confounders for regression results (Lin et al 1998), this method can be used to assess the sensitivity of regression results to unmeasured confounders by generating bias-adjusted estimates. Analyses can be conducted using the following information: the association between the unmeasured confounder and outcome among the unexposed and exposed, the prevalence of the unmeasured confounder among the unexposed and exposed, and the confounded effect estimate;

Method for unmeasured confounding for general outcomes, treatments, and confounders (VanderWeele and Arah 2011), this method can be used to control for the impact of unmeasured confounders under different situations, including studies with categorical or continuous exposures, unmeasured confounders, and outcomes, and effect estimates that are on the additive, risk-ratio, and odds-ratio scales. Analyses can be conducted using the following information: the associations between the unmeasured confounder and outcome and the distribution of the unmeasured confounder among different exposure levels.

6.3 Time-to-event (1)

Sensitivity analysis for unmeasured confounders for regression results (Lin et al 1998), this method can be used to assess the sensitivity of regression results to unmeasured confounders by generating bias-adjusted estimates. Analyses can be conducted using the following information: the association between the unmeasured confounder and outcome among the unexposed and exposed, the prevalence of the unmeasured confounder among the unexposed and exposed, and the confounded effect estimate.

5.2 Continuous (1)

Method for unmeasured confounding for general outcomes, treatments, and confounders (VanderWeele and Arah 2011), this method can be used to control for the impact of unmeasured confounders under different situations, including studies with categorical or continuous exposures, unmeasured confounders, and outcomes, and effect estimates that are on the additive, risk-ratio, and odds-ratio scales. Analyses can be conducted using the following information: the associations between the unmeasured confounder and outcome and the distribution of the unmeasured confounder among different exposure levels.

***2.2 Misclassification bias (14)***

***2.2.1 Exposure misclassification bias (11)***

3. Misclassification type

3.1 Differential exposure misclassification bias (7)

4. Exposure data type

4.1 Categorical (7)

5. Outcome data type

5.1 Categorical (7)

Method for misclassification bias in the estimation of relative risk (Copeland et al 1977), this method can be used to correct for differential or nondifferential misclassification of study subjects (i.e., disease occurrence in cohorts or exposure history in case-control studies,) in 2 by 2 tables. Analyses can be conducted using the following information: the four cells from 2 by 2 tables and estimates of specificity of the classification procedure;

Generalizations of Barren’s matrix correction method (Greenland and Kleinbaum 1983), this method can be used to generate bias-adjusted effect estimates in situations involving differential misclassification, matched-pair data, and arbitrary two-way tables. Analyses can be conducted by constructing a correction matrix using the misclassified cells in a 2 by 2 table and the probability of misclassification;

Method for using predictive values for misclassification (Marshall 1990), this method can be used to generate bias-adjusted effect estimates correcting for the differential or nondifferential exposure misclassification. Analyses can be conducted using the following information: the positive and negative predictive values and the prevalence of self-reported exposure and its variance;

Quality indices for exposure misclassification (Marshall 1994), this method can be used to generate bias-adjusted estimates correcting for misclassification by quality (i.e, rescaled sensitivity and specificity indices) based directly on the probabilities of disagreement between measured and true exposure. Analyses can be conducted using parameters on how cases were measured in a case-control studies;

Alloyed gold standard for exposure misclassification (Brenner 1996), this method can be used to correct for misclassification in the presence of an alloyed gold standard. Analyses can be conducted using the following information: the exposure prevalence in the population and the sensitivity (specificity) of exposure measurement by the alloyed gold standard from validation data and the routinely employed method from routinely-collected data;

Bayesian approach for misclassification (Chu et al 2006)*, this method can be used to correct for exposure misclassification in case-control studies. Analyses can be conducted using multiple parameters, including the observed proportions of cases and controls classified as exposed;

Method for differential measurement error (VanderWeele and Li 2019), this method can be used to assess how strong differential measurement error would have to be to completely explain away an observed exposure-outcome association. Analyses can be conducted using the following information: the observed and potential true risk ratio between exposure and outcome.

5.2 Continuous (1)

Method for differential measurement error (VanderWeele and Li 2019), this method can be used to assess how strong differential measurement error would have to be to completely explain away an observed exposure-outcome association. Analyses can be conducted using the following information: the observed and potential true risk ratio between exposure and outcome.

5.3 Time-to-event (0)

4.2 Continuous (1)

Method for differential measurement error (VanderWeele and Li 2019), this method can be used to assess how strong differential measurement error would have to be to completely explain away an observed exposure-outcome association. Analyses can be conducted using the following information: the observed and potential true risk ratio between exposure and outcome.

3.2 Nondifferential exposure misclassification bias (10)

4. Exposure data type

4.1 Categorical (10)

5.Outcome data type

5.1 Categorical (10)

Method for misclassification bias in the estimation of relative risk (Copeland et al 1977), this method can be used to correct for differential or nondifferential misclassification of study subjects (i.e., disease occurrence in cohorts or exposure history in case-control studies,) in 2 by 2 tables. Analyses can be conducted using the following information: the four cells from 2 by 2 tables and estimates of specificity of the classification procedure;

Method for misclassification bias in matched-pair studies (Greenland 1982), this method can be used to correct for misclassification bias in matched-pair case-control studies. Analyses can be conducted using the following information: the reported false negative rate, false positive rate, true negative rate, and true positive rate among cases and controls;

Generalizations of Barren’s matrix correction method (Greenland and Kleinbaum 1983), this method can be used to generate bias-adjusted effect estimates in situations involving differential misclassification, matched-pair data, and arbitrary two-way tables. Analyses can be conducted by constructing a correction matrix using the misclassified cells in a 2 by 2 table and the probability of misclassification;

Method for using predictive values for misclassification (Marshall 1990), this method can be used to generate bias-adjusted effect estimates correcting for the differential or nondifferential exposure misclassification. Analyses can be conducted using the following information: the positive and negative predictive values and the prevalence of self-reported exposure and its variance;

Sensitivity analysis for validation studies using an alloyed gold standard (Wacholder et al 1993), this method can be used to explore the influence of misclassification bias in an alloyed gold standard when a validation study is used to correct effect estimates. Analyses can be conducted using the following information: the coefficient estimate of the regression of the outcome on the measured exposure, and the correction factor from the validation study with the alloyed gold standard;

Quality indices for exposure misclassification (Marshall 1994), this method can be used to generate bias-adjusted estimates correcting for misclassification by quality (i.e, rescaled sensitivity and specificity indices) based directly on the probabilities of disagreement between measured and true exposure. Analyses can be conducted using parameters on how cases were measured in a case-control studies;

Alloyed gold standard for exposure misclassification (Brenner 1996), this method can be used to correct for misclassification in the presence of an alloyed gold standard. Analyses can be conducted using the following information: the exposure prevalence in the population and the sensitivity (specificity) of exposure measurement by the alloyed gold standard from validation data and the routinely employed method from routinely-collected data;

Bayesian approach for misclassification (Chu et al 2006)*, this method can be used to correct for exposure misclassification in case-control studies. Analyses can be conducted using multiple parameters, including the observed proportions of cases and controls classified as exposed;

Bayesian analysis of a matched case-control study (Liu et al 2019)*, this method can be used to assess the impact of misclassification of a binary exposure variable in a matched case–control study. Analyses can be conducted using the following information: the marginal probability of true exposure status and the sensitivity and specificity of the exposure classification among cases and controls;

Method for exposure misclassification bias in vaccine effectiveness (Baum et al 2021), this method can be used to assess biases under non-differential exposure misclassification when estimating vaccine effectiveness. Analyses can be conducted using various reported parameters, including the risk of the disease in those observed as vaccinated and unvaccinated, and the sensitivity and specificity of the vaccine.

5.2 Continuous (0)

5.3 Time-to-event (0)

4.2 Continuous (1)

Sensitivity analysis for validation studies using an alloyed gold standard (Wacholder et al 1993), this method can be used to explore the influence of misclassification bias in an alloyed gold standard when a validation study is used to correct effect estimates. Analyses can be conducted using the following information: the coefficient estimate of the regression of the outcome on the measured exposure, and the correction factor from the validation study with the alloyed gold standard.

***2.2.2 Outcome misclassification bias (4)***

3. Misclassification type

3.1 Differential exposure misclassification bias (2)

Generalizations of Barren’s matrix correction method (Greenland and Kleinbaum 1983), this method can be used to generate bias-adjusted effect estimates in situations involving differential misclassification, matched-pair data, and arbitrary two-way tables. Analyses can be conducted by constructing a correction matrix using the misclassified cells in a 2 by 2 table and the probability of misclassification;

Probabilistic bias analysis for two biases (Lash et al 2010)*, this method can be used to generate effect estimates that are corrected for bias from outcome misclassification. Analyses can be conducted using the following information: the number of observed cases and the distribution of sensitivities of the exposed and unexposed.

3.2 Nondifferential exposure misclassification bias (4)

4. Exposure data type

4.1 Categorical (4)

Generalizations of Barren’s matrix correction method (Greenland and Kleinbaum 1983), this method can be used to generate bias-adjusted effect estimates in situations involving differential misclassification, matched-pair data, and arbitrary two-way tables. Analyses can be conducted by constructing a correction matrix using the misclassified cells in a 2 by 2 table and the probability of misclassification; Probabilistic bias analysis for two biases (Lash et al 2010)*, this method can be used to generate effect estimates that are corrected for bias from outcome misclassification. Analyses can be conducted using the following information: the number of observed cases and the distribution of sensitivities of the exposed and unexposed;

Methods to adjust for outcome misclassification accounting for case-control sampling with external information (Jurek et al 2013)*, this method can be used to account for case-control sampling with external information and to adjust for outcome misclassification in case-control studies. Analyses can be conducted using reported parameters, including the probability of outcome being correctly classified and the probability of the non-outcome being correctly classified;

Bias correction methods for misclassification in test-negative designs (Endo et al 2020)*, this method can be used to correct for misclassification in test-negative study designs. Analyses can be conducted using the following information: the reported sensitivity and specificity of the test, number of cases among the unvaccinated and vaccinated participants, and number of controls among unvaccinated and vaccinated participants.

4.2 Continuous (0)

***2.2.3 Confounder misclassification bias (1)***

Method for nondifferential confounder misclassification (Savitz and Baron 1989), this method can be used to generate bias adjusted estimates corrected for nondifferential confounder misclassification in case control studies. Analyses can be conducted using the following information: the unadjusted, partially adjusted, and completely adjusted effect estimates.

***2.3 Selection bias (3)***

3. Result of interest

3.1 Explain-away (1)

E-value for selection bias (Smith and VanderWeele 2019, explain-away)*, this method can be used to assess the minimum strength of potential selection bias needed to explain away an observed effect. Analyses can be conducted using the crude effect estimate and corresponding 95% CIs.

3.2 Corrected estimates (2)

Quantifying biases for classical confounding and (or) collider-stratification bias (Greenland 2003), this method can be used to examine the relative magnitudes of selection biases under some simple causal models in which the stratification variable is graphically depicted as a collider. Analyses can be conducted using the following information: the reported associations between exposure, outcome, confounder, and collider;

E-value for selection bias (Smith and VanderWeele 2019, corrected estimates)*, this method can be used to generate selection bias-adjusted effect estimates. Analyses can be conducted using the following information: the reported crude effect estimate and various parameters related to the selection process.

***2.4 Multiple biases (2)***

3. Result of interest

3.1 Explain-away (1)

Bounding formulas for selection bias (Huang and Lee 2015), this method, which extends previous bounding formulas for unmeasured confounding and effect modification, can be used to assess if selection bias can explain away an observed finding. Analyses can be conducted using the associations between all variables, including the exposure, outcome, variable indicating the selection of study subjects, and unmeasured confounder.

3.2 Corrected estimates (1)

Bounding method for multiple biases (Smith et al 2021), this method can be used to estimate bounds for the total composite bias (selection bias, unmeasured confounding, and misclassification bias) under a variety of scenarios. Analyses can be conducted using different parameters related to the biases of interest.

**1.2.3 Cross-sectional studies (1)**

***2. Bias type***

***2.1 Unmeasured confounding (0)***

***2.2 Misclassification bias (1)***

Method for outcome misclassification (Green 1983), this method can be used to correct for relative risk estimates biased by misclassification of outcome status. Analyses can be conducted using the following information: the crude risk ratio, predictive value of a positive outcome, and true disease frequency in the unexposed group only.

***2.3 Selection bias (0)***

**1.3 Meta-analyses (4)**

***2. Bias type***

***2.1 Unmeasured confounding (4)***

3. Result of interest

3.1 Explain-away (2)

Sensitivity analysis for unmeasured confounding in meta-analyses (Mathur and VanderWeele 2020), this method can be used to assess if the unmeasured confounding can explain away findings in meta-analyses of more than 10 studies. Analyses can be conducted using the summary effect estimate from a meta-analysis.

Robust methods for point estimation and inference in meta-analyses (Mathur and VanderWeele 2020), this nonparametric method can be used to assess if the unmeasured confounding can explain away findings in meta-analyses of more than 10 studies. Analyses can be conducted using the effect estimate and standard error from each study included in a meta-analysis. This method makes no assumptions about the distribution of population effects, and can be used even when the proportion being estimated is close to 0 or 1. However, the robust method only accommodates bias whose strength is the same in all studies (i.e., homogeneous bias).

3.2 Corrected estimates (1)

Bayesian method for unmeasured confounding in meta-analyses (McCandless 2012), this method can be used to generate effect estimates from meta-analyses that are corrected for an unmeasured confounder. Analyses can be conducted using the following information from all component studies: the association between exposure and outcome, the association between unmeasured confounder and outcome, conditional on the exposure and measured covariates, and the prevalence of the unmeasured confounder in exposed and unexposed individuals.

***2.2 Misclassification bias (0)***

***2.3 Selection bias (0)***

***2.3 Multiple biases (1)***

Method for biases in meta-analyses (Turner et al 2009), this method can be used to generate effect estimates that are corrected for multiple biases at a time in a meta-analysis. Analyses can be conducted using mainly the specified range of possible biases in different categories.

### Unmeasured confounding (corrected estimate)

#### Bross 1966: size rule/array approach for unmeasured confounding

| **Name/description** | **Size rule/Array approach** |
| --- | --- |
| **DOI** | https://doi.org/10.1016/0021-9681(66)90062-2 |
| **Author and Year** | Original manuscript: Bross 1966^1^.  Additional information: Schneeweiss 2006^2^. This review article provides a clear overview of the methods and was used to populate the cells below. |
| **Crossed-referenced in QBA textbook^3^** | Referenced and explained |
| **QBA classification** | Simple sensitivity analysis |
| **Applicable study design scenarios described^b^** | Any observational study design |
| **Effect measure of interest** | RR or RD |
| **Result of interest** | Corrected estimates |
| **Data structure/format of the eligible exposure/intervention^b^** | Categorical |
| **Data structure/format of the confounders, if applicable^b^** | Categorical |
| **Data structure/format of eligible outcome^b^** | Categorical |
| **Source(s) of bias addressed** | Unmeasured confounding (single) |
| **Information/parameters needed to conduct the QBA method** | - **ARR** or **ARD**: apparent (or confounded/observed) RR or RD;  - **P_C0_**: Prevalence of confounder in the unexposed;  - **P_C1_**: Prevalence of confounder in the exposed;  - **RR_CD_** or **RD_CD_**: RR or RD between confounder and disease outcome. |
| **Mathematical formula** | $\mathbf{RR}=\frac{\mathrm{ARR}}{\left[ \frac{P_{\mathrm{Cl}}\left( \mathrm{RR}_{\mathrm{CD}}-1 \right)+1}{P_{\mathrm{Co}}\left( \mathrm{RR}_{\mathrm{CD}}-1 \right)+1} \right]}$;  Or$\mathbf{RD}=ARD-\left( P_{C1}-P_{C0} \right)\mathrm{RD}_{\mathrm{CD}}$.^2^ |
| **Data assumptions and additional required features** | Not mentioned |
| **Available and/or recommended software** | Excel spreadsheet (drugepi.org/dope/software#Sensitivity-analysis) |
| **Output of the QBA analysis** | Corrected estimates |
| **Interpretation of the QBA analysis output** | Effect estimate accounting for unmeasured confounding |
| **Considerations related to the interpretation of the results** | 1. The authors noted that “In realistic settings, ARR is already adjusted for a set of measured covariates and the interest is in assessing the residual confounding by additional covariates not measured in the main study.”^1^  2. The method is strictly constrained to only one binary confounder. |
| **Explicit discussions regarding the similarities and differences with other methods** | Zhang et al 2006 evaluated the E-value, array approach, and rule-out approach, and found that the results were numerically equivalent.^4^ |

#### Schlesselman 1978: method for unmeasured confounding

| **Name/description** | **Method for unmeasured confounding** |
| --- | --- |
| **DOI** | https://doi.org/10.1093/oxfordjournals.aje.a112581 |
| **Author and Year** | Schlesselman 1978^5^ |
| **Crossed-referenced in QBA textbook^3^** | Referenced and explained |
| **QBA classification** | Simple sensitivity analysis |
| **Applicable study design scenarios described^b^** | Cohort or case-control studies |
| **Effect measure of interest** | RR or OR |
| **Result of interest** | Corrected estimate |
| **Data structure/format of the eligible exposure/intervention^b^** | Categorical |
| **Data structure/format of the confounders, if applicable^b^** | Categorical |
| **Data structure/format of eligible outcome^b^** | Categorical |
| **Source(s) of bias addressed** | Unmeasured confounding (single) |
| **Information/parameters needed to conduct the QBA method** | - **p_1_, p_2_**: the prevalence of the unmeasured confounder among the exposed and the unexposed, respectively;  - **R_A_**: measured relative risk between exposure and outcome (with 95% CI);  **When there is no interaction between confounder and exposure,**  - **r**: relative risk due to unmeasured confounder in the absence of exposure;  **When there is an interaction between confounder and exposure,**  - $\boldsymbol{r}_{\boldsymbol{c}}\boldsymbol{,}\boldsymbol{r}_{\boldsymbol{c}}^{\mathbf{'}}:$relative risk due to unmeasured confounder in the absence and presence of exposure, respectively. |
| **Mathematical formula** | **When there is no interaction between confounder and exposure,**  $\begin{matrix} S= & \left[ rp_{1}+\left( 1-p_{1} \right) \right]/ \\ & \left[ rp_{2}+\left( 1-p_{2} \right) \right] \end{matrix};$  $r_{A}=\frac{R_{A}}{S.}$  **When there is an interaction between confounder and exposure,**  $\begin{matrix} S= & \left[ r_{c}^{'}p_{1}+\left( 1-p_{1} \right) \right]/ \\ & \left[ r_{c}p_{2}+\left( 1-p_{2} \right) \right] \end{matrix};$  $r_{A}=\frac{R_{A}}{S.}$ |
| **Data assumptions and additional required features** | An unstated assumption is that the relative risks between the unmeasured confounder and outcome among the unexposed and exposed should be the same; if this assumption is violated, the formula in Lin et al 1998’s paper can be used.^6^ |
| **Available and/or recommended software** | Not mentioned |
| **Output of the QBA analysis** | Corrected effect estimate $r_{A}$with 95% CIs |
| **Interpretation of the QBA analysis output** | Effect estimate accounting for unmeasured confounding |
| **Considerations related to the interpretation of the results** | 1. When multiple unmeasured confounders are present, the author noted that this method “would quickly lead to complications requiring either extensive knowledge or strong assumptions about the separate and joint effects of the variables.”^5^  2. When there is no interaction between confounder and exposure, if the exposure does not affect the risk of disease (i.e., $r_{A}=1$), then the author noted that the equation is equivalent to the result given first by Cornfield et al 1959^7^ and later by Bross 1966.^1^ |
| **Explicit discussions regarding the similarities and differences with other methods** | 1. The author noted that equation for no interaction between confounder and exposure is equivalent to the result first given by Cornfield et al 1959^7^ and Bross 1966^1^.  2. When the assumption that the relative risks between the unmeasured confounder and outcome among the unexposed and the exposed should be the same is violated, the formula in Lin et al 1998’s paper can be used.^6^ |

#### Rosenbaum and Rubin 1983: method for unmeasured confounding

| **Name/description** | **Method for unmeasured confounding** |
| --- | --- |
| **DOI** | 10.1111/j.2517-6161.1983.tb01242.x; https://doi.org/10.1093/ije/dyp332 |
| **Author and Year** | Original manuscript: Rosenbaum and Rubin 1983^8^.  Additional information: Groenwold et al 2010^2^. This review article provides a clear overview of the methods and was used to populate the cells below. |
| **Crossed-referenced in QBA textbook^3^** | Not referenced |
| **QBA classification** | Simple sensitivity analysis |
| **Applicable study design scenarios described^b^** | Any observational studies |
| **Effect measure of interest** | RR or OR |
| **Result of interest** | Corrected estimates |
| **Data structure/format of the eligible exposure/intervention^b^** | Categorical |
| **Data structure/format of the confounders, if applicable^b^** | Categorical |
| **Data structure/format of eligible outcome^b^** | Categorical |
| **Source(s) of bias addressed** | Unmeasured confounding (single) |
| **Information/parameters needed to conduct the QBA method** | - **P_X=0_**: proportion of the unexposed;  - **P_Z_**: prevalence of the unmeasured confounder;  - $\mathbf{OR}_{\boldsymbol{ZY}\boldsymbol{\mid}\boldsymbol{X=0}}$**,** $\mathbf{OR}_{\boldsymbol{ZY}\boldsymbol{\mid}\boldsymbol{X=1}}$: association between the unmeasured confounder and the outcome among unexposed and exposed, respectively;  - $\mathbf{OR}_{\mathbf{ZX}}$: association between the unmeasured confounder and the exposure.  *Notes: X: exposure; Y: outcome; Z: unmeasured confounder.* |
| **Mathematical formula** | The adjusted RR (or OR) is calculated with the risk for the outcome among the exposed ($R_{Y\mid X=1}$) and unexposed subjects ($R_{Y\mid X=0}$), i.e.,  RR = ($R_{Y\mid X=1}$/ $R_{Y\mid X=0}$).  The risk for the outcome given the exposure ($R_{Y\mid X}$) can be calculated using:  $R_{Y\mid X}=\frac{P_{Z=0}exp\left( \beta_{X}+ln\left( OR_{ZY\mid X} \right) \right)}{\left[ 1+exp\left( \beta_{X}+ln\left( OR_{ZY\mid X} \right) \right) \right]}+\frac{P_{Z=1}\left( exp\left( \beta_{X} \right)) \right.}{\left[ 1+exp\left( \beta_{X} \right) \right]}$.  The following steps need to be followed to generate$\beta_{X}$  Step 1: Solve the quadratic equation: $\begin{matrix} & \left[ P_{X=0}\cdot\mathrm{OR}_{ZX} \right]w^{2}+\left[ \left( P_{X=0}-P_{Z=0} \right)\mathrm{OR}_{\mathrm{ZX}} \right. \\ & \left. +P_{X=0}-1+P_{Z=0} \right]w+\left( P_{X=0}-1 \right)=0, \end{matrix}$  In which $w=exp(\Gamma)$. This quadratic equation has one positive solution, which provides $\Gamma$.  Step 2: Calculate the probability of absence of the unmeasured confounder, given exposure status, $P_{Z=0\mid X}$, by solving the equation for exposed and unexposed subjects (i.e., for the situations that X=0 and X=1):  $\begin{matrix} P_{Z=0\mid X}=P_{Z=0}/\left[ P_{Z=0}+P_{Z=1}\cdot exp\left( X\cdot ln\left( \mathrm{OR}_{ZX} \right) \right) \right. \\ \frac{1+\exp\left( \Gamma\right)}{\left( 1+\exp\left( \Gamma+\ln\left( \mathrm{OR}_{ZX} \right) \right) \right]}. \end{matrix}$  Step 3: For exposed and unexposed subjects (i.e., for X =1 and X =0), calculate the log(odds) for the absence of the outcome, given exposure status, $\beta_{X}=ln( w^{'})$, by solving the quadratic equation:  $\begin{matrix} & \left[ P_{X}\cdot\mathrm{OR}_{ZY\mid X} \right]{w'}^{2} \\ & +\left[ \left( P_{X}-P_{Z=0\mid X} \right)\mathrm{OR}_{ZY\mid X}+P_{X}-P_{Z=1\mid X} \right]w'+\left( P_{X}-1 \right)=0 \end{matrix}$  In which $P_{X}=P(Y=0\mid X=x),$ and $w'=exp\left( \beta_{X} \right)$. |
| **Data assumptions and additional required features** | Not mentioned |
| **Available and/or recommended software** | Not mentioned |
| **Output of the QBA analysis** | Corrected effect estimates |
| **Interpretation of the QBA analysis output** | Effect estimates accounting for unmeasured confounding |
| **Considerations related to the interpretation of the results** | Not mentioned |
| **Explicit discussions regarding the similarities and differences with other methods** | 1. The authors noted that compared to Cornfield et al 1959^7^ and Bross 1966^1^, this method has two major advantages: 1) it adjusts for measured covariates in addition to the unmeasured covariate; 2) unlike the previous methods, which investigated whether an unmeasured confounder can explain-away a finding, this method provides a bias-adjusted estimate.  2. The authors also noted that the approach outlined in Schlesselman et al 1978^5^ can be used as an alternative, but that it has limitations because it requires additional assumptions for unmeasured confounders.  3. Groenwold et al 2010^9^ noted that compared to the method outlined in Lin et al 1998,^6^ this method is more complicated but does not reply on rarity of outcomes to be applicable measure using in the estimation of ORs. They also noted that this method proposes to reduce measured confounders to stratification methods such as propensity scores. |

#### Flanders and Khoury: 1990 indirect assessment of confounding

| **Name/description** | **Indirect assessment of confounding** |
| --- | --- |
| **DOI** | 10.1097/00001648-199005000-00010 |
| **Author and Year** | Flanders and Khoury 1990^10^ |
| **Crossed-referenced in QBA textbook^3^** | Referenced and explained |
| **QBA classification** | Multidimensional analysis |
| **Applicable study design scenarios described^b^** | Cohort studies^b^ |
| **Effect measure of interest** | RR |
| **Result of interest** | Corrected estimate |
| **Data structure/format of the eligible exposure/intervention^b^** | Categorical |
| **Data structure/format of the confounders, if applicable^b^** | Categorical |
| **Data structure/format of eligible outcome^b^** | Categorical |
| **Source(s) of bias addressed** | Unmeasured confounding (single) |
| **Information/parameters needed to conduct the QBA method** | - **p_0_**: the prevalence of the covariate among the unexposed in the population;  - **OR_CE_**: the association between the exposure and the covariate;  - **RR_CD_**: the effect of the covariate on disease;  - **RR_Crude_**: the unadjusted association between the exposure and the outcome. |
| **Mathematical formula** | ${RR}_{Adjusted}=\frac{{RR}_{Crude}}{Bounding factor},$  Where the bounding factor is the **minimum** value of the following factors:  $\begin{matrix} \mathrm{OR}_{\mathrm{CE}}, \\ \mathrm{RR}_{\mathrm{CD}}, \\ \frac{1}{p_{0}}, \\ \frac{\mathrm{RR}_{\mathrm{CD}}}{\left[ \left( 1-p_{0} \right)+p_{0}\cdot\mathrm{RR}_{\mathrm{CD}} \right]}, \\ \frac{\mathrm{OR}_{\mathrm{CE}}}{\left[ \left( 1-p_{0} \right)+p_{0}\cdot\mathrm{OR}_{\mathrm{CE}} \right]}. \end{matrix}$ |
| **Data assumptions and additional required features** | 1. All relevant covariates have been identified and no other sources of bias or unidentified, residual confounding exist.  2. All bounding factors (including OR_CE_ and RR_CD_) must be greater than 1. When this is violated, one can consider recoding the variables.  3. The authors do not assume anything about the joint effects of confounder and exposure, such as additivity or multiplicativity. For RR or OR less than unity, different formulas can be used (see Appendix in the original paper^10^). |
| **Available and/or recommended software** | Not mentioned |
| **Output of the QBA analysis** | Corrected effect estimate |
| **Interpretation of the QBA analysis output** | The largest possible value for the RR due to confounding, assuming a valid bias model and given the bias parameters that are assigned values. |
| **Considerations related to the interpretation of the results** | Although results were derived for the RR in a cohort study, with proper modifications, the results extend directly to the incidence rate ratio in cohort studies or the ORs in case-cohort studies. |
| **Explicit discussions regarding the similarities and differences with other methods** | 1. The authors noted that the advantage of this method is that it requires specification of only one or two parameters, thus this method can be used even if accurate estimates for some of the relevant parameters are not available.  2. Fox et al 2021 noted that “this approach is most useful when little is known about the unmeasured confounder or when existing data on some of the factors (e.g., prevalence of the confounder in the exposed and unexposed groups) is believed not to apply to current study population.”^3^ |

#### Lin et al 1998: sensitivity analysis for unmeasured confounders for regression results

| **Name/description** | **Sensitivity analysis for unmeasured confounders for regression results** |
| --- | --- |
| **DOI** | PMID: 9750244 (DOI not found) |
| **Author and Year** | Lin et al 1998^6^ |
| **Crossed-referenced in QBA textbook^3^** | Referenced but not explained |
| **QBA classification** | Simple or multidimensional analysis |
| **Applicable study design scenarios described^b^** | Cohort or case-control studies |
| **Effect measure of interest** | RR, OR (when outcome is rare) or HR |
| **Result of interest** | Corrected estimate |
| **Data structure/format of the eligible exposure/intervention^b^** | Categorical |
| **Data structure/format of the confounders, if applicable^b^** | Categorical or continuous |
| **Data structure/format of eligible outcome^b^** | Categorical, time-to-event data, or continuous |
| **Source(s) of bias addressed** | Unmeasured confounding (single but can be generalized to multiple with modifications) |
| **Information/parameters needed to conduct the QBA method** | **For binary confounder,**  - $\boldsymbol{\Gamma}_{\mathbf{0}}\boldsymbol{,}\boldsymbol{\Gamma}_{\mathbf{1}}:$ the relative risks between the unmeasured confounder and outcome among the unexposed and exposed, respectively;  - **P_0_, P_1_**: the prevalence of the unmeasured confounder among the unexposed and exposed, respectively;  - **RR***: measured relative risk between exposure and outcome (with 95% CIs).  **For normally distributed confounder,**  - $\boldsymbol{\mu}_{\mathbf{0}}\boldsymbol{,}\boldsymbol{\mu}_{\mathbf{1}}\boldsymbol{:}$the mean values of unmeasured confounder among the unexposed and exposed, respectively;  - $\boldsymbol{\gamma}_{\mathbf{0}}\boldsymbol{, \gamma}_{\mathbf{1}}$: the beta coefficients between the unmeasured confounder and outcome among the unexposed and exposed, respectively;  - **RR***: measured relative risk between exposure and outcome (with 95% CIs). |
| **Mathematical formula** | **For binary confounder,**  $A=\frac{\Gamma_{1}P_{1}+\left( 1-P_{1} \right)}{\Gamma_{0}P_{0}+\left( 1-P_{0} \right)}$;  $RR=\frac{{RR}^{*}}{A}$.  **For normally distributed confounder,**  $RR={RR}^{*}/exp\left\{ \left( \gamma_{1}\mu_{1}-\gamma_{0}\mu_{0} \right)+0.5\left( \gamma_{1}^{2}-\gamma_{0}^{2} \right) \right\}$.  *Notes: formulas for survival data were presented in Section 3 of the original paper.* |
| **Data assumptions and additional required features** | 1. If the unmeasured confounder is a continuous variable, it should be normally distributed.  2. Conditional independence of unmeasured confounders and measured confounders given exposure is required. Alternatively, one can state the association between unmeasured confounders and measured confounders and incorporate that into the model (see Section 5 of the original manuscript).  3. Further simplification of the formula is possible, but the data assumptions also change (conditional independence of unmeasured confounders and measured confounders is no longer required, but the mean of an unmeasured confounder conditional on measured confounders and exposure ought to be additive in measured confounders and exposure). |
| **Available and/or recommended software** | Not mentioned |
| **Output of the QBA analysis** | Corrected effect estimates with 95% CIs |
| **Interpretation of the QBA analysis output** | Effect estimates accounting for unmeasured confounding |
| **Considerations related to the interpretation of the results** | When multiple unmeasured confounders are present, the author noted that “A more practical alternative is to simply treat U as a composite score constructed from a set of unmeasured confounders. In this regard, a normal U may represent a linear combination of several confounders and a binary U the dichotomy of high risk versus low risk determined by multiple risk factors.” ^6^ |
| **Explicit discussions regarding the similarities and differences with other methods** | 1. When $\Gamma_{0}=\Gamma_{1}$, the formula for binary confounder is the same as the formula (1) in Schlesselman 1978’s paper.^5^ Moreover, the formula is closely related to the bias analysis idea derived from Cornfield et al 1959^7^.  2. Groenwold et al 2010^9^ noted that compared to Rosenbaum and Rubin 1983^8^, Lin et al 1998’s method is easier to apply and more readily incorporates measured confounders but relies on the assumption of conditional independence of the unmeasured and measured confounders given exposure.  3. Vanderweele and Arah 2011^11^ noted that this method is less general than theirs, and that this method only applies to regression models. |

#### Greenland 2003: quantifying biases for classical confounding and (or) collider-stratification bias

| **Name/description** | **Quantifying biases for classical confounding and (or) collider-stratification bias** |
| --- | --- |
| **DOI** | 10.1097/01.EDE.0000042804.12056.6C |
| **Author and Year** | Greenland 2003^12^ |
| **Crossed-referenced in QBA textbook^3^** | Not referenced |
| **QBA classification** | Simple sensitivity analysis |
| **Applicable study design scenarios described^b^** | Any observational studies |
| **Effect measure of interest** | OR^b^ |
| **Result of interest** | Corrected estimate |
| **Data structure/format of the eligible exposure/intervention^b^** | Categorical |
| **Data structure/format of the confounders, if applicable^b^** | Categorical |
| **Data structure/format of eligible outcome^b^** | Categorical |
| **Source(s) of bias addressed** | Unmeasured confounding (single), selection bias |
| **Information/parameters needed to conduct the QBA method** | 1. **Classical confounding and Berksonian bias**:  - **crude OR** between E and D;  - **p**: p=P(C=1 \| D=E=0), the prevalence of C among the unexposed controls;  - **R_CD_, R_CE_**: the OR for C-D given E, for C-E given D (each assumed constant across strata of the given variable);  2. **M bias**:  - **R_AE_=R_AC_=R_BC_=R_BD_**: odds ratio of the association between A-E or A-C or B-C or B-D.  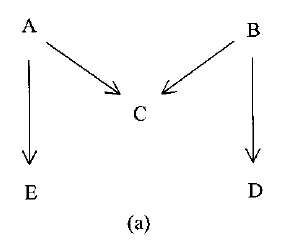  *Notes: A, B: variables on the E-D pathways as indicated in the picture; C: confounder/ collider; D: outcome; E: exposure.* |
| **Mathematical formula** | 1. **Classical confounding and Berksonian bias (selection bias)**:  $R_{ED}=crude OR/Bias\left( R_{CD},R_{CE},p \right)$,  where$\begin{matrix} & Bias\left( R_{CD},R_{CE},p \right)\equiv\left( R_{CD}R_{CE}p+1-p \right)/ \\ & \left( \left[ R_{CD}p+1-p \right]\left[ R_{CE}p+1-p \right] \right). \end{matrix}$  The authors noted that when p= 1/ (1+$\left( R_{CD}R_{CE} \right)^{1/2}$), the $Bias\left( R_{CD},R_{CE},p \right)$ reaches its maximum value. In addition, if $R_{CD}=R_{CE}$, $\mathrm{Bias} \left( R_{CD},R_{CE},p \right)$ is even larger.  2. **M bias**:  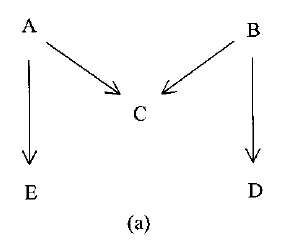  The lower bound of $R_{ED} is 16R(R+1)^{2}/\left( R^{2}+6R+1 \right)^{2}$, which is very close to 1, where $R_{AE}=R_{AC}=R_{BC}=R_{BD}=R$. |
| **Data assumptions and additional required features** | 1. **Classical confounding and Berksonian bias (selection bias)**:  R_CD_, R_CE_ and R_ED_ are assumed to be constant across strata of the given variable which is E, D, C respectively (i.e., for C-D given E and for C-E given D, for E-D given C, for).  Moreover, the author also noted that “To apply formula 1 to measure confounding by C, one must assume D=1 is rare in all C-E categories, or that the parameters refer to a case-cohort study in which the time of D is ignored.”^12^  2. **M bias**:  $R_{AE}=R_{AC}=R_{BC}=R_{BD}=R$.  *Notes: C: confounder/ collider; D: outcome; E: exposure.* |
| **Available and/or recommended software** | Not mentioned |
| **Output of the QBA analysis** | 1. **Classical confounding and Berksonian bias (selection bias)**: corrected effect estimate.  2. **M bias**: lower bound of $R_{ED}$. |
| **Interpretation of the QBA analysis output** | 1. **Classical confounding and Berksonian bias (selection bias)**: effect estimate accounting for the corresponding unmeasured confounding and (or) selection bias.  2. **M bias**: lower limit the magnitude of $R_{ED}$. |
| **Considerations related to the interpretation of the results** | 1. The author noted that if the prevalence of confounder is rare (i.e., under 5%) in all E-D categories, the R_ED_ would approximate the crude OR between E and D.  2. The author suggested that to avoid collider-stratification bias, one should not control variables affected by exposure or disease.  3. For M bias, the author noted that the lower bound of the OR between E and D should be very close to 1 unless the common effect R is very large. |
| **Explicit discussions regarding the similarities and differences with other methods** | Not mentioned |

#### Steenland and Greenland 2004: Monte Carlo sensitivity analysis and Bayesian analysis for unmeasured confounding

| **Name/description** | **Monte Carlo sensitivity analysis and Bayesian analysis for unmeasured confounding** |
| --- | --- |
| **DOI** | 10.1093/aje/kwh211 |
| **Author and Year** | Steenland and Greenland 2004^13^ |
| **Crossed-referenced in QBA textbook^3^** | Referenced but not explained |
| **QBA classification** | Bayesian analysis |
| **Applicable study design scenarios described^b^** | Cohort or case-control studies |
| **Effect measure of interest** | Rate ratio |
| **Result of interest** | Corrected estimates |
| **Data structure/format of the eligible exposure/intervention^b^** | Categorical |
| **Data structure/format of the confounders, if applicable^b^** | Categorical |
| **Data structure/format of eligible outcome^b^** | Categorical |
| **Source(s) of bias addressed** | Unmeasured confounding (single) |
| **Information/parameters needed to conduct the QBA method** | **Bayesian analysis:**  - the observed effect estimate (which adjusts for measured confounders but does not adjust for the unmeasured confounder);  Prior distributions needed (priors) for unknown parameters include:  - the exposure-outcome rate ratio after adjustment for the unmeasured confounder;  - the rate ratio for the effect of the unmeasured confounder on outcome (possibly from the literature);  - the estimated proportions of those with the confounder in exposed and nonexposed population (possibly from surveys).  **Monte Carlo sensitivity analysis:**  - the proportions of different confounder levels in the exposed and nonexposed populations;  - the exposure-outcome rate ratios within different confounder levels. |
| **Mathematical formula** | **Bayesian analysis:**  Step 1: Specify prior distributions (priors) for unknown parameters  Step 2: Construct a model for the probability of the data given these parameters (i.e., the likelihood function).  Step 3: Combine the priors for unknown parameters with the probability of the observed data, using Bayes’ theorem, to produce a posterior distribution for the parameter of interest (the bias-adjusted rate ratio).  **Monte Carlo sensitivity analysis:**  The procedure for **Monte Carlo sensitivity analysis** is very similar to **Bayesian analysis**. The major difference, as noted by the authors, is that for Monte Carlo sensitivity analysis, “a distribution for the parameter of interest is generated on the basis of repeated sampling from priors followed by adjustment, rather than sampling directly from a posterior distribution as in Bayesian analysis.”^13^ By adjustment, the authors mean dividing the observed (unadjusted) rate ratio by a bias factor which represents the confounding effect of the unmeasured confounder. The bias factor is a function of the proportions of participants with the confounder in the exposed and nonexposed populations as well as the rate ratio for the effect of unmeasured confounder on the outcome.  Bias $={RR}_{\exp}/RR_{\text{nonexp}}$. |
| **Data assumptions and additional required features** | The effect estimates do not vary across confounder levels. |
| **Available and/or recommended software** | WinBUGS (code in Appendix 2 of the original manuscript) |
| **Output of the QBA analysis** | Corrected effect estimates |
| **Interpretation of the QBA analysis output** | Effect estimate accounting for unmeasured confounding |
| **Considerations related to the interpretation of the results** | The authors noted that Monte Carlo sensitivity analysis and Bayesian analysis produce similar bias-adjusted results. |
| **Explicit discussions regarding the similarities and differences with other methods** | 1. The authors noted the difference and similarities between Monte Carlo sensitivity analysis and Bayesian analysis as follows: “In situations where the prior information is less precise, the bias distribution will lead to substantially wider Monte Carlo and Bayesian intervals relative to the conventional confidence intervals (which reflect only random error). Monte Carlo sensitivity analysis and Bayesian bias analysis will also produce much wider intervals when the conventional confidence intervals are narrow, as in large studies, pooling projects, and meta-analyses.”^13^ Moreover, they also noted that the shift (or location) in the final distribution can also determine the final results.  2. The authors noted that this bias analysis method has the advantage of explicitly and quantifiably accounting for likely sources of bias. |

#### Cabral 2007: integration of Rosenbaum and Greenland method for confounding

| **Name/description** | **Integration of Rosenbaum and Greenland method for confounding** |
| --- | --- |
| **DOI** | 10.1017/S0950268807008394;  10.1590/s0034-89102007000300017. |
| **Author and Year** | Cabral 2007^14,15^ |
| **Crossed-referenced in QBA textbook^3^** | Not referenced |
| **QBA classification** | Multidimensional analysis |
| **Applicable study design scenarios described^b^** | Any observational study designs |
| **Effect measure of interest** | RR or OR |
| **Result of interest** | Corrected estimates |
| **Data structure/format of the eligible exposure/intervention^b^** | Categorical or continuous |
| **Data structure/format of the confounders, if applicable^b^** | Categorical |
| **Data structure/format of eligible outcome^b^** | Categorical or continuous |
| **Source(s) of bias addressed** | Unmeasured confounding (single) |
| **Information/parameters needed to conduct the QBA method** | **Rosenbaum approach**:  - Total exposed individuals with the outcome;  - Total exposed individuals without the outcome;  - Total non-exposed individuals without the outcome;  - Total non-exposed individuals not without the outcome;  - **Γ Value**: the magnitude representing the effects of the unmeasured confounding variable on the exposure variable considered, which makes the observed association between exposure and outcome statistically non-significant.  **Greenland approach**:  - Speculated value of the OR between the outcome and the confounder;  - Proportion of the confounder Z among exposed individuals;  - Speculated value of the OR between the exposure and the confounder. |
| **Mathematical formula** | The idea is to use the Rosenbaum approach first, which can help identify the minimum value of Γ value. After the Γ value is determined, the Greenland approach is employed. Although no formulas were explicitly present by both papers, Table 3 and Table 4 indirectly presented some calculations.^15^ |
| **Data assumptions and additional required features** | Not mentioned |
| **Available and/or recommended software** | An Excel spreadsheet.^15^ |
| **Output of the QBA analysis** | A series of corrected ORs or RRs and corresponding 95% CIs. |
| **Interpretation of the QBA analysis output** | Corrected ORs or RRs under different settings (i.e., different magnitudes of the association between confounder and exposure, and the association between confounder and outcome.) |
| **Considerations related to the interpretation of the results** | 1. The authors noted that “Although a sensitivity analysis neither confirms nor eliminates the presence of an unmeasured confounder, it may be used by the researcher as a quantitative assessment that can be integrated with the analysis carried out during the validation of findings stage, particularly in observational studies.”^14^  2. The authors noted that the Greenland method approach focuses more on the epidemiological elements of the study, while the Rosenbaum method addresses the statistical significance of the findings observed. |
| **Explicit discussions regarding the similarities and differences with other methods** | 1. The original paper that proposed the method was a dissertation paper: *Cabral MDB. Sensitivity analysis in epidemiological*  *studies [Dissertation]. Rio de Janeiro, Brasil: Universidade Federal do Rio de Janeiro, 2005, 69 pp.*  2. The Greenland 1996^16^ approach was also presented separately in the table. |

#### Arah et al 2008: method for unmeasured confounders

| **Name/description** | **Method for unmeasured confounders** |
| --- | --- |
| **DOI** | 10.1016/j.annepidem.2008.04.003 |
| **Author and Year** | Arah et al 2008^17^ |
| **Crossed-referenced in QBA textbook^3^** | Referenced and explained |
| **QBA classification** | Simple or multidimensional sensitivity analysis |
| **Applicable study design scenarios described^b^** | Any observational study designs |
| **Effect measure of interest** | RR, OR, or RD |
| **Result of interest** | Corrected estimate |
| **Data structure/format of the eligible exposure/intervention^b^** | Categorical |
| **Data structure/format of the confounders, if applicable^b^** | Categorical |
| **Data structure/format of eligible outcome^b^** | Categorical |
| **Source(s) of bias addressed** | Unmeasured confounding (single) |
| **Information/parameters needed to conduct the QBA method** | - ${\boldsymbol{P}_{\mathbf{21}}\mathbf{,}\boldsymbol{P}}_{\mathbf{20}}\mathbf{,}\boldsymbol{P}_{\mathbf{2+}}$: prevalences of confounder among the exposed, unexposed, and total population, respectively.  Then, depending on the effect measure that one uses:  - $\boldsymbol{R}\boldsymbol{D}_{\boldsymbol{dxz}\left( \boldsymbol{T} \right)}$: risk difference for Z-stratified exposure-outcome relations among the total population;  -$\boldsymbol{RR}_{\boldsymbol{dxz}\mathbf{(}\boldsymbol{T}\mathbf{)}}$: risk ratio for Z-stratified exposure-outcome relations among the total population;  -$\boldsymbol{OR}_{\boldsymbol{dxz}\mathbf{(}\boldsymbol{T}\mathbf{)}}$: odds ratio for Z-stratified exposure-outcome relations among the total population.  *Note: D: disease or outcome; T: the total population; X: exposure; Z: unmeasured confounder.* |
| **Mathematical formula** | The formula depends on the effect measure that one uses. The formula here is for total population, and formulas for other target populations (i.e., exposed, and unexposed) can be found in the original manuscript:  $\begin{matrix} \boldsymbol{R}\boldsymbol{D}_{\boldsymbol{dx}\mathbf{+}}=RD_{dxz\left( T \right)}+ & RD_{d21}\left( P_{21}-P_{2+} \right) \\ & -RD_{d20}\left( P_{20}-P_{2+} \right) \end{matrix}$;  $\boldsymbol{R}\boldsymbol{R}_{\boldsymbol{dx}\mathbf{+}}={RR}_{dxz(T)}*\left( \frac{RR_{d21}P_{21}+1-P_{21}}{RR_{d21}P_{2+}+1-P_{2+}} \right)/\left( \frac{RR_{d20}P_{20}+1-P_{20}}{RR_{d20}P_{2+}+1-P_{2+}} \right)$;  $\boldsymbol{O}\boldsymbol{R}_{\boldsymbol{dx}\mathbf{+}}=OR_{dxz(T)}*\left( \frac{OR_{d21}p_{21}+1-P_{21}}{OR_{d21}p_{2+}+1-P_{2+}} \right)/\left( \frac{OR_{d20}p_{20}+1-p_{20}}{OR_{d20}p_{2+}+1-p_{2+}} \right)$.  *Note: D: disease or outcome; T: the total population; X: exposure; Z: unmeasured confounder. The + is used to represent summarizing over a variable or the applicable subscript;* ${RD}_{dx+}, RR_{dx+}, OR_{dx+}$*: adjusted risk difference, risk ratio or odds ratio; for example, in* $RD_{dxz\left( T \right)}$ *z = Z-stratified, X=1 exposed, X=0 is unexposed.* |
| **Data assumptions and additional required features** | When the effect measure of interest is an OR, the authors noted that “the odds ratios are estimating risk ratios (as when the controls are selected from an underlying cohort without respect to disease status, or when the outcome is rare at all exposure-confounder levels)”.^17^ |
| **Available and/or recommended software** | Not mentioned |
| **Output of the QBA analysis** | Corrected effect estimate |
| **Interpretation of the QBA analysis output** | Effect estimate accounting for unmeasured confounding |
| **Considerations related to the interpretation of the results** | The authors mentioned that prevalence of confounder among the exposed, unexposed, and the effect measure (i.e., RD) can be generated from external sources. |
| **Explicit discussions regarding the similarities and differences with other methods** | The authors noted that the formulas for RR is equivalent to the formulas derived by Flanders and Khoury 1990.^10^ |

#### Lash et al 2010: probabilistic bias analysis for two biases

| **Name/description** | **Probabilistic bias analysis for two biases** |
| --- | --- |
| **DOI** | 10.1002/pds.1938 |
| **Author and Year** | Lash et al 2010^18^ |
| **Crossed-referenced in QBA textbook^3^** | Referenced and explained |
| **QBA classification** | Probabilistic analysis |
| **Applicable study design scenarios described^b^** | Any observational study designs with multiple covariates (examples provided are case-control studies) |
| **Effect measure of interest** | RR, OR, HR, and rate ratio |
| **Result of interest** | Corrected estimate |
| **Data structure/format of the eligible exposure/intervention^b^** | Categorical |
| **Data structure/format of the confounders, if applicable^b^** | Categorical |
| **Data structure/format of eligible outcome^b^** | Categorical, or time-to-event data |
| **Source(s) of bias addressed** | Outcome misclassification (differential or nondifferential) (in **Method 1**), or unmeasured confounding (single) (in **Method 2**) |
| **Information/parameters needed to conduct the QBA method** | -$\hat{\mathbf{RR}}$or $\hat{\mathbf{OR}}$**:** observed RR or OR  **Method 1**:  - the number of observed cases;  - $\boldsymbol{S}_{\boldsymbol{i=1, j}} \mathrm{and} \boldsymbol{S}_{\boldsymbol{i=0, j}}$: distribution of sensitivities *S* of the exposed and unexposed respectively (the paper used beta distribution as the example);  **Method 2**:  - **OR_DZ_**: distribution of the association between the confounder and the outcome  - ***p*_Z, X=1_** or ***p*_Z, X=0_**: distribution of prevalence of the confounder in the exposed and the unexposed respectively.  *Note: D: disease; X: exposure; Z: confounder.* |
| **Mathematical formula** | **Method 1:** For disease misclassification:  $RR=\frac{\hat{RR}}{\frac{S_{i=1, j}}{S_{i=0, j}}}$;  **Method 2:** For unmeasured confounding:  $OR=\frac{\hat{OR}}{{RR}_{C}} ;{RR}_{C}=\frac{\mathrm{OR}_{DZ}\mathrm{OR}_{XZ}p_{Z,X=0}+1-p_{Z,X=0}}{\left( \mathrm{OR}_{DZ}p_{Z,X=0}+1-p_{Z,X=0} \right)\left( \mathrm{OR}_{XZ}p_{Z,X=0}+1-p_{Z,X=0} \right)}$;  $\mathrm{OR}_{XZ}$= [$p_{Z,X=1}$ (1- $p_{Z,X=0}$)]/[ $p_{Z,X=0}$ (1- $p_{Z,X=1}$)].  *Note: i: exposure status; j: stratum; OR: true odds ratio; RR: true relative risk;* ${RR}_{C}$*: relative risk due to confounding; S: sensitivity. Formula for 95% CIs were presented in the paper (equation (6)).* |
| **Data assumptions and additional required features** | 1. The bias is independent of confounding by measured variables, or requires that the dependence be incorporated into the bias model.  2. For disease misclassification specifically: perfect specificity of disease classification (i.e., presumably much greater than 95%). |
| **Available and/or recommended software** | SAS code available in the supplements |
| **Output of the QBA analysis** | Correct OR (95% CIs); corrected RR (95% CIs) |
| **Interpretation of the QBA analysis output** | OR adjusted for misclassification of the outcome, and/or unmeasured confounding |
| **Considerations related to the interpretation of the results** | 1.The authors mentioned that they proposed a solution to the assumption of the independence of the bias source from adjustments for measured covariates. The solution integrates the bias analysis results with a prior which is hard to parameterize. Therefore, it is critical that the analyst clearly explains the justification for the assigned values.  2. The two sensitivity parameters S can be drawn from their own beta distributions. |
| **Explicit discussions regarding the similarities and differences with other methods** | Earlier work focused on crude estimates,^1,7^ however application of these methods to estimates adjusted for multiple confounders is more difficult. Three solutions have been previously proposed,^19^ including using stratification, record level data, and empirical or Bayesian methods, but they have their own limitations as well. Unlike previous methods, this method does not resort to stratification, record-level corrections, empirical, or Bayesian methods. |

#### Chiba 2011: sensitivity analysis for unmeasured confounding of attributable fraction

| **Name/description** | **Sensitivity analysis for unmeasured confounding of attributable fraction** |
| --- | --- |
| **DOI** | 10.1097/EDE.0b013e31823b6409 |
| **Year** | Chiba 2011^20^ |
| **Crossed-referenced in QBA textbook^3^** | Not referenced |
| **QBA classification** | Simple or multidimensional sensitivity analysis |
| **Applicable study design scenarios described^b^** | Any observational study designs |
| **Effect measure of interest** | Attributable fraction (also known as the attributable risk and excess fraction) |
| **Result of interest** | Corrected estimate |
| **Data structure/format of the eligible exposure/intervention^b^** | Categorical |
| **Data structure/format of the confounders, if applicable^b^** | Categorical or continuous |
| **Data structure/format of eligible outcome^b^** | Categorical |
| **Source(s) of bias addressed** | Unmeasured confounding (single) |
| **Information/parameters needed to conduct the QBA method** | - **AF^O^**: the initial estimate adjusted only for X;  - **δ**: sensitivity parameter; among the strata X=x, the average difference in potential outcomes Y_0_ between those exposed and those unexposed;  - **Pr (Y=1)**: disease prevalence;  - **Pr (A=1)**: exposure prevalence.  *Notes: A: exposure status; AF: attributable fraction; X: measured confounder(s); Y: observed outcome; Y_0_: potential outcome if unexposed.* |
| **Mathematical formula** | $AF={AF}^{O}-\frac{Pr(A=1)}{Pr(A=0)}\delta$  *Notes: A: exposure status; AF: attributable fraction; X: measured confounder(s); Y: observed outcome; Y_0_: potential outcome if unexposed.* |
| **Data assumptions and additional required features** | δ does not vary between the strata of measured confounders X. |
| **Available and/or recommended software** | Not mentioned |
| **Output of the QBA analysis** | Corrected estimate |
| **Interpretation of the QBA analysis output** | Attributable fraction accounting for unmeasured confounding |
| **Considerations related to the interpretation of the results** | 1. The authors noted that “AF estimate adjusted only for measured confounders will give a good approximation to the true AF, if Pr(A = 1) is small and Pr(Y = 1) is large, even when unmeasured confounders exist.”^20^  2. The sensitivity parameter δ is set by the investigator according to what is thought to be plausible, which can be single or multiple value(s).  3. This method does not assume that the effect estimate has been adjusted for measured confounders. Therefore, the author noted that other causal inference methods, such as using doubly robust estimation, can be used. |
| **Strengths and limitations identified in studies comparing or commenting on QBA methods** | Not mentioned, but the concept of δ was derived from another paper.^21^ |

#### VanderWeele and Arah 2011: method for unmeasured confounding for general outcomes, treatments, and confounders

| **Name/description** | **Method for unmeasured confounding for general outcomes, treatments, and confounders** |
| --- | --- |
| **DOI** | 10.1097/EDE.0b013e3181f74493 |
| **Author and Year** | VanderWeele and Arah 2011^11^ |
| **Crossed-referenced in QBA textbook^3^** | Referenced but not explained |
| **QBA classification** | Simple sensitivity analysis |
| **Applicable study design scenarios described^b^** | Any observational study designs |
| **Effect measure of interest** | MD, RR, or OR |
| **Result of interest** | Corrected estimate |
| **Data structure/format of the eligible exposure/intervention^b^** | Categorical or continuous |
| **Data structure/format of the confounders, if applicable^b^** | Categorical or continuous |
| **Data structure/format of eligible outcome^b^** | Categorical or continuous |
| **Source(s) of bias addressed** | Unmeasured confounding (single) |
| **Information/parameters needed to conduct the QBA method** | **-** $E\left( Y\mid a_{1},x,u \right)-E\left( Y\mid a_{1},x,u^{'} \right), and E\left( Y\mid a_{0},x,u \right)-E\left( Y\mid a_{0},x,u^{'} \right):$the relation between U and Y, among those with treatment level A = a_1_ and A = a_0_, within each stratum of X;  - $\left\{ P\left( u\mid a_{1},x \right) \right.-P(u\mid x)\}, \{\left. P\left( u\mid a_{0},x \right)-P(u\mid x) \right\}$ : how the distribution of the unmeasured confounder U among those with treatment level A = a_1_ and A = a_0_ compares with the overall distribution of U, within each stratum of measured confounders X.  *Notes: A: exposure; X: measured confounders; U: unmeasured confounders; Y: outcome. The variable with lowercase letter refers to the actual value (e.g., u).* |
| **Mathematical formula** | $d_{a_{+}}=\sum_{x} \sum_{u} \left\{ E\left( Y\mid a_{1},x,u \right)-E\left( Y\mid a_{1},x,u^{'} \right) \right\}\left\{ P\left( u\mid a_{1},x \right) \right.-P(u\mid x)\}P(x)-\sum_{x} \sum_{u} \left\{ E\left( Y\mid a_{0},x,u \right)-E\left( Y\mid a_{0},x,u^{'} \right) \right\}\left. \{P\left( u\mid a_{0},x \right)-P(u\mid x) \right\}P(x)$; $d_{a_{1}}=\sum_{x} \sum_{u} \left\{ E\left( Y\mid a_{0},x,u \right)-E\left( Y\mid a_{0},x,u^{'} \right) \right\}\left\{ P\left( u\mid a_{1},x \right) \right.\left. -P\left( u\mid a_{0},x \right) \right\}P\left( x\mid a_{1} \right)$; $d_{a_{0}}=\sum_{x} \sum_{u} \left\{ E\left( Y\mid a_{1},x,u \right)-E\left( Y\mid a_{1},x,u^{'} \right) \right\}\left\{ P\left( u\mid a_{1},x \right) \right.\left. -P\left( u\mid a_{0},x \right) \right\}P\left( x\mid a_{0} \right)$.  *Note: the formulas apply to effect measure of MD; the formulas for other measures of effect can be found in the paper.* $d_{a_{+}}{, d}_{a_{1}}{, d}_{a_{0}}$*represent relevant bias (i.e.., the difference between the observed average outcome differences, adjusted for X, and*  *the true causal effect) when the target population is the total group, or those exposed to a_1_ or a_0_, respectively; u’ is any chosen reference value for the unmeasured confounder U.* |
| **Data assumptions and additional required features** | 1. Y_a_⫫A\|X, U.  2. If one makes simplifying assumption, for instance, that the relationship between U and Y, ie, $E\left( Y\mid a,x,u \right)-E\left( Y\mid a,x,u^{'} \right),$ does not vary between strata of A then the expression for $d_{a_{+}}$ simplifies to  $\begin{matrix} d_{a_{+}}=\sum_{x} \sum_{u} \left\{ E\left( Y \mid a,x,u \right)-E\left( Y\mid a,x,u^{'} \right) \right\} & \left\{ P\left( u\mid a_{1},x \right) \right. \\ & \left. -P\left( u\mid a_{0},x \right) \right\}P\left( x \right). \end{matrix}$  And it can be further simplified with more assumptions (see details in the paper). 3. This method does not assume the unmeasured confounders are independent of the measured confounders. |
| **Available and/or recommended software** | Not mentioned |
| **Output of the QBA analysis** | Bias terms (i.e., $d_{a_{+}}{, d}_{a_{1}}{, d}_{a_{0}}$) |
| **Interpretation of the QBA analysis output** | The difference between the observed average outcome differences, adjusted for X, and the true causal effect. One can use the observed MD minus the bias term to obtain the true MD. Similarly, the bias terms can be derived for RR or OR (see details in the paper). |
| **Considerations related to the interpretation of the results** | 1. The results do not presuppose a particular functional form relating the outcome and the observed covariates and treatment.  2. This method is applicable irrespective of how the initial adjusted estimates are obtained. |
| **Explicit discussions regarding the similarities and differences with other methods** | The authors noted that their formula can be used to replicate the results in Rosenbaum and Rubin 1983^8^. Moreover, the authors noted that the formula from VanderWeele and Arah 2011^11^ is more general since the approach from Rosenbaum and Rubin 1983^8^ was restricted to binary outcomes and effect measures (ORs). Moreover, the authors also noted that the VanderWeele and Arah 2011^11^ approach is more general then the Lin et al 1998^6^ one, since it does not require a regression context and does not have the no interaction assumption. |

#### McCandless 2012: Bayesian method for unmeasured confounding in meta-analyses

| **Name/description** | **Bayesian method for unmeasured confounding in meta-analyses** |
| --- | --- |
| **DOI** | 10.2202/1557-4679.1350 |
| **Year** | McCandless 2012^22^ |
| **Crossed-referenced in QBA textbook^3^** | Not referenced |
| **QBA classification** | Probabilistic analysis |
| **Applicable study design scenarios described^b^** | Meta-analyses of cohort, case-control, or cross-sectional studies |
| **Effect measure of interest** | Log RRs, log ORs, or log HRs (that have been adjusted for a set of confounders) |
| **Result of interest** | Corrected estimate |
| **Data structure/format of the eligible exposure/intervention^b^** | Categorical |
| **Data structure/format of the confounders, if applicable^b^** | Categorical |
| **Data structure/format of eligible outcome^b^** | Categorical, or time-to-event data |
| **Source(s) of bias addressed** | Unmeasured confounding (single) |
| **Information/parameters needed to conduct the QBA method** | - $\boldsymbol{\theta}_{\boldsymbol{j}}$: log relative risks for the association between the dichotomous exposure and disease in study j, adjusted for measured covariates in study j, and additionally, adjusted for a single binary unmeasured confounder. $\theta_{j}\overset{\text{ IID }}{\sim}N\left( \mu,\tau^{2} \right)$ for $j\in1:k$, as independent and identically normally distributed with mean $\mu$ and standard deviation τ.  - **RR_j_**: the relative risk for the association between the dichotomous unmeasured confounder and the outcome in study j, conditional on the exposure and measured covariates. $logRR_{j}\overset{\mathrm{IID}}{\sim}N\left( \mu_{RR},\tau_{RR}^{2} \right)$ for $j\in1:k$ as independent and identically normally distributed with mean $\mu_{RR}$ and standard deviation $t_{RR}.$  - **p0_j_** and **p1_j_**: the prevalence of the unmeasured confounder in unexposed versus exposed individuals in the j^th^ study.$\begin{matrix} & \mathrm{logit} \left( p_{0j} \right)\overset{\text{ IID }}{\sim}N\left( \mu_{p_{0}},\tau_{p_{0}}^{2} \right), \\ & logit\left( p_{1j} \right)\overset{\text{ IID }}{\sim}N\left( \mu_{p_{1}},\tau_{p_{1}}^{2} \right), \end{matrix}$  for $j\in1:k$ as independent and identically normally distributed with means $\mu_{p0}$, $\mu_{p1}$ and standard deviations $t_{po}, t_{p1},\mathrm{respectively}.$ |
| **Mathematical formula** | Procedures:  1. Run a preliminary analysis using a Bayesian random-effect model that ignores unmeasured confounding. One example method was provided^23^ and more details can be found in section 2.3 and 3.1 in the original manuscript.  2. Next, extend the analysis by incorporating uncertainty about unmeasured confounding. In particular, specify the priors and their distribution. The priors are hyperparameters $\left( \mu_{RR},\tau_{RR} \right),\left( \mu_{p_{0}},\tau_{p_{0}} \right)$ and $\left( \mu_{p_{1}},\tau_{p_{1}} \right)$. This can be accomplished by reading literature.  3. Then, the posterior distribution can be derived (key formula as follows):  $P\left( \boldsymbol{\theta},\boldsymbol{RR},\boldsymbol{p}_{\mathbf{1}},\boldsymbol{p}_{\mathbf{0}},\mu,\tau^{2}\mid\boldsymbol{y},\boldsymbol{\sigma} \right)\propto\prod_{j=1}^{k} exp\left[ -\frac{1}{2\sigma_{i}^{2}}\left\{ y_{i}-\left( \theta_{j}+\Omega\left( RR_{j},p_{1j},p_{0j} \right) \right) \right\}^{2} \right]\times$ (7) $P\left( \theta_{i}\mid\mu,\tau\right)P\left( RR_{j} \right)P\left( p_{1j} \right)P\left( p_{0j} \right)\times$ (8) $P(\mu)P(\tau)$,(9)  where equation $(7)$ is the likelihood function, equation $(8)$ is the prior distributions on $\left( \theta_{j},RR_{j},p_{0j},p_{1j} \right)$, and equation (9) is the prior on the hyperparameters $(\mu,\tau)$.  4. Lastly, run the Bayesian random-effect model again (key formula as follows).  $\theta_{j}\mid RR_{j},p_{1j},p_{0j},y_{j},\sigma_{j}\sim N\left( \frac{\tau^{2}\left\{ y_{i}-\Omega\left( RR_{j},p_{1j},p_{0j} \right) \right\}+\sigma_{j}^{2}\mu}{\sigma_{j}^{2}+\tau^{2}},\frac{\sigma_{j}^{2}\tau^{2}}{\sigma_{j}^{2}+\tau^{2}} \right)$. As noted by the author “This equation says that, given the bias parameters (RR_j_, p_0j_, p_1j_) and data (y_j_ , σ_j_ ), the posterior distribution of θ_j_ is normal with mean that is a weighted average of the prior mean of θ_j_, which is µ, and the bias-corrected exposure effect estimate, which is y_i_−Ω(RR_j_ , p_1j_, p_0j_).”^22^ |
| **Data assumptions and additional required features** | 1. The effect estimates as the basic unit of analysis are assumed to be roughly normally distributed with known standard error.  2. Each estimate y_j_ has been adjusted for a set of covariates that were measured and available for analysis in the j^th^ study.  3. The author noted that the standard error of the bias-corrected point estimator is not affected by unmeasured confouding. In other words, when there is unmeasured confounding in an observational study, then the location of the exposure effect estimate will shift, however its standard error will remain constant.  4. The unmeasured confounder does not interact with the exposure. |
| **Available and/or recommended software** | The author noted that the R code is available on their website (not presented). |
| **Output of the QBA analysis** | Corrected summary RR |
| **Interpretation of the QBA analysis output** | Summary RR which adjusts for the unmeasured confounding |
| **Considerations related to the interpretation of the results** | 1. The author highlighted the importance of correct specification of priors. They further illustrated that if the investigator can correctly determine the true distribution of bias from unmeasured confounding, then the Bayesian procedure will give interval estimates for the exposure effect with roughly nominal 95% coverage probability.  2. The author noted that the bias-corrected estimate $\theta_{j}$ does not necessarily have a causal interpretation, unless there are no additional unmeasured confounders other than the unmeasured and measured confounders that have been adjusted for.  3. The amount of unmeasured confounding in any particular study is independent of the exposure effect.  4. Lambert et al 2005 found that when the number of studies is small (≤ 5) then the prior distribution tends to greatly affects precision of the posterior distribution of $\mu$.^24^ |
| **Explicit discussions regarding the similarities and differences with other methods** | This method built upon the framework from Lin et al 1998.^6^ In particular, this method used the same algebraic adjustment formula from Lin et al 1998.^6^ |

#### VanderWeele et al 2012: sensitivity analysis for interactions under unmeasured confounding

| **Name/description** | **Sensitivity analysis for interactions under unmeasured confounding** |
| --- | --- |
| **DOI** | 10.1002/sim.4354 |
| **Author and Year** | VanderWeele et al 2012^25^ |
| **Crossed-referenced in QBA textbook^3^** | Not referenced |
| **QBA classification** | Simple sensitivity analysis |
| **Applicable study design scenarios described^b^** | Observational study design with interaction estimates:  1. Additive interaction: cohort studies  2. Multiplicative interaction and the relative excess risk due to interaction (RERI): cohort and case-control when the outcome is rare |
| **Effect measure of interest** | RR, OR, or relative excess risk |
| **Result of interest** | Corrected estimate |
| **Data structure/format of the eligible exposure/intervention^b^** | Categorical or continuous |
| **Data structure/format of the confounders, if applicable^b^** | Categorical |
| **Data structure/format of eligible outcome^b^** | Categorical or continuous |
| **Source(s) of bias addressed** | Unmeasured confounding for interactions (single or multiple) |
| **Information/parameters needed to conduct the QBA method** | **1. Interactions on the additive scale:**  - **additive interaction measure**:  $\mathbb{E}\left[ Y\mid g_{1},e_{1},c \right]-\mathbb{E}\left[ Y\mid g_{0},e_{1},c \right]-\mathbb{E}\left[ Y\mid g_{1},e_{0},c \right]+\mathbb{E}\left[ Y\mid g_{0},e_{0},c \right]$  - $\boldsymbol{\delta}_{\mathbf{1}}:$For E=e_1_, prevalence difference of U comparing G=g_1_ and G=g_0,_ $\delta_{1}=P\left( U=1\mid g_{1},e_{1},c \right)-P\left( U=1\mid g_{0},e_{1},c \right);$  - $\boldsymbol{\delta}_{\mathbf{0}}:$For E=e_0_, prevalence difference of U comparing G=g_1_ and G=g_0_, $\delta_{0}=P\left( U=1\mid g_{1},e_{0},c \right)-P\left( U=1\mid g_{0},e_{0},c \right);$  - $\boldsymbol{\gamma}_{\mathbf{1}}:$ For E=e_1_, the effect of U, $\gamma_{1}\mathbb{=E}\left[ Y\mid g,e_{1},c,U=1 \right]-\mathbb{E}\left[ Y\mid g,e_{1},c,U=0 \right]$;  - $\boldsymbol{\gamma}_{\mathbf{0}}:$ For E=e_0_, the effect of U, $\gamma_{0}=\mathbb{E}\left[ Y\mid g,e_{0},c,U=1 \right]-\mathbb{E}\left[ Y\mid g,e_{0},c,U=0 \right]$.  **2. Interactions on the multiplicative scale:**  - **multiplicative interaction measure:** $\frac{\mathbb{E}\left[ Y\mid g_{1},e_{1},c \right]}{\mathbb{E}\left[ Y\mid g_{0},e_{1},c \right]}/\frac{\mathbb{E}\left[ Y\mid g_{1},e_{0},c \right]}{\mathbb{E}\left[ Y\mid g_{0},e_{0},c \right]}$;  - $\boldsymbol{\gamma}_{\mathbf{1}}:$ For E=e_1_, the effect of U, $\gamma_{1}\mathbb{=E}\left[ Y\mid g,e_{1},c,U=1 \right]-\mathbb{E}\left[ Y\mid g,e_{1},c,U=0 \right]$;  - $\boldsymbol{\gamma}_{\mathbf{0}}:$ For E=e_0_, the effect of U, $\gamma_{0}=\mathbb{E}\left[ Y\mid g,e_{0},c,U=1 \right]-\mathbb{E}\left[ Y\mid g,e_{0},c,U=0 \right]$;  - $\boldsymbol{P}\left( \boldsymbol{U}\boldsymbol{=1\mid}\boldsymbol{g}_{\mathbf{i}}\mathbf{,}\boldsymbol{e}_{\mathbf{i}}\mathbf{,}\boldsymbol{c} \right):$prevalence of U among E=e_i_ and G=g_i_.  **3. Relative excess risk due to interaction (RERI):**  - **RERI**: $\frac{\mathbb{E}\left[ Y\mid g_{1},e_{1},c \right]}{\mathbb{E}\left[ Y\mid g_{0},e_{0},c \right]}-\frac{\mathbb{E}\left[ Y\mid g_{1},e_{0},c \right]}{\mathbb{E}\left[ Y\mid g_{0},e_{0},c \right]}-\frac{\mathbb{E}\left[ Y\mid g_{0},e_{1},c \right]}{\mathbb{E}\left[ Y\mid g_{0},e_{0},c \right]}+1$;  - $\boldsymbol{\gamma}_{\mathbf{1}}:$ For E=e_1_, the effect of U, $\gamma_{1}\mathbb{=E}\left[ Y\mid g,e_{1},c,U=1 \right]-\mathbb{E}\left[ Y\mid g,e_{1},c,U=0 \right]$;  - $\boldsymbol{\gamma}_{\mathbf{0}}:$ For E=e_0_, the effect of U, $\gamma_{0}=\mathbb{E}\left[ Y\mid g,e_{0},c,U=1 \right]-\mathbb{E}\left[ Y\mid g,e_{0},c,U=0 \right]$;  - $\boldsymbol{P}\mathbf{(}\boldsymbol{U}\boldsymbol{=1\mid}\boldsymbol{c}\mathbf{)}$: prevalence of U;  - $\boldsymbol{P}\left( \boldsymbol{U}\boldsymbol{=1\mid}\boldsymbol{e}_{\mathbf{i}}\mathbf{,}\boldsymbol{c} \right):$ prevalence of U among E=e_i_.  *Notes: G and E are two factors or exposures of interest (which can be genetic and environmental factors, or two genetic factors or two environmental factors); e_i_: status of exposure, i.e., e_1_, e_0_ refers to the exposed and the unexposed; U: unmeasured confounder; Y: outcome.* |
| **Mathematical formula** | **1. Interactions on the additive scale:**  ${Bias}_{\text{add }}=\delta_{1}\gamma_{1}-\delta_{0}\gamma_{0}$;  **2. Interactions on the multiplicative scale:**  ${Bias}_{\text{mult }}=\frac{1+\left( \gamma_{1}-1 \right)P\left( U=1\mid g_{1},e_{1},c \right)}{1+\left( \gamma_{1}-1 \right)P\left( U=1\mid g_{0},e_{1},c \right)}/\frac{1+\left( \gamma_{0}-1 \right)P\left( U=1\mid g_{1},e_{0},c \right)}{1+\left( \gamma_{0}-1 \right)P\left( U=1\mid g_{0},e_{0},c \right)}$;  **3. Relative excess risk due to interaction:**  - RERI**_c_**: causal relative excess risk due to interaction conditional on C=c,  $\mathrm{RERI}_{c}=\frac{1}{\kappa}\frac{\mathbb{E}\left[ Y\mid g_{1},e_{1},c \right]}{\mathbb{E}\left[ Y\mid g_{0},e_{0},c \right]}-\frac{\mathbb{E}\left[ Y\mid g_{1},e_{0},c \right]}{\mathbb{E}\left[ Y\mid g_{0},e_{0},c \right]}-\frac{1}{\kappa}\frac{\mathbb{E}\left[ Y\mid g_{0},e_{1},c \right]}{\mathbb{E}\left[ Y\mid g_{0},e_{0},c \right]}+1$;  where $\kappa=\frac{1+\left( \gamma_{1}-1 \right)P\left( U=1\mid e_{1},c \right)}{1+\left( \gamma_{1}-1 \right)P(U=1\mid c)}/\frac{1+\left( \gamma_{0}-1 \right)P\left( U=1\mid e_{0},c \right)}{1+\left( \gamma_{0}-1 \right)P(U=1\mid c)}$. |
| **Data assumptions and additional required features** | **These methods in the paper apply to causal interaction, which as defined by the authors, “examine the extent to which an outcome would change under interventions on both of the factors of interest.”^25^**  **1. Interactions on the additive scale:**  a) G does not interact with U on the additive scale.  b) The effect of G and E on Y are unconfounded conditional on {C, U}.  c) U is binary.  **2. Interactions on the multiplicative scale:**  a) U does not interact on the multiplicative scale with G or with E.  b) The effect of G and E on Y are unconfounded conditional on {C, U}.  **3. Relative excess risk due to interaction:**  a) G does not interact with U on the multiplicative scale.  b) Suppose that for all $g$ and $e,Y_{ge}$∐$\{G,E\}\mid\{C,U\}$ and we have $G\times E$ independence in the sense that $\{E,U\}\coprod G\mid C$.  c) U is binary.  *Notes: G and E are two factors or exposures of interest (which can be genetic and environmental factors, or two genetic factors or two environmental factors); U: unmeasured confounder; Y: outcome.* |
| **Available and/or recommended software** | Not mentioned |
| **Output of the QBA analysis** | **1. Interactions on the additive scale:**  Interactions on the additive scale: bias factor ${Bias}_{\text{add }}$;  **2. Interactions on the multiplicative scale:**  Interactions on the multiplicative scale: bias factor ${Bias}_{\text{mult }}$;  **3. Relative excess risk due to interaction:**  Relative excess risk due to interaction: $\mathrm{RERI}_{c}.$ |
| **Interpretation of the QBA analysis output** | **1. Interactions on the additive scale:**  Interactions on the additive scale: a corrected estimated of additive interaction can be obtained by subtracting the bias factor from the estimated additive interaction measure using the observed data.  **2. Interactions on the multiplicative scale:**  Interactions on the multiplicative scale: a corrected estimated of multiplicative interaction can be obtained by dividing the estimated multiplicative interaction measure by the bias factor.  **3. Relative excess risk due to interaction:**  Relative excess risk due to interaction: causal RERI conditional on C=c. |
| **Considerations related to the interpretation of the results** | 1.The authors highlighted that “if the two exposures of interest are independent, even under unmeasured confounding, if the estimate of the interaction is nonzero, then either there is a true interaction between the two factors or there is an interaction between one of the factors and the unmeasured confounder; an interaction must be present in either scenario.”^25^  2. The authors noted that additive interaction is most relevant for both public health purposes and for assessing mechanistic interaction, while the multiplicative scale is often used in practice. |
| **Explicit discussions regarding the similarities and differences with other methods** | Not mentioned |

#### Lubin et al 2018: indirect adjustment of relative risks of an exposure with multiple categories for an unmeasured confounder

| **Name/description** | **Indirect adjustment of relative risks of an exposure with multiple categories for an unmeasured confounder** |
| --- | --- |
| **DOI** | doi.org/10.1016/j.annepidem.2018.09.003 |
| **Author and Year** | Lubin et al 2018^26^ |
| **Crossed-referenced in QBA textbook^3^** | Not referenced |
| **QBA classification** | Multidimensional analysis |
| **Applicable study design scenarios described^b^** | Cohort studies^b^ |
| **Effect measure of interest** | RR, or HR |
| **Result of interest** | Corrected estimate |
| **Data structure/format of the eligible exposure/intervention^b^** | Categorical |
| **Data structure/format of the confounders, if applicable^b^** | Categorical |
| **Data structure/format of eligible outcome^b^** | Categorical, or time-to-event data |
| **Source(s) of bias addressed** | Unmeasured confounding (single) |
| **Information/parameters needed to conduct the QBA method** | - $\boldsymbol{R}\boldsymbol{R}_{\boldsymbol{X}}^{\boldsymbol{OBS}}\mathbf{(}\boldsymbol{i}\mathbf{)}$ **or** $\boldsymbol{ER}\boldsymbol{R}_{\boldsymbol{X}}^{\boldsymbol{OBS}}\mathbf{(}\boldsymbol{i}\mathbf{)}$: the RR or excess RR of disease for exposure level i;  - $\boldsymbol{R}\boldsymbol{R}_{\boldsymbol{C}}\mathbf{(}\boldsymbol{j}\mathbf{)}$: the RR of the confounder of confounder level j;  - $\boldsymbol{\pi}_{\mathbf{(}\boldsymbol{j}\boldsymbol{\mid}\boldsymbol{i}\mathbf{)}}$: for confounder level j, the two conditional probabilities of having the confounder for those with (i=1) and without exposure (i=0) or, equivalently, the probability of having the confounder for those without exposure and OR of X and C.  *Note: C: confounder; X: exposure.* |
| **Mathematical formula** | **Under a multiplicative model:**  $RR_{X}(i)=\frac{RR_{X}^{OBS}(i)}{\sum_{j=0}^{J} \pi_{(j\mid i)}RR_{C}(j)/\sum_{j=0}^{J} \pi_{(j\mid0)}RR_{C}(j)}$;  **Under an additive model:**  $\begin{matrix} \mathrm{ERR}_{X}(i)= & \sum_{j=0}^{J} \left\{ \pi_{(j\mid0)}\mathrm{ERR}_{X}^{OBS}(i)+\pi_{(j\mid0)}-\pi_{(j\mid i)} \right\}\mathrm{ERR}_{C}(j) \\ & +ERR_{X}^{OBS}\left( i \right). \end{matrix}$  *Note:* $RR_{X}\left( i \right) or ERR_{X}(i)$ *: for exposure level i, the true RR or excess RR.* |
| **Data assumptions and additional required features** | The true $RR_{X}\left( i \right)$ are null or equal to some predetermined values. |
| **Available and/or recommended software** | Excel spreadsheet (https://dceg.cancer.gov/tools/analysis/indirect-relative-risk-adjustment) |
| **Output of the QBA analysis** | Corrected estimate |
| **Interpretation of the QBA analysis output** | Effect estimate accounting for unmeasured confounding |
| **Considerations related to the interpretation of the results** | 1. Indirect adjustment rests on the validity of the external information for the factor in the study population under investigation.  2. The authors developed formulae for multiplicative and additive models for the joint association of X and C. However, other models are possible.  3. The authors noted that without information on the unmeasured factor, examination of model form is not feasible within the study data, and therefore the choice of model represents externally sourced information.  4. The authors noted that their method does not take into account uncertainties in the external information, nor does it provide confidence intervals for adjusted RRs that incorporate this uncertainty. |
| **Explicit discussions regarding the similarities and differences with other methods** | 1. The authors noted the alternative strategy by Schlesselman 1978, which “assumes no trend in the true exposure RRs and characterizes the conditions necessary for a proposed confounder to produce the observed trend.”^5^  2. The authors noted that the major difference between this method and the E-value and/or bounding factor is that the former gave a direct calculation of an adjusted RR while the latter equate the maximum RR. Hence, this method is useful when specific estimates are needed, such as for meta-analyses and risk assessments. |

### Unmeasured confounding (explain-away)

#### Cornfield et al 1959: sensitivity analysis for unmeasured confounding

| **Name/description** | **Sensitivity analysis for unmeasured confounding** |
| --- | --- |
| **DOI** | 10.1093/jnci/22.1.173 |
| **Year** | Cornfield et al 1959^7^ |
| **Crossed-referenced in QBA textbook^3^** | Referenced but not explained |
| **QBA classification** | Simple sensitivity analysis |
| **Applicable study design scenarios described^b^** | Any observational study design |
| **Effect measure of interest** | All relative measures (RR, OR etc.) |
| **Result of interest** | Explain-away |
| **Data structure/format of the eligible exposure/intervention^b^** | Categorical |
| **Data structure/format of the confounders, if applicable^b^** | Categorical |
| **Data structure/format of eligible outcome^b^** | Categorical |
| **Source(s) of bias addressed** | Unmeasured confounding (single) |
| **Information/parameters needed to conduct the QBA method** | - **p_1_/p_2_**: the prevalence of the confounder among the exposed, relative to the prevalence among those unexposed;  - **r**: crude association between the exposure and the outcome. |
| **Mathematical formula** | Any confounder (termed as the “common cause” to both exposure and outcome in the paper) must be at least r-fold more prevalent among the exposed than among nonexposed to explain away the observed effect. |
| **Data assumptions and additional required features** | Not mentioned |
| **Available and/or recommended software** | Not mentioned |
| **Output of the QBA analysis** | No |
| **Interpretation of the QBA analysis output** | Not mentioned |
| **Considerations related to the interpretation of the results** | Not mentioned |
| **Explicit discussions regarding the similarities and differences with other methods** | In Fox et al 2021,^3^ they noted that this paper is often cited as the first sensitivity analysis even if this is not in fact the first quantitative bias analysis. |

#### Greenland 1996: basic methods for sensitivity analysis of unmeasured confounding

| **Name/description** | **Basic methods for sensitivity analysis of unmeasured confounding** |
| --- | --- |
| **DOI** | https://doi.org/10.1093/ije/25.6.1107-a; or PMID: 9027513 |
| **Author and Year** | Greenland 1996^16^ |
| **Crossed-referenced in QBA textbook^3^** | Referenced but not explained |
| **QBA classification** | Simple sensitivity analysis |
| **Applicable study design scenarios described^b^** | Case-control or cohort studies |
| **Effect measure of interest** | OR^b^ |
| **Result of interest** | Corrected estimates |
| **Data structure/format of the eligible exposure/intervention^b^** | Categorical |
| **Data structure/format of the confounders, if applicable^b^** | Categorical |
| **Data structure/format of eligible outcome^b^** | Categorical |
| **Source(s) of bias addressed** | Unmeasured confounding (single) |
| **Information/parameters needed to conduct the QBA method** | - $\mathbf{A}_{\mathbf{1+,}}\mathbf{B}_{\mathbf{1+,}}\mathbf{A}_{\mathbf{0+}}\boldsymbol{,}\mathbf{B}_{\mathbf{0+}}$: cases among the exposed, controls among the exposed, cases among the unexposed, controls among the unexposed, respectively (See Table 1 and Table 2 in the manuscript);  - $\mathbf{OR}_{\mathbf{DZ}}$: OR between unmeasured confounder and outcome;  - $\boldsymbol{P}_{\boldsymbol{Z}\boldsymbol{0,}}\boldsymbol{P}_{\boldsymbol{Z}\boldsymbol{1}}$: prevalence of unmeasured confounder among the unexposed and the exposed, respectively.  *Notes: D: outcome; X: exposure; Z: confounder.* |
| **Mathematical formula** | $B_{11}=P_{Z1}B_{1+}$; $B_{01}=P_{Z0}B_{0+}$;  $A_{11}=\frac{\mathrm{OR}_{\mathrm{DZ}}A_{1+}B_{11}}{\left( \mathrm{OR}_{\mathrm{DZ}}B_{11}+B_{1+}-B_{11} \right)}$; $A_{01}=\frac{\mathrm{OR}_{\mathrm{DZ}}A_{0+}B_{01}}{\left( \mathrm{OR}_{\mathrm{DZ}}B_{01}+B_{0+}-B_{01} \right)}$;  $\mathrm{OR}_{\mathrm{DX}}=A_{11}{\text{ }B}_{01}/A_{01}{\text{ }B}_{11}=\left( A_{1+}-A_{11} \right)\left( B_{0+}-B_{01} \right)/\left( A_{0+}-A_{01} \right)\left( B_{1+}-B_{01} \right)$. |
| **Data assumptions and additional required features** | The exposure-outcome associations are constant across different strata of unmeasured confounder. |
| **Available and/or recommended software** | Not mentioned |
| **Output of the QBA analysis** | Stratum-specific $\mathrm{OR}_{\mathrm{DX}}$ |
| **Interpretation of the QBA analysis output** | Since no effect modification is assumed, the stratum-specific OR is the adjusted OR which accounts for the effect of unmeasured confounding. |
| **Considerations related to the interpretation of the results** | 1. The author noted that “the prevalence must be obtained externally; occasionally (and preferably) they may be obtained from a survey of the underlying source population from which the subjects were selected.”^16^  2. This method can extend to data with for instance person-time denominators or count denominators. |
| **Explicit discussions regarding the similarities and differences with other methods** | As one of the first sensitivity analyses methods, like previous methods including Bross 1966^1^ and Schlesselman 1978^5^, this method has the assumption that the ORs or RRs between the exposure and the outcome should be constant across strata of the unmeasured confounder. |

#### Frank 2000: impact thresholds for regression models

| **Name/description** | **Impact thresholds for regression models** |
| --- | --- |
| **DOI** | 10.1177/0049124100029002001 |
| **Author and Year** | Frank 2000^27^ |
| **Crossed-referenced in QBA textbook^3^** | Not referenced |
| **QBA classification** | Simple sensitivity analysis |
| **Applicable study design scenarios described^b^** | Any observational studies |
| **Effect measure of interest** | Regression coefficients |
| **Result of interest** | Explain-away |
| **Data structure/format of the eligible exposure/intervention^b^** | Categorical or continuous |
| **Data structure/format of the confounders, if applicable^b^** | Categorical or continuous |
| **Data structure/format of eligible outcome^b^** | Continuous [categorical (information below is for linear, but authors note that the method could be extended for logistic)] |
| **Source(s) of bias addressed** | Unmeasured confounding (single or multiple) |
| **Information/parameters needed to conduct the QBA method** | - **n**: sample size;  - **q**: number of parameters estimated in a regression model;  - $\boldsymbol{r}_{\boldsymbol{y}\boldsymbol{\cdot}\boldsymbol{x}}$: the sample correlation between the outcome and the predictor of interest;  - **t** ratio: calculated as $t=t\left( \hat{\beta}_{1} \right)=\frac{\hat{\beta}_{1}}{se\left( \hat{\beta}_{1} \right)},$ where $\hat{\beta}_{1}$ is the estimated regression coefficient for the predictor variable. |
| **Mathematical formula** | Impact threshold for a confounding variable (ITCV) is defined as the product of the two variables -$r_{y\cdot cv}$, which is the relationship between outcome and confounder, and $r_{x\cdot cv}$ , which is the relationship between the predictor of interest and confounder: $k=ITCV=r_{y\cdot cv}\times r_{x\cdot cv}$; and k is calculated as: $d=t^{2}+(n-q-1)$, then $k=\frac{t^{2}+t\sqrt{d}}{-(n-q-1)}+\left[ \frac{-d-t\sqrt{d}}{-(n-q-1)} \right]r_{y\cdot x}$  *Note: ITCV: impact thresholds for a confounding variable.* |
| **Data assumptions and additional required features** | Not mentioned |
| **Available and/or recommended software** | Not mentioned |
| **Output of the QBA analysis** | k=ITCV |
| **Interpretation of the QBA analysis output** | k is defined as the minimum impact of the unobserved confounder that is necessary not to reject the null hypothesis of zero effect. Therefore, the impact of confounder should be greater than k in order to explain away the observed finding. |
| **Considerations related to the interpretation of the results** | The author noted that the magnitude of ITCV depends in part on sample size and significant level (i.e., the values of ITCV are larger for larger sample sizes and/or larger significant level).  The authors noted that the “ideas in this article could also be extended beyond the general linear model. For example, one could obtain the ITCV for multilevel models or logistic regressions” |
| **Explicit discussions regarding the similarities and differences with other methods** | 1. The author mentioned the Size rule method by Bross 1966^1^, which accounts for the impact of a confounder on a coefficient in log-linear models for contingency tables. In contrast, the ITCV applies to linear models and focuses on the impact on inference rather than the change in the size of the coefficient.  2. Compared to sensitivity analyses by Rosenbaum 1986^28^, the author noted that “ITCV neatly summarizes the threshold value as a function of the two critical pieces of information associated with the confound” which greatly reduces the information required.^27^ Moreover, the author noted that the concise expression also facilitates the extension of ITCV to the multivariate case and other topics.^27^ 3. Cinelli and Hazlett 2019^29^ , who proposed robustness value method, noted that for the impact thresholds method, for given $q$ and $R_{Y\sim D\mid\mathbf{X}}$, the impact that is necessary to bring about a relative bias of magnitude $q$ depends on the sensitivity parameter $R_{D\sim Z\mid\mathbf{X}}^{2}$ (which is the partial R^2^ from regressing exposure D on unobserved covariates Z after controlling for observed covariates X), except when $q=1$. However, the robustness value does not depend on $R_{D\sim Z\mid\mathbf{X}}^{2}.$ |

#### Phillips 2003: rule-out approach (target-adjustment sensitivity analysis)

| **Name/description** | **Rule-out approach (target-adjustment sensitivity analysis)** |
| --- | --- |
| **DOI** | 10.1097/01.ede.0000072106.65262.ae |
| **Author and Year** | Original manuscript: Phillips 2003^30^.  Additional information: Schneeweiss 2006^2^. This review article provides a clear overview of the methods and was used to populate the cells below. |
| **Crossed-referenced in QBA textbook^3^** | Referenced but not explained |
| **QBA classification** | Simple sensitivity analysis |
| **Applicable study design scenarios described^b^** | Any observational study designs |
| **Effect measure of interest** | RR |
| **Result of interest** | Explain-away |
| **Data structure/format of the eligible exposure/intervention^b^** | Categorical |
| **Data structure/format of the confounders, if applicable^b^** | Categorical |
| **Data structure/format of eligible outcome^b^** | Categorical |
| **Source(s) of bias addressed** | Unmeasured confounding (single) |
| **Information/parameters needed to conduct the QBA method** | - **ARR**: apparent (or observed) RR;  - **P_C_**: Prevalence of confounder in the total population;  - **P_E_**: Prevalence of exposure in the total population;  - **RR_CD_**: RR between measured confounder and disease outcome. |
| **Mathematical formula** | $P_{C}=\frac{P_{E}-P_{E}^{2}-\left( P_{E}P_{C}\mathrm{RR}\mathrm{CD}_{C}\mathrm{ARR} \right)+\left( P_{E}P_{C}\mathrm{ARR} \right)+\left( P_{E}^{2}\mathrm{ARR} \right)-\left( P_{E}\mathrm{ARR} \right)}{\left( P_{E}\mathrm{ARR} \right)-\left( P_{E}\mathrm{RR}_{\mathrm{CD}}\mathrm{ARR} \right)-\mathrm{RR}_{\mathrm{CD}}+1+\left( P_{E}\mathrm{RR}\mathrm{CD}_{\mathrm{CD}} \right)-P_{E}}$;  $\mathrm{OR}_{\mathrm{EC}}=\frac{P_{C1}\left[ 1-P_{C}-P_{E}+P_{C1} \right]}{\left[ P_{C}-P_{C1} \right]\left[ P_{E}-P_{C1} \right]}$.^2^  *Note:* ${OR}_{EC}$*: the association between exposure and confounder.* |
| **Data assumptions and additional required features** | Rule-out approach assumes that the true RR is null (RR=1) and explores how strong an unmeasured confounder has to be to fully explain the observed findings. |
| **Available and/or recommended software** | Excel spreadsheet (https://www.drugepi.org/dope/software#Sensitivity-analysis) |
| **Output of the QBA analysis** | $\mathrm{OR}_{\mathrm{EC}}$ |
| **Interpretation of the QBA analysis output** | The association between exposure and confounder. For a given ARR, RR_CD_, P_C_, and P_E_, the relationship between OR_EC_ and RR_CD_ can help understand if ARR can be explained away. |
| **Considerations related to the interpretation of the results 1.1.** | 1. This method is constrained to only one binary confounder.  2. Schneeweiss 2006 noted that “these methods, unlike external adjustment, do not provide an assessment of the magnitude of the existing residual confounding in a specific study, or the choice of reference group.”^2^ |
| **Explicit discussions regarding the similarities and differences with other methods** | 1. The author noted that this method can be limited and fails to assess the interaction of multiple biases. Moreover, it can create the impression that this method tries to make the effect go away rather than tries to improve the measurement.  2. Zhang et al 2006 evaluated the E-value, array approach, and rule-out approach, and found that the results from these methods are numerically equivalent.^4^ |

#### Yu and Gastwirth 2005: sensitivity analysis for trend tests

| **Name/description** | **Sensitivity analysis for trend tests** |
| --- | --- |
| **DOI** | 10.1093/biostatistics/kxi003 |
| **Author and Year** | Yu and Gastwirth 2005^31^ |
| **Crossed-referenced in QBA textbook^3^** | Not referenced |
| **QBA classification** | Multidimensional analysis |
| **Applicable study design scenarios described^b^** | Case-control studies with trend or dose-response tests |
| **Effect measure of interest** | OR |
| **Result of interest** | Explain-away |
| **Data structure/format of the eligible exposure/intervention^b^** | Categorical |
| **Data structure/format of the confounders, if applicable^b^** | Categorical or continuous |
| **Data structure/format of eligible outcome^b^** | Categorical |
| **Source(s) of bias addressed** | Unmeasured confounding (single) |
| **Information/parameters needed to conduct the QBA method** | - **δ**: the association between X and U;  - $\boldsymbol{\omega}_{\boldsymbol{1\vert0}}$**,** $\boldsymbol{\omega}_{\boldsymbol{1\vert1}}$: prevalence of U at different exposure level among the control and among the exposure group, respectively;  - $\boldsymbol{e}^{\boldsymbol{\gamma}}$: OR between U and R.  *Notes: R: outcome; U: unmeasured confounder; X: exposure.* |
| **Mathematical formula** | $P\left( Z_{\mathrm{CA}}\leq t \right)=\Phi\left( \frac{s_{0}t-B_{0}}{\sigma_{0}} \right)$  where $s_{0}=\left[ \hat{\pi}(1-\hat{\pi})\sum_{j=1}^{J} n_{j}\left( x_{j}-\bar{x} \right)^{2} \right]^{\frac{1}{2}}$ only depends on observed data and $B_{0}=\sum_{j=1}^{J} n_{j}\pi_{j}^{(0)}\left( x_{j}-\bar{x} \right)$ and $\sigma_{0}=\left[ \sum_{j=1}^{J} \pi_{j}^{(0)}\left( 1-\pi_{j}^{(0)} \right)n_{j}\left( x_{j}-\bar{x} \right)^{2} \right]^{\frac{1}{2}}$ depend on observed data as well as the parameters $(\delta,\gamma, \omega)$.  The detailed formula can be found in the original paper and the Appendix of Yu and Gastwirth 2005^31^. |
| **Data assumptions and additional required features** | There is a same increasing or decreasing trend in the prevalences of unmeasured confounder U across the control, low-dose to high-dose groups. |
| **Available and/or recommended software** | Not mentioned |
| **Output of the QBA analysis** | The p-values of the trend test with different values of (ω, δ, γ) for U and required odds ratio of U to set the p-value to 0.05. |
| **Interpretation of the QBA analysis output** | The same as above: the p-values of the trend test with different values of (ω, δ, γ) for U and required odds ratio of U to set the p-value to 0.05. |
| **Considerations related to the interpretation of the results** | Not mentioned. |
| **Explicit discussions regarding the similarities and differences with other methods** | 1. The authors noted that “The analysis complements that of Rosenbaum 2003^32^ who considered exposures that were measured on a continuous scale.  2. The authors noted that “This paper utilizes an approach similar to that of Rosenbaum 2002^33^ to develop a sensitivity analysis for testing data for a dose–response or trend.”. However, the main difference is that this paper follows Cornfield et al 1959^34^ who consider the conditional distribution of the unobserved variable U given the dose or exposure level X, rather than the propensity score. |

#### Lee and Wang 2007: bounding formulas for population stratification bias

| **Name/description** | **Bounding formulas for population stratification bias** |
| --- | --- |
| **DOI** | 10.1093/aje/kwm257 |
| **Author and Year** | Lee and Wang 2007^35^ |
| **Crossed-referenced in QBA textbook^3^** | Not referenced |
| **QBA classification** | Simple sensitivity analysis |
| **Applicable study design scenarios described^b^** | Case-control studies for genetic associations |
| **Effect measure of interest** | OR |
| **Result of interest** | Explain-away |
| **Data structure/format of the eligible exposure/intervention^b^** | Categorical |
| **Data structure/format of the confounders, if applicable^b^** | Categorical |
| **Data structure/format of eligible outcome^b^** | Categorical |
| **Source(s) of bias addressed** | Unmeasured confounding by population stratification (single) |
| **Information/parameters needed to conduct the QBA method** | - **B**: the ratio of the largest background disease rate and the smallest background disease rate;  - **G**: the ratio of the largest frequency odds and the smallest frequency odds of the susceptibility genotype. |
| **Mathematical formula** | U and L denote the upper and lower bound for the confounding rate ratio, which computed as follows:  $U=\frac{\sqrt{G\times B}\times(\sqrt{G\times B}+1)^{2}}{(\sqrt{G\times B}+G)\times(\sqrt{G\times B}+B)}\geq1;$  $L=(1/U)\leq1$.  *Note: The authors also proposed formulas for the upper bound of the type I error rate, which is not presented in this table since it does not focus on bias quantification. Readers can resort to it based on their interest.* |
| **Data assumptions and additional required features** | Relative rate of disease on the susceptibility genotype is constant across the different strata of a population. |
| **Available and/or recommended software** | Not mentioned |
| **Output of the QBA analysis** | U and L, which are the bounds for the confounding rate ratio. |
| **Interpretation of the QBA analysis output** | If the upper bound of confounding rate ratio U is less than the observed OR, then it suggests that the finding cannot be explained away by population stratification bias alone. |
| **Considerations related to the interpretation of the results** | 1. The authors noted that “population stratification bias is the largest when there are two strata in the study population, the first stratum having frequency odds of susceptibility genotype g and background disease rate b and the second stratum having frequency odds of susceptibility genotype g*G and background disease rate b*B.”^35^  2. The authors also noted their understanding for the bounds: “The above bounds are not intended to be close to the true magnitude of population stratification bias. Rather, they serve to accommodate the magnitude of bias for every possible population stratification scenario conceivable, under the G and B constraints. G and B themselves are to be determined in an ad hoc manner. To err on the safe side, one can overexaggerate the values of G and B (to obtain more conservative bounds for population stratification bias) on the basis of one’s best (but perhaps scant) knowledge of the stratified population under study.”^35^ |
| **Explicit discussions regarding the similarities and differences with other methods** | Wacholder et al 2000^36^ and Wang et al 2004 ^37^ (the magnitude of bias), and Heiman et al 2004^38^ and Gorroochurn et al 2004^39^ (the magnitude of the false-positive rate) also studied population stratification bias by using computer simulation to demonstrate the impacts of bias in many different situations. |

#### Lee 2011: bounding formulas for unmeasured confounding with effect-modifying potentials

| **Name/description** | **Bounding formulas for unmeasured confounding with effect-modifying potentials** |
| --- | --- |
| **DOI** | 10.1002/sim.4151 |
| **Author and Year** | Lee 2011^40^ |
| **Crossed-referenced in QBA textbook^3^** | Not referenced |
| **QBA classification** | Simple sensitivity analysis |
| **Applicable study design scenarios described^b^** | Any observational studies with non-null findings |
| **Effect measure of interest** | RR, OR, rate ratio (for person-time data) |
| **Result of interest** | Explain-away |
| **Data structure/format of the eligible exposure/intervention^b^** | Categorical |
| **Data structure/format of the confounders, if applicable^b^** | Categorical or continuous |
| **Data structure/format of eligible outcome^b^** | Categorical |
| **Source(s) of bias addressed** | Unmeasured confounding (single or multiple) |
| **Information/parameters needed to conduct the QBA method** | - **crude RR**;  - $\boldsymbol{U}_{\boldsymbol{D}}$: the maximum strength of association (in terms of odds ratio) between unmeasured confounder and outcome;  - $\boldsymbol{U}_{\boldsymbol{E}}$: the maximum strength of association (in terms of risk ratio) between unmeasured confounder and exposure；  - $\boldsymbol{U}_{\boldsymbol{M}}$: the maximum modifying effect (in terms of ratio of risk ratios) unmeasured confounder has on the association between exposure and outcome. |
| **Mathematical formula** | 1. **Bounding formulas for confounding risk ratios (CRR), which is defined as CRR=crude RR/standardized risk ratio (SRR)**:  **Bounds for CRR^E+^**:  $\mathrm{Upper}^{E+}=\frac{1}{\text{ Lower }^{E+}}=\left( \frac{\sqrt{U_{E}U_{D}}+1}{\sqrt{U_{E}}+\sqrt{U_{D}}} \right)^{2};$  **Bounds for CRR^E-^**:  $\text{ Upper }^{E-}=\frac{1}{\text{ Lower }^{E-}}=\left( \frac{\sqrt{U_{E}U_{D}U_{M}}+1}{\sqrt{U_{E}}+\sqrt{U_{D}U_{M}}} \right)^{2}.$  *Notes: D: outcome; E: exposure; M: effect-modifier;*  2$.$ **Conditions to explain away a finding**:  $\left( \left( \sqrt{U_{E}U_{D}}+1 \right)/\left( \sqrt{U_{E}}+\sqrt{U_{D}} \right) \right)^{2}>RR$;  Solving the above inequality while satisfying the following conditions can obtain the value of RR which will be used to calculate $T_{L}{and T}_{H}$.  $\mathrm{Min}\left( U_{E},U_{D} \right)>T_{L}=RR$ and $\max\left( U_{E},U_{D} \right)>T_{H}=(\sqrt{\mathrm{RR}}+\sqrt{RR-1})^{2}$. |
| **Data assumptions and additional required features** | The formula for explain-away only applied to RR>1. |
| **Available and/or recommended software** | Not mentioned |
| **Output of the QBA analysis** | 1. **Bounds for CRR^E+^, CRR^E-^**;  2$.$ **Conditions to explain away a finding:** $T_{L}, T_{H}$ (which are calculated from RR) |
| **Interpretation of the QBA analysis output** | 1. **Interpretation for bounds for CRR^E+^, CRR^E-^**: In the presence of unmeasured confounding, if the sensitivity analysis parameter take values (i.e., $U_{E},U_{D}, U_{M}$), bounds for CRR^E+^, CRR^E-^ stands for the possible range for CRR, which is used to quantify the bias associated with U. In particular, CRR^E+^=crude RR/SRR^E+^, where SRR^E+^ can be interpreted as ‘proportionate increase due to the exposure in the disease risk of the exposed population’; ^40^ likewise, CRR^E-^=crude RR/SRR^E-^, where SRR^E-^ can be interpreted as ‘proportionate increase in the disease risk that would be expected to occur in the unexposed population if they were exposed’.^40^  2$.$ **Interpretation for**$\boldsymbol{T}_{\boldsymbol{L}}\boldsymbol{,}\boldsymbol{T}_{\boldsymbol{H}}$**:** For an unmeasured factor to explain away an RR, its strengths of association with the exposure ($U_{E}$ ) and with the disease ($U_{D}$) must both exceed $T_{L}$ , and additionally, at least one of these two strength parameters must exceed $T_{H}$. As a helpful resource, Table I in the original article presents $T_{L}$and $T_{H}$ for various values of RR. |
| **Considerations related to the interpretation of the results** | 1. The author noted that “If one has difficulties in gauging U in any one aspect of its strengths, one can simply set the corresponding constraint to infinity.”^40^  2. The author noted that “This study considers the situations when the exposed group and the unexposed group, respectively,  are taken as the standard population. When the total group is used instead as the standard population, Appendix B shows that CRR is a weighted harmonic mean and is therefore a value between CRR^E+^ and CRR^E−^.”^40^ |
| **Explicit discussions regarding the similarities and differences with other methods** | 1. The authors mentioned that Lee and Wang 2007^35^ proposed a similar bounding formula which corresponds exactly to the special case of U_M_=1in this paper.  2. The authors noted that their results are in agreement with the signs of the bias described in VanderWeele 2008^41^ and Chiba 2009^42^.  3. Flanders and Ye 2019^43^ compared their method to this method, and found that Lee’s result is a special and limiting case. |

#### Hasegawa and Small 2017: average case calibration

| **Name/description** | **Average case calibration** |
| --- | --- |
| **DOI** | 10.1111/biom.12688 |
| **Author and Year** | Hasegawa and Small 2017^44^ |
| **Crossed-referenced in QBA textbook^3^** | Not referenced |
| **QBA classification** | Simple sensitivity analysis |
| **Applicable study design scenarios described^b^** | Matched-pair observational studies |
| **Effect measure of interest** | OR |
| **Result of interest** | Explain away |
| **Data structure/format of the eligible exposure/intervention^b^** | Categorical |
| **Data structure/format of the confounders, if applicable^b^** | Categorical |
| **Data structure/format of eligible outcome^b^** | Categorical |
| **Source(s) of bias addressed** | Unmeasured confounding (single) |
| **Information/parameters needed to conduct the QBA method** | -$\bar{\mathbf{p}}:$the sample average of p_s_, which is the probability that the unit with positive outcome, receives treatment in pair s, i.e., $\bar{\mathbf{p}}=\left( \sum_{s=1}^{S} p_{s} \right)/S$, where S refers to the number of matched pairs. |
| **Mathematical formula** | $\Gamma_{\text{ }\text{truth}}^{'}=\bar{\mathbf{p}}/(1-\bar{\mathbf{p}})$.  *Notes: in the paper* $\Gamma_{\text{ }}^{'}$*denotes average calibration bias, while Γ or Γ_sens_ denotes the worst-case bias (i.e.,* *the largest value of the worst-case bias across matched pairs that does not invalidate the finding of evidence for a treatment effect.). The subscript truth refers to the true unknown parameter, for instance,* $\Gamma_{\text{ }\text{truth}}^{'}$*refers to the true unknown average calibration bias.* |
| **Data assumptions and additional required features** | The authors noted that “The proof relies on Schur-convexity of the distribution function of the test statistic with respect to p which requires that it be symmetric in p.”^44^ |
| **Available and/or recommended software** | Not mentioned |
| **Output of the QBA analysis** | $\Gamma_{\text{ }\text{truth}}^{'}$ |
| **Interpretation of the QBA analysis output** | $\Gamma_{\text{ }\text{truth}}^{'}$ denotes the true unknown average calibration bias (which is less conservative than the worst-case bias) and is interpretated as the average value of the worst-case bias across matched pairs that does not invalidate the finding of evidence for a treatment effect. For instance, an $\Gamma_{\text{ }\text{truth}}^{'}$ value = 2.1 would be interpretated as: if the average probability of being exposed is at most 2.1 times that of being unexposed, then it is plausible that the observed finding cannot be explained away by the unmeasured confounding at significance level α set by the researchers. |
| **Considerations related to the interpretation of the results** | Not mentioned |
| **Explicit discussions regarding the similarities and differences with other methods** | 1. This method builds upon on Rosenbaum 1987^45^’s method. Average case calibration is able to handle violations to the assumptions in worst-case sensitivity analysis by Rosenbaum^46^, such as the violation to positivity assumption.  2. The authors mentioned that the results in Rosenbaum 2002^47^ shows that it may be possible to extend average case calibration analysis to a study with non-binary outcomes. |

#### VanderWeele and Ding 2017 E-value and Ding and VanderWeele 2016 bounding factor

| **Name/description** | **E-value** |
| --- | --- |
| **DOI** | 10.7326/m16-2607; 10.1097/EDE.0000000000000457 |
| **Year** | VanderWeele and Ding 2017; Ding and VanderWeele 2016^48,49^ |
| **Crossed-referenced in QBA textbook^3^** | Referenced and explained |
| **QBA classification** | Simple sensitivity analysis |
| **Applicable study design scenarios described^b^** | Any observational study designs |
| **Effect measure of interest** | RR, OR, HR, rate ratio, or RD |
| **Result of interest** | Explain-away |
| **Data structure/format of the eligible exposure/intervention^b^** | Categorical |
| **Data structure/format of the confounders, if applicable^b^** | Categorical, or continuous |
| **Data structure/format of eligible outcome^b^** | Categorical, continuous, or time-to-event data |
| **Source(s) of bias addressed** | Unmeasured confounding (single or multiple) |
| **Information/parameters needed to conduct the QBA method** | **Bounding factor:**  - $\mathbf{RR}_{\boldsymbol{EU}}$: observed RR for the exposure and unmeasured confounder  -$\mathbf{RR}_{\boldsymbol{UD}}$: observed RR for the unmeasured confounder and outcome  - $\mathbf{RR}_{\boldsymbol{ED}}^{\text{obs }}$: observed RR for the exposure and outcome  **E-value:**  - **RR**: observed RR for the exposure and outcome (or other effect measures)  - **CI closest to the null** |
| **Mathematical formula** | **Bounding factor:**  $bounding factor= \frac{\mathrm{RR}_{EU}\times\mathrm{RR}_{UD}}{\mathrm{RR}_{EU}+\mathrm{RR}_{UD}-1}\geq\frac{\mathrm{RR}_{ED}^{\text{obs }}}{\mathrm{RR}_{ED}^{\text{true }}}$;  **E-value:**  1. When RR>1  E-value for the effect estimate:  $E-value=RR+\sqrt{RR\times\left( RR-1 \right)}$;  E-value for the lower limit of the CI:  $E-value=RR+\sqrt{RR\times\left( RR-1 \right)}$ , if lower limit >1;  $E-value=1$ , if lower limit ≤1.  2. When RR<1,  E-value for the effect estimate: $E-value=\frac{1}{RR}+\sqrt{\frac{1}{RR}\times\left( \frac{1}{RR}-1 \right)}$;  E-value for the upper limit of the CI:  $E-value=\frac{1}{RR}+\sqrt{\frac{1}{RR}\times\left( \frac{1}{RR}-1 \right)}$ , if upper limit <1;  $E-value=1$ , if upper limit ≥1.  Please find formula for other effect measures in the original paper in Table 2 of the original manuscript. |
| **Data assumptions and additional required features** | 1. When researchers calculate the E-value, it is assumed that the association between confounder and exposure is of the same strength as the association between confounder and outcome.  2. It is clearly stated in other articles that the assumption of no interaction between the exposure and the confounder is not required in the E-value method.^50^ |
| **Available and/or recommended software** | Online calculator (https://www.evalue-calculator.com/evalue/); R package Evalue^51^. |
| **Output of the QBA analysis** | Bounding factor; E-value for the point estimate and E-value for the CI |
| **Interpretation of the QBA analysis output** | **Interpretation for bounding factor:**  In the presence of unmeasured confounding, if the sensitivity analysis parameter takes specified values (i.e., $\mathrm{RR}_{EU}$, $\mathrm{RR}_{UD}$), the true RR must be as large as the value of ${(RR}_{ED}^{\text{obs }}$/bouding factor).  **Interpretation for E-value in general:**  The minimum strength of association, on the risk ratio scale, that an unmeasured confounder would need to have with both the treatment and outcome to fully explain away a specific treatment–outcome association, conditional on the measured covariates. |
| **Considerations related to the interpretation of the results** | 1. Interpretation of E-value depends on context (e.g., measured covariates for which adjustment has been made).^52^  2. E-value does not guarantee that if a confounder with parameters of a particular strength existed, then it necessarily would explain away the effect, only that it is possible to construct scenarios in which it could.^49^ Moreover, the lowest possible E-value is 1.0.  3. The authors noted that reporting the E-value for the limit of the CI closest to the null is good practice.  4. E-value requires interpretations along with other strengths and weaknesses of the study.  5. E-value depends on the magnitude of the association; it cannot be made arbitrarily large simply by increasing the sample size. The E-value for the CI does depend on the sample size. |
| **Strengths and limitations identified in studies comparing or commenting on QBA methods** | 1. MacLehose et al 2021 noted that one unstated assumption of the E-value method is that the prevalence of the confounder among the exposed is 100%.^53^  2. Ioannidis et al 2019 and VanderWeele et al 2019 provided overviews to the potential misinterpretations of E-value.^54,55^  3. Cusson and Infante-Rivard 2020 provided modification to the E-value method, which covered different scenarios including an opposite direction of associations between unmeasured confounder and exposure, and between unmeasured confounder and outcome.^50^  4. Zhang et al 2006 evaluated E-value, array approach, and rule-out approach, and found that the results are numerically equivalent.^4^  5. Ding and VanderWeele 2006 provided proof and formula for the E-value method.^49^ |

#### Cinelli and Hazlett 2019: robustness value

| **Name/description** | **Robustness value** |
| --- | --- |
| **DOI** | https://rss.onlinelibrary.wiley.com/doi/10.1111/rssb.12348 |
| **Author and Year** | Cinelli and Hazlett 2019^29^ |
| **Crossed-referenced in QBA textbook^3^** | Not referenced |
| **QBA classification** | Simple sensitivity analysis |
| **Applicable study design scenarios described^b^** | Any observational studies with non-null findings |
| **Effect measure of interest** | Linear regression coefficients |
| **Result of interest** | Explain away |
| **Data structure/format of the eligible exposure/intervention^b^** | Categorical or continuous |
| **Data structure/format of the confounders, if applicable^b^** | Categorical or continuous |
| **Data structure/format of eligible outcome^b^** | Continuous |
| **Source(s) of bias addressed** | Unmeasured confounding (single or multiple) |
| **Information/parameters needed to conduct the QBA method** | The following parameters from a linear regression model are required:  - **Coefficient estimate;**  - **Standard error;**  - **Degrees of freedom;**  - **Significance level (**default is 0.05);  - **q**: percent change of the effect estimate that would be deemed problematic. The default value for q is 1, which means a reduction of 100% of the current effect estimate (bring estimate to zero)**.**  *Note: the authors also introduced a procedure to bound the strength of confounders formally on the basis of a comparison with observed covariates, which can be found in the original manuscript section 4.4.* |
| **Mathematical formula** | **R package** sensemak (https://cran.r-project.org/package=sensemakr) or **online calculator** can be used to directly obtain the results (<https://carloscinelli.shinyapps.io/robustness_value/>).  The formula for robustness value is:  $\mathrm{RV}_{q}=\frac{1}{2}\left\{ \sqrt{\left( f_{q}^{4}+4f_{q}^{2} \right)}-f_{q}^{2} \right\}$  where $f_{q}:=q\left\vert f_{Y\sim D\mid X} \right\vert$ is the partial Cohen’s $f$ of the treatment with the outcome multiplied bv the proportion of reduction $q$ on the treatment coefficient which would be deemed problematic and $f_{Y\sim D\mid\mathbf{X}}$ can be obtained by simply dividing the treatment coefficient $t$-value by $\sqrt{df}$. |
| **Data assumptions and additional required features** | Calculation of the robustness value assumes that there are equal associations between the confounder and the treatment and the confounder and the outcome. Moreover, this method does not require assumptions on the functional form of the treatment assignment mechanism nor on the distribution of the unobserved confounder and can be used to assess the sensitivity of multiple confounders, whether they influence the treatment and outcome linearly or not. |
| **Available and/or recommended software** | R package sensemakr, or online calculator (https://carloscinelli.shinyapps.io/robustness_value/). |
| **Output of the QBA analysis** | Robustness value for the point estimate and one for the t-value*;$R_{Y\sim D\mid\mathbf{X}}^{2};$ sensitivity contour plots.  *Notes: D: treatment; X: observed confounders; Y: outcome. *Bounds on confounding strength relative to a measured confounder and robustness value for confidence intervals can also be calculated if needed.* |
| **Interpretation of the QBA analysis output** | **Robustness value for the point estimate and the t-value**: robustness value for the point estimate describes the minimum strength of association that unobserved confounding would need to have, both with the treatment and with the outcome, to bring the effect estimate down to exactly 0.*  If the confounders’ association to the treatment and to the outcome (measured in terms of partial R^2^) are both assumed to be less than the robustness value, then such confounders cannot ‘explain away’ the observed effect. A robustness value that is close to 1 means that the treatment effect can handle strong confounders explaining almost all residual variation of the treatment and the outcome and a robustness value close to 0 means that even very weak confounders could eliminate the results.  The robustness value for the t-value describes the minimum strength of association (measured in terms of partial R^2^) that brings the point estimate not to exactly 0 but rather into a range where it is no longer ‘statistically different’ from 0.  $\boldsymbol{R}_{\boldsymbol{Y}\boldsymbol{\sim}\boldsymbol{D}\boldsymbol{\mid X}}^{\mathbf{2}}$: the proportion of variation in the outcome explained uniquely by the treatment, which shows how strongly confounders that explain 100% of the residual variance of the outcome would have to be associated with the treatment to eliminate the effect.^29^ For example, the partial R^2^ of the treatment with the outcome being 2.23% would be interpreted as: if confounders explained 100% of the residual variance of the outcome, they would need to explain at least 2.23% of the residual variance of the treatment to fully account for the observed estimated effect.  **Sensitivity contour plots**: how the effect estimate would vary depending on hypothetical strengths of the confounder.  *Note: ** ${RV}_{q}$ *describes how strong that association must be to reduce the estimated effect by* $100q\%$*. Here we presented the interpretation for robustness value* ${RV}_{q}$*, when q=1. Readers can also change values of q based on their research interest.* |
| **Considerations related to the interpretation of the results** | For $R_{Y\sim D\mid\mathbf{X}}^{2}$, the author noted that it “is not only the determinant of the robustness of the treatment effect coefficient but can also be interpreted as the result of an ‘extreme scenario’ sensitivity analysis.” |
| **Explicit discussions regarding the similarities and differences with other methods** | The authors discussed the relation of this method to two other methods: impact thresholds^27^ and the E-value^48^.  For the **impact thresholds method**, the authors noted that for given $q$ and $R_{Y\sim D\mid\mathbf{X}}$, the impact that is necessary to bring about a relative bias of magnitude $q$ depends on the sensitivity parameter $R_{D\sim Z\mid\mathbf{X}}^{2}$ (which is the partial R2 from regressing exposure D on unobserved covariates Z after controlling for observed covariates X), except when $q=1$. The robustness value does not depend on $R_{D\sim Z\mid\mathbf{X}}^{2}.$  For the **E-value**, the authors noted two things: 1) the robustness value is an exact value if the researcher uses linear regression to obtain an estimate. However, the E-value is only designed for risk ratio; for other effect measures, the E-value is an approximation; 2) the E- value and robustness value served for different effect measures.  The authors also noted that compared to previous methods, including Rosenbaum and Rubin 1983^8^, this method does not require specifying the distribution of the confounder as well as modelling the treatment assignment mechanism. |

#### Cusson and Infante-Rivard 2020: E-value extensions

| **Name/description** | **E-value extensions (1. For opposite relations of unmeasured confounders with exposure and outcome 2. For odds ratio scale)** |
| --- | --- |
| **DOI** | 10.1093/ije/dyaa127 |
| **Author and Year** | Cusson and Infante-Rivard 2020^50^ |
| **Crossed-referenced in QBA textbook^3^** | Not referenced |
| **QBA classification** | Simple sensitivity analysis |
| **Applicable study design scenarios described^b^** | Any observational study designs with non-null findings |
| **Effect measure of interest** | OR, RR, or HR |
| **Result of interest** | Explain-away |
| **Data structure/format of the eligible exposure/intervention^b^** | Categorical |
| **Data structure/format of the confounders, if applicable^b^** | Categorical |
| **Data structure/format of eligible outcome^b^** | Categorical, or time-to-event data |
| **Source(s) of bias addressed** | Unmeasured confounding (single or multiple) |
| **Information/parameters needed to conduct the QBA method** | - Observed **RR** or **OR** |
| **Mathematical formula** | 1. For RR scale,  B = observed RR/ true RR, where true RR is usually hypothesized to be 1.  For opposite relations of unmeasured confounders with exposure and outcome (Table 2 in the original manuscript):  a. ${RR}_{UY}>1$ and ${RR}_{XU}<1$  $E=[1+sqrt(1-B)]/B$;  b. ${RR}_{UY}<1$ and ${RR}_{XU}>1$  $E=\left[ \left( B+1 \right)+\mathrm{sqrt} \left( (B+1)^{2}-4*B^{2} \right) \right]/(2B)$.  2. For OR scale,  The full formula can be found in Table 3.  *Note: B: the maximum value of bias factor; E: E-value; U: unmeasured confounder; X: exposure; Y: outcome. Example* ${RR}_{UY}$ *is the risk ratio between unmeasured confounder and the outcome* |
| **Data assumptions and additional required features** | RR_XU_ and RR_UY_ do not need to be equal, as required by the original E-value method. |
| **Available and/or recommended software** | Not mentioned |
| **Output of the QBA analysis** | E-value |
| **Interpretation of the QBA analysis output** | The same as original E-value method: the minimum strength of association, on the risk ratio scale, that an unmeasured confounder would need to have with both the treatment and outcome to fully explain away a specific treatment–outcome association, conditional on the measured covariates. |
| **Considerations related to the interpretation of the results** | Similar to original E-value method^48^ (See VanderWeele and Ding 2017: E-value method) |
| **Explicit discussions regarding the similarities and differences with other methods** | 1. The authors discussed the bias factor, the maximum bias B, and the E-value methods. The bias factor, proposed by Bross 1966^1^ and Schlesselman 1978^5^, requires no triple interaction assumption (essentially no interaction between the exposure and the confounder) and parameters including ${RR}_{UY}$, P(U\|X=1) and P(U\|X=0). The maximum bias B, proposed by Ding and VanderWeele 2006^49^, requires no assumption and only two parameters ${RR}_{UY}$ and ${RR}_{XU}.$ The Cusson and Infante-Rivard 2020^50^ method extends the E-value. The E-value primarily focuses on RRs as effect measures and assumes that ${RR}_{UY}$ = ${RR}_{XU}$. The method by Cusson and Infante-Rivard 2020^50^ develops new E-value formulas for situations when different ${RR}_{UY}$ and ${RR}_{XU}$values are present, and when ORs are the effect measures.  2. One unstated assumption, as it was in the original E-value paper, is that the prevalence of the confounder among the exposed is 100%.^53^ |

#### Mathur and VanderWeele 2020: sensitivity analysis for unmeasured confounding in meta-analyses

| **Name/description** | **Method for unmeasured confounding in meta-analyses** |
| --- | --- |
| **DOI** | 10.1080/01621459.2018.1529598 |
| **Year** | Mathur and VanderWeele 2020^56^ |
| **Crossed-referenced in QBA textbook^3^** | Not referenced |
| **QBA classification** | Simple sensitivity analysis |
| **Applicable study design scenarios described^b^** | Random-effects meta-analyses |
| **Effect measure of interest** | 1. Sensitivity analysis for the proportion of meaningfully strong effects  summary RR, OR, MD, log-RR or log-OR  2. Sensitivity analysis for the point estimate: pooled RR, OR, HR or MD |
| **Result of interest** | Explain-away |
| **Data structure/format of the eligible exposure/intervention^b^** | Categorical |
| **Data structure/format of the confounders, if applicable^b^** | Categorical |
| **Data structure/format of eligible outcome^b^** | Categorical, or time-to-event |
| **Source(s) of bias addressed** | Unmeasured confounding (single or multiple) |
| **Information/parameters needed to conduct the QBA method** | **1. Sensitivity analysis for the proportion of meaningfully strong effects**  - ${\hat{\boldsymbol{y}}}_{\boldsymbol{R}}^{\boldsymbol{c}}\boldsymbol{:}$ the summary effect estimate from a meta-analysis (on the log or non-log scale);  - $\boldsymbol{q}$: threshold for meaningfully strong effect size on the same scale of $\hat{y}_{R}^{c}$;  - **r**: proportion below which strong effects are to be reduced;  - $\boldsymbol{\mu}\boldsymbol{B}^{\mathbf{*}}$: mean bias factor across studies on the same scale of $\hat{y}_{R}^{c}$;  - $\boldsymbol{\tau}_{\boldsymbol{c}}^{\mathbf{2}}$: heterogeneity estimate from a meta-analysis;  - $\boldsymbol{\sigma}_{\boldsymbol{B}^{\mathbf{*}}}^{\mathbf{2}}$: proportion of heterogeneity due to variation in confounding bias.  **2. Sensitivity analysis for the point estimate**  - ${\hat{\boldsymbol{y}}}_{\boldsymbol{R}}^{\boldsymbol{c}} :$ the summary effect estimate from a meta-analysis (on the log or non-log scale). |
| **Mathematical formula** | When confounded pooled RR >1  **1. Sensitivity analysis for the proportion of meaningfully strong effects:** $\hat{p}(q)=1-\Phi\left( \frac{q+\mu B^{*}-\hat{y}_{R}^{c}}{\sqrt{\tau_{c}^{2}-\sigma_{B^{*}}^{2}}} \right),\tau_{c}^{2}>\sigma_{B^{*}}^{2}$;  $\hat{T}(r,q)=exp\left\{ \Phi^{-1}(1-r)\sqrt{\tau_{c}^{2}}-q+\hat{y}_{R}^{c} \right\}$;  $\hat{G}(r,q)=\hat{T}(r,q)+\sqrt{(\hat{T}(r,q))^{2}-\hat{T}(r,q)}$ ;  **2. Sensitivity analysis for the point estimate:**  E-value for the point estimate=$exp\left( \hat{y}_{R}^{c} \right)+\sqrt{exp\left( \hat{y}_{R}^{c} \right)\left[ exp\left( \hat{y}_{R}^{c} \right)-1 \right]}$.  *Note: 1. Φ denotes the standard normal cumulative distribution function; 2. Formulas for confounded RR <1 can be found in the paper; 3. E-value formula can also be applied to 95% CIs.* |
| **Data assumptions and additional required features** | **For both sensitivity analysis for the proportion of meaningfully strong effects, and sensitivity analysis for the point estimate:**  a. The bias factor is assumed to be normally distributed across studies or is assessed across a range of fixed values.  b. The bias factor is independent of the true effects but not the confounded effects.  c. Estimated mean and estimated heterogeneity are consistent and unbiased, asymptotically normal, and asymptotically independent;  d. The point estimates’ expectations are independent of their standard errors;  e. There are approximately 10 or more studies included in a meta-analysis. |
| **Available and/or recommended software** | Online tool (https://www.evalue-calculator.com/meta/) or R package Evalue. |
| **Output of the QBA analysis** | 1. $\hat{p}(q)$, $\hat{T}(r,q)$, $\hat{G}(r,q)$;  2. E-value for the point estimate. |
| **Interpretation of the QBA analysis output** | 1. $\hat{p}(q)$: the proportion of studies with true effect sizes surpassing a threshold q;  $\hat{T}(r,q)$: the minimum bias to reduce to less than r the proportion of studies with population causal effects above q;  $\hat{G}(r,q)$: the minimum confounding association strength in all studies that would be required to reduce to below a threshold r the proportion of studies with effect sizes above q;  2. E-value for the point estimate: the minimum confounding strength, on average across the studies in a meta-analysis, to completely shift the pooled point estimate to the null. |
| **Considerations related to the interpretation of the results** | 1. The authors noted the interpretation for a large $\hat{p}(q)$: “a large proportion of true effect sizes stronger than a threshold of scientific importance in a meta-analysis suggests that, although the true causal effects may be heterogeneous across studies, there is evidence that overall, many of these effects are strong enough to merit scientific interest.”  2. The authors noted that recommendations on how to assign values to bias parameters $q$, r, $\mu B^{*}$, $\sigma_{B^{*}}^{2}$ can be found in the paper and other literature.^57^  3. The authors noted that a large value of either $\hat{T}(r,q)$, $\hat{G}(r,q)$ could indicate that it would take substantial unmeasured confounding to “explain away” the results of the meta-analysis in this sense, and that weaker unmeasured confounding could not do so. Thus, the results may be considered relatively robust to unmeasured confounding.  4. The authors noted that “If a well-designed meta-analysis yields a low value of $\hat{T}(r,q)$, and $\hat{G}(r,q)$ and thus, is relatively sensitive to unmeasured confounding, this indicates that future research on the topic should prioritize randomized trials or designs and data collection that reduce unmeasured confounding.”  5. Although individual studies included in the meta-analyses are sometimes not robust to confounding bias, the conclusions of the meta-analysis may in fact be robust to moderate degrees of unmeasured confounding.  6. Formulas for effect measures other than RR are discussed in the full manuscript and in other articles.^48^ |
| **Explicit discussions regarding the similarities and differences with other methods** | 1. Alternatively, one can use Ding and VanderWeele^49^’s bounding formula on each individual study to compute the proportion of studies with effect sizes more extreme than q given a particular bias factor. However, this approach has several limitations noted in the paper.  2. This approach subsumes several earlier approaches (Cornfield et al 1959^7^; Schlesselman’s 1978^5^; Flanders and Khoury 1990)^10^ and, in contrast to Lin et al 1998^6^ and certain results of VanderWeele and Arah 2011^11^, this does not make any no-interaction assumptions between the exposure and the unmeasured confounder(s). |

#### Mathur and VanderWeele 2020: robust methods for point estimation and inference in meta-analyses

| **Name/description** | **Robust methods for point estimation and inference in meta-analyses** |
| --- | --- |
| **DOI** | 10.1097/EDE.0000000000001180 |
| **Year** | Mathur and VanderWeele 2020^58^ |
| **Crossed-referenced in QBA textbook^3^** | Not referenced |
| **QBA classification** | Simple sensitivity analysis |
| **Applicable study design scenarios described^b^** | Random-effects meta-analyses |
| **Effect measure of interest** | summary RR, OR, MD, log-RR or log-OR |
| **Result of interest** | Explain-away |
| **Data structure/format of the eligible exposure/intervention^b^** | Categorical or continuous |
| **Data structure/format of the confounders, if applicable^b^** | Categorical or continuous |
| **Data structure/format of eligible outcome^b^** | Categorical or continuous |
| **Source(s) of bias addressed** | Unmeasured confounding (single or multiple) |
| **Information/parameters needed to conduct the QBA method** | **Calibrated estimation method for the bias-corrected estimates and explain-away metrics:**  - $\hat{\boldsymbol{\mu}}$ , ${\hat{\boldsymbol{\tau}}}^{\mathbf{2}}$: the classical Dersimonian–Laird meta-analytic estimates of the mean and variance of the true population effects;  -${\hat{\boldsymbol{\theta}}}_{\boldsymbol{i}}$: the point estimate in study i;  - ${\hat{\boldsymbol{\sigma}}}_{\boldsymbol{i}}$: the estimated standard error of study i;  - $\boldsymbol{q}$: threshold for meaningfully strong effect size on the same scale of${\hat{\boldsymbol{\theta}}}_{\boldsymbol{i}}$. |
| **Mathematical formula** | **Procedures for the calibrated estimation method***  1. Choose a bias factor B:  $\mathrm{bias} factor B\leq\frac{\mathrm{RR}_{EU}\times\mathrm{RR}_{UD}}{\mathrm{RR}_{EU}+\mathrm{RR}_{UD}-1}$,  where $\mathrm{RR}_{EU}$ is the observed RR for the exposure and unmeasured confounder; $\mathrm{RR}_{UD}$ is the observed RR for the unmeasured confounder and outcome; more in Ding and VanderWeele 2016.^49^  2. Calculate the calibrated point estimate for an individual study i on the $\log$ relative risk scale:  $\hat{\theta}_{i}^{'}=\hat{\theta}_{i}-logB$ if $\hat{\mu}>0$ (i.e., the confounded pooled point estimate on the log relative risk scale is apparently causative)  or as $\hat{\theta}_{i}^{'}=\hat{\theta}_{i}+logB$ if $\hat{\mu}<0$ (i.e., the confounded pooled point estimate is apparently preventive).  3. Calculate the bias-corrected pooled point estimate:  $\hat{\mu}^{'}=\hat{\mu}-logB$ if $\hat{\mu}>0$ and as $\hat{\mu}^{'}=\hat{\mu}+logB$ if $\hat{\mu}<0$.  4. Calculate the bias-corrected calibrated estimates on the log relative risk scale:  $\tilde{\theta}_{i}^{'}=\hat{\mu}^{'}+\sqrt{\hat{\tau}^{2}/\left( \hat{\tau}^{2}+\hat{\sigma}_{i}^{2} \right)}\left( \hat{\theta}_{i}^{'}-\hat{\mu}^{'} \right)$. 5. Calculate the proportion of unconfounded effects stronger than q when all studies have bias factor B:  $\hat{P}_{>q}(B)=\hat{P}\left( \tilde{\theta}_{i}^{'}>q \right)$.  6. Bootstrap pairs of $\left( \hat{\theta}_{i}^{'},\hat{\sigma}_{i} \right)$ by drawing with replacement from the original sample and estimating in turn $\hat{\mu}^{'}$ and $\hat{\tau}^{2},\tilde{\theta}_{i}^{'}$ for each study, and finally $\hat{P}_{>q}(B)$. Bootstrapping is a suggested but optional step.  7. Construct a bias-corrected and accelerated (BCa) confidence interval from the bootstrapped values of $\hat{P}_{>q}(B)$.  8. Calculate two explain-away metrics $\hat{T}(r,q)$, $\hat{G}(r,q)$:  $\hat{T}(r,q)$ is defined a bias factor such that $\hat{P}_{>q}(B)$ is exactly equal to the chosen proportion $r$; that is, $\hat{T}(r,q)=B:\hat{P}_{>q}(B)=r$.  $\hat{G}(r,q)$ is a simple transformation of $\hat{T}(r,q)$, calculated as $\hat{G}(r,q)=\hat{T}(r,q)+\sqrt{(\hat{T}(r,q))^{2}-\hat{T}(r,q)}$.  9. Construct confidence intervals for $\hat{T}(r,q)$, $\hat{G}(r,q)$ via bootstrapping by resampling the bias-corrected point estimates for each study $\hat{\theta}_{i}^{'}$ and using the BCa method.  *Note: *The other method termed sign test method, which calculated* $\hat{P}_{>q}$*in another way, was detailed in the supplement of Mathur and VanderWeele 2020.^58^* *Based on the simulation results comparing the calibrated estimation method and sign test method, Mathur and VanderWeele noted that the calibrated estimation method “was usually the least biased for all distributions, though its root mean square error was sometimes higher than that of other methods.”. They recommended “…by default constructing the confidence interval by applying the bias-corrected and accelerated bootstrap as described above (“BCa-calibrated”)”.* |
| **Data assumptions and additional required features** | 1. There are approximately 10 or more studies included in a meta-analysis.  2. The bias factor is homogeneous across studies in a meta-analysis. As noted by the authors, “all studies are subject to the same degree of unmeasured confounding, albeit possibly due to different unmeasured confounders.”. |
| **Available and/or recommended software** | Online tool (https://www.evalue-calculator.com/meta/) or R package MetaUtility (function prop_stronger). |
| **Output of the QBA analysis** | Point estimates and confidence intervals for $\hat{P}_{>q}\left( B \right), \hat{T}(r,q)$, $\hat{G}(r,q)$. |
| **Interpretation of the QBA analysis output** | $\hat{P}_{>q}\left( B \right)$: the proportion of studies with true effect sizes surpassing a threshold q when all studies have bias factor B;  $\hat{T}(r,q)$: the minimum bias to reduce to less than r the proportion of studies with population causal effects above q;  $\hat{G}(r,q)$: the minimum confounding association strength in all studies that would be required to reduce to below a threshold r the proportion of studies with effect sizes above q. |
| **Considerations related to the interpretation of the results** | 1. The authors noted that “relatively imprecise estimates $\hat{\theta}_{i}$ (i.e., those with large $\hat{\sigma}_{i}$) receive strong shrinkage toward $\hat{\mu}$, while relatively precise estimates receive less shrinkage and remain closer to their original values.”  2. For the calibrated estimation method, the authors proposed the proportion of scientifically meaningful effect sizes as the sample proportion of calibrated estimates *above* q, and noted that “analogous methods can be used to estimate the proportion of effects *below* another threshold.”  3. One limitation that the authors noted was that “the BCa-calibrated method did sometimes lose considerable precision compared with the parametric method in certain scenarios in which the latter achieved approximately nominal coverage, so for large meta-analyses with apparently normal effects and in which $\hat{P}_{>q}$ is close to 0.50, one might reasonably choose to substitute the parametric interval for the default BCa-calibrated interval.”.  4. The authors noted that the other limitation of the method is that “the BCa-calibrated interval may sometimes fail to converge for small meta-analyses”. |
| **Explicit discussions regarding the similarities and differences with other methods** | 1. The authors noted that another similar analytic method using parametric methods was Mathur and VanderWeele 2020.^56^  2. The authors noted that “Wang and Lee^59^ demonstrated that the calibrated estimates can be used to construct approximately unbiased prediction intervals for small meta-analyses and for non-normal true effect distributions.” |

#### MacLehose et al 2021: generalized E-value (G-value)

| **Name/description** | **Generalized E-value (G-value)** |
| --- | --- |
| **DOI** | 10.1097/EDE.0000000000001381 |
| **Author and Year** | MacLehose et al 2021^53^ |
| **Crossed-referenced in QBA textbook^3^** | Referenced and explained |
| **QBA classification** | Simple sensitivity analysis |
| **Applicable study design scenarios described^b^** | Any observational study designs |
| **Effect measure of interest** | RR, OR, HR, or RD |
| **Result of interest** | Explain-away |
| **Data structure/format of the eligible exposure/intervention^b^** | Categorical |
| **Data structure/format of the confounders, if applicable^b^** | Categorical |
| **Data structure/format of eligible outcome^b^** | Categorical, or time-to-event data |
| **Source(s) of bias addressed** | Unmeasured confounding (single or multiple) |
| **Information/parameters needed to conduct the QBA method** | - **RR_obs_** (and 95% CIs): observed RR (and 95% CIs);  - **p_1_**: prevalence of confounder among the exposed. |
| **Mathematical formula** | G-value $\left( p_{1} \right)=\frac{RR_{obs}+p_{1}+RR_{obs}p_{1}-1+\sqrt{\begin{matrix} \left( 1-p_{1}-R_{obs}-R_{obs}p_{1} \right)^{2}-4R_{obs}p_{1}^{2} \\ \end{matrix}}}{2p_{1}}$. |
| **Data assumptions and additional required features** | When calculating the G-value, RR_eu_= RR_ud_ >1.  *Note: RR_eu_: the association between exposure and unmeasured confounder on a risk ratio scale; RR_ud_: the association between unmeasured confounder and outcome on a risk ratio scale* |
| **Available and/or recommended software** | Not mentioned |
| **Output of the QBA analysis** | Generalized E-value, or also known as G-value |
| **Interpretation of the QBA analysis output** | When the prevalence of confounder among exposed is p_1_, the output is the minimum strength of association, on the risk ratio scale, that an unmeasured confounder would need to have with both the treatment and outcome to fully explain away a specific treatment–outcome association, conditional on the measured covariates. |
| **Considerations related to the interpretation of the results** | 1. When p_1_=1, G-value=E-value; otherwise, when p_1_<1, G-value>E-value.  2. The authors noted that it is possible to derive a G-formula under more general conditions (e.g., such as a nonbinary confounder).  3. The authors noted that like the E-value method, the G value method is defined for RR_obs_ >1 and reverse coding can be used to accommodate observed effects less than 1.  4. The authors noted that unlike E-value, which provides a bound on the bias, G-value gives the exact size that the confounder associations would need to have to completely nullify the observed association between exposure and outcome. |
| **Explicit discussions regarding the similarities and differences with other methods** | 1. The G-value formula was derived using Schlesselman 1978^5^’s formula.  2. G-value extends the E-value method by VanderWeele and Ding 2017^48^, by removing the assumption that the prevalence of the uncontrolled confounder among the exposed is 100%. |

### Misclassification bias (exposure misclassification bias)

#### Copeland et al 1977 method for misclassification bias in the estimation of relative risk

| **Name/description** | **Method for misclassification bias in the estimation of relative risk** |
| --- | --- |
| **DOI** | 10.1093/oxfordjournals.aje.a112408 |
| **Year** | Copeland et al 1977^60^ |
| **Crossed-referenced in QBA textbook^3^** | Referenced but not explained |
| **QBA classification** | Simple sensitivity analysis |
| **Applicable study design scenarios described^b^** | Cohort or case-control studies |
| **Effect measure of interest** | RR, and OR |
| **Result of interest** | Corrected estimate |
| **Data structure/format of the eligible exposure/intervention^b^** | Categorical |
| **Data structure/format of the confounders, if applicable^b^** | Not mentioned |
| **Data structure/format of eligible outcome^b^** | Categorical |
| **Source(s) of bias addressed** | Misclassification of study subjects (i.e., disease occurrence in cohorts or exposure history in case-control studies, differential or nondifferential) |
| **Information/parameters needed to conduct the QBA method** | **-** $\boldsymbol{a}^{\mathbf{'}}$: the observed number of study subjects that are exposed and have the outcome (i.e., the cells in a 2 by 2 table);  **-** $\boldsymbol{c}^{\mathbf{'}}$: the observed number of study subjects that are not exposed and have the outcome (i.e., the cells in a 2 by 2 table);  **-** $\boldsymbol{N}_{\mathbf{0}}$: total number of unexposed;  **-** $\boldsymbol{N}_{\mathbf{1}}$: total number of exposed;  **-** $\boldsymbol{Sp}$: specificity of classification procedure (i.e., disease occurrence in cohorts or exposure history in case-control studies). |
| **Mathematical formula** | $RR=\frac{\left[ a^{'}-N_{1}(1-Sp) \right]/N_{1}}{\left[ c^{'}-N_{0}(1-Sp) \right]/N_{0}}$ |
| **Data assumptions and additional required features** | 1. The misclassification affects only the comparison criterion—that is, disease occurrence in cohort or follow-up studies, and exposure history in case-control studies.  2. The misclassified variable is dichotomous.  3. There is no bias due to selection or confounding in the data from which the estimates are calculated, thereby allowing the use of simple, unadjusted ratio measures of effect. |
| **Available and/or recommended software** | Not mentioned |
| **Output of the QBA analysis** | Corrected estimate |
| **Interpretation of the QBA analysis output** | Corrected effect estimate that accounts for misclassification bias. |
| **Considerations related to the interpretation of the results** | Not mentioned |
| **Explicit discussions regarding the similarities and differences with other methods** | Not mentioned |

#### Barron 1977 method for misclassification for relative risks

| **Name/description** | Method for misclassification for relative risks |
| --- | --- |
| **DOI** | https://doi.org/10.2307/2529795 |
| **Year** | Barron 1977^61^ |
| **Crossed-referenced in QBA textbook^3^** | Referenced but not explained |
| **QBA classification** | Simple sensitivity analysis |
| **Applicable study design scenarios described^b^** | Cohort studies |
| **Effect measure of interest** | RRs |
| **Result of interest** | Corrected effect estimate |
| **Data structure/format of the eligible exposure/intervention^b^** | Categorical |
| **Data structure/format of the confounders, if applicable^b^** | Not applicable |
| **Data structure/format of eligible outcome^b^** | Categorical |
| **Source(s) of bias addressed** | Exposure and/or outcome misclassification (differential) |
| **Information/parameters needed to conduct the QBA method** | - **p_11_**: probability of an individual classified as having both A and B;  - **p_10_**: probability of an individual classified as having A but not B;  - **p_01_**: probability of an individual classified as having B but not A;  - **p_00_**: probability of an individual classified as having neither A nor B.  *Notes: A: exposure; B: outcome.* |
| **Mathematical formula** | the corrected RR: $R_{p}=\left[ \frac{p_{11}}{p_{10}} \right]\left[ \frac{p_{10}+p_{00}}{p_{11}+p_{01}} \right]$.  *Notes: If the probabilities of systematically misclassifying a given element of the sample with respect to A and B are known, formula with matrix involved can be used which were presented in the section 2 of the paper as formula (1).* |
| **Data assumptions and additional required features** | It is assumed that a study sample is drawn from the population and each element of the sample is classified according to the presence or absence of A and B. |
| **Available and/or recommended software** | Not mentioned |
| **Output of the QBA analysis** | R_p_ |
| **Interpretation of the QBA analysis output** | The RR corrected for misclassification |
| **Considerations related to the interpretation of the results** | Not mentioned |
| **Explicit discussions regarding the similarities and differences with other methods** | This method is flexible in the sense that it allows simultaneous adjustment of misclassification of exposure and outcome.^3^ |

#### Greenland 1982 method for misclassification bias in matched-pair studies

| **Name/description** | **Method for misclassification bias in matched-pair studies** |
| --- | --- |
| **DOI** | 10.1093/oxfordjournals.aje.a113424 |
| **Year** | Greenland 1982^62^ |
| **Crossed-referenced in QBA textbook^3^** | Not referenced |
| **QBA classification** | Simple sensitivity analysis |
| **Applicable study design scenarios described^b^** | Matched-pair case control studies |
| **Effect measure of interest** | OR |
| **Result of interest** | Corrected estimate |
| **Data structure/format of the eligible exposure/intervention^b^** | Categorical |
| **Data structure/format of the confounders, if applicable^b^** | Not mentioned |
| **Data structure/format of eligible outcome^b^** | Categorical |
| **Source(s) of bias addressed** | Exposure misclassification (nondifferential) |
| **Information/parameters needed to conduct the QBA method** | - **Fn_1_:** false negative rate among cases;  - **Fp_1_:** false positive rate among cases;  - **Fn_0_:** false negative rate among controls;  - **Fp_0_:** false positive rate among controls.  - **Tp_1_:** true positive rate (sensitivity) among cases, **Tp_1_** = Pr (exposed case is correctly classified) = 1-**Fn1**;  - **Tn_1_:** true negative rate (specificity) among cases, **Tn_1_** = Pr (unexposed case is correctly classified) = 1-**Fp_1_**;  - **Tp_0_:** true positive rate among controls, **Tp0** = Pr (exposed control is correctly classified) = 1-**Fn_0_**;  - **Tn_0_:** true negative rate among controls, **Tn_0_** = Pr (unexposed control is correctly classified) = 1-**Fp_0_**.  - **T’, U’, V’, W’**: observed counts in 2 by 2 table  *Notes: T, U, V, W: true counts in 2 by 2 table (see Table 1 for details)* |
| **Mathematical formula** | $\begin{matrix} T^{'}= & Tp_{1}Tp_{0}T+Tp_{1}Fp_{0}U+Fp_{1}Tp_{0}V; \\ & +Fp_{1}Fp_{0}W, \\ U^{'}= & Tp_{1}Fn_{0}T+Tp_{1}Tn_{0}U+Fp_{1}Fn_{0}V; \\ & +Fp_{1}Tn_{0}W, \\ V^{'}= & Fn_{1}Tp_{0}T+Fn_{1}Fp_{0}U+Tn_{1}Tp_{0}V; \\ & +Tn_{1}Fp_{0}W, \\ \text{ and } & \\ W^{'}= & Fn_{1}Fn_{0}T+Fn_{1}Tn_{0}U+Tn_{1}Fn_{0}V \\ & +Tn_{1}Tn_{0}W. \end{matrix}$  $corrected OR: \hat{OR}=\frac{\hat{U}}{\hat{V}}$*.*  *Notes:* $\hat{U}and \hat{V}$*: estimated U and V.* |
| **Data assumptions and additional required features** | If the classification rates Tp_1_, Tn_1_, Tp_0_, and Tn_0_ can be estimated with some degree of accuracy, their estimated values and the observed values of T’, U’, V, and W can be substituted into the above formulae, resulting in a set of four linear equations in the four unknowns, T, U, V, and W. |
| **Available and/or recommended software** | Not mentioned |
| **Output of the QBA analysis** | Corrected OR |
| **Interpretation of the QBA analysis output** | OR adjusted for misclassification (OR_t_) |
| **Considerations related to the interpretation of the results** | The authors noted that “If the classification rates are only known with error (as is usually the case), these corrections should be regarded with caution in direct proportion to the uncertainty in the estimated classification rates.” |
| **Explicit discussions regarding the similarities and differences with other methods** | The authors mentioned that Copeland et al 1977^60^ provided equations for unmatched studies, while the focus of this method was on matched studies.^62^ |

#### Greenland and Kleinbaum 1983 generalizations of Barron’s matrix correction method

| **Name/description** | **Generalizations of Barron’s matrix correction method** |
| --- | --- |
| **DOI** | 10.1093/ije/12.1.93 |
| **Year** | Greenland and Kleinbaum 1983^63^ |
| **Crossed-referenced in QBA textbook^3^** | Referenced but not explained |
| **QBA classification** | Simple sensitivity analysis |
| **Applicable study design scenarios described^b^** | Case-control studies^b^ with 2 by 2 table (in **Method 1**) or matched-pair case control studies (in **Method 2**) |
| **Effect measure of interest** | OR |
| **Result of interest** | Corrected estimate |
| **Data structure/format of the eligible exposure/intervention^b^** | Categorical |
| **Data structure/format of the confounders, if applicable^b^** | Not mentioned |
| **Data structure/format of eligible outcome^b^** | Categorical^b^ |
| **Source(s) of bias addressed** | Exposure and/or outcome misclassification (differential or nondifferential) |
| **Information/parameters needed to conduct the QBA method** | **Method 1**:  - **m:** misclassified cells stratified by exposure and outcome status;  - **P_rxy_:** probability that study subject with true value x of X and y of Y is classified as having value r for X;  - **Q_syx_**: probability that study subject with true value y of Y and x of X is classified as having value s for Y.  *Notes: X: exposure; Y: outcome; Prxy and/or Qsyx will be used to construct correction matrix C.*  **Method 2**:  - **m**: misclassified cells stratified by exposure and outcome status;  - **P_rxy_**: probability that pair member 1 is classified as having response value r;  - **Q_syx_**: probability that pair member 2 is classified as having response value s.  *Notes: X: response value of the first pair-member; Y: response value of the second pair-member; Prxy and/or Qsyx will be used to construct correction matrix C.* |
| **Mathematical formula** | $m_{rs}=\sum_{x,y} P_{rxy}Q_{syx}t_{xy}$;  These equations can be compactly written in matrix form, m = Ct,  If C is invertible, a correction formula can be derived immediately from equation,  t= C^-1^ m.  *Notes: C: correction matrix; m: misclassified variables; t: true variables.* |
| **Data assumptions and additional required features** | Misclassification of each variable is independent of misclassification of the other variables (i.e., the misclassification probabilities for a variable do not depend on whether or not misclassification took place for another variable). |
| **Available and/or recommended software** | Not mentioned |
| **Output of the QBA analysis** | Corrected OR |
| **Interpretation of the QBA analysis output** | OR adjusted for misclassification (OR_m_) |
| **Considerations related to the interpretation of the results** | 1. The authors noted that estimate of C must be derived either from earlier studies involving similar classification methods, or from a validation study carried out on a subsample of the study subjects at hand.  2. The authors noted that C will vary from study to study, especially when considering questionnaire items administered to separate populations at separate times.  3. The authors noted that when classification of one of both variables takes place before sampling, C will depend heavily on the study design, and a fallacy in estimating C from data external to the study can arise.  4. The authors noted that complete validation is rarely possible, and corrected estimates should not be viewed as being fully corrected.  *Note: the study also provided a valid correction of C if the ratio R of the case-control sampling fractions is known.* |
| **Explicit discussions regarding the similarities and differences with other methods** | Previous methods on misclassification including Barron 1977^61^ and Copeland et al 1977^60^, had different assumptions that misclassification of each variable was nondifferential; and misclassification of each variable was independent of misclassification of the other variables. None of the methods dealt with situation of pair-matched variable. |

#### Marshall 1990 method for using predictive values for misclassification

| **Name/description** | **Method for using predictive values for misclassification** |
| --- | --- |
| **DOI** | 10.1016/0895-4356(90)90077-3 |
| **Year** | Original manuscript: Marshall 1990^64^. Additional information: Fox et al 2021.^3^ This textbook provides a clear overview of the methods and was used to populate the cells below. |
| **Crossed-referenced in QBA textbook^3^** | Referenced and explained |
| **QBA classification** | Direct bias modeling method |
| **Applicable study design scenarios described^b^** | Case-control studies with validation data |
| **Effect measure of interest** | OR |
| **Result of interest** | Corrected estimate |
| **Data structure/format of the eligible exposure/intervention^b^** | Categorical |
| **Data structure/format of the confounders, if applicable^b^** | Not mentioned |
| **Data structure/format of eligible outcome^b^** | Categorical |
| **Source(s) of bias addressed** | Exposure misclassification (differential or nondifferential) |
| **Information/parameters needed to conduct the QBA method** | - **NPV_i_**: negative predictive value among cases (i=1) and controls (i=0);  - **PPV_i_**: positive predictive value among cases (i=1) and controls (i=0);  - **P(e_i_) and Var(P(e_i_))**: prevalence of self-reported exposure among cases (i=1) and controls (i=0), and its variance. |
| **Mathematical formula** | Step 1: Calculate $P\left( E_{i} \right)=PPV_{i}P\left( e_{i} \right)+\left( 1-NPV_{i} \right)\left( 1-P\left( e_{i} \right) \right)$  where $P\left( E_{i} \right)$ is the proportion of cases (i=1) or controls (i=0) that are truly exposed;  Step 2: obtain the bias-adjusted odds ratio (OR_b)_ and corresponding variance (ln(OR_b_)):^3^  $OR_{b}=\frac{\frac{P\left( E_{1} \right)}{1-P\left( E_{1} \right)}}{\frac{P\left( E_{0} \right)}{1-P\left( E_{0} \right)}}$;  $Var\left[ ln\left( OR_{b} \right) \right]=\frac{a_{0}}{\left[ P\left( E_{0} \right)\left( 1-P\left( E_{0} \right) \right) \right]^{2}}+\frac{a_{1}}{\left[ P\left( E_{1} \right)\left( 1-P\left( E_{1} \right) \right) \right]^{2}}$  where $a_{i}=\left[ PPV_{i}-\left( 1-NPV_{i} \right) \right]^{2}\mathrm{Var} \left[ P\left( e_{i} \right) \right]+\mathrm{Var} \left( PPV_{i} \right)P\left( e_{i} \right)^{2}+\mathrm{Var} \left( NPV_{i} \right)\left[ 1-P\left( e_{i} \right) \right]^{2};$  Step 3: Using the information from Step 1 and 2, calculate the upper and lower limits of the 95% CI of OR using:  $e^{\ln{OR}_{b}\pm1.96*\surd Var\left[ ln\left( OR_{b} \right) \right]}.$ |
| **Data assumptions and additional required features** | As noted in the Fox et al 2021 textbook,^3^ “Sampling error from a validation study is the source of uncertainty about the predictive values measured in the validation study.” |
| **Available and/or recommended software** | Not mentioned |
| **Output of the QBA analysis** | Corrected effect estimate and the corresponding 95% CI |
| **Interpretation of the QBA analysis output** | Effect estimate and corresponding 95% CI accounting for misclassification |
| **Considerations related to the interpretation of the results** | As noted in the Fox et al 2021 textbook,^3^ this method:  1. cannot easily be extrapolated to bias analysis for individual participant level data or multiple bias analysis;  2. “...can only be used in conjunction with OR and does not admit correlation between predictive probabilities.” |
| **Explicit discussions regarding the similarities and differences with other methods** | The author mentioned that Barron 1977^65^ was the most common and now fairly standard method, and it was also one of the two methods that the author intensively discussed in the paper. |

#### Checkoway et al 1991 method for nondifferential exposure misclassification

| **Name/description** | **Method for nondifferential exposure misclassification** |
| --- | --- |
| **DOI** | 10.1080/1047322X.1991.10387923 |
| **Year** | Checkoway et al 1991^66^ |
| **Crossed-referenced in QBA textbook^3^** | Not referenced |
| **QBA classification** | Simple sensitivity analysis |
| **Applicable study design scenarios described^b^** | Cohort studies^b^ |
| **Effect measure of interest** | RR |
| **Result of interest** | Corrected estimate |
| **Data structure/format of the eligible exposure/intervention^b^** | Categorical^b^ |
| **Data structure/format of the confounders, if applicable^b^** | Categorical |
| **Data structure/format of eligible outcome^b^** | Categorical^b^ |
| **Source(s) of bias addressed** | Exposure misclassification (nondifferential) |
| **Information/parameters needed to conduct the QBA method** | - **a’**: the observed value of exposed cases;  - **b’**: the observed value of nonexposed cases;  - **L**: the total number of cases;  - **N_0_’**: the observed person-years among the exposed;  - **N_1_’**: the observed person-years among the nonexposed;  - **S**: the common value for sensitivity and specificity;  - **T**: the total number of person-years.  *Notes: Please check table VI*^66^ *for more details.* |
| **Mathematical formula** | $a=\frac{L*S-b^{'}}{2S-1}$;$b=\frac{L*S-a^{'}}{2S-1};$ $N_{1}=\frac{T*S-N_{0}^{'}}{2S-1}$;$N_{0}=\frac{T*S-N_{1}^{'}}{2S-1}$;  $RR=\frac{\frac{a}{N_{1}}}{\frac{b}{N_{0}}}$.  *Notes: a, b, N_1_, N_0_ are the corrected versions of a’, b’, N_1_’, N_0_’.* |
| **Data assumptions and additional required features** | Not mentioned |
| **Available and/or recommended software** | Not mentioned |
| **Output of the QBA analysis** | Corrected effect estimate (e.g., RR, standardized mortality ratio (SMR)) |
| **Interpretation of the QBA analysis output** | Effect estimate accounting for nondifferential exposure misclassification |
| **Considerations related to the interpretation of the results** | 1. The authors noted that this approach can be refined by allowing for time-dependent variation in exposure misclassification or for more complex patterns of misclassification between exposure strata than those demonstrated here.  2. The authors noted that “The method described above for assessing bias resulting from misclassification of the main study exposure can be adapted to estimate the extent of residual confounding caused by misclassification of confounders.” |
| **Explicit discussions regarding the similarities and differences with other methods** | The author mentioned that this method was meant to be straightforward analytic methods with simplifying assumptions, focusing on nondifferential misclassification bias. More mathematical approaches can be found by Kaldor and Clayton 1985,^67^ and Espeland and Hui 1987.^68^ |

#### Wacholder et al 1993 sensitivity analysis for validation studies using an alloyed gold standard

| **Name/description** | **Sensitivity analysis for validation studies using an alloyed gold standard** |
| --- | --- |
| **DOI** | 10.1093/oxfordjournals.aje.a116627 |
| **Year** | Wacholder et al 1993^69^ |
| **Crossed-referenced in QBA textbook^3^** | Referenced but not explained |
| **QBA classification** | Simple sensitivity analysis |
| **Applicable study design scenarios described^b^** | Case-control and cohort studies with validation data |
| **Effect measure of interest** | RR, OR, and HR |
| **Result of interest** | Corrected estimate |
| **Data structure/format of the eligible exposure/intervention^b^** | Categorical, or continuous |
| **Data structure/format of the confounders, if applicable^b^** | Categorical, or continuous |
| **Data structure/format of eligible outcome^b^** | Categorical, or time-to-event data |
| **Source(s) of bias addressed** | Misclassification bias (nondifferential) |
| **Information/parameters needed to conduct the QBA method** | -${\hat{\boldsymbol{\beta}}}_{\boldsymbol{W}}$: the coefficient estimate of the regression of the outcome on the measured exposure;  - $\boldsymbol{r}_{\boldsymbol{zw}}^{\mathbf{2}}$: the correction factor from the validation study with the alloyed gold standard, which is the square of the correlation between Z and W.  Notes: W: measured exposure; Z: more sensitive and specific measure than W; X: the true value. |
| **Mathematical formula** | We define $\hat{\beta}_{W(Z)}$ as the estimate obtained by correcting $\beta_{W}$ using the squared correlation $r_{zw}^{2}$ from the validation study with the alloyed gold standard i.e.,  $\hat{\beta}_{W(Z)}=\frac{\hat{\beta}_{W}}{r_{zw}^{2}}$;  Then the formula can be used to covert beta coefficient to risk estimates (e.g., OR):  $OR=e^{\hat{\beta}_{W(Z)}}$. |
| **Data assumptions and additional required features** | 1. The putative gold standard is in fact measured with error.  2. For continuous variables, the distributions of the true exposure and errors are normally distributed. |
| **Available and/or recommended software** | Not mentioned |
| **Output of the QBA analysis** | Corrected effect estimate |
| **Interpretation of the QBA analysis output** | Effect estimate accounting for misclassification |
| **Considerations related to the interpretation of the results** | 1. The authors noted that “It is important to consider the likely direction of the correlation in designing and interpreting validation studies, since these quantities cannot be known or estimated without a measured gold standard.”  2. The authors also mentioned that independent errors between W and Z seem  likely when two different sources are used, such as self-report and medical records. |
| **Explicit discussions regarding the similarities and differences with other methods** | 1. The authors noted that “Rosner et al^70^ appears to be the most robust against use of alloyed instead of actual gold standards, as it does give an unbiased estimate when the errors in the two measures being compared in the validation study are independent.”  2. The authors pointed out other correction methods when a gold standard is absent as long as an assumption of independent errors is made. |

#### Marshall 1994 quality indices for exposure misclassification

| **Name/description** | **Quality indices for exposure misclassification** |
| --- | --- |
| **DOI** | 10.1097/00001648-199405000-00009 |
| **Year** | Marshall 1994^71^ |
| **Crossed-referenced in QBA textbook^3^** | Not referenced |
| **QBA classification** | Simple sensitivity analysis |
| **Applicable study design scenarios described^b^** | Case-control studies |
| **Effect measure of interest** | OR |
| **Result of interest** | Corrected estimates |
| **Data structure/format of the eligible exposure/intervention^b^** | Categorical |
| **Data structure/format of the confounders, if applicable^b^** | Not mentioned |
| **Data structure/format of eligible outcome^b^** | Categorical |
| **Source(s) of bias addressed** | Exposure misclassification (differential or nondifferential) |
| **Information/parameters needed to conduct the QBA method** | - **A, B, C, D**: four fundamental measures for cases, as defined as $A=$ $Pr(X,E),B=Pr(\bar{X},E),C=Pr(X,\bar{E})$, and $D=Pr(\bar{X},\bar{E})$*;  - **A’, B’, C’, D’**: the corresponding control probabilities as A, B, C, D in case population**;**  -$\boldsymbol{OR}_{\boldsymbol{X}}$: odds ratio with respect to the measured exposure.  *Note:* $X$ *and* $\bar{X}$ *denote presence and absence, respectively, of measured exposure;* $E$ *and* $\bar{E}$ *denote presence and absence, respectively, of actual exposure; *: alternatively, sensitivity and specificity or positive predictive value and negative predictive value can be used (details see original paper).* |
| **Mathematical formula** | $\mathrm{OR}_{E}=bias factor*{OR}_{X}$;  $where bias factor=\frac{Q_{0}Q_{1}^{'}}{Q_{0}^{'}Q_{1}}$ can be computed as follows:  $Q_{1}=1-\frac{B}{(B+A)(B+D)}$*;  $Q_{0}=1-\frac{C}{\left( C+A \right)\left( C+D \right)}*$;  $Q_{1}^{'}=1-\frac{B^{'}}{(B^{'}+A^{'})(B^{'}+D^{'})}$*;  $Q_{0}^{'}=1-\frac{C^{'}}{\left( C^{'}+A^{'} \right)\left( C^{'}+D^{'} \right)}*$.  *Note: *: alternatively, sensitivity and specificity or positive predictive value and negative predictive value can be used to construct* quality indexes $Q_{1}, Q_{0}, Q_{1}^{'}, Q_{0}'$*(for details see original paper).* |
| **Data assumptions and additional required features** | The information on true exposure may be obtained by subsample validation study. |
| **Available and/or recommended software** | Not mentioned |
| **Output of the QBA analysis** | $\mathrm{OR}_{E}$ and quality indices |
| **Interpretation of the QBA analysis output** | $\mathrm{OR}_{E}:$Odds ratio with respect to the actual exposure.  Quality indices: rescaled sensitivity and specificity indices, which are based directly on the probabilities of disagreement between measured and true exposure. They are symmetrical with respect to measured and true exposure variables. |
| **Considerations related to the interpretation of the results** | 1. The author noted that if quality indices are nondifferential, that is, if $Q_{1}=$ $Q_{1}^{'}$ and $Q_{0}=Q_{0}^{'}$, or case quality indices are proportional to corresponding control indices, that is, if $Q_{0}=cQ_{0}^{'}$ and $Q_{1}=cQ_{1}^{'}$ for a constant c, then ${OR}_{X}={OR}_{E}.$ |
| **Explicit discussions regarding the similarities and differences with other methods** | Not mentioned |

#### Brenner 1996 alloyed gold standard for exposure misclassification

| **Name/description** | **Alloyed gold standard for exposure misclassification** |
| --- | --- |
| **DOI** | 10.1097/00001648-199607000-00011 |
| **Year** | Brenner 1996^72^ |
| **Crossed-referenced in QBA textbook^3^** | Not referenced |
| **QBA classification** | Simple sensitivity analysis |
| **Applicable study design scenarios described^b^** | Case-control studies^b^ with validation data |
| **Effect measure of interest** | OR |
| **Result of interest** | Corrected estimates |
| **Data structure/format of the eligible exposure/intervention^b^** | Categorical |
| **Data structure/format of the confounders, if applicable^b^** | Not mentioned |
| **Data structure/format of eligible outcome^b^** | Categorical |
| **Source(s) of bias addressed** | Exposure misclassification (differential or nondifferential) |
| **Information/parameters needed to conduct the QBA method** | - **P**: exposure prevalence in the population;  - **Se_as_** (**Sp_as_**) and **Se_rm_** (**Sp_rm_**): the sensitivity (specificity) of exposure measurement by the alloyed gold standard (AS) from validation data and the routinely employed method (RM) from routinely-collected data. |
| **Mathematical formula** | The authors proposed formula for two correlation structures:  1. Independence of classification by AS and RM conditional on true exposure status:  $Se_{ind}=\frac{P\cdot Se_{as}\cdot Se_{rm}+(1-P)\cdot\left( 1-Sp_{as} \right)\cdot\left( 1-Sp_{rm} \right)}{P\cdot Se_{as}+(1-P)\cdot\left( 1-Sp_{as} \right)}$;  $Sp_{\text{ind }}=\frac{P\cdot\left( 1-Se_{as} \right)\cdot\left( 1-Se_{rm} \right)+(1-P)\cdot Sp_{as}\cdot Sp_{rm}}{P\cdot\left( 1-Se_{as} \right)+(1-P)\cdot Sp_{as}}$.  2. Maximum possible positive correlation of classification errors given the individual misclassification rates:  $Se_{pos}=\frac{P\cdot Se_{rm}+(1-P)\cdot\left( 1-Sp_{as} \right)}{P\cdot Se_{as}+(1-P)\cdot\left( 1-Sp_{as} \right)}$;  $Sp_{pos}=\frac{P\cdot\left( 1-Se_{as} \right)+(1-P)\cdot Sp_{rm}}{P\cdot\left( 1-Se_{as} \right)+(1-P)\cdot Sp_{as}}.$ |
| **Data assumptions and additional required features** | A validation study is carried out in which routinely employed measures of a binary exposure variable are validated by an alloyed gold standard in a group of study participants from a population. |
| **Available and/or recommended software** | Not mentioned |
| **Output of the QBA analysis** | Se_ind_ (Sp_ind_) and Se_pos_ (Sp_pos_) |
| **Interpretation of the QBA analysis output** | Se_ind_ (Sp_ind_): the sensitivity (specificity) of exposure measurement given independence of classification by AS and RM conditional on true exposure status;  Se_pos_ (Sp_pos_): the sensitivity (specificity) of exposure measurement given maximum possible positive correlation of classification errors. |
| **Considerations related to the interpretation of the results** | 1. The authors noted that “for both independent and positively correlated classification errors, apparent sensitivity and specificity not only depend on the individual misclassification rates of both classification methods but also on the prevalence of exposure in the sample.”  2. The authors illustrated that Se_ind_ and Sp_ind_ are underestimated in the case of independent classification errors, whereas Se_pos_ and Sp_pos_ are overestimated in the scenarios with maximum possible positive correlation of classification errors.  3. With information of sensitivity and specificity under different scenarios, the readers can construct corrected ORs for the study. |
| **Explicit discussions regarding the similarities and differences with other methods** | Different from Wacholder et al 1993^69^ that assessed both binary and continuous exposure variable, this method is restricted to a binary exposure and a categorical outcome. It has an additional requirement that a validation study is carried out in which routinely employed measures of a binary exposure variable are validated by an alloyed gold standard in a group of study participants from a population. |

#### Weinkam 1999 sensitivity analysis when multilevel exposure and covariates are both misclassified

| **Name/description** | **Sensitivity analysis when multilevel exposure and covariates are both misclassified** |
| --- | --- |
| **DOI** | 10.1093/oxfordjournals.aje.a010094 |
| **Year** | Weinkam 1999^73^ |
| **Crossed-referenced in QBA textbook^3^** | Not referenced |
| **QBA classification** | Simple sensitivity analysis |
| **Applicable study design scenarios described^b^** | Any observational studies using RR as effect measure |
| **Effect measure of interest** | RR^b^ |
| **Result of interest** | Corrected estimates |
| **Data structure/format of the eligible exposure/intervention^b^** | Categorical |
| **Data structure/format of the confounders, if applicable^b^** | Categorical |
| **Data structure/format of eligible outcome^b^** | Categorical |
| **Source(s) of bias addressed** | Misclassification bias for both categorical exposure and confounder/effect modifiers (nondifferential) |
| **Information/parameters needed to conduct the QBA method** | - **M_CB_**: misclassification matrix for C with respect to B;  - **P_WB_**: the distribution of the population at risk with respect to W and B;  - Relative risks for X and C in all strata by the exposure and confounder/effect modifier level;  - Assumed rate of outcome among the unexposed.  - **M_XW_**: misclassification matrix for X with respect to W (Optional).  *Notes: C: confounder/ effect modifiers; D: outcome; X: exposure; W and B are surrogate variables used to estimate X and C, respectively.* |
| **Mathematical formula** | Step 1: Generate a four-dimensional array of the population classified by W, B, X, and C- the population distribution P_WB_, along with the misclassification matrices M_CB_, M_XW_, can be used to generate the array (see the example in the original paper).  Step 2: Compute the number of cases with respect to W and B: the assumed rate of outcome among the unexposed along with the assumed relative risks can then be used to compute the number of cases in each cell and the resulting four-dimensional array collapsed along the X and C axes to yield the expected number of cases with respect to W and B, D_WB_.  Step 3: Raw RRs are obtained by dividing the total number of cases in each row of D_WB_ by the total population at risk in the corresponding row of P_WB_. These are then normalized by dividing the values by the risk for the reference category (i.e., the lowest exposed level).”  Step 4: Compute the risk ratios for W standardized with respect to B: these are obtained by multiplying the matrix R_WB_ of estimated risks with respect to W and B by the vector S_B_ and normalizing as above. R_WB_ is obtained by dividing each element of D_WB_ by the corresponding element of P_WB_.  Step 5: If M_XW_ is also available, one can recover the true RRs by multiplying the standardized risk ratios on the left by M_XW_^T-1^ and then dividing by the relative risks for X and C. |
| **Data assumptions and additional required features** | 1. Misclassification of the exposure (i.e., X) by the surrogate (i.e., W) is independent of the other predictor variables and likewise for the misclassification of C by B.  2. The surrogate (i.e., W or B) should be highly correlated with the true variable, and its relation to the true variable should not depend on other variables included in the analysis.  3. If the surrogate variable is the measurement of another substance to which the subjects may be exposed, that substance should not be a risk factor in and of itself.  3. The authors stressed that "X and C are both risk factors for D but that their surrogates are not in themselves risk factors.”  4. The authors noted that “any misclassification matrix actually used in solving for the true risks is invertible.” |
| **Available and/or recommended software** | Not mentioned |
| **Output of the QBA analysis** | Corrected effect estimates |
| **Interpretation of the QBA analysis output** | Effect estimate accounting for misclassification of both exposure and confounder |
| **Considerations related to the interpretation of the results** | The authors noted that the fact that the dose response of the final result is not monotonic would suggest that this is not the correct misclassification matrix. Implausible or impossible true RRs can help rule out patterns of misclassification that are unlikely to be correct. |
| **Explicit discussions regarding the similarities and differences with other methods** | The authors mentioned that “Fung and Howe^74^ analyzed the effects of joint misclassification of exposure and confounding variables on the magnitude of the bias.” This paper extended previous results on nondifferential misclassification bias for both categorical exposure and confounder/effect modifiers. |

#### Chu et al 2006 Bayesian approach for misclassification

| **Name/description** | **Bayesian approach for misclassification** |
| --- | --- |
| **DOI** | 10.1016/j.annepidem.2006.04.001 |
| **Year** | Chu et al 2006^75^ |
| **Crossed-referenced in QBA textbook^3^** | Referenced and explained |
| **QBA classification** | Bayesian analysis |
| **Applicable study design scenarios described^b^** | Case-control studies |
| **Effect measure of interest** | OR |
| **Result of interest** | Corrected estimate |
| **Data structure/format of the eligible exposure/intervention^b^** | Categorical |
| **Data structure/format of the confounders, if applicable^b^** | Not mentioned |
| **Data structure/format of eligible outcome^b^** | Categorical |
| **Source(s) of bias addressed** | Exposure misclassification (differential or nondifferential) |
| **Information/parameters needed to conduct the QBA method** | - **a***: number of cases that are classified as exposed;  - **c***: number of controls that are classified as exposed;  - **N_1,_ N_0_**: the total numbers of cases and controls;  **- P_1,_ P_0_**: the observed proportions of cases and controls classified as exposed;  - **Se_1_, Se_0_**: probability of being classified as exposed when truly exposed among the cases and controls;  - **Sp_1_, Sp_0_**: probability of being classified as unexposed when truly unexposed among the cases and controls; |
| **Mathematical formula** | Step 1. Assuming a random sampling of cases and controls, the observed counts of misclassified exposed subjects would follow binomial distributions as a*~Bin (N_1_, P_1_) and c*~Bin (N_0_, P_0_);  Step 2. Obtain prior distributions of Se_1_, Se_0_, Sp_1_, Sp_0,_ π_0_ and π_1_ based on information from previous studies and expert opinions. The joint distribution of Se and Sp can be estimated by a bivariate meta-regression analysis (Appendix B of the original paper^75^);  3. Calculate the posterior distribution using:  $f\left( \alpha_{1},\alpha_{0},Se_{1},Sp_{1},Se_{0},Sp_{0}\mid\right.$ data $)$ $\propto\prod_{i=0}^{1} \left\{ \left[ 1-Sp_{i}+\pi_{i}\left( Se_{i}+Sp_{i}-1 \right) \right]^{A_{i}} \right.$ $\left. \left[ Sp_{i}-\pi_{i}\left( Se_{i}+Sp_{i}-1 \right) \right]^{N_{i}-A_{i}} \right\}\times Prior\left( Se_{1},Sp_{1},Se_{0},Sp_{0} \right)\times Prior\left( \pi_{1},\pi_{0} \right)$; where A_1_=a*, A_0_=c*.  4. Incorporate the uncertainty about misclassification parameters into the results by using methods such as Monte-Carlo sensitivity analysis and (usually with the code) compute the true (corrected) OR:  $\begin{matrix} OR_{c} & =f\left( Se_{0},Sp_{0},Se_{1},Sp_{1} \right) \\ & =\frac{\pi_{1}/\left( 1-\pi_{1} \right)}{\pi_{0}/\left( 1-\pi_{0} \right)} \\ & =\frac{\left( P_{1}+Sp_{1}-1 \right)\left( Se_{0}-P_{0} \right)}{\left( P_{0}+Sp_{0}-1 \right)\left( Se_{1}-P_{1} \right)} \end{matrix}$ |
| **Data assumptions and additional required features** | 1. Cases and controls are randomly sampled.  2. $Se_{1}+Sp_{1}\geq1$, $Se_{0}+Sp_{0}\geq1$*.* |
| **Available and/or recommended software** | WinBUGS (code available upon request to the authors of the original paper) |
| **Output of the QBA analysis** | Corrected effect estimate*  *Note: If the readers are interested in accompanying 95% intervals, formula for standard error of ln (OR_c_) is also provided in the paper, as equation 3, with additional assumptions)* |
| **Interpretation of the QBA analysis output** | Effect estimate accounting for misclassification |
| **Considerations related to the interpretation of the results** | Not mentioned |
| **Explicit discussions regarding the similarities and differences with other methods** | 1. The authors noted that the Gustafson et al 2001 method^76^ assumes that Se and Sp are uncorrelated. However, Chu et al 2006’s method is an extension to Gustafson et al’s method where they directly modeled uncertainty about Se and Sp and their correlation in the prior distributions.  2. The authors noted that “Fox et al 2005^77^ provided an alternative specification for correlated prior for Se between case and control and correlated prior for Sp between case and control, which include triangular and trapezoidal probability densities. However, they did not include a correlation between Se and Sp or a meta-regression to estimate the joint distribution of Se and Sp.”^75^ Moreover, Fox et al 2005^77^ is more computationally intensive than this method, which takes much less time to run a simulation.  3. In the QBA textbook,^3^ the authors compared the results among crude analysis, probabilistic analysis, and Bayesian analysis, and found that the point estimate is identical, and the 95% intervals are similar to each other but the 95% intervals are the largest in Bayesian analysis. However, the authors noted that the results from different bias analyses could differ too. |

#### Liu et al 2019 Bayesian analysis of a matched case-control study

| **Name/description** | **Bayesian analysis of a matched case-control study** |
| --- | --- |
| **DOI** | 10.1002/sim.3694 |
| **Year** | Liu et al 2019^78^ |
| **Crossed-referenced in QBA textbook^3^** | Not referenced |
| **QBA classification** | Bayesian analysis |
| **Applicable study design scenarios described^b^** | Matched case-control studies |
| **Effect measure of interest** | OR |
| **Result of interest** | Corrected effect estimates |
| **Data structure/format of the eligible exposure/intervention^b^** | Categorical |
| **Data structure/format of the confounders, if applicable^b^** | Not mentioned |
| **Data structure/format of eligible outcome^b^** | Categorical |
| **Source(s) of bias addressed** | Exposure misclassification (nondifferential)*  *Note: *The authors noted that with sufficient prior information or validation data, extensions to differential misclassification are straightforward.* |
| **Information/parameters needed to conduct the QBA method** | - $\boldsymbol{\theta}_{\boldsymbol{ij}}$: the marginal probability of true exposure status; It is defined as $\theta_{ij}=P\left( E_{1k}=i,E_{2k}=j \right)$, for $i,j=0,1$, where 0 indicates unexposed and 1 indicates exposed; moreover, (E_1k_ , E_2k_ ) representing the true exposure status for the case and control, respectively, in the kth of n matched pairs.  - **SN, SP**: the sensitivity (SN), and specificity (SP) of the exposure classification among cases (d=1) and controls (d=2); |
| **Mathematical formula** | Step 1: Set up a joint prior distribution for parameters $\left( \theta_{00},\theta_{10},\theta_{01},\theta_{11},SN,SP \right)$ as follows: $\begin{matrix} \left( \theta_{01}+\theta_{10},\theta_{11},\theta_{00} \right) & \sim Dirichlet\left( s,c_{11},c_{00} \right) \\ log\left( \frac{\theta_{10}}{\theta_{01}} \right)=log(OR) & \sim logit\left( Beta\left( c_{10},c_{01} \right) \right) \\ \mathrm{SN} & \sim Beta\left( \alpha_{0},\alpha_{1} \right) \\ \mathrm{SP} & \sim Beta\left( \alpha_{2},\alpha_{3} \right) \end{matrix}$  Step 2: Generate a posterior distribution using the joint model and prior specification to the parameters (typically with WinBUGS).  *Note: Determination of hyperparameters including* $\left( s,c_{11},c_{00} \right), Beta\left( c_{10},c_{01} \right)$*,*  $\left( \alpha_{0},\alpha_{1},\alpha_{2},\alpha_{3} \right)$ *is discussed in Section 2.3.^78^* |
| **Data assumptions and additional required features** | 1. Multiple exposures are evaluated individually in the study; moreover, all available covariates are used for matching and none entering the model.  2. In the prior distribution, $\left( \theta_{01}+\theta_{10},\theta_{11},\theta_{00} \right)$ and $\frac{\theta_{10}}{\theta_{01}}$ are set to be independent of one another in the prior, except that  $\theta_{11}\theta_{00}\geq\theta_{10}\theta_{01}$.  3. SN+SP>1, which usually indicates a good assessment scheme.  4. When the exposure assessment derives from a job-exposure matrix, the nondifferential misclassification assumption  is tantamount to assuming that occupation and disease status are conditionally independent given the exposure status. |
| **Available and/or recommended software** | WinBUGS code is available from the authors of the original paper upon request. |
| **Output of the QBA analysis** | Corrected effect estimates |
| **Interpretation of the QBA analysis output** | Effect estimates accounting for nondifferential exposure misclassification |
| **Considerations related to the interpretation of the results** | Not mentioned |
| **Explicit discussions regarding the similarities and differences with other methods** | 1. The authors noted that Prescott and Garthwaite 2005^79^’s random effect model is the starting point for this method. Moreover, Gustafson et al 2001^80^ reported a similar finding in the unmatched case-control setting.  2. The authors noted that their approach differs from others “in its focus on settings where validation data or multiple exposure assessments are not available”, and it utilizes expert prior knowledge about exposure–disease association and misclassification parameters. |

#### VanderWeele and Li 2019 method for differential measurement error

| **Name/description** | **Method for differential measurement error** |
| --- | --- |
| **DOI** | 10.1093/aje/kwz133 |
| **Year** | VanderWeele and Li 2019^81^ |
| **Crossed-referenced in QBA textbook^3^** | Referenced but not explained |
| **QBA classification** | Simple sensitivity analysis |
| **Applicable study design scenarios described^b^** | Any observational study designs |
| **Effect measure of interest** | RR, and OR |
| **Result of interest** | Explain-away |
| **Data structure/format of the eligible exposure/intervention^b^** | Categorical or continuous^b^ |
| **Data structure/format of the confounders, if applicable^b^** | Not mentioned |
| **Data structure/format of eligible outcome^b^** | Categorical or continuous^b^ |
| **Source(s) of bias addressed** | Exposure measurement error (in **Method 2, differential**) |
| **Information/parameters needed to conduct the QBA method** | - observed **RR** (or **OR**) (and 95% CIs)  **Method 2**:  - **{p1/(1 – p1)}1/ {p0/(1 – p0)}**: true risk ratio between A and Y;  - **{p1’/(1 – p1’)}/ {p0’/(1 – p0’)}**: observed odds ratio between A* and Y*;  - the odds ratio related to the sensitivity of the measurement A* for A conditional on the outcome being present versus absent: {s1’/(1 – s1’)}/{s0’/(1 – s0’)};  - the odds ratio of the false-positive probabilities for the exposure: {f1’/(1 – f1’)}/{f0’/(1 – f0’)};  - correct classification ratio r_c_ ={s1’/s0’}/{(1 – f1’)/(1 – f0’)};  - incorrect classification ratio r_i_ ={f1’/f0’}/{(1 – s1’)/(1 – s0’)}.  *Note: A: exposure; A*: measured exposure; C: observed confounders; f: false positive probability; s: sensitivity; Y: outcome; Y*: measured outcome.* |
| **Mathematical formula** | **Method 2**:  if p1/p0 ≥ 1, then {p1/(1 – p1)}/{p0/(1 – p0)} ≥ {p1’/(1 – p1’)}/{p0’/(1 – p0’)}/max[{s1’/(1 – s1’)}/{s0’/(1 – s0’)}, {f1’/(1 – f1’)}/{f0’/(1 – f0’)}, r_c_, r_i_]; and if p1/p0 ≤ 1, then {p1/(1 – p1)}/{p0/(1 – p0)} ≤ {p1’/(1 – p1’)}/{p0’/(1 – p0’)}/min[{s1’/(1 – s1’)}/{s0’/(1 – s0’)}, {f1’/(1 – f1’)}/{f0’/(1 – f0’)}, r_c_, r_i_]. |
| **Data assumptions and additional required features** | **For both Method 1 and Method 2**:  No unmeasured confounders  **For Method 2 only**:  the correct and incorrect classification ratios, r_c_ and r_i_, are not larger than the differential measurement error direct effects of Y on A* captured by the odds ratios {s1’/(1 – s1’)}/{s0’/(1 – s0’)}and {f1’/(1 −f1’)}/{f0’/(1−f0’)}. |
| **Available and/or recommended software** | Not mentioned |
| **Output of the QBA analysis** | Observed RR or OR divided by the maximum strength of the differential measurement error. The latter is further interpreted as follows:  **Method 2**: the maximum of the effect of Y on A* conditional on A (i.e., the maximum of the sensitivity odds ratio for Y on A*, {s1’/(1 – s1’)}/{s0’/(1 – s0’)}), and the false-positive probability odds ratio for the effect of Y on A*, {f1’/(1 −f1’)}/{f0’/(1−f0’)}. |
| **Interpretation of the QBA analysis output** | **Method 2**:  For the observed association to be completely explained away (i.e., reduced to the null) by differential measurement error so there is no true causative effect, the true effect on the odds ratio scale of the exposure on the outcome must be at least as large as the observed odds ratio between the mismeasured exposure and the outcome divided by the maximum odds ratio of the effect of the outcome on mismeasured exposure conditional on the true exposure. |
| **Considerations related to the interpretation of the results** | 1. The authors noted that “The results do not necessarily yield definitive conclusions but can be helpful in establishing a general sense as to the robustness of results.”  2. The authors noted that if data are available on the necessary sensitivities and specificities concerning the differentially mismeasured exposure or outcome, then such data could be used to obtain more precise inferences about the magnitude of the true effect.  3. The authors noted that for exposure misclassification, if the outcome is rare, then the formula could also be employed with risk ratios, because the odds ratios for the outcome will then approximate risk ratios. |
| **Explicit discussions regarding the similarities and differences with other methods** | The author mentioned that the E-value method by VanderWeele and Ding 2017^48^ is an analogous approach that addresses the minimum strength of unmeasured confounding necessary to explain away an observed exposure-outcome association. |

#### Baum et al 2021 method for exposure misclassification bias in vaccine effectiveness

| **Name/description** | **Method for exposure misclassification bias in vaccine effectiveness** |
| --- | --- |
| **DOI** | <https://doi.org/10.1371/journal.pone.0251622> |
| **Year** | Baum et al 2021^82^ |
| **Crossed-referenced in QBA textbook^3^** | Not referenced |
| **QBA classification** | Simple sensitivity analysis |
| **Applicable study design scenarios described^b^** | Any observational study on vaccine effectiveness |
| **Effect measure of interest** | Vaccine effectiveness (VE), which is the vaccine-induced relative reduction in the risk of disease |
| **Result of interest** | Corrected estimate |
| **Data structure/format of the eligible exposure/intervention^b^** | Categorical |
| **Data structure/format of the confounders, if applicable^b^** | Not mentioned |
| **Data structure/format of eligible outcome^b^** | Categorical |
| **Source(s) of bias addressed** | Exposure misclassification (nondifferential) |
| **Information/parameters needed to conduct the QBA method** | - **γ**: the true vaccination coverage, i.e., the proportion of truly vaccinated individuals.  - **P_1_**: the risk of the disease in those observed as vaccinated;  - **P_0_**: the risk of the disease in those observed as unvaccinated;  - $\boldsymbol{P}\mathbf{(}\tilde{\boldsymbol{V}}\mathbf{=1)}$: observed vaccination coverage;  - **SE**: the sensitivity, i.e., probability of measuring correctly the vaccination status of a vaccinated individual;  - **SP**: the specificity, i.e., probability of measuring correctly the vaccination status of an unvaccinated individual; |
| **Mathematical formula** | $\gamma=P(V=1)=\frac{P(\tilde{V}=1)+SP-1}{SE+SP-1}$;  $\begin{matrix} & \\ & VE=1-\frac{1-\gamma}{\gamma}\cdot\frac{p_{1}\cdot SP\cdot(SE\cdot\gamma+(1-SP)\cdot(1-\gamma))-p_{0}\cdot(1-SP)\cdot(1-SE\cdot\gamma-(1-SP)\cdot(1-\gamma))}{p_{0}\cdot SE\cdot(1-SE\cdot\gamma-(1-SP)\cdot(1-\gamma))-p_{1}\cdot(1-SE)\cdot(SE\cdot\gamma+(1-SP)\cdot(1-\gamma))}. \end{matrix}$ |
| **Data assumptions and additional required features** | 1. There are no other sources of bias.  2. The sensitivity and specificity of exposure measurement are known or are estimable from external sources |
| **Available and/or recommended software** | Not mentioned |
| **Output of the QBA analysis** | VE |
| **Interpretation of the QBA analysis output** | The vaccine-induced relative reduction in the risk of disease. |
| **Considerations related to the interpretation of the results** | The authors noted that if misclassification in the exposure status is not accurately considered, expressions quantifying the magnitude of bias will be wrong. |
| **Explicit discussions regarding the similarities and differences with other methods** | This model on exposure misclassification is a special case of the model by Tang et al 2015^83^ for misclassification in both exposure and outcome. |

### Misclassification bias (outcome misclassification bias)

#### Copeland et al 1977 method for misclassification bias in the estimation of relative risk

| **Name/description** | **Method for misclassification bias in the estimation of relative risk** |
| --- | --- |
| **DOI** | 10.1093/oxfordjournals.aje.a112408 |
| **Year** | Copeland et al 1977^60^ |
| **Crossed-referenced in QBA textbook^3^** | Referenced but not explained |
| **QBA classification** | Simple sensitivity analysis |
| **Applicable study design scenarios described^b^** | Cohort or case-control studies |
| **Effect measure of interest** | RR, and OR |
| **Result of interest** | Corrected estimate |
| **Data structure/format of the eligible exposure/intervention^b^** | Categorical |
| **Data structure/format of the confounders, if applicable^b^** | Not mentioned |
| **Data structure/format of eligible outcome^b^** | Categorical |
| **Source(s) of bias addressed** | Misclassification of study subjects (i.e., disease occurrence in cohorts or exposure history in case-control studies, differential or nondifferential) |
| **Information/parameters needed to conduct the QBA method** | **-** $\boldsymbol{a}^{\mathbf{'}}$: the observed number of study subjects that are exposed and have the outcome (i.e., the cells in a 2 by 2 table);  **-** $\boldsymbol{c}^{\mathbf{'}}$: the observed number of study subjects that are not exposed and have the outcome (i.e., the cells in a 2 by 2 table);  **-** $\boldsymbol{N}_{\mathbf{0}}$: total number of unexposed;  **-** $\boldsymbol{N}_{\mathbf{1}}$: total number of exposed;  **-** $\boldsymbol{Sp}$: specificity of classification procedure (i.e., disease occurrence in cohorts or exposure history in case-control studies). |
| **Mathematical formula** | $RR=\frac{\left[ a^{'}-N_{1}(1-Sp) \right]/N_{1}}{\left[ c^{'}-N_{0}(1-Sp) \right]/N_{0}}$ |
| **Data assumptions and additional required features** | 1. The misclassification affects only the comparison criterion—that is, disease occurrence in cohort or follow-up studies, and exposure history in case-control studies.  2. The misclassified variable is dichotomous.  3. There is no bias due to selection or confounding in the data from which the estimates are calculated, thereby allowing the use of simple, unadjusted ratio measures of effect. |
| **Available and/or recommended software** | Not mentioned |
| **Output of the QBA analysis** | Corrected estimate |
| **Interpretation of the QBA analysis output** | Corrected effect estimate that accounts for misclassification bias. |
| **Considerations related to the interpretation of the results** | Not mentioned |
| **Explicit discussions regarding the similarities and differences with other methods** | Not mentioned |

#### Barron 1977 method for misclassification for relative risks

| **Name/description** | Method for misclassification for relative risks |
| --- | --- |
| **DOI** | https://doi.org/10.2307/2529795 |
| **Year** | Barron 1977^61^ |
| **Crossed-referenced in QBA textbook^3^** | Referenced but not explained |
| **QBA classification** | Simple sensitivity analysis |
| **Applicable study design scenarios described^b^** | Cohort studies |
| **Effect measure of interest** | RRs |
| **Result of interest** | Corrected effect estimate |
| **Data structure/format of the eligible exposure/intervention^b^** | Categorical |
| **Data structure/format of the confounders, if applicable^b^** | Not applicable |
| **Data structure/format of eligible outcome^b^** | Categorical |
| **Source(s) of bias addressed** | Exposure and/or outcome misclassification (differential) |
| **Information/parameters needed to conduct the QBA method** | - **p_11_**: probability of an individual classified as having both A and B;  - **p_10_**: probability of an individual classified as having A but not B;  - **p_01_**: probability of an individual classified as having B but not A;  - **p_00_**: probability of an individual classified as having neither A nor B.  *Notes: A: exposure; B: outcome.* |
| **Mathematical formula** | the corrected RR: $R_{p}=\left[ \frac{p_{11}}{p_{10}} \right]\left[ \frac{p_{10}+p_{00}}{p_{11}+p_{01}} \right]$.  *Notes: If the probabilities of systematically misclassifying a given element of the sample with respect to A and B are known, formula with matrix involved can be used which were presented in the section 2 of the paper as formula (1).* |
| **Data assumptions and additional required features** | It is assumed that a study sample is drawn from the population and each element of the sample is classified according to the presence or absence of A and B. |
| **Available and/or recommended software** | Not mentioned |
| **Output of the QBA analysis** | R_p_ |
| **Interpretation of the QBA analysis output** | The RR corrected for misclassification |
| **Considerations related to the interpretation of the results** | Not mentioned |
| **Explicit discussions regarding the similarities and differences with other methods** | This method is flexible in the sense that it allows simultaneous adjustment of misclassification of exposure and outcome.^3^ |

#### Green 1983 method for outcome misclassification

| **Name/description** | **Method for outcome misclassification** |
| --- | --- |
| **DOI** | 10.1093/oxfordjournals.aje.a113521 |
| **Year** | Green 1983^84^ |
| **Crossed-referenced in QBA textbook^3^** | Not referenced |
| **QBA classification** | Simple sensitivity analysis |
| **Applicable study design scenarios described^b^** | Prospective epidemiologic studies or population surveys^b^ |
| **Effect measure of interest** | RR |
| **Result of interest** | Corrected estimate |
| **Data structure/format of the eligible exposure/intervention^b^** | Categorical |
| **Data structure/format of the confounders, if applicable^b^** | Not mentioned |
| **Data structure/format of eligible outcome^b^** | Categorical |
| **Source(s) of bias addressed** | Outcome misclassification (nondifferential) |
| **Information/parameters needed to conduct the QBA method** | - **RR’:** observed risk ratio;  - **Pred_2_:** predictive value of a positive outcome (i.e., the probability that a subject with a positive test result actually has the outcome) in the unexposed group only;  - **I_2_:** true disease frequency in the unexposed group only. |
| **Mathematical formula** | $RR=\frac{{[RR^{'}-(1-{Pred}_{2})}/{(1-I_{2})]}}{{[1-(1-{Pred}_{2})}/{(1-I_{2})]}}$;  When the I_2_ is considered low relative to Pred_2_, then we have  $RR=\frac{RR^{'}-1+{Pred}_{2}}{{Pred}_{2}}$.  *Note: RR: true relative risk.* |
| **Data assumptions and additional required features** | 1. For the sake of simplicity, any other possible biases such as those due to confounding or selection have been ignored.  2. Exposure status can be determined without error. |
| **Available and/or recommended software** | Not mentioned |
| **Output of the QBA analysis** | Corrected RR |
| **Interpretation of the QBA analysis output** | RR adjusted for nondifferential outcome misclassification |
| **Considerations related to the interpretation of the results** | 1. The authors noted that “The determination of specificity and sensitivity of the classification procedure depends on information on the performance of the test in those individuals classified both as cases and as noncases.” When the information is not obtained on noncases, this paper provides a method to adjust the bias by use of the predictive value of positive outcome and the disease frequency in the unexposed group.  2. The authors provided a method when disease frequency is low relative to the predictive value.  3. This method could be adapted to case-control studies with appropriate modifications. |
| **Explicit discussions regarding the similarities and differences with other methods** | Not mentioned |

#### Greenland and Kleinbaum 1983 generalizations of Barron’s matrix correction method

| **Name/description** | **Generalizations of Barron’s matrix correction method** |
| --- | --- |
| **DOI** | 10.1093/ije/12.1.93 |
| **Year** | Greenland and Kleinbaum 1983^63^ |
| **Crossed-referenced in QBA textbook^3^** | Referenced but not explained |
| **QBA classification** | Simple sensitivity analysis |
| **Applicable study design scenarios described^b^** | Case-control studies^b^ with 2 by 2 table (in **Method 1**) or matched-pair case control studies (in **Method 2**) |
| **Effect measure of interest** | OR |
| **Result of interest** | Corrected estimate |
| **Data structure/format of the eligible exposure/intervention^b^** | Categorical |
| **Data structure/format of the confounders, if applicable^b^** | Not mentioned |
| **Data structure/format of eligible outcome^b^** | Categorical^b^ |
| **Source(s) of bias addressed** | Exposure and/or outcome misclassification (differential or nondifferential) |
| **Information/parameters needed to conduct the QBA method** | **Method 1**:  - **m:** misclassified cells stratified by exposure and outcome status;  - **P_rxy_:** probability that study subject with true value x of X and y of Y is classified as having value r for X;  - **Q_syx_**: probability that study subject with true value y of Y and x of X is classified as having value s for Y.  *Notes: X: exposure; Y: outcome; Prxy and/or Qsyx will be used to construct correction matrix C.*  **Method 2**:  - **m**: misclassified cells stratified by exposure and outcome status;  - **P_rxy_**: probability that pair member 1 is classified as having response value r;  - **Q_syx_**: probability that pair member 2 is classified as having response value s.  *Notes: X: response value of the first pair-member; Y: response value of the second pair-member; Prxy and/or Qsyx will be used to construct correction matrix C.* |
| **Mathematical formula** | $m_{rs}=\sum_{x,y} P_{rxy}Q_{syx}t_{xy}$;  These equations can be compactly written in matrix form, m = Ct,  If C is invertible, a correction formula can be derived immediately from equation,  t= C^-1^ m.  *Notes: C: correction matrix; m: misclassified variables; t: true variables.* |
| **Data assumptions and additional required features** | Misclassification of each variable is independent of misclassification of the other variables (i.e., the misclassification probabilities for a variable do not depend on whether or not misclassification took place for another variable). |
| **Available and/or recommended software** | Not mentioned |
| **Output of the QBA analysis** | Corrected OR |
| **Interpretation of the QBA analysis output** | OR adjusted for misclassification (OR_m_) |
| **Considerations related to the interpretation of the results** | 1. The authors noted that estimate of C must be derived either from earlier studies involving similar classification methods, or from a validation study carried out on a subsample of the study subjects at hand.  2. The authors noted that C will vary from study to study, especially when considering questionnaire items administered to separate populations at separate times.  3. The authors noted that when classification of one of both variables takes place before sampling, C will depend heavily on the study design, and a fallacy in estimating C from data external to the study can arise.  4. The authors noted that complete validation is rarely possible, and corrected estimates should not be viewed as being fully corrected.  *Note: the study also provided a valid correction of C if the ratio R of the case-control sampling fractions is known.* |
| **Explicit discussions regarding the similarities and differences with other methods** | Previous methods on misclassification including Barron 1977^61^ and Copeland et al 1977^60^, had different assumptions that misclassification of each variable was nondifferential; and misclassification of each variable was independent of misclassification of the other variables. None of the methods dealt with situation of pair-matched variable. |

#### Farrington 1990 method for misclassification bias in cohort studies of vaccine efficacy

| **Name/description** | **Method for misclassification bias in cohort studies of vaccine efficacy** |
| --- | --- |
| **DOI** | https://doi.org/10.1002/sim.4780091110 |
| **Year** | Farrington 1990^85^ |
| **Crossed-referenced in QBA textbook^3^** | Not referenced |
| **QBA classification** | Multidimensional analysis |
| **Applicable study design scenarios described^b^** | Prospective cohort studies and non-randomized interventional studies of vaccine efficacy |
| **Effect measure of interest** | RR |
| **Result of interest** | Corrected estimate |
| **Data structure/format of the eligible exposure/intervention^b^** | Categorical^b^ |
| **Data structure/format of the confounders, if applicable^b^** | Not mentioned |
| **Data structure/format of eligible outcome^b^** | Categorical^b^ |
| **Source(s) of bias addressed** | Disease misclassification (differential) |
| **Information/parameters needed to conduct the QBA method** | - **a**: the sensitivities of the ascertainment procedure in the vaccinated group compared to that in the control group;  - ***l***: proportions of cases ascertained during follow-up which are reported in the vaccinated group compared to that in the control group;  - **PV**: the approximate predictive value of the ascertainment procedure in the control group (proportion of ascertained cases which are true cases);  - **r**: the proportions of individuals at the start of the study in the vaccinated group compared to that in the control group who are susceptible;  - **R**: true value of relative risk;  - **R***: observed value of relative risk. |
| **Mathematical formula** | R = (l/ar) {1 – (1 – R*/*l*)/PV}.  *Note: Advanced formula for age-specific vaccine efficacy can be found in the paper.* |
| **Data assumptions and additional required features** | 1. The model assumes that follow-up of each individual terminates upon ascertainment of disease.  2. The specificity of the case ascertainment procedure is independent of vaccination status. |
| **Available and/or recommended software** | Not mentioned |
| **Output of the QBA analysis** | Corrected effect estimate |
| **Interpretation of the QBA analysis output** | Effect estimate accounting for disease misclassification |
| **Considerations related to the interpretation of the results** | The authors noted that “The model does not attempt to deal with other types of misclassifications, such as misclassification of vaccination status, which arise more commonly in case-control studies. Nor does the model correct for errors other than misclassification, such as susceptibility bias due to causes other than prior exposure.” |
| **Explicit discussions regarding the similarities and differences with other methods** | Not mentioned |

#### Magder 2003 response probability ratio for missing outcome

| **Name/description** | **Response probability ratio for missing outcome** |
| --- | --- |
| **DOI** | 10.1016/S0197-2456(03)00021-7 |
| **Year** | Magder 2003^86^ |
| **Crossed-referenced in QBA textbook^3^** | Not referenced |
| **QBA classification** | Multidimensional analysis |
| **Applicable study design scenarios described^b^** | Non-randomized interventional or observational study designs (examples are clinical trials) |
| **Effect measure of interest** | RR, OR, and RD |
| **Result of interest** | Corrected estimates |
| **Data structure/format of the eligible exposure/intervention^b^** | Categorical^b^ |
| **Data structure/format of the confounders, if applicable^b^** | Not mentioned |
| **Data structure/format of eligible outcome^b^** | Categorical |
| **Source(s) of bias addressed** | Missing data (outcome information) |
| **Information/parameters needed to conduct the QBA method** | - **a, b, c, d**: the observed number of study subjects stratified by exposure and outcome status (i.e., the cells in a 2 by 2 table);  - **RPR_A_**: the response probability ratio for treatment A; the result of response probabilities for those with treatment A who had treatment successes divided by response probabilities for those with treatment A who had treatment failures;  - **RPR_B_**: the response probability ratio for treatment B; the result of response probabilities for those with treatment B who had treatment successes divided by response probabilities for those with treatment B who had treatment failures;  *Notes: response probability is defined as the probability that a patient will not have a missing value for the outcome variable* |
| **Mathematical formula** | $\begin{matrix} \mathrm{RR}_{adj} & =\frac{a/\left( a+\mathrm{RPR}_{A}b \right)}{c/\left( c+\mathrm{RPR}_{B}d \right)}; \\ \mathrm{OR}_{adj} & =\frac{ad}{bc}\frac{\mathrm{RPR}_{B}}{\mathrm{RPR}_{A}}; \\ \mathrm{RD}_{adj} & =\frac{a}{a+\mathrm{RPR}_{A}b}-\frac{c}{c+\mathrm{RPR}_{B}d}. \end{matrix}$ |
| **Data assumptions and additional required features** | Study subjects should be randomly allocated to one of two groups (treatment A and treatment B), and there are three possible outcomes for each study subject (success, failure, and missing). |
| **Available and/or recommended software** | Not mentioned |
| **Output of the QBA analysis** | Estimates accounting for specified values of the RPRs |
| **Interpretation of the QBA analysis output** | RPR quantifies the departure from the missing at random assumption. |
| **Considerations related to the interpretation of the results** | 1. The authors noted that “If there is no association between outcome and response probabilities, the complete-case estimates are asymptotically unbiased.” (i.e., if RPR_A_=RPR_B_ = 1, then it is an example of data that are “missing at random.”)  2. The authors noted that “The complete-case OR estimates are asymptotically unbiased under certain plausible conditions, even if the response probabilities are associated with both treatment and outcome.” In many cases it might be reasonable to make this assumption. For example, if it is thought that the probability that a study participant has a missing outcome is the product of the probabilities of being missing due to background factors, treatment factors, and outcome-related factors, then the two RPRs would be equivalent.  3. The authors noted that reasonable parameter values (e.g., RPRs) should be chosen in the range supported by the data or set by subject-matter experts and should also be set prior to the unblinding the data.  4. The authors noted that other approaches (e.g., Bayesian approaches) can be combined to obtain the single best estimate of the effect of treatment. |
| **Explicit discussions regarding the similarities and differences with other methods** | Although this method was described with reference to clinical trial data, the method can be used for observational data when there is complete information about a predictor, but some missing information about outcomes. |

#### Lash et al 2010 probabilistic bias analysis for two biases

| **Name/description** | **Probabilistic bias analysis for two biases** |
| --- | --- |
| **DOI** | 10.1002/pds.1938 |
| **Year** | Lash et al 2010^18^ |
| **Crossed-referenced in QBA textbook^3^** | Referenced and explained |
| **QBA classification** | probabilistic analysis |
| **Applicable study design scenarios described^b^** | Any observational study designs with multiple covariates (examples are case-control studies) |
| **Effect measure of interest** | RR, OR, HR, and rate ratio |
| **Result of interest** | Corrected estimate |
| **Data structure/format of the eligible exposure/intervention^b^** | Categorical |
| **Data structure/format of the confounders, if applicable^b^** | Categorical |
| **Data structure/format of eligible outcome^b^** | Categorical, or time-to-event data |
| **Source(s) of bias addressed** | Outcome misclassification (differential or nondifferential) (in **Method 1**), or unmeasured confounding (single) (in **Method 2**) |
| **Information/parameters needed to conduct the QBA method** | -$\hat{\mathbf{RR}}$or $\hat{\mathbf{OR}}$**:** observed RR or OR  **Method 1**:  - the number of observed cases;  - $\boldsymbol{S}_{\boldsymbol{i=1, j}} \mathrm{and} \boldsymbol{S}_{\boldsymbol{i=0, j}}$: distribution of sensitivities *S* of the exposed and unexposed respectively (the paper used beta distribution as the example);  **Method 2**:  - **OR_DZ_**: distribution of the association between the; confounder and the outcome  - ***p*_Z, X=1_** or ***p*_Z, X=0_**: distribution of prevalence of the confounder in the exposed and the unexposed respectively.  *Note: D: disease; X: exposure; Z: confounder.* |
| **Mathematical formula** | **Method 1:** For disease misclassification:  $RR=\frac{\hat{RR}}{\frac{S_{i=1, j}}{S_{i=0, j}}}$;  **Method 2:** For unmeasured confounding:  $OR=\frac{\hat{OR}}{{RR}_{C}} ;{RR}_{C}=\frac{\mathrm{OR}_{DZ}\mathrm{OR}_{XZ}p_{Z,X=0}+1-p_{Z,X=0}}{\left( \mathrm{OR}_{DZ}+p_{Z,X=0}1-p_{Z,X=0} \right)\left( \mathrm{OR}_{XZ}p_{Z,X=0}+1-p_{Z,X=0} \right)}$;  $\mathrm{OR}_{XZ}$= [$p_{Z,X=1}$ (1- $p_{Z,X=0}$)]/[ $p_{Z,X=0}$ (1- $p_{Z,X=1}$)].  *Note: i: exposure status; j: stratum; OR: true odds ratio; RR: true relative risk;* ${RR}_{C}$*: relative risk due to confounding; S: sensitivity. Formula for 95% CIs were presented in the paper (equation (6)).* |
| **Data assumptions and additional required features** | 1. The bias is independent of confounding by measured variables, or requires that the dependence be incorporated into the bias model.  2. For disease misclassification specifically: perfect specificity of disease classification (i.e., presumably much greater than 95%). |
| **Available and/or recommended software** | SAS code available in the supplements |
| **Output of the QBA analysis** | Correct OR (95% CIs); corrected RR (95% CIs) |
| **Interpretation of the QBA analysis output** | OR adjusted for misclassification of the outcome, and/or unmeasured confounding |
| **Considerations related to the interpretation of the results** | 1.The authors mentioned that they proposed a solution to the assumption of the independence of the bias source from adjustments for measured covariates. The solution integrates the bias analysis results with a prior which is hard to parameterize. Therefore, it is critical that the analyst clearly explains the justification for the assigned values.  2. The two sensitivity parameters S can be drawn from their own beta distributions. |
| **Explicit discussions regarding the similarities and differences with other methods** | Earlier work focused on crude estimates,^1,7^ however application of these methods to estimates adjusted for multiple confounders is more difficult. Three solutions have been previously proposed,^19^ including using stratification, record level data and empirical or Bayesian methods, but they have their own limitations as well. Unlike previous, this method does not resort to stratification, record-level corrections, empirical, or Bayesian methods. |

#### Jurek et al 2013 methods to adjust for outcome misclassification accounting for case-control sampling with external information

| **Name/description** | **Methods to adjust for outcome misclassification accounting for case-control sampling with external information** |
| --- | --- |
| **DOI** | <http://dx.doi.org/10.1016/j.annepidem.2012.12.007> |
| **Year** | Jurek et al 2013^87^ |
| **Crossed-referenced in QBA textbook^3^** | Referenced and explained |
| **QBA classification** | Multidimensional analysis |
| **Applicable study design scenarios described^b^** | Case-control studies |
| **Effect measure of interest** | OR |
| **Result of interest** | Corrected estimate |
| **Data structure/format of the eligible exposure/intervention^b^** | Categorical^b^ |
| **Data structure/format of the confounders, if applicable^b^** | Categorical^b^ |
| **Data structure/format of eligible outcome^b^** | Categorical |
| **Source(s) of bias addressed** | Outcome misclassification (nondifferential) |
| **Information/parameters needed to conduct the QBA method** | - **f_C_**: sampling fraction for apparent cases;  - **f_N_**_:_ sampling fraction for apparent controls;  - **Se_p_**: probability of outcome being correctly classified in the source population;  - **Sp_p_**_:_ probability of non-outcome being correctly classified in source population.  *Note: 1. Se_p_, Sp_p_ requires prior information from validation study; 2. If Se_p_, Sp_p_ are Not mentioned but predictive values are, then the paper also provided formula using predictive values.* |
| **Mathematical formula** | $A^{*}=\frac{a^{*}}{f_{C}}$ and $B^{*}=\frac{b^{*}}{f_{N}}$;  $A=\frac{A^{*}-Fp_{p}\cdot N}{d};$  $B=\frac{B^{*}-Fn_{\text{P }}\cdot N}{d}$;  $\left[ \begin{matrix} A \\ B \end{matrix} \right]=\left[ \begin{matrix} Se_{p} & Fp_{p} \\ Fn_{p} & S_{p} \end{matrix} \right]^{-1}\cdot\left[ \begin{matrix} A^{*} \\ B^{*} \end{matrix} \right]$;OR${}_{\text{adjusted }}=\frac{A_{1/B_{1}}}{A_{0/B_{0}}}$.  *Note: A: Expected number in source population with actual outcome (i.e., true cases in the source population); A*: Expected number classified as cases in source population; B: Expected number in source population without the outcome (i.e., true noncases in the source population); B*: Expected number classified as noncases in source population; a*: Case counts in study under outcome misclassification (i.e., observed cases in the sample); b*: Control counts in study under outcome misclassification (i.e., observed controls in the sample);; Fn_p_: false-negative probabilities in the source population (Fn_P_ =1-Se_P_); Fp_p_: false-positive probabilities in the source population (Fp_P_ =1-Sp_P_); N: total population; subscript 1: among the exposed population; subscript 0: among the unexposed population.* |
| **Data assumptions and additional required features** | 1. It is assumed that the cases and controls in the sample have been subject to identical outcome diagnosis and classification criteria as the source population from which the classification probabilities (Se_p_, Sp_p_), predictive values (PVP, PVN), and the outcome risk r* were derived.  2. It is required that prior information of sensitivity and specificity or predictive values from validation study |
| **Available and/or recommended software** | Excel program available on request from the first author |
| **Output of the QBA analysis** | Corrected odds ratio and 95% CIs |
| **Interpretation of the QBA analysis output** | Odds ratios adjusted for outcome misclassification (OR_adjusted_) with and without accounting for case-control sampling. |
| **Considerations related to the interpretation of the results** | 1. Adjustment can only be conducted for variables with sampling fractions available.  2. Researchers had to rely on validation data for prior information but the quality of prior information varies.  3. The authors assumed the sampling fractions for cases and controls did not vary with exposure (no exposure-related selection bias), especially if sampling was done with no knowledge of exposure status.  4. If the original dataset is not available, matching is not possible to consider. |
| **Explicit discussions regarding the similarities and differences with other methods** | The authors noted that Greenland and Kleinbaum 1983^63^ proposed bias outcome adjustment formulas that assume either complete enrollment of subjects into the study or that the sampling fractions are the same for the numerators (e.g., cases) and denominators (e.g., noncases) of the outcome-frequency measures used in the study. Moreover, since case-control studies sample based on outcome status, formulas from Greenland and Kleinbaum 1983^63^ may not suitable for adjusting outcome misclassification in a case-control study. |

#### VanderWeele and Li 2019 method for differential measurement error

| **Name/description** | **Method for differential measurement error** |
| --- | --- |
| **DOI** | 10.1093/aje/kwz133 |
| **Year** | VanderWeele and Li 2019^81^ |
| **Crossed-referenced in QBA textbook^3^** | Referenced but not explained |
| **QBA classification** | Simple sensitivity analysis |
| **Applicable study design scenarios described^b^** | Any observational study designs |
| **Effect measure of interest** | RR, and OR |
| **Result of interest** | Explain-away |
| **Data structure/format of the eligible exposure/intervention^b^** | Categorical or continuous^b^ |
| **Data structure/format of the confounders, if applicable^b^** | Not mentioned |
| **Data structure/format of eligible outcome^b^** | Categorical or continuous^b^ |
| **Source(s) of bias addressed** | Outcome measurement error (in **Method 1, differential**) |
| **Information/parameters needed to conduct the QBA method** | - observed **RR** (or **OR**) (and 95% CIs)  **Method 1**:  - **p1/p0**: true risk ratio between A and Y;  - **p1*/p0***: observed risk ratio between A* and Y*;  - **s1/s0**: the ratio of the sensitivity parameter conditional on the exposure being present versus absent: s1/s0;  - **f1/f0**: the ratio of the false-positive probabilities conditional on the exposure being present versus absent: f1/f0.  *Note: A: exposure; A*: measured exposure; C: observed confounders; f: false positive probability; s: sensitivity; Y: outcome; Y*: measured outcome.* |
| **Mathematical formula** | **Method 1**:  if p1/p0 ≥ 1, then p1/p0 ≥(p1*/p0*)/max(s1/s0 , f1/f0); and if p1/p0 ≤ 1, then p1/p0 ≤ (p1*/p0*)/min(s1/s0 , f1/f0). |
| **Data assumptions and additional required features** | **For both Method 1 and Method 2**:  No unmeasured confounders |
| **Available and/or recommended software** | Not mentioned |
| **Output of the QBA analysis** | Observed RR or OR divided by the maximum strength of the differential measurement error. The latter is further interpreted as follows:  **Method 1**: “maximum direct effect” of A on Y* not through Y”: the maximum between sensitivity risk ratio for A on Y*, s1/s0 or false-positive probability ratio for A on Y*, f1/f0; |
| **Interpretation of the QBA analysis output** | **Method 1**:  For the observed association to be completely explained away (i.e., reduced to the null) by differential measurement error so there is no true causative effect, the true effect of the exposure on the outcome on the risk ratio scale must be at least as large as the observed association between the exposure and the mismeasured outcome divided by the maximum strength of differential measurement error. |
| **Considerations related to the interpretation of the results** | 1. The authors noted that “The results do not necessarily yield definitive conclusions but can be helpful in establishing a general sense as to the robustness of results.”  2. The authors noted that if data are available on the necessary sensitivities and specificities concerning the differentially mismeasured exposure or outcome, then such data could be used to obtain more precise inferences about the magnitude of the true effect.  3. The authors noted that for exposure misclassification, if the outcome is rare, then the formula could also be employed with risk ratios, because the odds ratios for the outcome will then approximate risk ratios. |
| **Explicit discussions regarding the similarities and differences with other methods** | The author mentioned that the E-value method by VanderWeele and Ding 2017^48^ is an analogous approach that addresses the minimum strength of unmeasured confounding necessary to explain away an observed exposure-outcome association. |

#### Endo et al 2020 bias correction methods for misclassification in test-negative designs

| **Name/description** | **Bias correction methods for misclassification in test-negative designs** |
| --- | --- |
| **DOI** | 10.1017/S0950268820002058 |
| **Year** | Endo et al 2020^88^ |
| **Crossed-referenced in QBA textbook^3^** | Not referenced |
| **QBA classification** | Simple sensitivity analysis |
| **Applicable study design scenarios described^b^** | Vaccine effectiveness  (VE) studies with test-negative designs (which can be regarded as a modified version of a case-control study) |
| **Effect measure of interest** | OR |
| **Result of interest** | Corrected estimates |
| **Data structure/format of the eligible exposure/intervention^b^** | Categorical |
| **Data structure/format of the confounders, if applicable^b^** | Categorical or continuous |
| **Data structure/format of eligible outcome^b^** | Categorical |
| **Source(s) of bias addressed** | Outcome misclassification (nondifferential)*  *Notes: *The paper proposed bias correction in both univariate and multivariate analysis, here we presented the method for univariate analysis.* |
| **Information/parameters needed to conduct the QBA method** | - $\boldsymbol{\alpha}$**,** $\boldsymbol{\beta}$: the sensitivity and specificity of the test, respectively;  - $\boldsymbol{X}_{\boldsymbol{U}}$***,*** $\boldsymbol{X}_{\boldsymbol{V}}$: number of cases among unvaccinated and vaccinated participants, respectively;  - $\boldsymbol{Y}_{\boldsymbol{U}}$***,*** $\boldsymbol{Y}_{\boldsymbol{V}}$: number of controls among unvaccinated and vaccinated participants, respectively. |
| **Mathematical formula** | Corrected OR $\gamma^{*}$ is calculated as follows*:  $\gamma^{*}=\frac{X_{V}-((1-\beta)/\beta)Y_{V}}{Y_{V}-((1-\alpha)/\alpha)X_{V}}\cdot\frac{Y_{U}-((1-\alpha)/\alpha)X_{U}}{X_{U}-((1-\beta)/\beta)Y_{U}}$；  The confidence interval is calculated using formula: $\gamma^{*}\exp\left( \pm1.96\sigma^{*} \right),$ where  $\begin{matrix} \sigma^{*} & =SD\left( log\left( \gamma^{*} \right) \right) \\ & =c\sqrt{\frac{\left[ \alpha Y_{V}-(1-\alpha)X_{V} \right]^{2}\left[ \beta X_{V}-(1-\beta)Y_{V} \right]^{2}}{\frac{X_{U}Y_{U}\left( X_{U}+Y_{U} \right)}{\left[ \alpha Y_{U}-(1-\alpha)X_{U} \right]^{2}\left[ \beta X_{U}-(1-\beta)Y_{U} \right]^{2}}}}. \end{matrix}$  $c=\alpha+\beta-1.$  *Notes: *The paper proposed bias correction in both univariate and multivariate analysis, here we presented the method for univariate analysis.* |
| **Data assumptions and additional required features** | 1. The occurrence of target diseases and non-target diseases are mutually independent.  2. Vaccines have no effect on the risk of non-target diseases.  3. The probability of medical attendance in vaccinated and unvaccinated population given infection is constant regardless of the disease (i.e., target disease or the non-target disease).  4. To acquire the confidence interval, log-normality of the γ is assumed.  5. When confounding is present, no interaction between covariates is assumed.  6. Other biases in the studies are non-existent or properly addressed. |
| **Available and/or recommended software** | R code is available on GitHub (https://github.com/akira-endo/TND-biascorrection/) |
| **Output of the QBA analysis** | $\gamma^{*}$and its 95% CIs |
| **Interpretation of the QBA analysis output** | An asymptotically unbiased estimate of the relative risk of the disease in the vaccinated population relative to the unvaccinated, after adjusting for misclassification. |
| **Considerations related to the interpretation of the results** | 1. In every simulation setting, the authors noted that their bias correction method was able to yield unbiased estimates whose median almost corresponds to the true VE.  2. The authors noted that the bias correction methods could be extended to other types of studies with misclassification bias.  3. The method depends on the assumed test sensitivity and specificity, misspecifying those values can lead to improper calculations. |
| **Explicit discussions regarding the similarities and differences with other methods** | Not mentioned |

### Misclassification bias (confounder misclassification bias)

#### Savitz and Baron 1989 method for nondifferential confounder misclassification

| **Name/description** | **Method for nondifferential confounder misclassification** |
| --- | --- |
| **DOI** | 10.1093/oxfordjournals.aje.a115210 |
| **Year** | Savitz and Baron 1989^89^ |
| **Crossed-referenced in QBA textbook^3^** | Not referenced |
| **QBA classification** | Simple sensitivity analysis |
| **Applicable study design scenarios described^b^** | Case control studies^b^ |
| **Effect measure of interest** | OR |
| **Result of interest** | Corrected estimate |
| **Data structure/format of the eligible exposure/intervention^b^** | Categorical^b^ |
| **Data structure/format of the confounders, if applicable^b^** | Categorical^b^ |
| **Data structure/format of eligible outcome^b^** | Categorical^b^ |
| **Source(s) of bias addressed** | Confounder misclassification (nondifferential) |
| **Information/parameters needed to conduct the QBA method** | - **OR_c_(e)**: the crude (unadjusted) effect estimate;  - **OR_pa_(e)**: the effect estimate with adjustment for the misclassified confounder (partially adjusted);  - **OR_ca_(e)**: the completely adjusted effect estimate, which equals to the stratum-specific effect estimate. |
| **Mathematical formula** | $\begin{matrix} & \text{ Per Cent } \\ & \text{ Adjustment } \end{matrix}=\frac{\mathrm{OR}_{c}(e)-\mathrm{OR}_{\mathrm{pa}}(e)}{\mathrm{OR}_{c}(e)-\mathrm{OR}_{\mathrm{ca}}(e)}\times100$  *Note: OR_ca_(e): the completely adjusted effect estimate.* |
| **Data assumptions and additional required features** | The confounder and exposure are assumed to have multiplicative effects on disease risk in generating expected cell frequencies. |
| **Available and/or recommended software** | Not mentioned |
| **Output of the QBA analysis** | Per Cent Adjustment |
| **Interpretation of the QBA analysis output** | Per Cent Adjustment: amount of confounding bias removed using the misclassified measure relative to the total confounding bias present. Per Cent Adjustment is a function of sensitivity and specificity and can also be derived from sensitivity and specificity (see Figure 1 in the paper). Moreover, (1 – Per Cent Adjustment) indicates the proportion of residual confounding. |
| **Considerations related to the interpretation of the results** | 1. The authors noted that when confounder and exposure do not have multiplicative effects on disease risk, the results can change considerably.  2. No other sources of error or misclassification are considered in the model.  3. The authors noted that investigators can rarely quantify derivation of accurate estimates for confounder misclassification, but may have some intuitive knowledge regarding the quality of their confounder measure. |
| **Explicit discussions regarding the similarities and differences with other methods** | Checkoway et al 1991^66^ presented the Per Cent Adjustment method as an example method for confounder misclassification. |

#### Nab et al 2020 bias analysis for a misclassified confounder

| **Name/description** | **Bias analysis for a misclassified confounder** |
| --- | --- |
| **DOI** | 10.1097/EDE.0000000000001239 |
| **Year** | Nab et al 2020^90^ |
| **Crossed-referenced in QBA textbook^3^** | Not referenced |
| **QBA classification** | Probabilistic analysis |
| **Applicable study design scenarios described^b^** | Point treatment observational study with a continuous outcome analyzed using a marginal structural model estimated using inverse probability weighting (MSM-IPW) or a conditional regression model |
| **Effect measure of interest** | Not mentioned |
| **Result of interest** | Bias illustration |
| **Data structure/format of the eligible exposure/intervention^b^** | Categorical |
| **Data structure/format of the confounders, if applicable^b^** | Categorical |
| **Data structure/format of eligible outcome^b^** | Continuous |
| **Source(s) of bias addressed** | Confounder misclassification (nondifferential) |
| **Information/parameters needed to conduct the QBA method** | - **p_1_**: sensitivity of L*;  - (**1-p_0_**): specificity of L*;  - **π_1_***: probability of receiving treatment if L* is present;  - **π_0_***: probability of receiving treatment if L* is not present;  - ***l***: prevalence of L*­;  - **γ***: association between L* and Y, given that A=0.  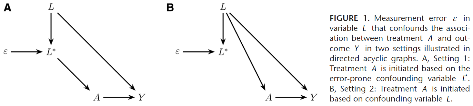  Figure 1 in the paper that illustrates the two settings.  A, setting 1: Treatment A is initiated based on the error-prone confounding variable L*. B, setting 2: Treatment A is initiated based on confounding variable L.  *Notes: ε: measurement error; A: treatment; L: confounder; L*: measured/observed confounder; Y: outcome.* |
| **Mathematical formula** | The authors recommend using the online tool to obtain the results (https://lindanab.shinyapps.io/SensitivityAnalysis/). |
| **Data assumptions and additional required features** | 1. **Consistency assumption**: we would observe Y = Y^a=0^ if the individual is not exposed or Y = Y^a=1^ if the individual is exposed.  2. **Stable-Unit-Treatment-Value-assumption**: the potential outcome Y^a^ for an individual does not depend on treatments received by other individuals and that there are not multiple versions of treatment.  3. **Conditional exchangeability**: given any level of L, if the untreated group had in fact received treatment, then their expected outcome would have been the same as that in the treated, and vice versa.  4. **Positivity**: we assume π_L_ (i.e., prevalence of confounder L)>0 for L =0, 1. |
| **Available and/or recommended software** | Online tool (https://lindanab.shinyapps.io/SensitivityAnalysis/) |
| **Output of the QBA analysis** | Bias plots |
| **Interpretation of the QBA analysis output** | Illustration of the potential range of misclassification bias. |
| **Considerations related to the interpretation of the results** | If the assumption of no treatment effect modification can be relaxed, then future research could extend the results to more general settings. |
| **Explicit discussions regarding the similarities and differences with other methods** | Not mentioned |

### Selection bias

#### Greenland 2003 quantifying biases for classical confounding and (or) collider-stratification bias

| **Name/description** | **Quantifying biases for classical confounding and (or) collider-stratification bias** |
| --- | --- |
| **DOI** | 10.1097/01.EDE.0000042804.12056.6C |
| **Year** | Greenland 2003^12^ |
| **Crossed-referenced in QBA textbook^3^** | Not referenced |
| **QBA classification** | Simple sensitivity analysis |
| **Applicable study design scenarios described^b^** | Any observational studies |
| **Effect measure of interest** | OR^b^ |
| **Result of interest** | Corrected estimate |
| **Data structure/format of the eligible exposure/intervention^b^** | Categorical |
| **Data structure/format of the confounders, if applicable^b^** | Categorical |
| **Data structure/format of eligible outcome^b^** | Categorical |
| **Source(s) of bias addressed** | Unmeasured confounding (single), selection bias |
| **Information/parameters needed to conduct the QBA method** | 1. **Classical confounding and Berksonian bias**:  -**crude OR** between E and D;  -**p**: p=P(C=1 \| D=E=0), the prevalence of C among the unexposed controls;  -**R_CD_, R_CE_**: the OR for C-D given E, for C-E given D (each assumed constant across strata of the given variable);  2. **M bias**:  -**R_AE_=R_AC_=R_BC_=R_BD_**: odds ratio of the association between A-E or A-C or B-C or B-D.  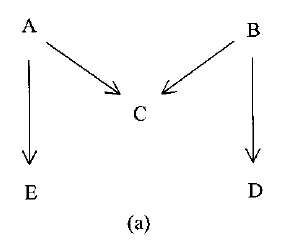  *Notes: A, B: variables on the E-D pathways as indicated in the picture; C: confounder/ collider; D: outcome; E: exposure.* |
| **Mathematical formula** | 1. **Classical confounding and Berksonian bias (selection bias)**:  $R_{ED}=crude OR/Bias\left( R_{CD},R_{CE},p \right)$,  where$\begin{matrix} & Bias\left( R_{CD},R_{CE},p \right)\equiv\left( R_{CD}R_{CE}p+1-p \right)/ \\ & \left( \left[ R_{CD}p+1-p \right]\left[ R_{CE}p+1-p \right] \right). \end{matrix}$  Let $G=\left( R_{CD}R_{CE} \right)^{1/2}$ be the geometric mean of $R_{CD}$ and $R_{CE}$The authors noted that when p= 1/ (1+G), the $Bias\left( R_{CD},R_{CE},p \right)$ reaches its maximum value.  $\mathrm{Bias}_{\max}\left( R_{CD},R_{CE} \right)\equiv(G+1)^{2}/\left( R_{CD}+2G+R_{CE} \right)$  In addition, if $R_{CD}=R_{CE}$, $\mathrm{Bias} \left( R_{CD},R_{CE},p \right)$ is even larger.  2. **M bias**:  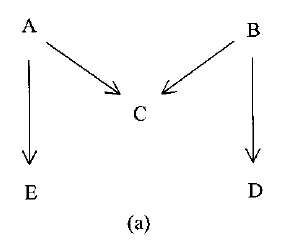  The lower bound of $R_{ED} is 16R(R+1)^{2}/\left( R^{2}+6R+1 \right)^{2}$, which is very close to 1, where $R_{AE}=R_{AC}=R_{BC}=R_{BD}=R$. |
| **Data assumptions and additional required features** | The author noted that all the biases discussed in the paper correspond to large-sample (asymptotic) biases.  1. **Classical confounding and Berksonian bias (selection bias)**:  R_CD_, R_CE_ and R_ED_ are assumed to be constant across strata of the given variable which is E, D, C respectively.  Moreover, the author also noted that “To apply formula 1 to measure confounding by C, one must assume D=1 is rare in all C-E categories, or that the parameters refer to a case-cohort study in which the time of D is ignored.”  2. **M bias**:  $R_{AE}=R_{AC}=R_{BC}=R_{BD}=R$.  *Notes: C: confounder/ collider; D: outcome; E: exposure.* |
| **Available and/or recommended software** | Not mentioned |
| **Output of the QBA analysis** | 1. **Classical confounding and Berksonian bias (selection bias)**: corrected effect estimate.  2. **M bias**: lower bound of $R_{ED}$. |
| **Interpretation of the QBA analysis output** | 1. **Classical confounding and Berksonian bias (selection bias)**: effect estimate accounting for the corresponding unmeasured confounding and (or) selection bias.  2. **M bias**: lower limit the magnitude of $R_{ED}$. |
| **Considerations related to the interpretation of the results** | 1. The author noted that if the prevalence of confounder is rare (i.e., under 5%) in all E-D categories, the R_ED_ would approximate the crude OR between E and D.  2. The author suggested that in order to avoid collider-stratification bias, one should not control variables affected by exposure or disease.  3. For M bias, the author noted that the lower bound of the OR between E and D should be very close to 1 unless the common effect R is very large. |
| **Explicit discussions regarding the similarities and differences with other methods** | Not mentioned |

#### Flanders and Ye 2019 bounding formulas for m-bias and other structural selection bias

| **Name/description** | **Bounding formulas for m-bias and other structural selection bias** |
| --- | --- |
| **DOI** | 10.1097/EDE.0000000000001031 |
| **Year** | Flanders and Ye 2019^43^ |
| **Crossed-referenced in QBA textbook^3^** | Not referenced |
| **QBA classification** | Simple sensitivity analysis |
| **Applicable study design scenarios described^b^** | Cohort studies |
| **Effect measure of interest** | RR, and OR |
| **Result of interest** | Explain-away |
| **Data structure/format of the eligible exposure/intervention^b^** | Categorical |
| **Data structure/format of the confounders, if applicable^b^** | Not mentioned |
| **Data structure/format of eligible outcome^b^** | Categorical |
| **Source(s) of bias addressed** | Selection bias |
| **Information/parameters needed to conduct the QBA method** | -$\boldsymbol{R}_{\boldsymbol{E}}\mathbf{,}\boldsymbol{R}_{\boldsymbol{D}}$**, and** $\boldsymbol{R}_{\boldsymbol{C}}$: the associations between the factors $E,C,N,V$, and $D$, in Figure $1C$ :  $\begin{matrix} & R_{E}=\max_{v,e} \frac{p(V=v\mid E=e)}{P(V=v\mid E=1-e)}, \\ & R_{D}=\frac{\max_{n} P(D=1\mid E=0,N=n)}{\min_{n} P(D=1\mid E=0,N=n)}, \\ & R_{C}=\frac{\max_{v,n} P(C=c\mid V=v,N=n)}{\min_{v,n}P(C=c\mid V=v,N=n)} \end{matrix}$  Figure 1 C:  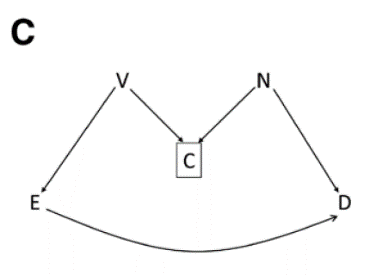  *Notes: C: collider; D: disease; E: exposure; N, V: variables on the E-D pathways as indicated in the picture.* |
| **Mathematical formula** | The upper bound of the bias is Bd, and the lower bound of the bias is 1/Bd:  $\begin{matrix} Bd & =\frac{R_{E}\left( \sqrt{R_{D}}R_{C}+1 \right)+\left( \sqrt{R_{D}}+R_{C} \right)}{R_{E}\left( \frac{R_{C}}{\sqrt{R_{D}}}+1 \right)+\left( \frac{1}{\sqrt{R_{D}}}+R_{C} \right)}\times\frac{\left( \frac{R_{C}}{\sqrt{R_{D}}}+1 \right)+R_{E}\left( \frac{1}{\sqrt{R_{D}}}+R_{C} \right)}{\left( \sqrt{R_{D}}R_{C}+1 \right)+R_{E}\left( \sqrt{R_{D}}+R_{C} \right)} \\ & =\left( \frac{R_{E}\left( \sqrt{R_{D}}R_{C}+1 \right)+\left( R_{C}+\sqrt{R_{D}} \right)}{\left( \sqrt{R_{D}}R_{C}+1 \right)+R_{E}\left( R_{C}+\sqrt{R_{D}} \right)} \right)^{2} \end{matrix}$  where $R_{E},R_{D}$, and $R_{C}$ measure the strengths of association between the factors $E,C,N,V$, and $D$, in Figure $1C$ *:  $\begin{matrix} & R_{E}=\max_{v,e} \frac{p(V=v\mid E=e)}{P(V=v\mid E=1-e)}, \\ & R_{D}=\frac{\max_{n} P(D=1\mid E=0,N=n)}{\min_{n} P(D=1\mid E=0,N=n)}, \\ & R_{C}=\frac{\max_{v,n} P(C=c\mid V=v,N=n)}{\min_{v,n}P(C=c\mid V=v,N=n)} \end{matrix}$  Figure 1:  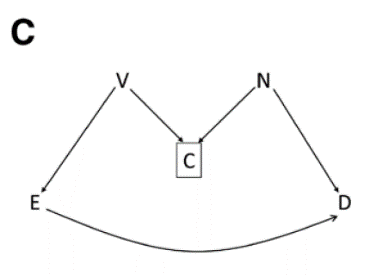  *Notes: C: collider; D: disease; E: exposure; N.V: variables on the E-D pathways as indicated in the picture. * Bound Bd can also be applied to other situations listed in Figure 2 of the paper.* |
| **Data assumptions and additional required features** | 1. This method does not require homogeneity assumption (i.e., all associations as measured by odds ratios are uniform) which is required by the method in Greenland 2003.^12^  2. Time-varying confounding and selection bias without colliders are not considered. |
| **Available and/or recommended software** | R program (eAppendix 4 of the paper) |
| **Output of the QBA analysis** | The bound Bd. |
| **Interpretation of the QBA analysis output** | The upper bound Bd presents the maximum selection bias and the lower bound 1/Bd presents the minimum selection bias. |
| **Considerations related to the interpretation of the results** | 1. The authors noted that “The approach works best and most plausibly if the variables are recognized but perhaps unmeasured, and information about the associations $R_{E},R_{D}$, and $R_{C}$ is available in the literature.”^43^  2. The authors noted that eAppendix 5 “also includes summaries of comparing *Bd* to *BdH*, focusing on robustness to more extreme violations of the homogeneity assumption used to derive *BdH*.”^43^  3. The authors noted that this method provides a tool for conducting sensitivity analyses without having to specify either the frequency of the potentially unobserved variables (U,V,C) or absolute risks. |
| **Explicit discussions regarding the similarities and differences with other methods** | 1. The author noted that their method is analogous to the work of Ding and VanderWeele,^49^ in the sense that they all developed bounds for the magnitude of confounding and used association between variables that create the bias. Moreover, the bound this method generated can also be used as the basis of the E-value.^48^  2. The authors compared their method to Greenland 2003’s method,^12^ and found absent homogeneity, their bound is meaningfully tighter than the bound generated by Greenland’s method. Moreover, the authors also compared their method to Lee 2011’s method,^40^ and found that Lee’s result is a special and limiting case.  3. The authors noted that their method is complementary to Huang and Lee 2015’s method, which only applies to ORs from case-control studies.^91^ |

#### Smith and VanderWeele 2019 E-value for selection bias

| **Name/description** | **E-value for selection bias** |
| --- | --- |
| **DOI** | 10.1097/EDE.0000000000001032 |
| **Year** | Smith and VanderWeele 2019^92^ |
| **Crossed-referenced in QBA textbook^3^** | Not referenced |
| **QBA classification** | Simple sensitivity analysis |
| **Applicable study design scenarios described^b^** | Any observational study designs |
| **Effect measure of interest** | RR, OR, HR, and RD |
| **Result of interest** | Corrected estimates (in **Method 1**)  Explain-away (in **Method 2**) |
| **Data structure/format of the eligible exposure/intervention^b^** | Categorical or continuous |
| **Data structure/format of the confounders, if applicable^b^** | Categorical or continuous |
| **Data structure/format of eligible outcome^b^** | Categorical, or time-to-event data |
| **Source(s) of bias addressed** | Selection bias |
| **Information/parameters needed to conduct the QBA method** | **Method 1**:  - **RR_UY\|(A =Sa)_** **parameters**: interpreted as the maximum relative risks for Y = 1 comparing any two values of U within strata of A = 1 and A = 0, respectively;  - **RR_SU\|(A =a)_** **parameters**: interpreted as maximum factors by which selection is associated with an increased prevalence of some value of U within stratum A = 1 and by which non-selection is associated with an increased prevalence of some value of U within stratum A = 0.  - crude **RR** and its 95% CIs.  **Method 2**:  - crude **RR** and its 95% CIs;  *Note: A: exposure; U: unmeasured confounder; S: selection; Y: outcome. Formula for risk difference is provided in the eTable of Appendix of the original manuscript.* |
| **Mathematical formula** | **Method 1**:  $Bounding factor=\left( \frac{\mathrm{RR}_{UY(A=1)}\times\mathrm{RR}_{SU\mid(A=1)}}{\mathrm{RR}_{UY\mid(A=1)}+\mathrm{RR}_{SU\mid(A=1)}-1} \right)\times\left( \frac{\mathrm{RR}_{UY\mid(A=0)}\times\mathrm{RR}_{SU\mid(A=0)}}{\mathrm{RR}_{UY\mid(A=0)}+\mathrm{RR}_{SU(A=0)}-1} \right)$;  Adjusted RR= Crude RR/bias factor (These formulas can be applied to the effect estimate and the corresponding 95% CIs).  **Method 2**:  $\mathrm{RR}_{UY\mid(A=0)}=\mathrm{RR}_{UY\mid(A=1)}=\mathrm{RR}_{SU\mid(A=0)}$ $=\mathrm{RR}_{SU\mid(A=1)}\geq\sqrt{\mathrm{RR}_{AY}^{\text{obs }}}+\sqrt{\mathrm{RR}_{AY}^{\text{obs }}-\sqrt{\mathrm{RR}_{AY}^{\text{obs }}}}$. |
| **Data assumptions and additional required features** | **Method 1 and 2**:  1. $Y\coprod S\mid\{A,U\}$  Which means outcome Y is independent of selection S conditional on exposure A and unmeasured confounder C.  2. $Y_{a}\coprod A\mid C$  No unmeasured confounding.  **Method 2 only**:  $\mathrm{RR}_{UY\mid(A=0)}=\mathrm{RR}_{UY\mid(A=1)}=\mathrm{RR}_{SU\mid(A=0)}=\mathrm{RR}_{SU\mid(A=1)}$, that is the maximum relative risks for Y = 1 comparing any two values of U within strata of A = 1 and A = 0, and then maximum factors by which selection is associated with an increased prevalence of some value of U within stratum A = 1 and by which non-selection is associated with an increased prevalence of some value of U within stratum A = 0 are equal. |
| **Available and/or recommended software** | R package EValue; Online calculator (http://selection-bias.louisahsmith.com/) |
| **Output of the QBA analysis** | **Method 1**: Adjusted RR and 95% CIs;  **Method 2**: minimum magnitude for any of the four (equal) parameters below  $\mathrm{RR}_{UY\mid(A=0)}=\mathrm{RR}_{UY\mid(A=1)}=\mathrm{RR}_{SU\mid(A=0)}=\mathrm{RR}_{SU\mid(A=1)}$ |
| **Interpretation of the QBA analysis output** | **Method 1**: RRs adjusted for sufficient selection bias  **Method 2**: To have selection bias fully explain away the observed association, the minimum magnitude of any of the four (equal) parameters below  $\mathrm{RR}_{UY\mid(A=0)}=\mathrm{RR}_{UY\mid(A=1)}=\mathrm{RR}_{SU\mid(A=0)}=\mathrm{RR}_{SU\mid(A=1)}$ |
| **Considerations related to the interpretation of the results** | 1. The authors noted that the result is context specific and should be interpreted relative to the selection mechanism in a given study and conditional on whatever confounders have been controlled for in the analysis.  2. The results describe the maximum bias that could result from the bias parameters. This conservative approach is useful when less is known about the selection mechanism and a simple exploration of the possible bias is desired.  3. The authors noted that this article only addresses bias due to selection and assumes that other criteria for causal inference, such as control of exposure–outcome confounding and lack of measurement error, have been met.  4. The methods can also be adapted to other effect measurements (e.g., ORs) and control selection bias only situations.^93^ |
| **Explicit discussions regarding the similarities and differences with other methods** | Previous QBA methods on selection bias including Huang and Lee 2015^91^, are limited by computational or mathematical complexity, the need for strong assumptions or the specification of a large number of parameters, or applicability only to certain selection mechanisms or study designs. Moreover, the authors noted that this simplified approach “to assessing selection bias makes clear the target of interference and limits the number of parameters and assumptions that determine the possible magnitude of the bias.” |

### Multiple biases

#### Turner et al 2009: method for biases in meta-analyses

| **Name/description** | **Method for biases in meta-analyses** |
| --- | --- |
| **DOI** | 10.1111/j.1467-985X.2008.00547.x |
| **Author and Year** | Turner et al 2009^94^ |
| **Crossed-referenced in QBA textbook^3^** | Not referenced |
| **QBA classification** | Multiple bias modeling |
| **Applicable study design scenarios described^b^** | Meta-analyses of observational study designs or randomized or nonrandomized trials |
| **Effect measure of interest** | RR or OR^b^ |
| **Result of interest** | Corrected estimates |
| **Data structure/format of the eligible exposure/intervention^b^** | Categorical^b^ |
| **Data structure/format of the confounders, if applicable^b^** | Categorical^b^ |
| **Data structure/format of eligible outcome^b^** | Categorical^b^ |
| **Source(s) of bias addressed** | Combination of selection bias, unmeasured confounding (single or multiple), exposure or outcome misclassification |
| **Information/parameters needed to conduct the QBA method** | Distributions for each possible bias in each category are required to be specified by each assessor(s) through marking estimated ranges on elicitation scales (a visual analog scale)  - $\boldsymbol{a}_{\boldsymbol{i\delta}}$: for additive bias δ in study i, the range limits $a_{i\delta}$ on the left-hand half of the elicitation scale;  - $\boldsymbol{b}_{\boldsymbol{i\delta}}$: for additive bias δ in study i, the range limits $b_{i\delta}$ on the right-hand half of the elicitation scale;  - $\boldsymbol{a}_{\boldsymbol{i\beta}}$: for proportional bias β in study i, the range limits $a_{i\beta}$ on the left-hand half of the elicitation scale;  - $\boldsymbol{b}_{\boldsymbol{i\beta}}$: for proportional bias β in study i, the range limits $b_{i\beta}$ on the right-hand half of the elicitation scale;  - $\boldsymbol{y}_{\boldsymbol{i}}$: effect estimate in the individual study i;  - $\boldsymbol{s}_{\boldsymbol{i}}^{\boldsymbol{2}}$: sampling variances of $y_{i}$.  *Note: According to the authors, additive biases are biases independent of the magnitude of the intervention effect, and proportional biases are biases which depend on the magnitude of the intervention effect.* |
| **Mathematical formula** | Procedures:  1. Define the *target* question.  2. Write down a mini-protocol for an *idealized* version of each study.  3. Identify each category of internal biases by comparing the original study against the *idealized* study. A checklist for sources of internal bias is presented in Table 1 of Turner et al 2009.^94^  4. Identify external biases by comparing the *idealized* study against the *target* question. A checklist for sources of external bias is presented in Table 2 of Turner et al 2009.^94^  5. Identify additive and proportional biases among the internal and external biases listed in Step 3 and Step 4.  6. Specify distributions of each bias in each study included in the meta-analysis. In particular, each assessor is given a copy of an elicitation scale for additive biases (in Figure 2 in Turner et al 2009^94^) for each bias and the other scale for proportional biases (in Figure 7 in Turner et al 2009^94^). As noted by the authors, the assessors were “asked to independently mark a 67% range for the relative risk of the adverse outcome chosen in that study such that they feel that the true answer to the question is twice as likely to lie inside rather than outside this range.” It is recommended by the authors that “assessors first make a qualitative judgement of the severity of bias, before quantifying their opinion as a 67% range. Assessors write down their judgement of the severity of bias (as none, low, medium or high) in favor of the intervention and, separately, the severity of bias in favor of the control. Th authors suggested the following correspondence between qualitative judgements of severity and choices for the range limit: none (1); low (0.9–1); medium (0.7–0.9); high (less than 0.7).”^94^  7. Combine the effects of additive biases (δ) and proportional biases (β) during adjustment of each study i’s results for each assessor. Formulas are listed below:  $\begin{matrix} \mu_{i\delta} & =\frac{log\left( a_{i\delta} \right)+log\left( 1/b_{i\delta} \right)}{2} \\ \sigma_{i\delta} & =\frac{log\left( 1/b_{i\delta} \right)-log\left( a_{i\delta} \right)}{2} \end{matrix}$  $\begin{matrix} \mu_{i\beta} & =\frac{log\left( a_{i\beta} \right)+log\left( 1/b_{i\beta} \right)}{2} \\ \sigma_{i\beta} & =\frac{log\left( 1/b_{i\beta} \right)-log\left( a_{i\beta} \right)}{2}. \end{matrix}$  Intervention effect in study i: $\hat{\theta}_{i}=\left( y_{i}-\mu_{i\beta}^{I}\mu_{i\delta}^{E}-\mu_{i\delta}^{I} \right)/\mu_{i\beta}^{I}\mu_{i\beta}^{E}$.  Standardized error of $\hat{\theta}_{i},$ $SE(\hat{\theta}_{i})=\left( \frac{1}{\mu_{i\beta}^{I}\mu_{i\beta}^{E}} \right)^{2}\left\{ s_{i}^{2}+\left( {\mu_{i\beta}^{I}}^{2}+\sigma_{i\beta}^{I}{}^{2} \right)\left( \hat{\theta}^{2}{\sigma_{i\beta}^{E}}^{2}+{\sigma_{i\delta}^{E}}^{2} \right)+{\sigma_{i\beta}^{I}}^{2}\left( \hat{\theta}\mu_{i\beta}^{E}+\mu_{i\delta}^{E} \right)^{2}+{\sigma_{i\delta}^{I}}^{2} \right\}$.  8. Pool range of biases across different assessors using a standard random-effects model for each study.  9. Pool bias-adjusted effect estimates across studies to obtain a bias-adjusted effect estimate.  *Note: the superscript of I and E are used to note internal and external biases, respectively.* |
| **Data assumptions and additional required features** | 1. All biases can be categorized as either additive or proportional. This is a simplification, noted by the authors.  2. The effect estimate in the individual study i, $y_{i}$, follow the distribution below, where the notation f(μ, $\sigma^{2}$) represents a general distribution with mean μ and variance $\sigma^{2}$. $y_{i}\sim f\left\{ \mu_{i\beta}^{I}\left( \mu_{i\beta}^{E}\theta+\mu_{i\delta}^{E} \right)+\mu_{i\delta}^{I},s_{i}^{2}+\left( \mu_{i\beta}^{I}2+\sigma_{i\beta}^{I}{}^{2} \right)\left( \theta^{2}\sigma_{i\beta}^{E2}+\sigma_{i\delta}^{E2} \right)+\sigma_{i\beta}^{I}2\left( \theta\mu_{i\beta}^{E}+\mu_{i\delta}^{E} \right)^{2}+{\sigma_{i\delta}^{I}}^{2}+\tau^{2} \right\}$. |
| **Available and/or recommended software** | Not mentioned |
| **Output of the QBA analysis** | Corrected effect estimates |
| **Interpretation of the QBA analysis output** | Effect estimates accounting for internal biases (selection bias, unmeasured confounding, exposure or outcome misclassification) and external biases (population bias, intervention bias, etc.) |
| **Considerations related to the interpretation of the results** | 1. The authors noted that although they have based all distributions on elicited opinion alone, the bias distributions can also be informed by empirical evidence obtained from meta-epidemiological studies.  2. The authors noted that they chose to model biases as either additive or proportional, as a simplification for many circumstances, and that it is possible to incorporate more complex forms of adjustment for certain biases, to their approach.  3. The authors noted that it is sufficient to elicit opinions from four or five assessors.  4. The authors noted that although it is possible to give more weight to certain assessors’ opinions on the basis of their greater  knowledge in particular areas, selection of such weights would be difficult in practice. In cases of divergent opinions between assessors, it should be acknowledged through discussion and presentation of bias-adjusted meta-analyses for each assessor separately, rather than to combine the opinions statistically.  5. Because the authors did not take into account potential correlation of biases included in their checklist, they expect their degree of variance adjustment to be conservative, meaning that the study results should be down-weighted further.  6. The authors noted that “When deciding how to handle biases in evidence synthesis, researchers should consider whether their aim is to answer a specific target question by using the evidence which is currently available, or to summarize the literature.”. They also noted that their method is developed for the former (that is to answer a specific target question by using the evidence which is currently available, and not to merely summarize the literature).  7. The authors noted that their method is presented in the context of a two-arm comparison of a binary outcome, but it could be modified for other settings, for example, a non-comparative target setting.  8. The authors noted that Step 8 and Step 9 could be conducted alternatively by pooling the data first across different studies for each assessor first, and then pooling the data across different assessors. |
| **Explicit discussions regarding the similarities and differences with other methods** | Mathur and VanderWeele 2020^56^ noted that it “may be difficult for meta-analysts to accurately specify numerical ranges of bias for each study”. They also noted that “moments-based meta-analysis estimation can perform poorly”. |

#### Huang and Lee 2015 bounding formulas for selection Bias

| **Name/description** | **Bounding formulas for selection bias** |
| --- | --- |
| **DOI** | 10.1093/aje/kwv130 |
| **Year** | Huang and Lee 2015^91^ |
| **Crossed-referenced in QBA textbook^3^** | Not referenced |
| **QBA classification** | Multiple bias modeling^a^ |
| **Applicable study design scenarios described^b^** | Any observational study designs (examples are case-control studies) |
| **Effect measure of interest** | OR^b^ |
| **Result of interest** | Explain-away |
| **Data structure/format of the eligible exposure/intervention^b^** | Categorical^b^ |
| **Data structure/format of the confounders, if applicable^b^** | Categorical |
| **Data structure/format of eligible outcome^b^** | Categorical |
| **Source(s) of bias addressed** | Selection bias  Unmeasured confounding |
| **Information/parameters needed to conduct the QBA method** | - **S_E_**: odds ratio for the association between U and E  - **S_D_**: odds ratio for the association between U and D  - **S_M_**: when U is also an effect modifier, the modifying effect U has on the association between E and D  - **S_F_**: odds ratio for the association between U and R  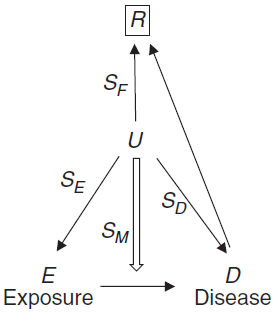*Note: D: disease; E: exposure; R: recruitment of study subjects; U: unmeasured confounder.*  DAG: |
| **Mathematical formula** | When the unexposed is taken as standard population, the upper and lower bounds of the odds ratio are:  Upper $=\frac{1}{\text{ Lower }}\leq\left( \frac{\sqrt{S_{E}\times S_{D}\times S_{M}\times S_{F}}+1}{\sqrt{S_{E}}+\sqrt{S_{D}\times S_{M}\times S_{F}}} \right)^{2}$;  *Note: special conditions:*  *1) The authors mentioned that if one has difficulties in assigning values of the four parameters, one can simply set the value to infinity but this may result in broader bounds.*  *2) If U is not associated with the recruitment process for the controls, then one can set SF = 1.* |
| **Data assumptions and additional required features** | Not specified |
| **Available and/or recommended software** | R code available in supplementary materials of the original manuscript |
| **Output of the QBA analysis** | The upper and lower bounds of the OR/RR |
| **Interpretation of the QBA analysis output** | If the bound overlaps with the effect estimate and/or its corresponding 95% CI, then the possibility of selection and/or unmeasured confounding cannot be ruled out. |
| **Considerations related to the interpretation of the results** | 1. This is a conservative way to think about the effects of biases, as noted in the original manuscript: “the formulas established in this paper are to be used in a worst-case scenario for bounding every possible unknown or unmeasured factor having the potential to create biases.”  2. The exact values of the 4 strength parameters are not required to use the formulas, therefore exaggerated values can be used to err on the conservative side.  3. One can set the corresponding strength parameter to infinity if one has difficulties in gauging U in any aspects of its strengths. |
| **Explicit discussions regarding the similarities and differences with other methods** | 1. Different from previous methods including Rosenbaum and Rubin 1983^8^, VanderWeele and Arah 2011^11^, and Steenland and Greenland 2004^13^, this method does not require one to know the exact values of the 4 strength parameters and one can err on the side by exaggerating the values.  2. The authors extended the bounding formulas for unmeasured confounding and effect modification in Lee 2011^40^ to include selection bias.  3. Flanders and Ye 2019^43^ noted that their method is complementary to this method since their method applies to RRs from cohort studies while this one works for ORs from case-control studies. |

#### Smith et al 2021 bounding method for multiple biases

| **Name/description** | **Bounding method for multiple biases** |
| --- | --- |
| **DOI** | 10.1097/EDE.0000000000001380 |
| **Year** | Smith et al 2021^95^ |
| **Crossed-referenced in QBA textbook^3^** | Not referenced |
| **QBA classification** | Multiple bias modeling^a^ |
| **Applicable study design scenarios described^b^** | Any observational study designs |
| **Effect measure of interest** | RR, and OR |
| **Result of interest** | Corrected estimate |
| **Data structure/format of the eligible exposure/intervention^b^** | Categorical^b^ |
| **Data structure/format of the confounders, if applicable^b^** | Categorical or continuous |
| **Data structure/format of eligible outcome^b^** | Categorical^b^ |
| **Source(s) of bias addressed** | Any combination of selection bias, unmeasured confounding, and misclassification bias (differential or nondifferential) |
| **Information/parameters needed to conduct the QBA method** | **Misclassification parameter:**  - **RR_AY* \|y. S=1_**: interpreted as the maximum of the false-positive probability ratio or sensitivity ratio within the selected population  **Selection bias parameters:**  - **RR_UsY \|A=a_**: interpreted as the maximum factors by which the outcome risk differs by values of Us, within strata of A  - **RR_SUs \|A=a_**: interpreted as the maximum factors by which some level of Us differs between the selected and nonselected groups, within strata of A  **Unmeasured confounding parameters:**  - **RR_UcY_**: interpreted as the maximum factor by which Uc increases the outcome risk, conditional on A  - **RR_AUc_**: interpreted as the maximum factor by which exposure is associated with some value of Uc  *Note: A: exposure; A*: misclassified exposure; C: measured covariates; S: selection; Uc: unmeasured confounder(s); Y: outcome; Y*: misclassified outcome.* |
| **Mathematical formula** | Specific bounding formulas regarding different combinations of biases can be found in Table of the paper.  Note:$g(a,b)=\frac{a\times b}{a+b-1}$. |
| **Data assumptions and additional required features** | General assumptions:  1. conditional exchangeability: potential outcomes are independent of exposure status conditional on C and U_c_  $Y_{a}\coprod A\mid C,U_{c}$  2. They consider only misclassification of the exposure or the outcome but not both.  Specific assumptions regarding different combinations of biases can be found in the Table of the original manuscript. |
| **Available and/or recommended software** | R package EValue |
| **Output of the QBA analysis** | The upper and lower bounds of the OR/RR which can be used to calculate the bias-corrected estimates |
| **Interpretation of the QBA analysis output** | The effect estimates adjusted for corresponding biases |
| **Considerations related to the interpretation of the results** | 1. When specifying values for the bias parameters, one should clearly specify related components. For instance, when adjusting unmeasured confounding, one should be clear about the confounders that are taken into account.  2. The authors noted that the bound should be interpreted as the bias that could result from parameters of a given magnitude not that necessarily would result, which describes a “worst-case scenario” for the bias.  3. Although this approach takes into account all three biases, if any of the three is judged not to threaten a given study or to bias toward the null, that bias can be omitted.  4. If data exist to justify estimates of the alternative parameters (which are available in the eAppendix of the original paper), different ordering of the biases may be possible (in the original paper, the authors proposed to adjust confounding last, and selection or misclassification bias first, depending on the specific study design).  5. The bound for exposure misclassification relies on the rare outcome assumption.  6. It is noted that the interpretation of the bias-adjusted 95% CIs pertains to the application of the adjustment to repeated samples with the same sets of biases; if the biases were truly resolved in the design or analysis, the bounds would differ. |
| **Explicit discussions regarding the similarities and differences with other methods** | The approach is built upon previous bounding methods, including VanderWeele and Li 2019^81^, Smith and VanderWeele 2019^92^, VanderWeele and Ding 2017^48^, and Ding and VanderWeele 2016^49^. |

**Footnote**: ^a^: multiple bias modeling but with simple analyses methods, not computationally intensive.

^b^: modification to the original method(s) can be made so that it may also adapt to other designs or variables with other data structure.

References

1.	Bross ID. Spurious effects from an extraneous variable. *J. Chronic Dis.* 1966;19(6):637-647.

2.	Schneeweiss S. Sensitivity analysis and external adjustment for unmeasured confounders in epidemiologic database studies of therapeutics. *Pharmacoepidemiol. Drug Saf.* 2006;15(5):291-303.

3.	Fox MP, MacLehose RF, Lash TL. *Applying Quantitative Bias Analysis to Epidemiologic Data.* Springer Nature; 2021.

4.	Zhang MF, Falconer M, Taylor L. A quantitative bias analysis of the confounding effects due to smoking on the association between fluoroquinolones and risk of aortic aneurysm. *Pharmacoepidemiol. Drug Saf.* 2020;29(8):958-961.

5.	Schlesselman JJ. Assessing effects of confounding variables. *Am. J. Epidemiol.* 1978;108(1):3-8.

6.	Lin DY, Psaty BM, Kronmal RA. Assessing the sensitivity of regression results to unmeasured confounders in observational studies. *Biometrics.* 1998;54(3):948-963.

7.	Cornfield J, Haenszel W, Hammond EC, Lilienfeld AM, Shimkin MB, Wynder EL. Smoking and Lung Cancer: Recent Evidence and a Discussion of Some Questions. *JNCI: Journal of the National Cancer Institute.* 1959;22(1):173-203.

8.	Rosenbaum PR, Rubin DB. Assessing Sensitivity to an Unobserved Binary Covariate in an Observational Study with Binary Outcome. *Journal of the Royal Statistical Society. Series B (Methodological).* 1983;45(2):212-218.

9.	Groenwold RHH, Nelson DB, Nichol KL, Hoes AW, Hak E. Sensitivity analyses to estimate the potential impact of unmeasured confounding in causal research. *Int. J. Epidemiol.* 2009;39(1):107-117.

10.	Flanders WD, Khoury MJ. Indirect assessment of confounding: graphic description and limits on effect of adjusting for covariates. *Epidemiology.* 1990;1(3):239-246.

11.	Vanderweele TJ, Arah OA. Bias formulas for sensitivity analysis of unmeasured confounding for general outcomes, treatments, and confounders. *Epidemiology.* 2011;22(1):42-52.

12.	Greenland S. Quantifying Biases in Causal Models: Classical Confounding vs Collider-Stratification Bias. *Epidemiology.* 2003;14(3):300-306.

13.	Steenland K, Greenland S. Monte Carlo sensitivity analysis and Bayesian analysis of smoking as an unmeasured confounder in a study of silica and lung cancer. *Am. J. Epidemiol.* 2004;160(4):384-392.

14.	Cabral MD, Luiz RR. Use of sensitivity analysis to assess the effects on anti-hepatitis A virus antibodies of access to household water supply. *Epidemiol. Infect.* 2008;136(3):334-340.

15.	Cabral MD, Luiz RR. Sensitivity analysis for unmeasured confounders using an electronic spreadsheet. *Rev. Saude Publica.* 2007;41(3):446-452.

16.	GREENLAND S. Basic Methods for Sensitivity Analysis of Biases. *Int. J. Epidemiol.* 1996;25(6):1107-1116.

17.	Arah OA, Chiba Y, Greenland S. Bias Formulas for External Adjustment and Sensitivity Analysis of Unmeasured Confounders. *Ann. Epidemiol.* 2008;18(8):637-646.

18.	Lash TL, Schmidt M, Jensen AØ, Engebjerg MC. Methods to apply probabilistic bias analysis to summary estimates of association. *Pharmacoepidemiol. Drug Saf.* 2010;19(6):638-644.

19.	Rothman KJ, Greenland S, Lash TL. *Modern epidemiology.* Vol 3: Wolters Kluwer Health/Lippincott Williams & Wilkins Philadelphia; 2008.

20.	Chiba Y. Sensitivity Analysis for Unmeasured Confounding of Attributable Fraction. *Epidemiology.* 2012;23(1):175-176.

21.	Brumback BA, Hernán MA, Haneuse SJ, Robins JM. Sensitivity analyses for unmeasured confounding assuming a marginal structural model for repeated measures. *Stat. Med.* 2004;23(5):749-767.

22.	McCandless LC. Meta-analysis of observational studies with unmeasured confounders. *Int. J. Biostat.* 2012;8(2).

23.	Carlin JB. Meta-analysis for 2 x 2 tables: a Bayesian approach. *Stat. Med.* 1992;11(2):141-158.

24.	Lambert PC, Sutton AJ, Burton PR, Abrams KR, Jones DR. How vague is vague? A simulation study of the impact of the use of vague prior distributions in MCMC using WinBUGS. *Stat. Med.* 2005;24(15):2401-2428.

25.	Vanderweele TJ, Mukherjee B, Chen J. Sensitivity analysis for interactions under unmeasured confounding. *Stat. Med.* 2012;31(22):2552-2564.

26.	Lubin JH, Hauptmann M, Blair A. Indirect adjustment of relative risks of an exposure with multiple categories for an unmeasured confounder. *Ann. Epidemiol.* 2018;28(11):801-807.

27.	Frank KA. Impact of a confounding variable on a regression coefficient. *Sociological Methods & Research.* 2000;29(2):147-194.

28.	Rosenbaum PR. Dropping out of High School in the United States: An Observational Study. *Journal of Educational Statistics.* 1986;11(3):207-224.

29.	Cinelli C, Hazlett C. Making sense of sensitivity: Extending omitted variable bias. *Journal of the Royal Statistical Society: Series B (Statistical Methodology).* 2020;82(1):39-67.

30.	Phillips CV. Quantifying and reporting uncertainty from systematic errors. *Epidemiology.* 2003;14(4):459-466.

31.	Yu B, Gastwirth JL. Sensitivity analysis for trend tests: application to the risk of radiation exposure. *Biostatistics.* 2005;6(2):201-209.

32.	Rosenbaum PR. Does a dose-response relationship reduce sensitivity to hidden bias? *Biostatistics.* 2003;4(1):1-10.

33.	Rosenbaum PR. *Observational Studies.* Springer; 2002.

34.	Cornfield J, Haenszel W, Hammond EC, Lilienfeld AM, Shimkin MB, Wynder EL. Smoking and lung cancer: recent evidence and a discussion of some questions. *J. Natl. Cancer Inst.* 1959;22(1):173-203.

35.	Lee W-C, Wang L-Y. Simple Formulas for Gauging the Potential Impacts of Population Stratification Bias. *Am. J. Epidemiol.* 2007;167(1):86-89.

36.	Wacholder S, Rothman N, Caporaso N. Population stratification in epidemiologic studies of common genetic variants and cancer: quantification of bias. *J. Natl. Cancer Inst.* 2000;92(14):1151-1158.

37.	Wang Y, Localio R, Rebbeck TR. Evaluating bias due to population stratification in case-control association studies of admixed populations. *Genet. Epidemiol.* 2004;27(1):14-20.

38.	Heiman GA, Hodge SE, Gorroochurn P, Zhang J, Greenberg DA. Effect of population stratification on case-control association studies. I. Elevation in false positive rates and comparison to confounding risk ratios (a simulation study). *Hum. Hered.* 2004;58(1):30-39.

39.	Gorroochurn P, Hodge SE, Heiman G, Greenberg DA. Effect of population stratification on case-control association studies. II. False-positive rates and their limiting behavior as number of subpopulations increases. *Hum. Hered.* 2004;58(1):40-48.

40.	Lee WC. Bounding the bias of unmeasured factors with confounding and effect-modifying potentials. *Stat. Med.* 2011;30(9):1007-1017.

41.	VanderWeele TJ. The sign of the bias of unmeasured confounding. *Biometrics.* 2008;64(3):702-706.

42.	Chiba Y. The sign of the unmeasured confounding bias under various standard populations. *Biom. J.* 2009;51(4):670-676.

43.	Flanders WD, Ye D. Limits for the Magnitude of M-bias and Certain Other Types of Structural Selection Bias. *Epidemiology.* 2019;30(4):501-508.

44.	Hasegawa R, Small D. Sensitivity analysis for matched pair analysis of binary data: From worst case to average case analysis. *Biometrics.* 2017;73(4):1424-1432.

45.	Rosenbaum PR. Sensitivity Analysis for Certain Permutation Inferences in Matched Observational Studies. *Biometrika.* 1987;74(1):13-26.

46.	Rosenbaum PR. Overt Bias in Observational Studies. In: Rosenbaum PR, ed. *Observational Studies*. New York, NY: Springer New York; 2002:71-104.

47.	Rosenbaum PR. Attributing effects to treatment in matched observational studies. *Journal of the American statistical Association.* 2002;97(457):183-192.

48.	VanderWeele TJ, Ding P. Sensitivity Analysis in Observational Research: Introducing the E-Value. *Ann. Intern. Med.* 2017;167(4):268-274.

49.	Ding P, VanderWeele TJ. Sensitivity Analysis Without Assumptions. *Epidemiology.* 2016;27(3):368-377.

50.	Cusson A, Infante-Rivard C. Bias factor, maximum bias and the E-value: insight and extended applications. *Int. J. Epidemiol.* 2020;49(5):1509-1516.

51.	Linden A, Mathur MB, VanderWeele TJ. Conducting sensitivity analysis for unmeasured confounding in observational studies using E-values: The evalue package. *STATA JOURNAL.* 2020;20(1):162-175.

52.	Haneuse S, VanderWeele TJ, Arterburn D. Using the E-Value to Assess the Potential Effect of Unmeasured Confounding in Observational Studies. *JAMA.* 2019;321(6):602-603.

53.	MacLehose RF, Ahern TP, Lash TL, Poole C, Greenland S. The Importance of Making Assumptions in Bias Analysis. *Epidemiology.* 2021;32(5).

54.	Ioannidis JPA, Tan YJ, Blum MR. Limitations and Misinterpretations of E-Values for Sensitivity Analyses of Observational Studies. *Ann. Intern. Med.* 2019;170(2):108-111.

55.	VanderWeele TJ, Mathur MB, Ding P. Correcting Misinterpretations of the E-Value. *Ann. Intern. Med.* 2019;170(2):131-132.

56.	Mathur MB, VanderWeele TJ. Sensitivity Analysis for Unmeasured Confounding in Meta-Analyses. *J Am Stat Assoc.* 2020;115(529):163-172.

57.	Mathur MB, VanderWeele TJ. New metrics for meta-analyses of heterogeneous effects. *Stat. Med.* 2019;38(8):1336-1342.

58.	Mathur MB, VanderWeele TJ. Robust Metrics and Sensitivity Analyses for Meta-analyses of Heterogeneous Effects. *Epidemiology.* 2020;31(3):356-358.

59.	Wang CC, Lee WC. A simple method to estimate prediction intervals and predictive distributions: Summarizing meta-analyses beyond means and confidence intervals. *Res Synth Methods.* 2019;10(2):255-266.

60.	Copeland KT, Checkoway H, McMichael AJ, Holbrook RH. Bias due to misclassification in the estimation of relative risk. *Am. J. Epidemiol.* 1977;105(5):488-495.

61.	Barron BA. The effects of misclassification on the estimation of relative risk. *Biometrics.* 1977;33(2):414-418.

62.	GREENLAND S. THE EFFECT OF MISCLASSIFICATION IN MATCHED-PAIR CASE-CONTROL STUDIES. *Am. J. Epidemiol.* 1982;116(2):402-406.

63.	GREENLAND S, KLEINBAUM DG. Correcting for Misclassification in Two-Way Tables and Matched-Pair Studies. *Int. J. Epidemiol.* 1983;12(1):93-97.

64.	Marshall RJ. Validation study methods for estimating exposure proportions and odds ratios with misclassified data. *J. Clin. Epidemiol.* 1990;43(9):941-947.

65.	Barron BA. The Effects of Misclassification on the Estimation of Relative Risk. *Biometrics.* 1977;33(2):414-418.

66.	Checkoway H, Savitz DA, Heyer NJ. Assessing the effects of nondifferential misclassification of exposures in occupational studies. *Appl. Occup. Environ. Hyg.* 1991;6(6):528-533.

67.	Kaldor J, Clayton D. Latent class analysis in chronic disease epidemiology. *Stat. Med.* 1985;4(3):327-335.

68.	Espeland MA, Hui SL. A General Approach to Analyzing Epidemiologic Data that Contain Misclassification Errors. *Biometrics.* 1987;43(4):1001-1012.

69.	Wacholder S, Armstrong B, Hartge P. Validation Studies using an Alloyed Gold Standard. *Am. J. Epidemiol.* 1993;137(11):1251-1258.

70.	Rosner B, Willett WC, Spiegelman D. Correction of logistic regression relative risk estimates and confidence intervals for systematic within-person measurement error. *Stat. Med.* 1989;8(9):1051-1069; discussion 1071-1053.

71.	Marshall RJ. Misclassification of exposure in case-control studies: assessment by quality indices. *Epidemiology.* 1994;5(3):309-314.

72.	Brenner H. Correcting for exposure misclassification using an alloyed gold standard. *Epidemiology.* 1996;7(4):406-410.

73.	Weinkam JJ, Rosenbaum WL, Sterling TD. Recovering true risks when multilevel exposure and covariables are both misclassified. *Am. J. Epidemiol.* 1999;150(8):886-891.

74.	Fung KY, Howe GR. Methodological issues in case-control studies. III--The effect of joint misclassification of risk factors and confounding factors upon estimation and power. *Int. J. Epidemiol.* 1984;13(3):366-370.

75.	Chu H, Wang Z, Cole SR, Greenland S. Sensitivity Analysis of Misclassification: A Graphical and a Bayesian Approach. *Ann. Epidemiol.* 2006;16(11):834-841.

76.	Gustafson P, Le ND, Saskin R. Case-control analysis with partial knowledge of exposure misclassification probabilities. *Biometrics.* 2001;57(2):598-609.

77.	Fox MP, Lash TL, Greenland S. A method to automate probabilistic sensitivity analyses of misclassified binary variables. *Int. J. Epidemiol.* 2005;34(6):1370-1376.

78.	Liu J, Gustafson P, Cherry N, Burstyn I. Bayesian analysis of a matched case-control study with expert prior information on both the misclassification of exposure and the exposure-disease association. *Stat. Med.* 2009;28(27):3411-3423.

79.	Prescott GJ, Garthwaite PH. Bayesian analysis of misclassified binary data from a matched case–control study with a validation sub‐study. *Stat. Med.* 2005;24(3):379-401.

80.	Gustafson P, Le ND, Saskin R. Case–control analysis with partial knowledge of exposure misclassification probabilities. *Biometrics.* 2001;57(2):598-609.

81.	VanderWeele TJ, Li Y. Simple Sensitivity Analysis for Differential Measurement Error. *Am. J. Epidemiol.* 2019;188(10):1823-1829.

82.	Baum U, Kulathinal S, Auranen K. Exposure misclassification bias in the estimation of vaccine effectiveness. *PLoS One.* 2021;16(5):e0251622.

83.	Tang L, Lyles RH, King CC, Celentano DD, Lo Y. Binary regression with differentially misclassified response and exposure variables. *Stat. Med.* 2015;34(9):1605-1620.

84.	GREEN MS. USE OF PREDICTIVE VALUE TO ADJUST RELATIVE RISK ESTIMATES BIASED BY MISCLASSIFICATION OF OUTCOME STATUS1. *Am. J. Epidemiol.* 1983;117(1):98-105.

85.	Farrington CP. Quantifying misclassification bias in cohort studies of vaccine efficacy. *Stat. Med.* 1990;9(11):1327-1337.

86.	Magder LS. Simple approaches to assess the possible impact of missing outcome information on estimates of risk ratios, odds ratios, and risk differences. *Control. Clin. Trials.* 2003;24(4):411-421.

87.	Jurek AM, Maldonado G, Greenland S. Adjusting for outcome misclassification: the importance of accounting for case-control sampling and other forms of outcome-related selection. *Ann. Epidemiol.* 2013;23(3):129-135.

88.	Endo A, Funk S, Kucharski AJ. Bias correction methods for test-negative designs in the presence of misclassification. *Epidemiol. Infect.* 2020;148:e216.

89.	Savitz DA, Barón AE. Estimating and correcting for confounder misclassification. *Am. J. Epidemiol.* 1989;129(5):1062-1071.

90.	Nab L, Groenwold RHH, van Smeden M, Keogh RH. Quantitative Bias Analysis for a Misclassified Confounder: A Comparison Between Marginal Structural Models and Conditional Models for Point Treatments. *Epidemiology.* 2020;31(6):796-805.

91.	Huang TH, Lee WC. Bounding formulas for selection bias. *Am. J. Epidemiol.* 2015;182(10):868-872.

92.	Smith LH, VanderWeele TJ. Bounding Bias Due to Selection. *Epidemiology.* 2019;30(4):509-516.

93.	Smith LH, VanderWeele TJ. Simple sensitivity analysis for control selection bias. *Epidemiology.* 2020;31(5):e44-e45.

94.	Turner RM, Spiegelhalter DJ, Smith GC, Thompson SG. Bias modelling in evidence synthesis. *Journal of the Royal Statistical Society Series A: Statistics in Society.* 2009;172(1):21-47.

95.	Smith LH, Mathur MB, VanderWeele TJ. Multiple-bias Sensitivity Analysis Using Bounds. *Epidemiology.* 2021;32(5):625-634.
